# U.S. state-level COVID-19 transmission insights from a mechanistic mobility-incidence model

**DOI:** 10.1101/2022.06.21.22276712

**Authors:** Edward W. Thommes, Zahra Mohammadi, Darren Flynn-Primrose, Sarah Smook, Gabriela Gomez, Sandra S. Chaves, Laurent Coudeville, Robertus Van Aalst, Cedric Mahé, Monica G. Cojocaru

**Affiliations:** Sanofi; University of Guelph, Guelph, Ontario, Canada; York University, Toronto, Ontario, Canada; McMaster University, Hamilton, Ontario, Canada; London School of Hygiene and Tropical Medicine, London, U.K.; Brown University School of Public Health, Providence, Rhode Island, USA; University of Groningen, Groningen, Netherlands

## Abstract

**Background:** Throughout the COVID-19 pandemic, human mobility has played a central role in shaping disease transmission. In this study, we develop a mechanistic model to calculate disease incidence from commercially-available US mobility data over the course of 2020. We use it to study, at the US state level, the lag between infection and case report. We examine the evolution of per-contact transmission probability, and its dependence on mean air temperature. Finally, we evaluate the potential of the model to produce short-term incidence forecasts from mobility data.

**Methods:** We develop a mechanistic model that relates COVID-19 incidence to time series contact index (CCI) data collected by mobility data vendor Cuebiq. From this, we perform maximum-likelihood estimates of the transmission probability per CCI event. Finally, we retrospectively conduct forecasts from multiple dates in 2020 forward.

**Findings:** Across US states, we find a median lag of 19 days between transmission and case report. We find that the median transmission probability from May onward was about 20% lower than it was during March and April. We find a moderate, statistically significant negative correlation between mean state temperature and transmission probability, *r* = − .57, *N* = 49, *p* = 2 × 10^−5^. We conclude that for short-range forecasting, CCI data would likely have performed best overall during the first few months of the pandemic.

**Interpretation:** Our results are consistent with associations between colder temperatures and stronger COVID-19 burden reported in previous studies, and suggest that changes in the per-contact transmission probability play an important role. Our model displays good potential as a short-range (2 to 3 week) forecasting tool during the early stages of a future pandemic, before non-pharmaceutical interventions (NPIs) that modify per-contact transmission probability, principally face masks, come into widespread use. Hence, future development should also incorporate time series data of NPI use.

## 1. Introduction

As of end of early June 2022, the global COVID-19 pandemic has produced 530M recorded cases and 6.3M recorded deaths worldwide^1^. Throughout its course, the complex epidemiology of the disease has been shaped, above all, by the changes in human behavior it has elicited. Indeed, in a counterfactual world that took no measures against it, the course of the pandemic would have been simple and catastrophic; it is estimated [1]^2^ that about 90% of the world’s population would have been infected in a single massive wave lasting roughly two months, with a death toll of about 40 million.

The most immediate response consisted of the near-universal lock-downs which began in rapid succession around the world in spring of 2020. The publication of freely-available worldwide human mobility data by Google^3^, Apple^4^ and Facebook^5^, and the re-purposing of business intelligence mobility data from vendors such as Cuebiq^6^ and Safegraph^7^, has made it possible to trace changes in mobility with high spatial and temporal resolution, and to directly observe the results of mobility-related measures enacted to counter disease transmission. Numerous studies have examined the connection between mobility and COVID-19 epidemiology. Some use statistical models to characterize associations between measures of mobility and measures of disease burden (e.g. [2], [3], [4], [5]). Others use hybrid approaches that combine statistical and mechanistic models (e.g. [6] [7] [8]), in some cases with the help of artificial intelligence (e.g. [9]). Many further examples are given in the systematic review of Zhang et al.[10].

Our approach here is almost entirely mechanistic. Changes in human mobility affect disease transmission by modifying the rate of person-to-person contacts. Most available mobility data is in the form of indices that are indirect proxies for contact rate, and which require additional work (e.g. [11]) to infer contact rate itself. Here, we use data from US mobility data provider Cuebiq, which probes person-to-person contact rate more directly (see Section 3), thus lending itself better to use in a mechanistic model. This allows us to estimate, within a proportionality constant, the per-contact transmission probability of the disease. We restrict our analysis to 2020 in order to avoid complication due to i) emergence of new variants, ii) vaccination and iii) significant accumulation of post-infection natural immunity in the population.

In 2020, prior to the availability of COVID vaccines, the evolution of per-contact transmission probability over time within a given region reflected the time-varying practice of non-pharmaceutical interventions (NPIs), notably mask-wearing and short-range social distancing (i.e. maintaining a minimum separation of e.g. 6ft among people). Note that the latter is technically encompassed within the contact rate, however the mobility data we use does not have a high enough spatial resolution to discern the degree to which distancing on the scale of a few meters is practiced.

An association of colder temperatures/climates with different measures of COVID-19 burden has been reported in multiple studies (see [12] for a 2020 systematic review; more recent studies include [13], [14] and [15]). Using our model results, we compare the transmission probability across US states during spring of 2020, at the onset of the pandemic.

Finally, we take an exploratory look at the potential of Cuebiq mobility data for short-term forecasting.

## 2. Methods

In this section we provide an overview of our model; the full derivation is given in Appendix A.

We assume that the disease dynamics are adequately described by a Susceptible-Infected-Recovered (SIR) compartmental model [16] of a homogeneously mixed population. The equation for the rate of change of disease prevalence is then

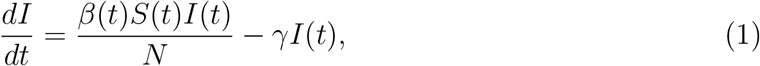

where *β*(*t*) is the (time-dependent) rate of effective contacts, *γ* is the recovery rate from the infectious state, and *N* is the size of the total population. Effective contacts are ones which would transmit disease if they involved an infectious person. As long as a small enough proportion of the population has been infected that *S/N* ≈ 1—as was the case in 2020 for COVID throughout the US—the SIR model solution for the prevalence is

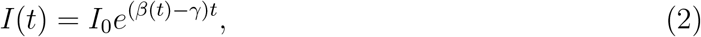

see e.g. [17]. The incidence, i.e. the rate of new cases, is given by the first term on the right-hand side of Equation 1 alone. Substituting, we obtain

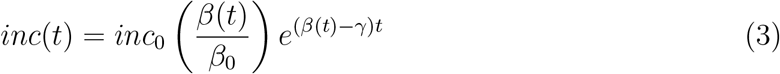

Furthermore, can decompose *β*(*t*) into

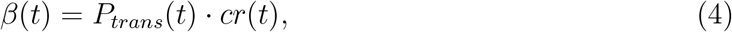

where *cr*(*t*) is the contact rate, while *P*_*trans*_(*t*) is the transmission probability per contact.

Suppose we have time series data of incidence, *inc*_0_, *inc*_1_, …, *inc*_*n*_, and contact rate, *cr*_0_, *cr*_1_, …, *cr*_*n*_ at evenly-spaced times *t*_0_, *t*_1_, …, *t*_*n*_. Suppose further that this time interval is sufficiently short that we can consider *P*_*trans*_ to be approximately constant throughout. With some more manipulation (see Appendix A), we obtain an expression for the incidence at time *t*_*n*_ in terms of the contacts occurring between times *t*_0_ and *t*_*n*_:

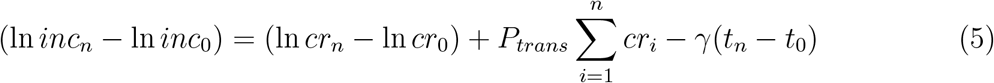

In reality, reporting delays and the incubation and latent periods of the disease will together impose a distribution of delays between the time that transmission occurs and the time that the resulting cases are captured by surveillance. We can account for this by replacing the time series of *cr*_*i*_ with an appropriately lagged version (see Appendix A).

## 3. Data

For incidence, we use the US COVID-19 surveillance data compiled by the New York Times, available at https://github.com/nytimes/covid-19-data, aggregated at the state level.

We obtain contact data from Cuebiq, a vendor of US mobility data sourced from mobile phone users who have opted into sharing location data through a California Consumer Privacy Act (CCPA) compliant process, hereafter referred to as Cuebiq users. Several previous studies have assessed the representativeness of the data by calculating the correlation between the spatial distribution Cuebiq user home locations, and the spatial distribution of the population as captured by US Census data. The studies found high correlations at the census tract level in Washington State [18], the Boston metropolitan area [19] and Philadelphia [20], and U.S.-wide at the county level [21], with Pearson correlation coefficients of 0.91, 0.8, 0.72 and 0.94, respectively.

We make use of the so-called Cuebiq contact index, hereafter CCI. The CCI is a 7-day rolling average of the daily number of encounters that a Cuebiq user has with other Cuebiq users in a given county. An encounter is registered for every instance of two devices occupying the same 50-foot geohash region within the same 5 minute interval; hereafter we refer to this as a CCI encounter. The CCI index is described in more detail on Cuebiq’s website^8,9^.

A CCI encounter thus amounts to a *contact opportunity* rather than an actual contact; in practice only a fraction *f*_*contact*_ of them will consist of two Cuebiq users encountering each other at a small enough separation to be meaningful for disease transmission (e.g. *<* 6 feet for COVID-19). At the same time, only a fraction of the population are Cuebiq users. We thus calculate (Equation B.2) an adjusted index, CCI_100&_, which is the estimated rate of Cuebiq encounters if the entire population were Cuebiq users, under the assumption (see above) that Cuebiq users constitute a representative sample of the population. The relationship between contact rate and CCI is then

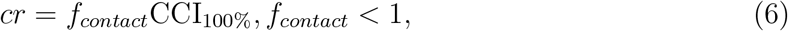

and so we can write Equation 5 as

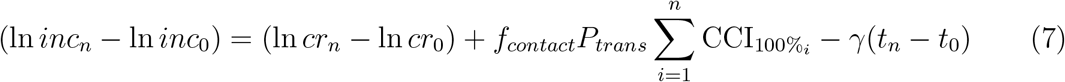

Appendix B describes the details of how a time series of *P*_*CCI*_ is obtained for each state by fitting Equation 5 to COVID-19 case reports.

## 4. Results

Using Equation 5, we first determine the best-fit lag between CCI_100%_ and incidence for each state, as described in Appendix B. Figure 1 shows the time series of CCI_100%_ B.2) together with its lagged version for four example states. Using this lag, we then perform maximum-likelihood fits of the scaled transmission probability (*f*_*contact*_*P*_*trans*_) and initial incidence *inc*_0_ to observed incidence over successive 6-week intervals, again using Equation 5. Results are shown in Figure 2, while Figure 3 shows the model-derived incidence using the maximum-likelihood values, together with the observed incidence. Fits for all 51 states are presented in the Supplementary Material.

**Figure 1.**
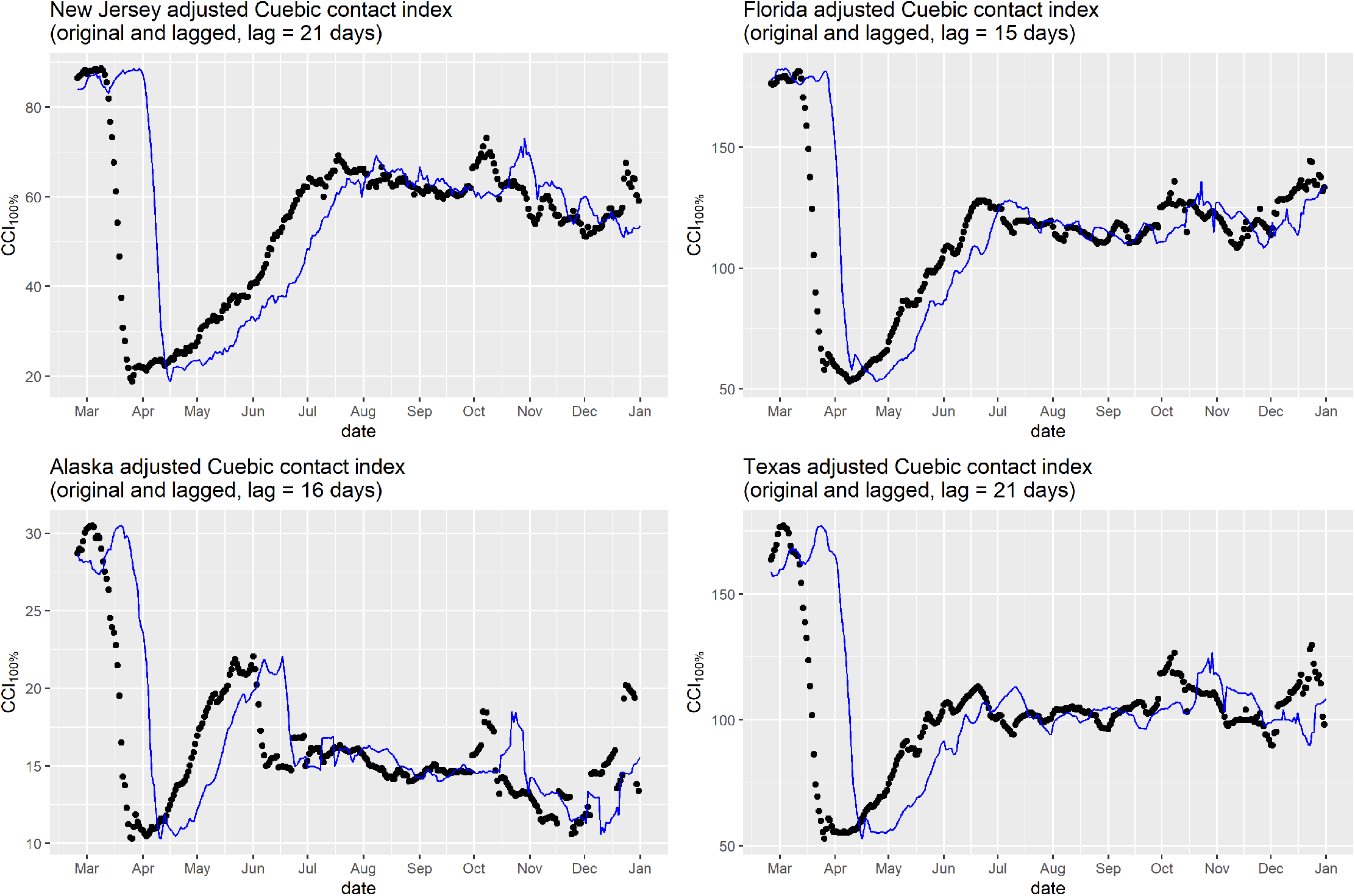
The rescaled seven-day rolling average Cuebiq contact index, CCI_100%_, for four states during 2020 (black points). Also shown is the same data lagged by the best-fit mobility-incidence delay for the given state, obtained as described in Appendix B

**Figure 2.**
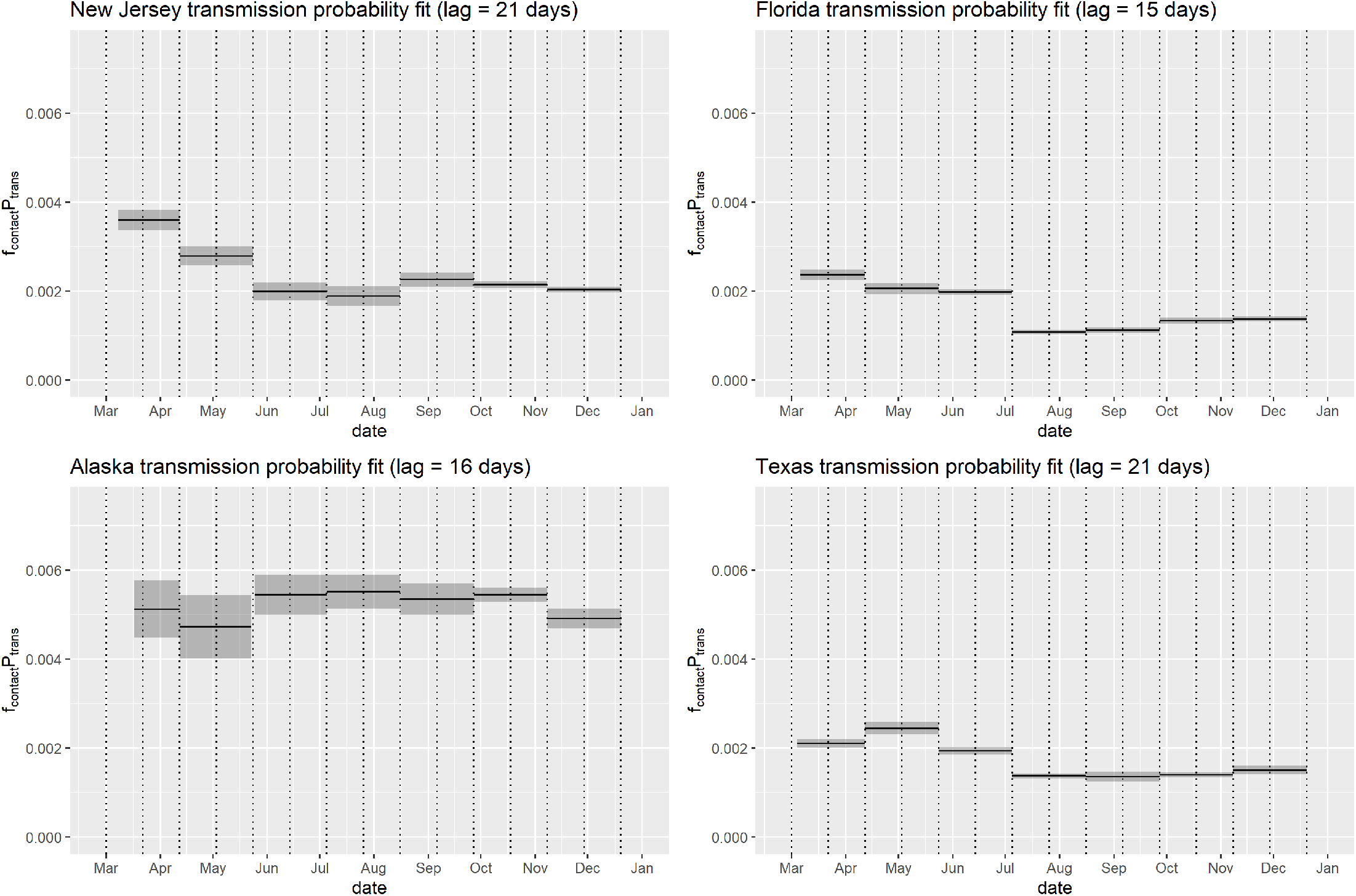
*P*_*CCI*_, the probability of an effective Cuebiq contact (see Equation 6) from the same rolling fits computed computed at six-week intervals (gray dotted lines) for Figure 3. Gray bands denote the 95% confidence interval for each interval.

**Figure 3.**
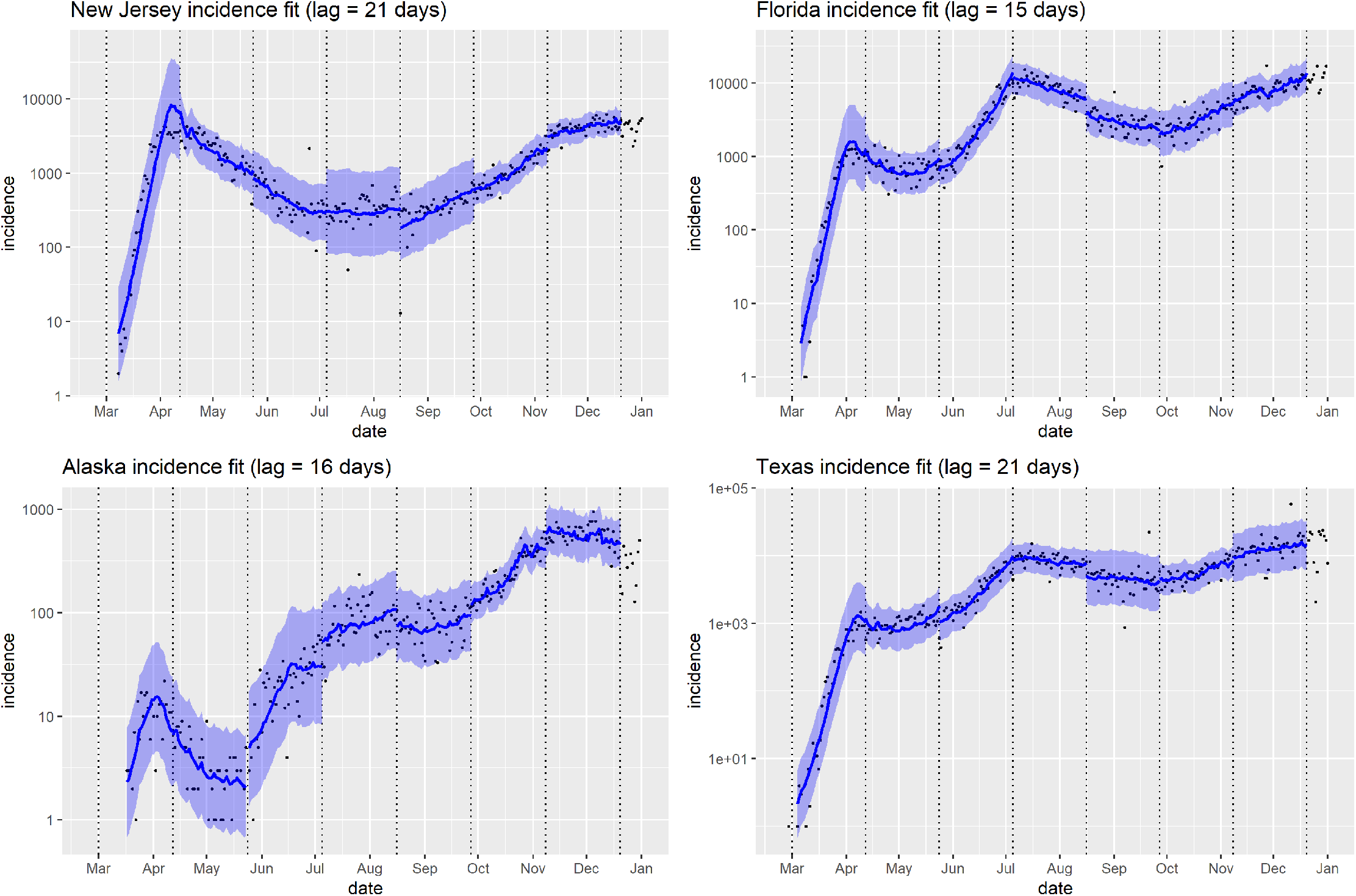
Model incidence (blue curve segments; blue bands show 95% confidence interval) from Equation 5, using the maximum-likelihood fits of *P*_*CCI*_ (see Figure 2) and initial incidence *inc*_0_ to the time series of observed incidence (black dots) and CCI_100%_ (see Figure 1) for four states. Fits are performed over successive six-week intervals (delimited by dotted vertical lines).

Figure 4 shows the distributions across all states of (*f*_*contact*_*P*_*trans*_) averaged over March and April 2020, (*f*_*contact*_*P*_*trans*_)_*early*_, together with the average across the rest of 2020, (*f*_*contact*_*P*_*trans*_)_*RoY*_. The former reflects a largely pre-mask measure of the transmission probability, while for the latter, transmission probability is modified by subsequent widespread yet heterogeneous adoption of masks across US. This figure also shows the distribution of best-fit mobility-transmission lags; the median is 19 days.

**Figure 4.**
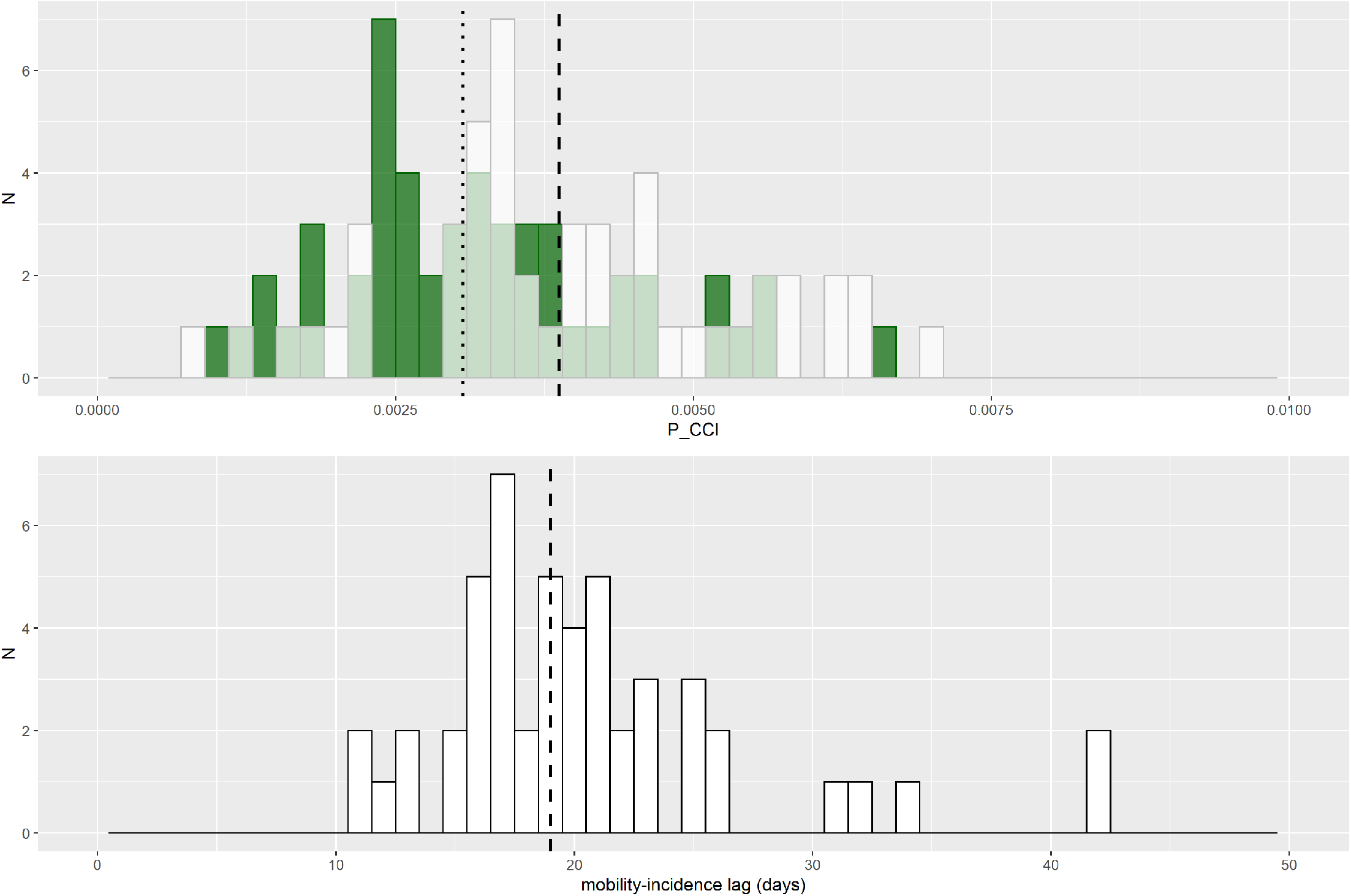
Top: Distribution across all 51 states of early (March 1 to April 30, 2020) average *P*_*CCI*_ (white; dashed line shows median = 0.0039), and rest-of-year (May 1 to December 31, 2020) average *P*_*CCI*_ (green, dotted line shows median = 0.0031). Bottom: distribution across all 51 states of best-fit lag between reported incidence and mobility (dashed line shows median = 19 days).

Figure 5 shows (*f*_*contact*_*P*_*trans*_)_*early*_ versus mean spring temperature for the states (excluding the District of Columbia)^10^ Computing the Pearson product-moment correlation coefficient of the two quantities, we find a moderate, statistically significant negative correlation, *r* = − .57, *N* = 49, *p* = 2 × 10^−5^. That is, colder temperatures tended to be associated with higher transmission probabilities in the initial stage of the pandemic, before differences in mask adoption among the states obscured the picture.

**Figure 5.**
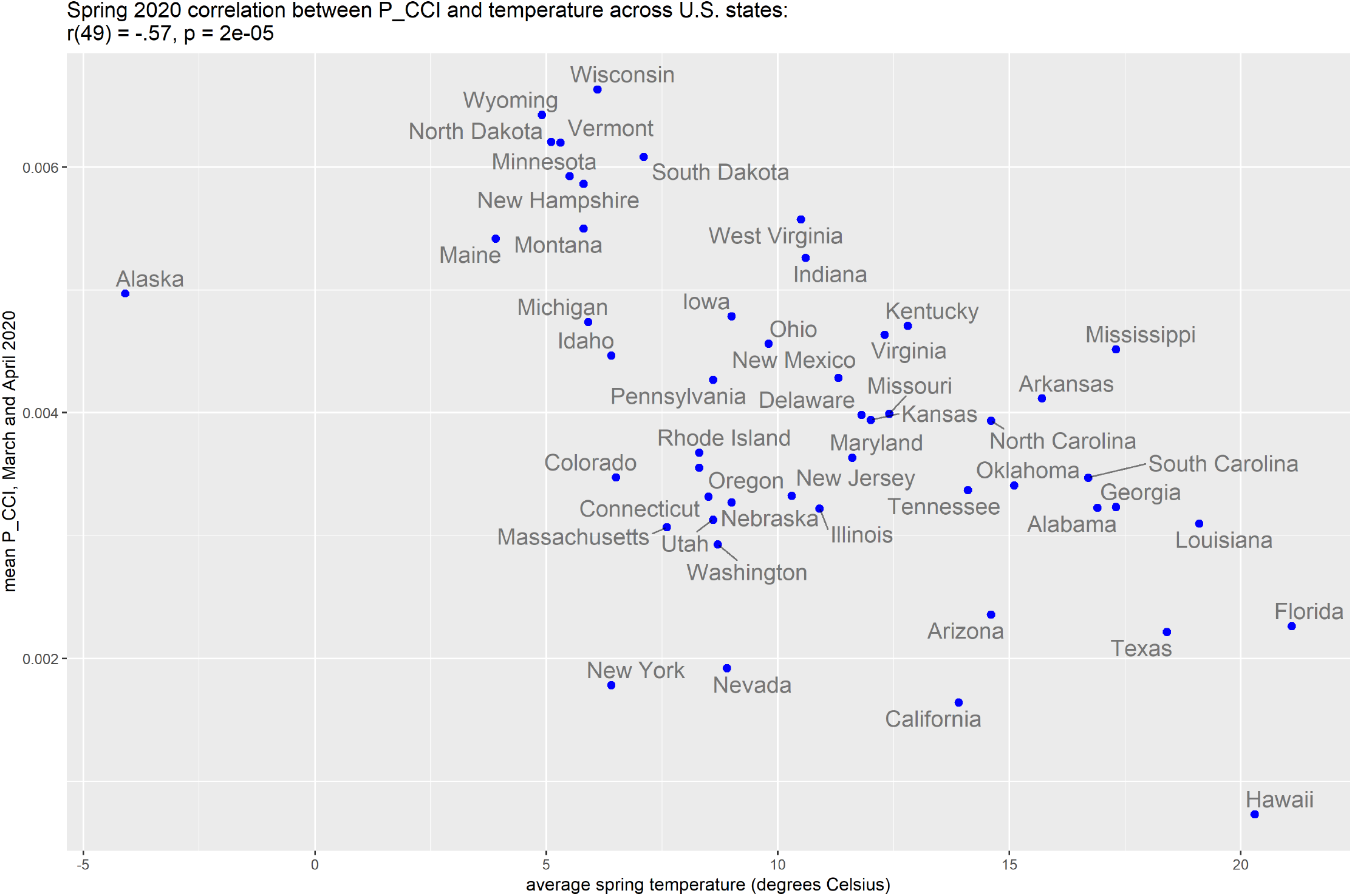
Early (1 March to 30 April) *P*_*CCI*_ versus average winter temperature by state (excluding DC). A moderate, statistically significant negative correlation exists, *r*(49) = − .57, *p* = 2 × 10^−5^, i.e. lower temperatures tend to be associated with higher *P*_*CCI*_.

Figure 6 shows a simple demonstration of how the model developed here could be used for short-range forecasting, using Florida as an example. Additional examples are shown in the Supplementary Material. To perform a forecast starting from a given date *T* forward, we first fit our model to the previous six weeks, [*T* − 6*w, T*], of incidence data and lagged CCI_100%_ data to obtain a maximum-likelihood estimate of (*f*_*contact*_*P*_*trans*_). The best-fit lag between incidence and CCI is 15 days for Florida. This means that at time *T*, we still have lagged CCI data up to date *T* + 15*d*. Using this data, together with the estimate of (*f*_*contact*_*P*_*trans*_), we are thus able to run the model 15 days into the future. We retrospectively perform forecasts from dates *T*_1_ = 1 April 2020, *T*_2_ = 1 June, *T*_3_ =1 July and *T*_4_ = 25 August, each date chosen to come just before a turnover in incidence from growth to decay or vice versa. The quality of each forecast can be visually assessed by comparing it to the actual incidence of cases reported over the forecast horizon.

**Figure 6.**
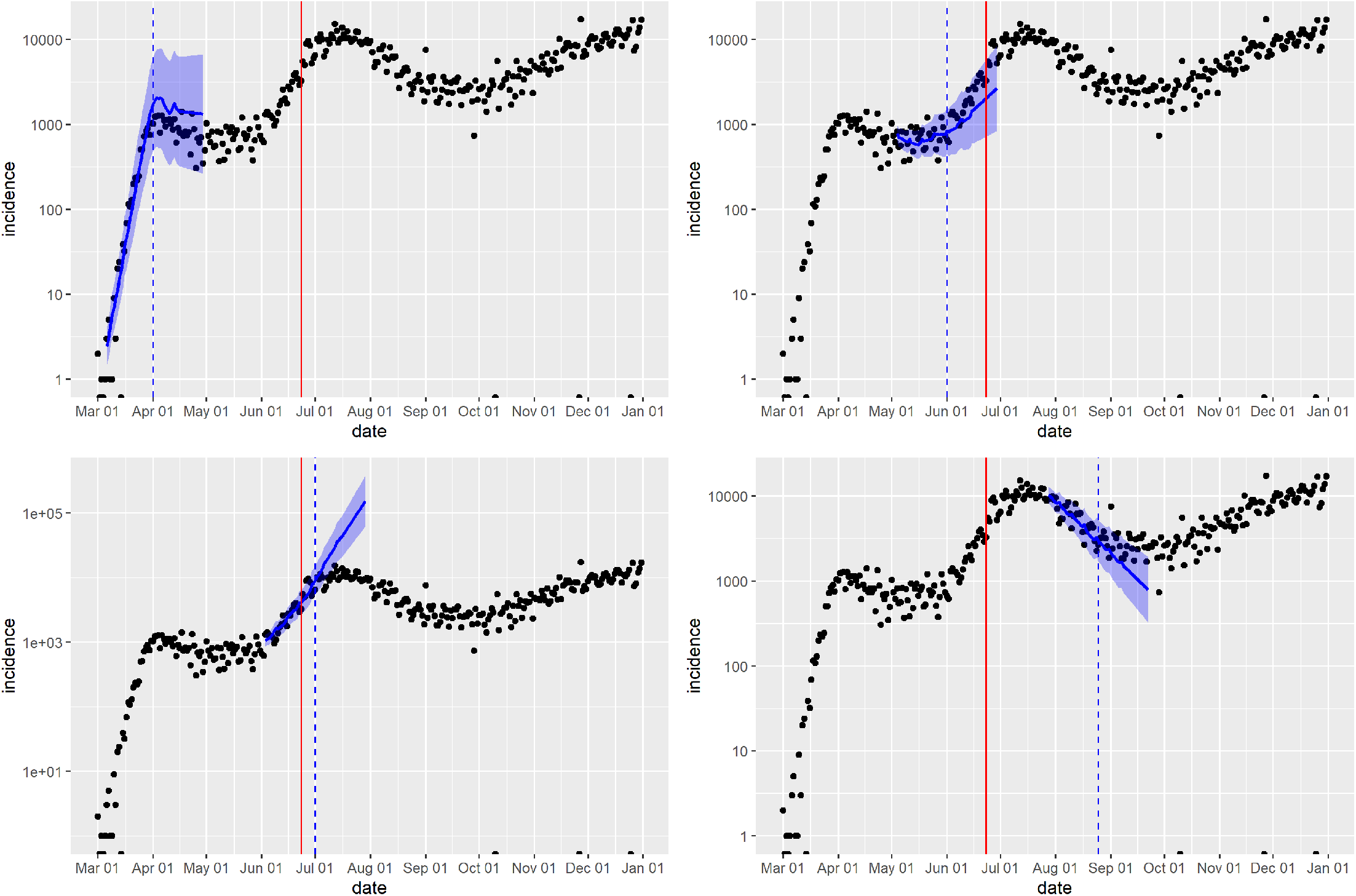
Illustration of the application of our fitting methodology to short-range forecasting, using the state of Florida as an example. Forecasts are performed at different times (dashed vertical lines), each time using a fitting time window of the previous six weeks to estimate *P*_*CCI*_. The forecast time horizon is equal to the mobility-transmission lag, which for Florida is estimated as 15 days. Also shown for comparison is 23 June (solid vertical line), the date on which widespread mask mandates started coming into effect in Florida, starting with Miami-Dade County.

## 5. Discussion

Across the 51 states, the scaled transmission probability averaged over the first two months of the pandemic (March and April 2020), (*f*_*contact*_*P*_*trans*_)_*early*_, has a median value of 0.0039. The transmission probability across the rest of the year, (*f*_*contact*_*P*_*trans*_)_*RoY*_, has a median value of 0.0031, about 20% lower (Figure 4, top). A marked decrease in transmission probability is consistent with the overall increasing level of NPI adoption, principally maskwearing, as the year progressed. Seasonality may also play a role. However, looking at the individual state time series of (*f*_*contact*_*P*_*trans*_) (see Figure 2 for four examples, and the Supplementary Material for the remaining 47 states) reveals significant heterogeneity, with some states (e.g. New Jersey, Florida) showing a clear reduction in *P*_*CCI*_ after spring of 2020, yet others (e.g. Alaska) showing no clear time trend. This may be reflective of the heterogeneity in the practice of NPIs that reduce per-contact transmission probability, primarily the adoption level of masks and the level of compliance with social distancing (which, since it occurs on a scale of a few meters, is not resolved in Cuebiq’s mobility data). Given the evidence of temperature dependence in COVID transmissibility, it may also be reflective of heterogeneity in seasonal weather patterns among states.

Across all states, the median best-fit time lag between CCI_100%_ and observed incidence is 19 days, though here, again, there is significant heterogeneity (Figure 4, bottom). The time between infection and case report is the sum of the incubation period of a disease, the diagnostic delay and the reporting delay. In the hypothetical case of instantaneous diagnosis and reporting, the delay would be due to the incubation period alone. Thus the smallest lag we observe, 11 days, constitutes an upper limit to the median incubation period. Meta-analyses have variously reported a mean COVID-19 incubation period of 6.5 (95% CI: 5.9–7.1) days[22], 5.8 (95% CI: 5.0-6.7) days[23], 5.6 (95% CI: 5.2–6.0) days or 6.7 (95% CI: 6.0–7.4) days[24], 5.74 (95% CI: 5.18-6.30) days[25], and 6.2 (95% CI 5.4, 7.0) days[26], all of which fall below 11 days. One source of variability may be heterogeneity in state-level reporting practices. Also, since the latent period is determined by in-host interaction, it may vary systematically by population characteristics (age distribution, comorbidity profile etc.), which may also contribute to the heterogeneity.

Multiple studies have reported associations between colder temperatures and various metrics of COVID burden (see Introduction). Our results suggest, specifically, an association between temperature and per-contact transmission probability. Although prior studies have found evidence that the COVID-19 virus half-life is reduced at higher temperatures ([27], [28], [29]), it is important to note that our results do not by themselves imply that this particular mechanism is responsible. Behavior could also contribute: People in warmer climates tend to spend a larger proportion of their time outdoors, thus a larger proportion of daily contacts will occur outdoors in, for example, springtime California versus springtime Alaska. And there is strong evidence to suggest that the outdoor risk of COVID transmission is substantially lower than the indoor risk (see [30] for a systematic review). Both these causal pathways, and more, could be operating together.

In the forecasting demonstration shown in Figure 6, the sharp downturn in Florida incidence just after 1 April 2020 is reasonably well predicted, as is the return to incidence growth after 1 June. However, neither the downturn after 1 July nor the upturn beginning in late August are predicted. Indeed, Florida’s CCI_100%_ (Figure 1) varies much less after the beginning of July than it does before. However, widespread mask mandates started coming into effect in Florida on 23 June; with a 15-day lag (i.e. 8 July) this is close to Florida’s second incidence peak. This suggests that at later time, variations in mask use, rather than in contact rate, may have played the dominant role in driving changes in transmission. We leave to future work a more sophisticated forecast model that incorporates mask use, where such data is available.

The work presented here is subject to a number of limitations: i) Both our model and the data we use lack any stratification, thus any effects arising from heterogeneous demography, health status etc. within a given state are not accounted for. ii) Though previous studies all found high correlation between the geographic distribution of Cuebiq users and that of the population as a whole, this does not fully guarantee the represenativeness of Cuebiq users. iii) In comparing states to each other, we have made the assumption that the scaling (Equation B.1) between true contact rate and adjusted Cuebiq contact index is the same across all states. iv) We have assumed that within a given state, the lag between mobility and incidence, which we estimate using only the first four months of the pandemic, remains constant. v) We have argued that transmission probability changes more slowly over time than mobility, and thus approximated *P*_*trans*_ as constant within successive 6-week periods. However, this approximation may not always hold well, in particular when a change in mask mandates falls within a given period. Also, during the phase of gradual relaxation after the initial lock-downs, mask use may have increased at a similar rate to mobility as businesses, public spaces etc. re-opened while at the same time requiring masking. vi) In our forecasting experiment, we have unfairly granted ourselves fore-knowledge of the state’s mobility-incidence lag, which was actually fit using the first four months of data. In practice, in the very beginning of a pandemic we would have to resort to using a range of plausible lags, the lower bound being the mean incubation period of the disease.

## 6. conclusion

Using a mobility index that can be considered a direct proxy of contact rate has allowed us to construct a fully mechanistic model that derives disease incidence from this data. As a result, we have been able to get direct insight into the variability of per-contact COVID-19 transmission in the U.S. both by state and by date. Our findings are consistent with associations between colder temperatures and stronger COVID-19 burden reported in previous studies, and suggest that it is specifically changes in the per-contact transmission probability which play a role. As a forecast tool, the model would have performed best before NPIs that modified per-contact transmission probability—principally masks—came into widespread use. To lift this limitation, future development should also incorporate time series data of NPI use. Our methodology is also readily extensible to other respiratory diseases such as influenza or RSV, contingent on the availability of good-quality surveillance data. Indeed, in a non-pandemic setting forecasting will be aided by the (likely) absence of NPIs, and by mobility following more predictable seasonal patterns rather than being driven by reaction to epidemiology. The availability of mobility data that even more directly probes person-to-person contacts, e.g. through Bluetooth proximity detection of the sort used in COVID exposure-notification apps, would also benefit the performance of this model.

## Supporting information

Supplementary Material: Retrospective forecasts for all states

Supplementary Material: Fits for all states

## Data Availability

All data produced in the present study are available upon reasonable request to the authors

## Contributors

All authors were involved in the conception and design of the study. EWT, ZM and MGC developed the methodology, with input from all other authors. CM acquired the funding to purchase the commercial (Cuebiq) data used. EWT and MGC accessed, verified and collected the data. EWT, ZM and MGC contributed to the analysis, including the development of the software used therein. EWT wrote the original draft. All authors critically reviewed and edited the manuscript for scientific content. All authors have access to the data and software used, and are thus able to validate the analysis.

## Appendix A: The model in detail

In an SIR model, the equation for rate of change of disease prevalence is

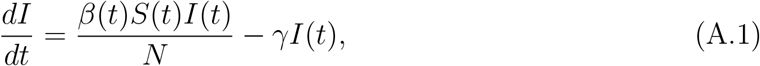

where *β*(*t*) is the time-dependent average number of effective contacts per person per unit time, *γ* is the recovery rate from the infectious state, and *N* is the size of the total population. Effective contacts are ones which would transmit disease if they involved an infectious person. As long as only a small fraction of the total population has become infected—as was the case in the US and most of the world throughout 2022—*S/N* ≈ 1, and the SIR model solution for the prevalence is

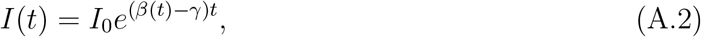

see e.g. [17]. The incidence, i.e. the rate of change of cumulative cases *C*(*t*), is given by the first term on the right-hand side of Equation A.1 alone:

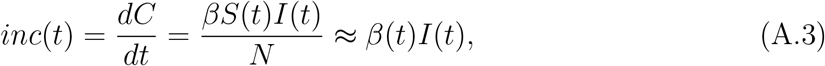

where the approximate equality holds when, again, *S/N* ≈ 1. Substituting, we obtain

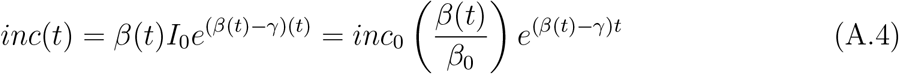

where

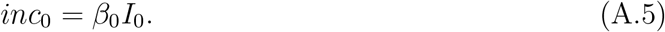

Since for an SIR model the instantaneous effective reproduction number is

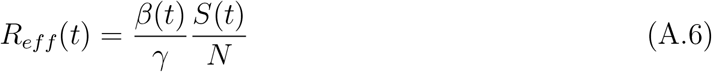

thus we can also write the incidence in terms of the reproduction number:

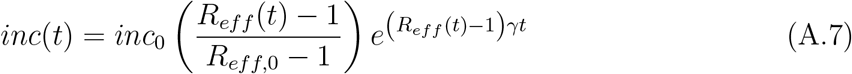

Taking the log of Equation A.4, we have

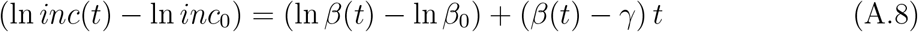

Considering now an infinitesimally small time interval, *dt*, this becomes

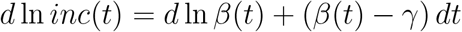

or

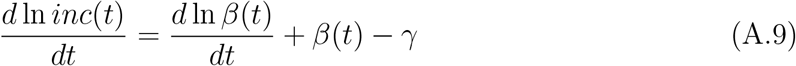

Integrating with respect to *t*, we obtain, over a time interval [*t*_0_, *t*],

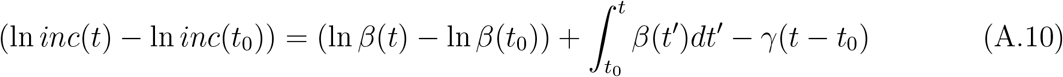

We can decompose *β*(*t*) into

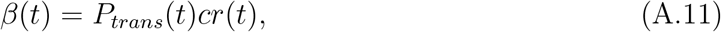

where *cr*(*t*) is the contact rate, while *P*_*trans*_(*t*) is the transmission probability per contact. Note that we can then express the reproduction number as:

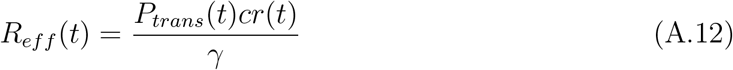

In general, both the contact rate and the transmission probability change over time, the latter due to changes in the practice of non-pharmaceutical interventions (NPIs) such as mask-wearing, as well as changes intrinsic to the disease, e.g. emergence of new variants. Since widespread changes in NPIs and in the relative distribution of variants are usually gradual, whereas contact patterns can change significantly from one day to the next (for example between a weekday and the weekend, or as the result of a mass gathering event), we expect *P*_*trans*_(*t*) to generally vary more slowly than *cr*(*t*). If *P*_*trans*_ can be considered constant over the time interval [*t*_0_, *t*], then Equation A.10 becomes

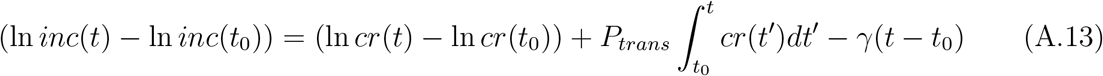

Suppose we have time series data of incidence and contact rate reported with constant time interval *δt*, so that *cr*_*i*_ and *inc*_*i*_ are the contacts per person and the total number of new cases, respectively, occurring within the time interval *t*_*i*−1_ *< t* ≤ *t*_*i*_. We can then apply Equation A.13 in discrete form to obtain the change in incidence between a time *t*_0_ and time *t*_*n*_ in terms of the contacts occurring during this time:

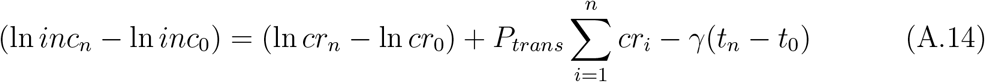

In practice, disease incidence captured by surveillance will be subject to under-reporting, i.e. the reported incidence is

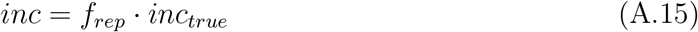

where *inc*_*true*_ is the true underlying incidence, and *f*_*rep*_ is the fraction of cases reported. If *f*_*rep*_ can be considered constant over the time interval [*t*_*a*_, *t*_*b*_], then if we now replace the reported incidence with the true incidence in Equation A.14, the left-hand side becomes

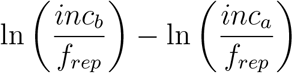

or

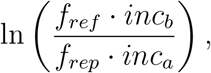

thus *f*_*rep*_ cancels out and we recover the original equation. Therefore, when considering time intervals over which the degree of under-reporting can be considered constant, the relationship described by Equation A.13 is independent of under-reporting.

A simplification we have made thus far is to assume that there is no lag between contacts and their effect on reported incidence. In reality, reporting delays and the incubation and latent periods of the disease will together impose a distribution of delays between the time that transmission occurs and the time that the resulting cases are captured by surveillance. If cases reported at time *t*_*i*_ depend on contacts occurring between times *t*_*i*−*q*_ and *t*_*i*−*p*_, with *q > p*, then we can account for this by replacing *cr*_*i*_ with an appropriately lagged version,

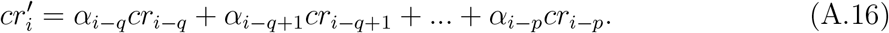

## Appendix B: Fitting the model to data

The CCI of a given region is the daily number of instances of two Cuebiq users occupying the same 50ft x 50ft geohash grid cell within the same 5 minute interval, divided by the total number of Cuebiq users within that region. From this, we want to estimate the rate of encounters between Cuebiq users at distances ≤ *d*_*trans*_, where *d*_*trans*_ is the maximum distance for potentially disease-transmitting contacts. Assuming an average movement speed *v*, the time to pass through a *d*_*trans*_ × *d*_*trans*_ cell is approximately *d*_*trans*_*/v*. The proportionality constant between CCI and the contact rate within distance *d*_*trans*_ between Cuebiq users is given by the ratio of their associated space-time volumes. And, assuming that *v* is approximately constant, the contact rate is linearly proportional to CCI:

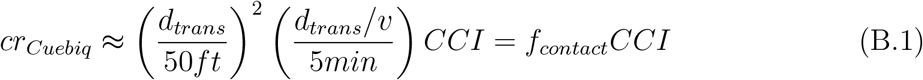

Since only a fraction of the total population are Cuebiq users, the CCI only captures a fraction of the total contacts experienced by a person per day. In order to estimate CCI_100%_, the hypothetical CCI which would be measured if the entire population were Cuebiq users, we additionally obtain from Cuebiq the time series of total number of Cuebiq user devices seen on day *i* across a given region, 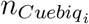. Insofar as the Cuebiq users can be considered a representative sample of the general population, we can then estimate CCI_100%_ on day *i* across a given region by rescaling the CCI as follows:

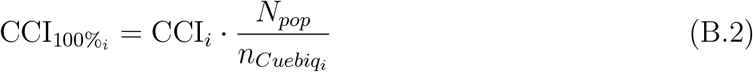

where *N*_*pop*_ is the population size of the region. Our estimate for the total contact rate is then

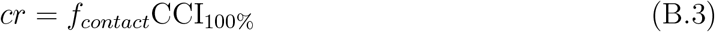

We can then write Equation 5 as

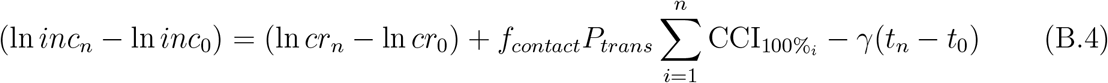

where *P*_*CCI*_ = *f*_*contact*_*P*_*trans*_ is the transmission probability per Cuebiq encounter.

We conduct our analysis at the state level, and thus aggregate incidence and Cuebiq data (both of which are provided at the county level) accordingly. In order to calculate the lagged version of the time series of 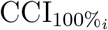 as per Equation A.16, we make the simplifying assumption that *q* = *p* + 7. Given that the CCI is already computed as a 7-day rolling average, we then only need to find *p* for each state. To do so, we perform a two-step optimization. First, we select a time window *t*_*j*_ < *t* ≤ *t*_*k*_ within the available data. We then lag the time series of 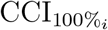 by values of *L* = 0, 1, 2, …, 50. For each value of *L*, we use the R optimization function `optim()` to find the value of *P*_*CCI*_ which produces the best fit of Equation B.4 to the observed incidence over the time window, in the sense of minimizing the negative log likelihood (NLL).

We take as *t*_*j*_ the day on which cumulative cases first reached or exceeded 10 in a given state. We repeat the above fitting procedure with *t*_*k*_ = 1 May 2020, *t*_*k*_ = 1 June 2020, and *t*_*k*_ = 1 July 2020, each time computing a best-fit lag. We then take the average of these three as the best-fit lag *p* for the given state. Using *p*, we compute the lagged CCI time series for the state as per Equation A.16, which we then use to fit Equation B.4 to reported incidence.

We do so over successive 6-week time intervals, going from *t*_*j*_ until the end of 2020, thus obtaining a best-fit *P*_*CCI*_ for each interval.

## Footnotes

1 https://covid19.who.int/

2 The original report published by the Collaborating Centre for Infectious Disease Modelling and collaborators: https://www.imperial.ac.uk/mrc-global-infectious-disease-analysis/covid-19/report-12-global-impact-covid-19/

3 https://www.google.com/covid19/mobility/

4 https://covid19.apple.com/mobility

5 https://dataforgood.facebook.com/dfg/covid-19

6 https://www.cuebiq.com/

7 https://www.safegraph.com/

8 https://www.cuebiq.com/visitation-insights-contact-index/

9 https://help.cuebiq.com/hc/en-us/articles/360041285051-Mobility-Insights-Mobility-Index-CMI

10 Data taken from https://www.currentresults.com/Weather/US/average-state-weather.php

