## Supplementary figures and images for "U.S. state-level COVID-19 transmission insights from a mechanistic mobility-incidence model"

### Alabama_Peff_plot.png

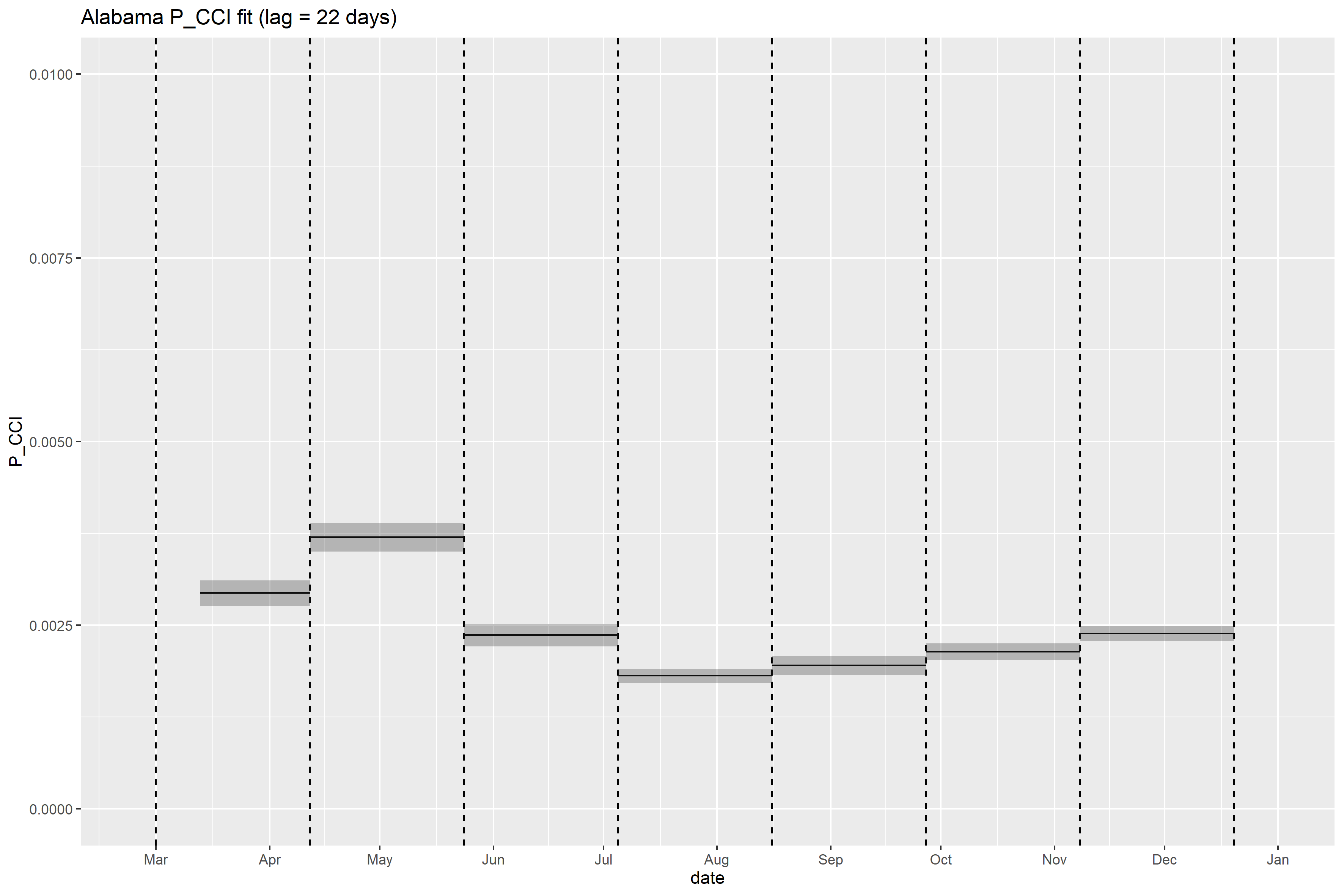

### Alabama_retrospective_forecasts.png

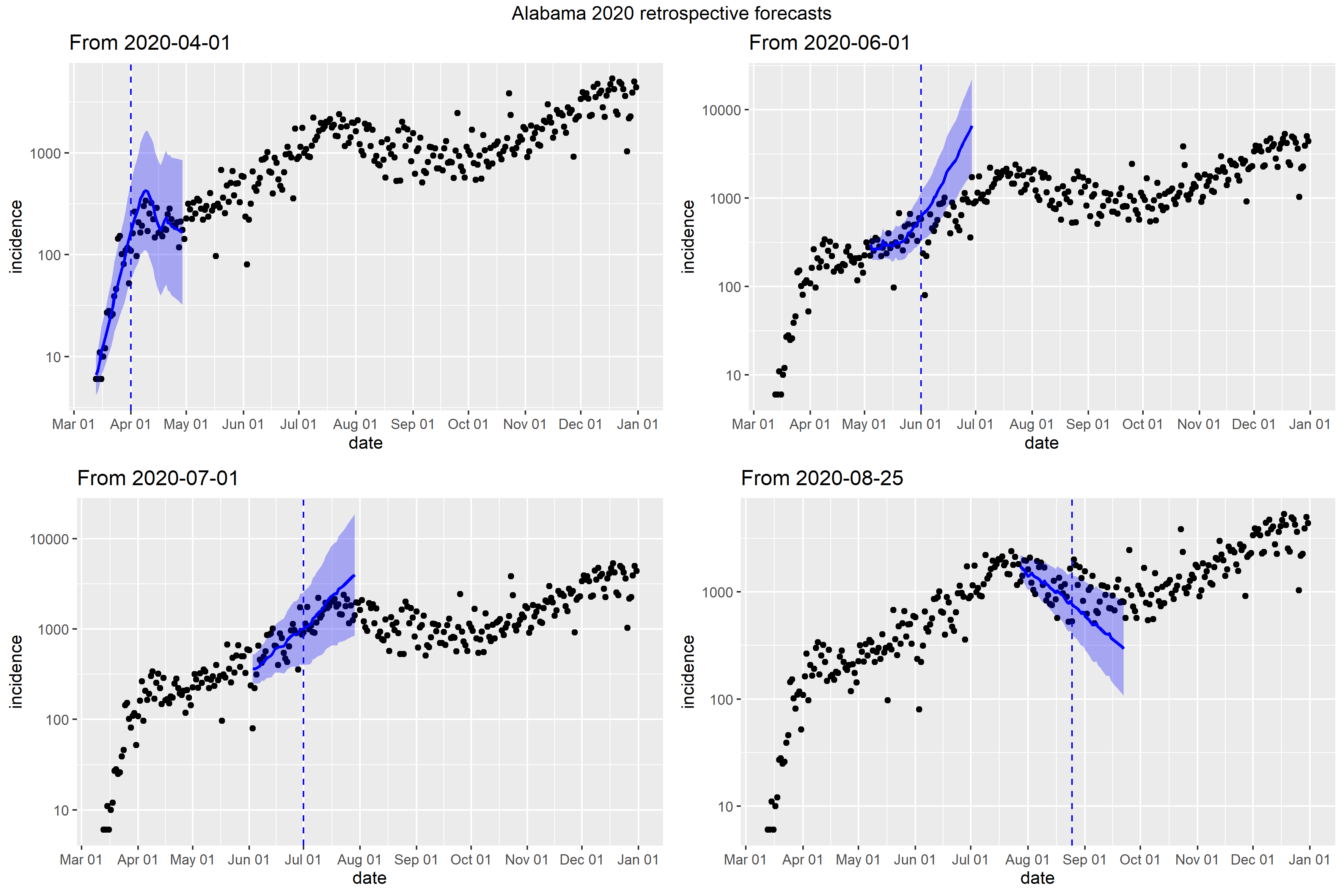

### Alaska_full_fit_plot.png

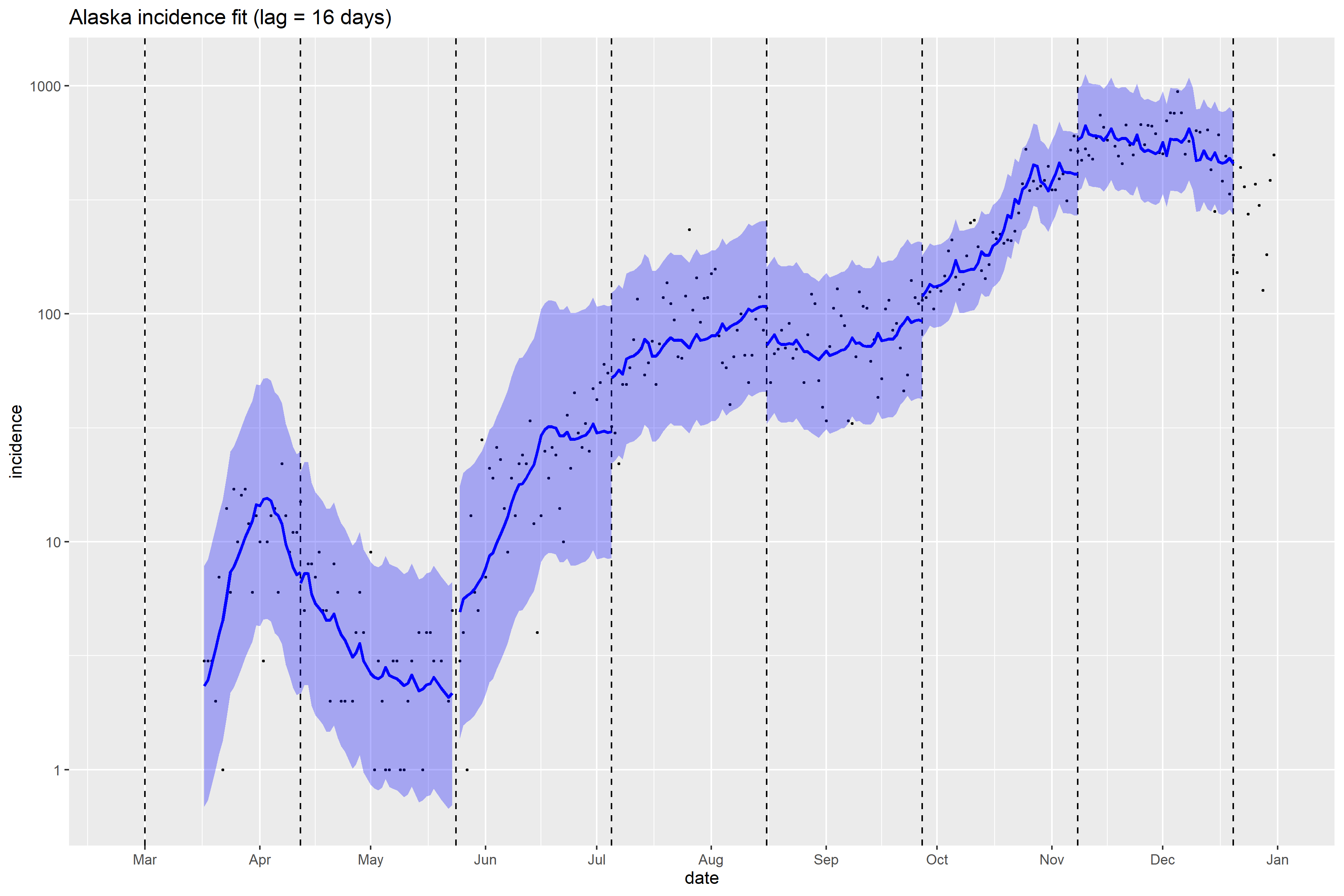

### Alaska_Peff_plot.png

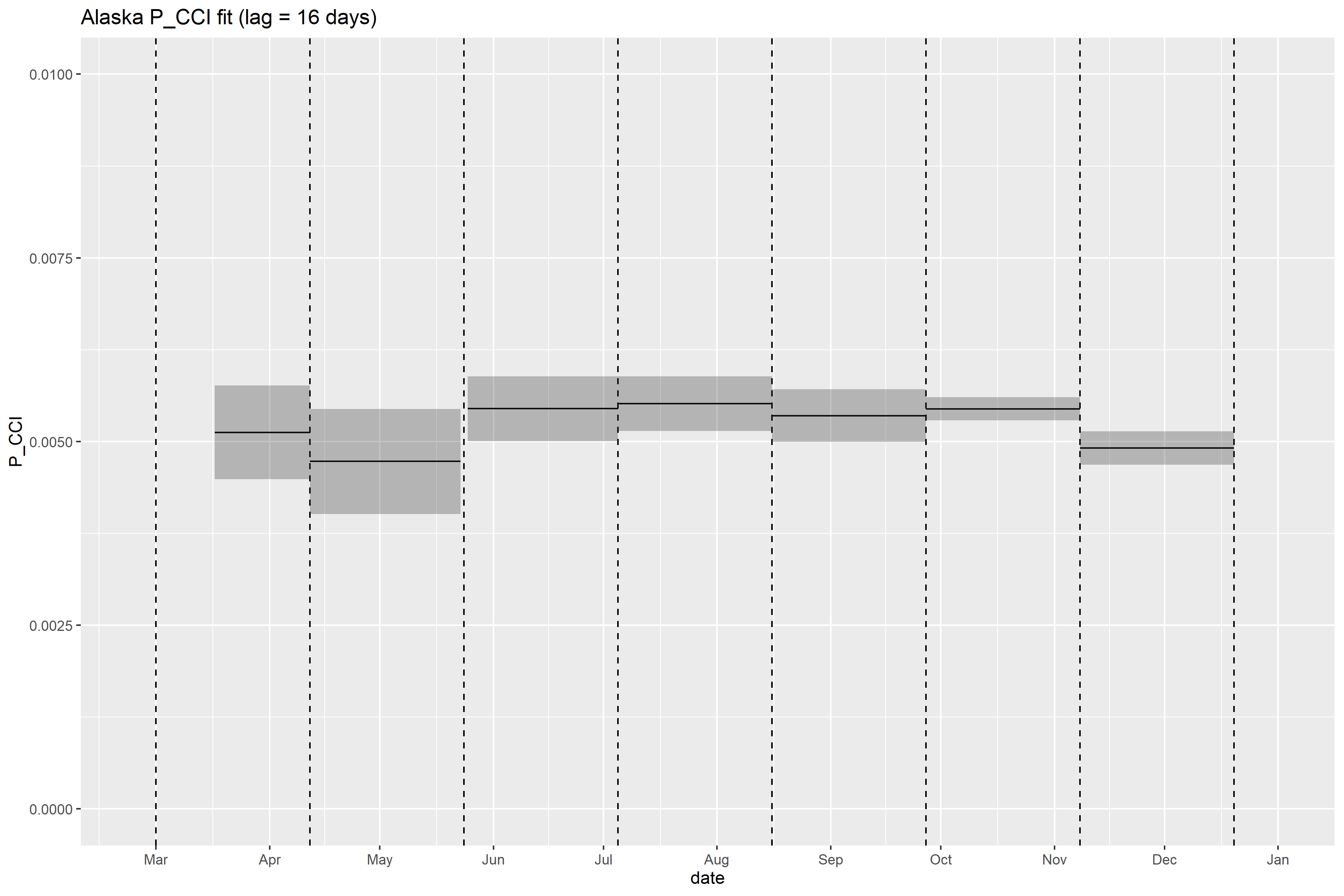

### Alaska_retrospective_forecasts.png

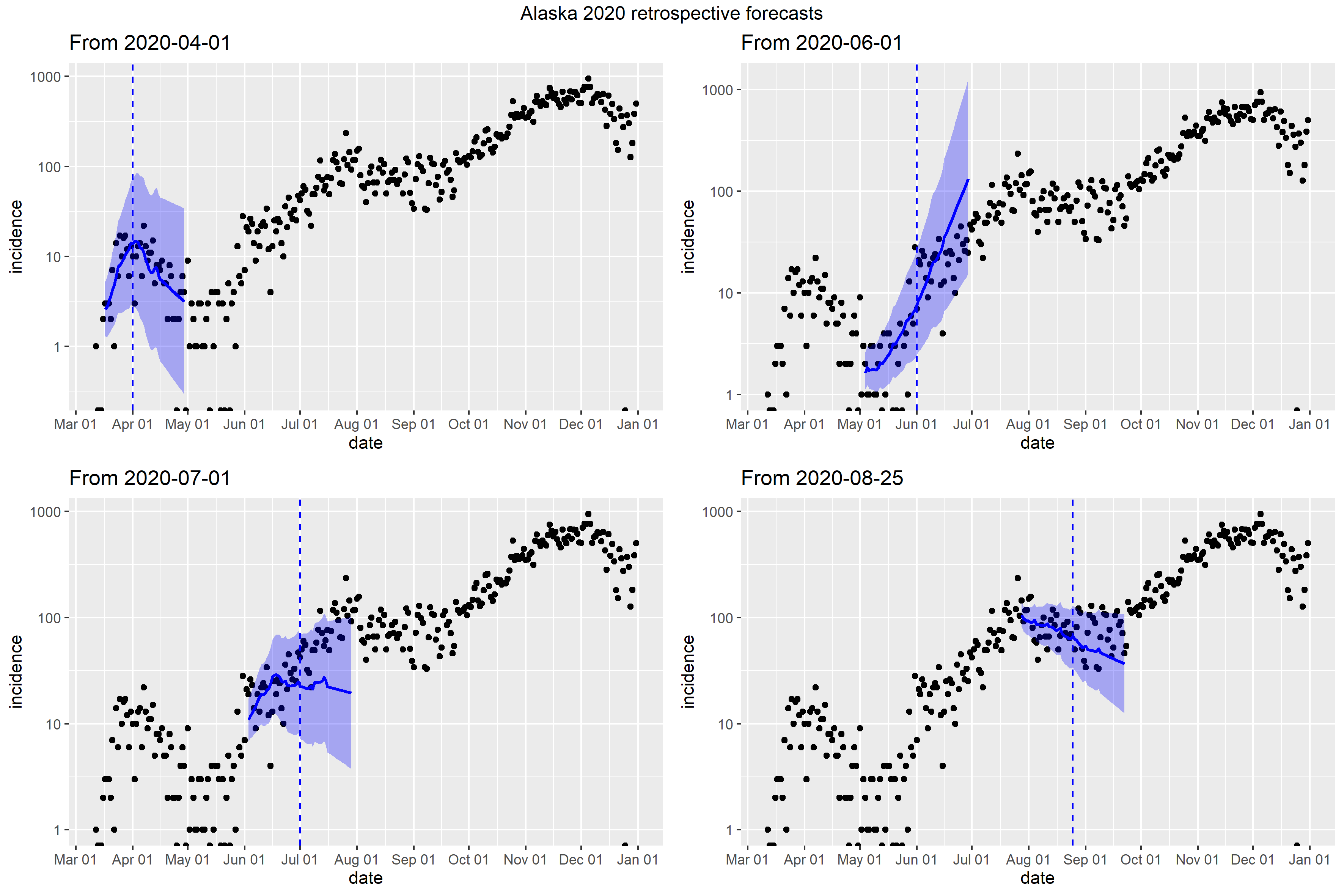

### Arizona_Peff_plot.png

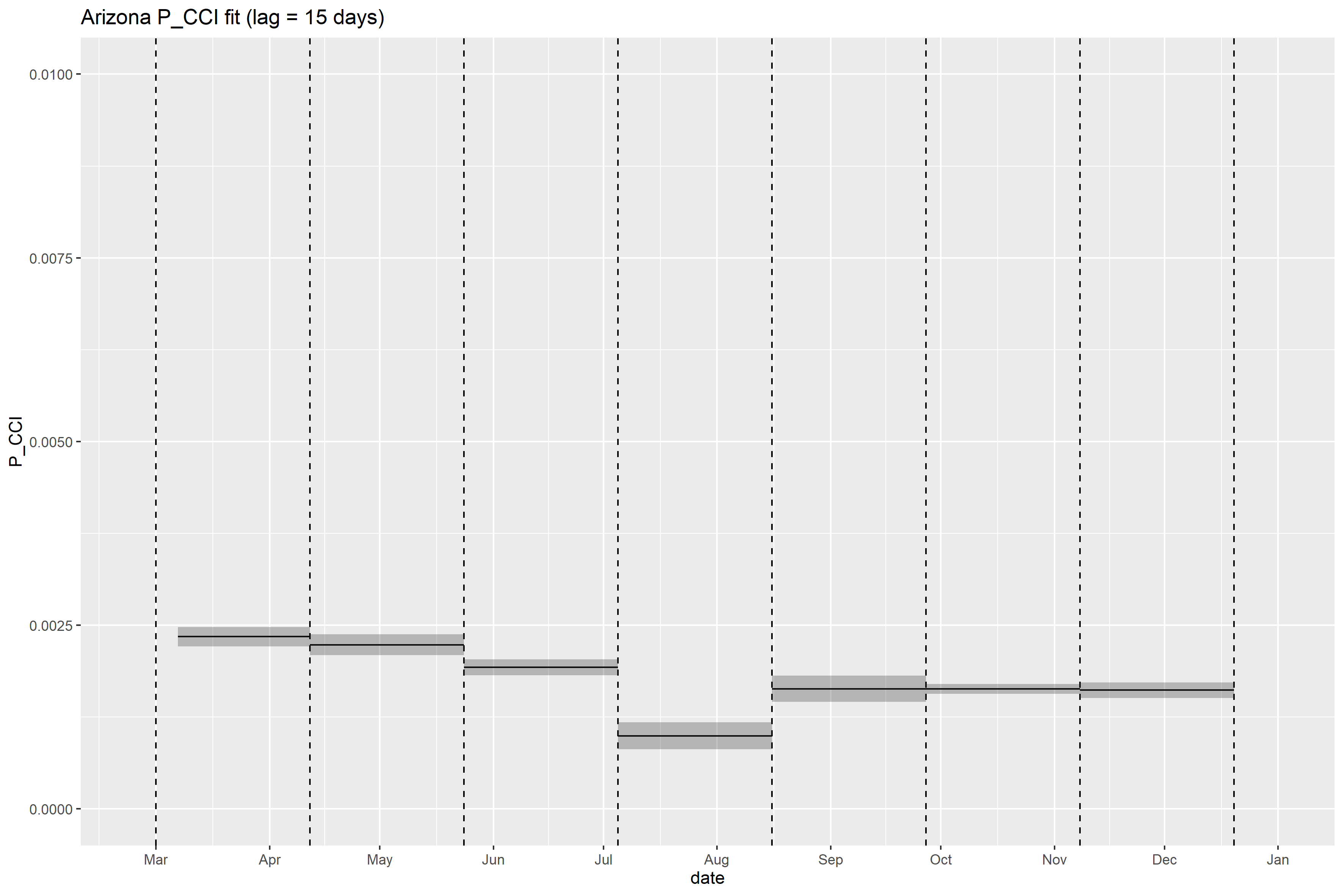

### Arizona_retrospective_forecasts.png

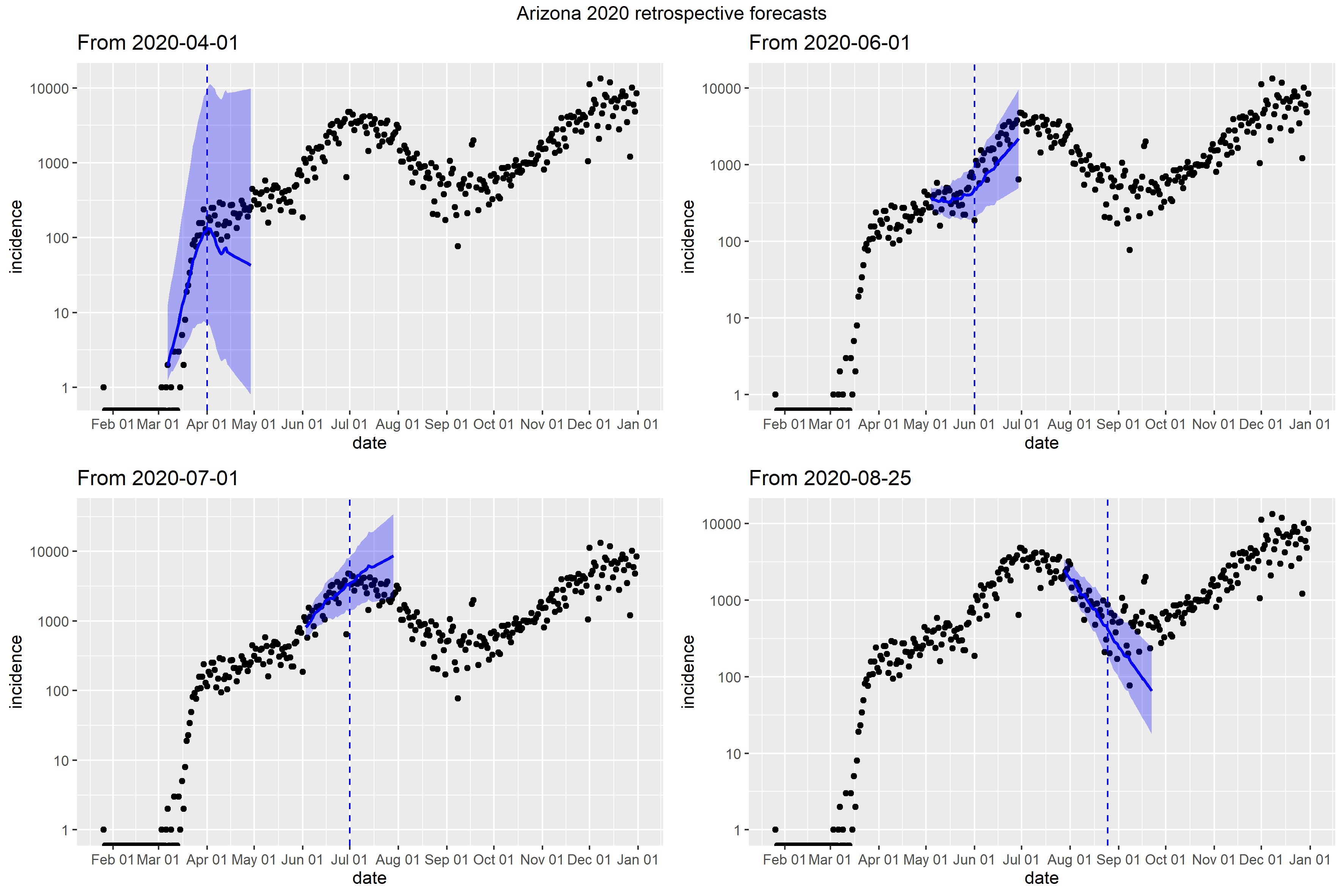

### Arkansas_Peff_plot.png

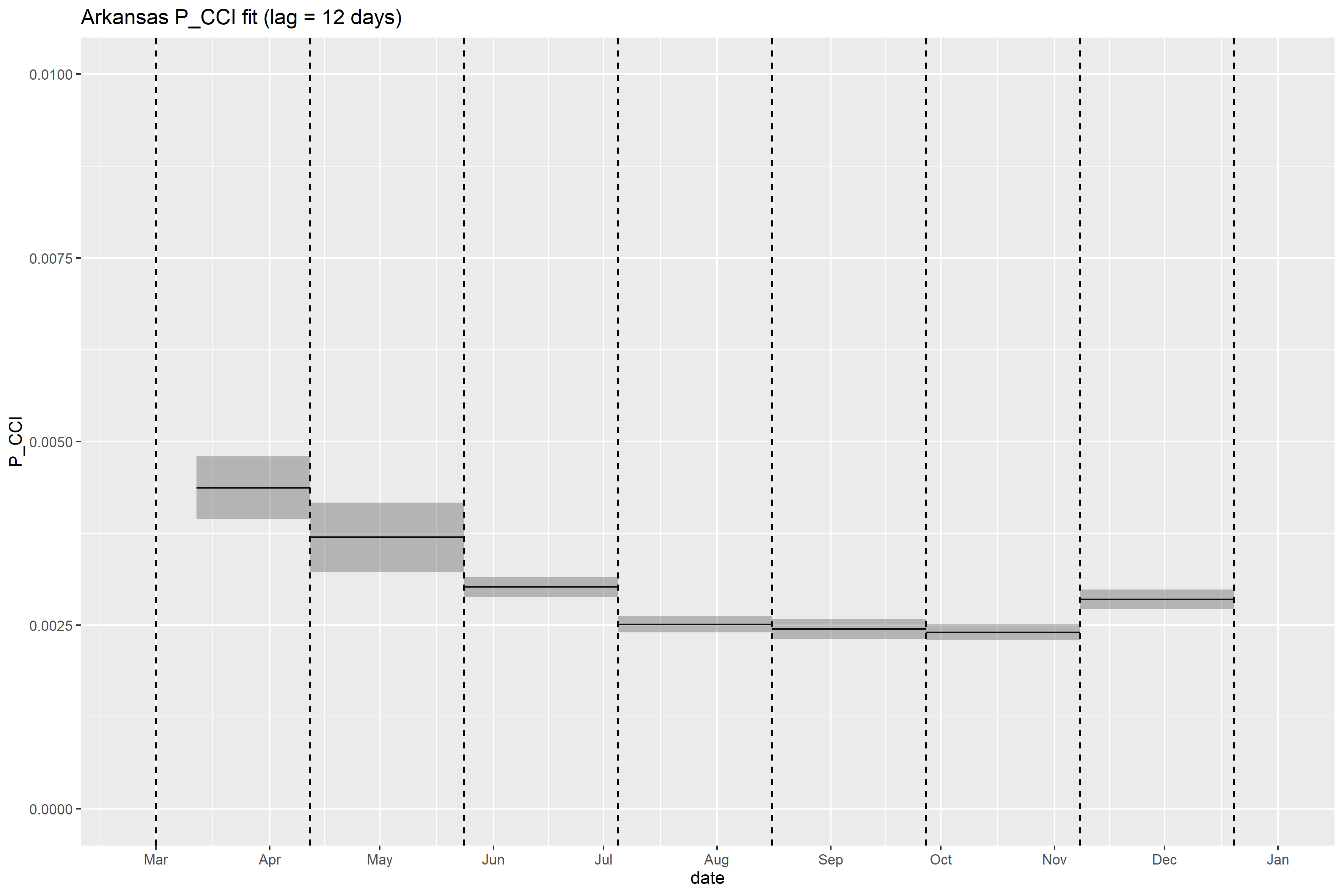

### Arkansas_retrospective_forecasts.png

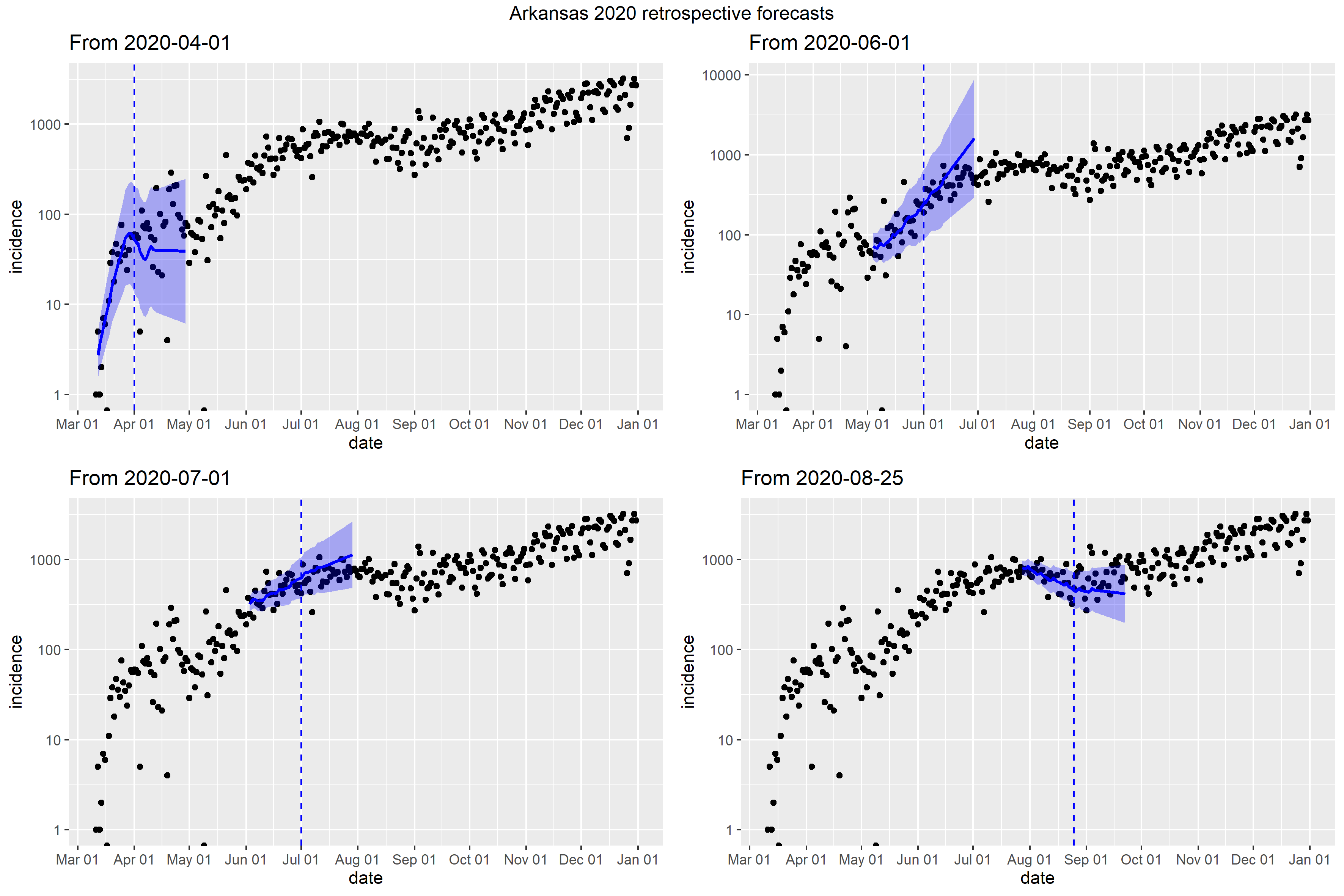

### California_Peff_plot.png

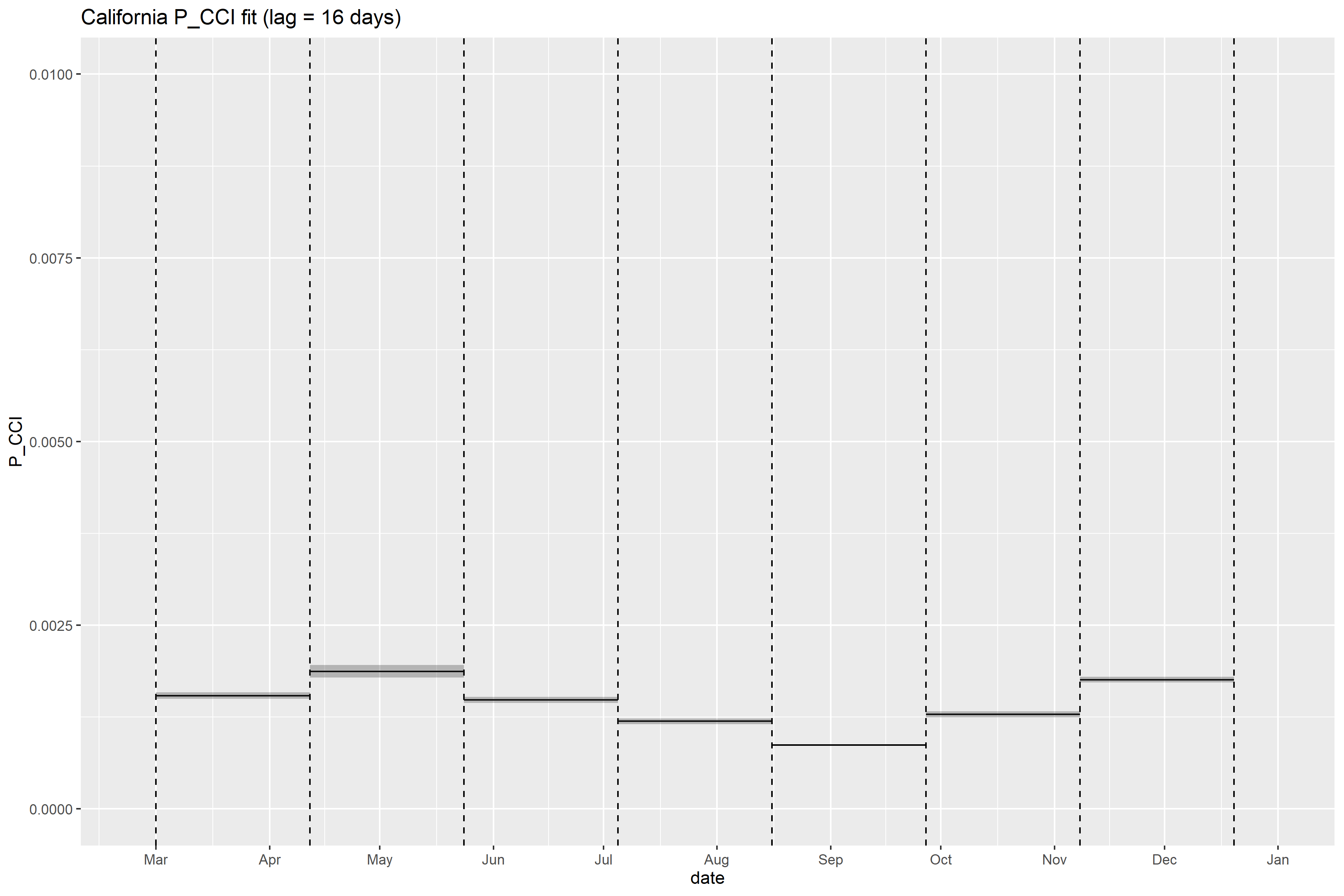

### California_retrospective_forecasts.png

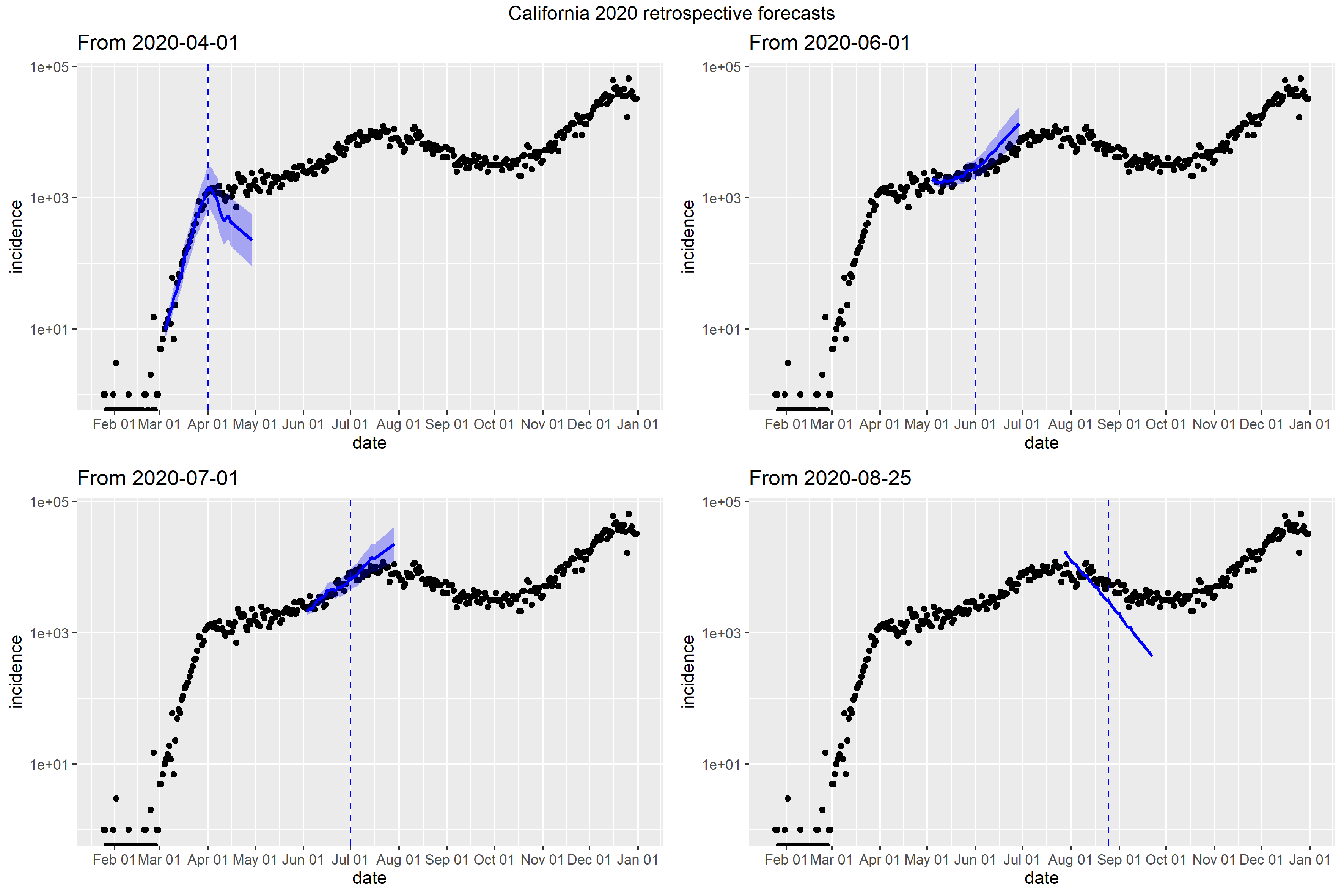

### Colorado_Peff_plot.png

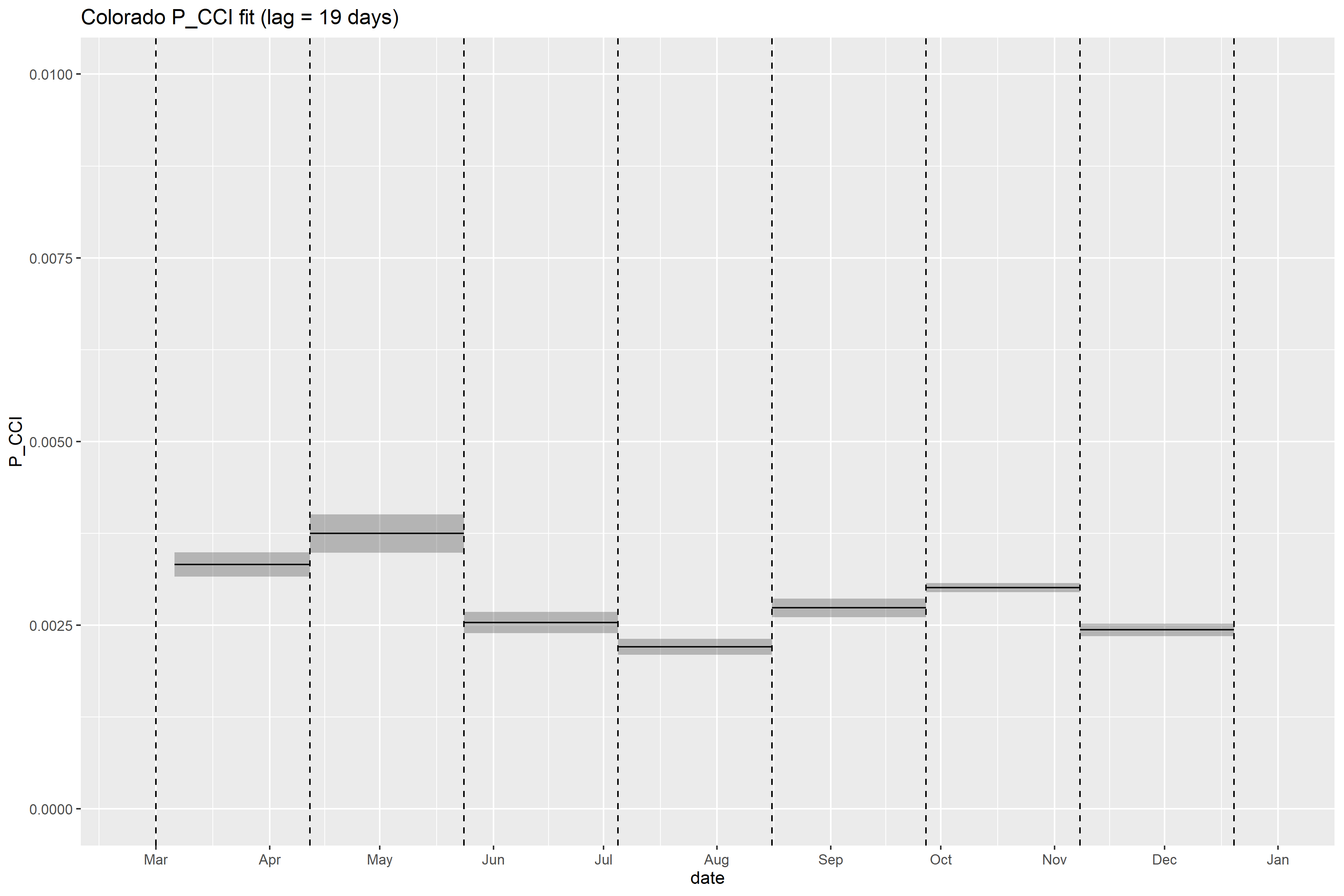

### Colorado_retrospective_forecasts.png

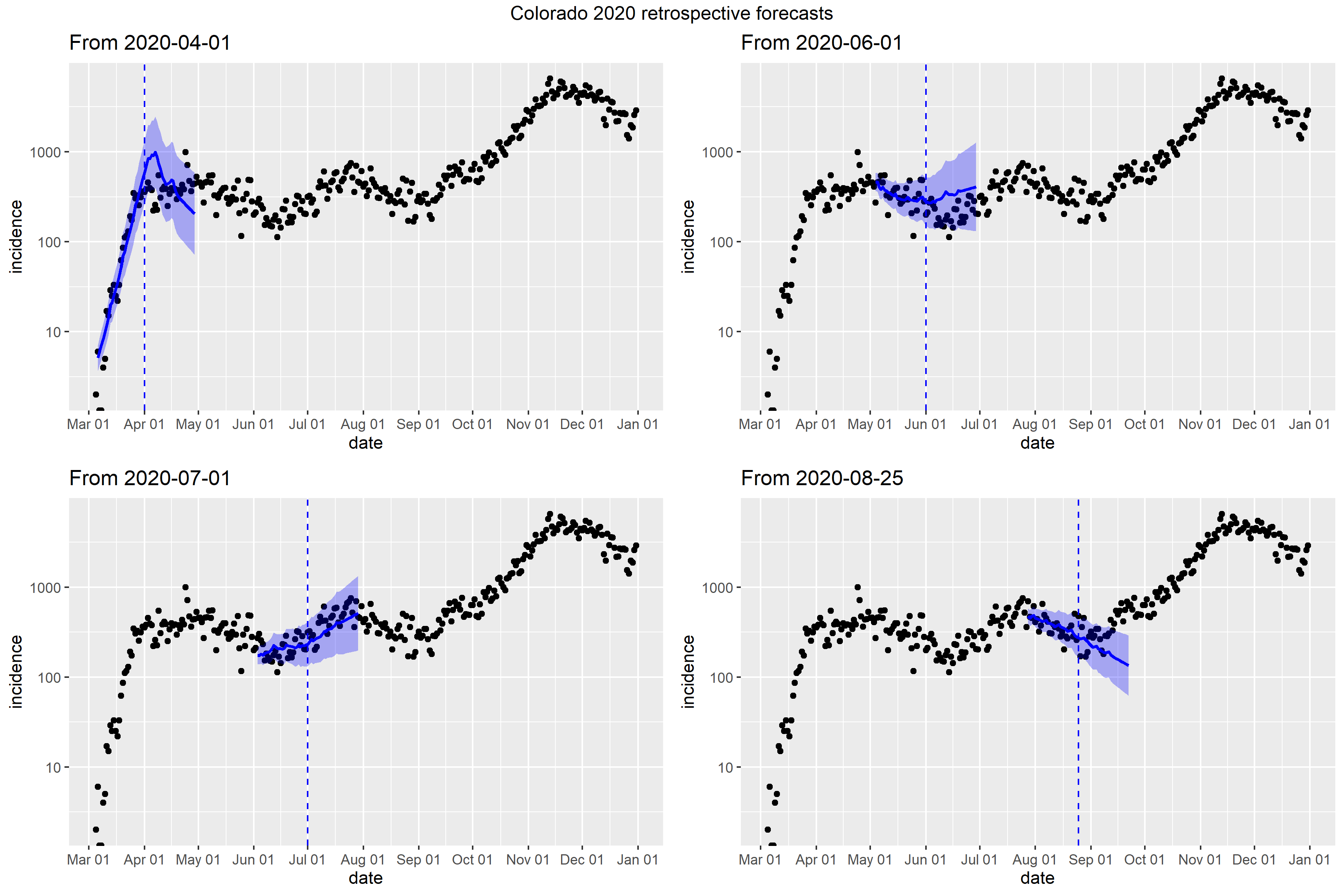

### Connecticut_retrospective_forecasts.png

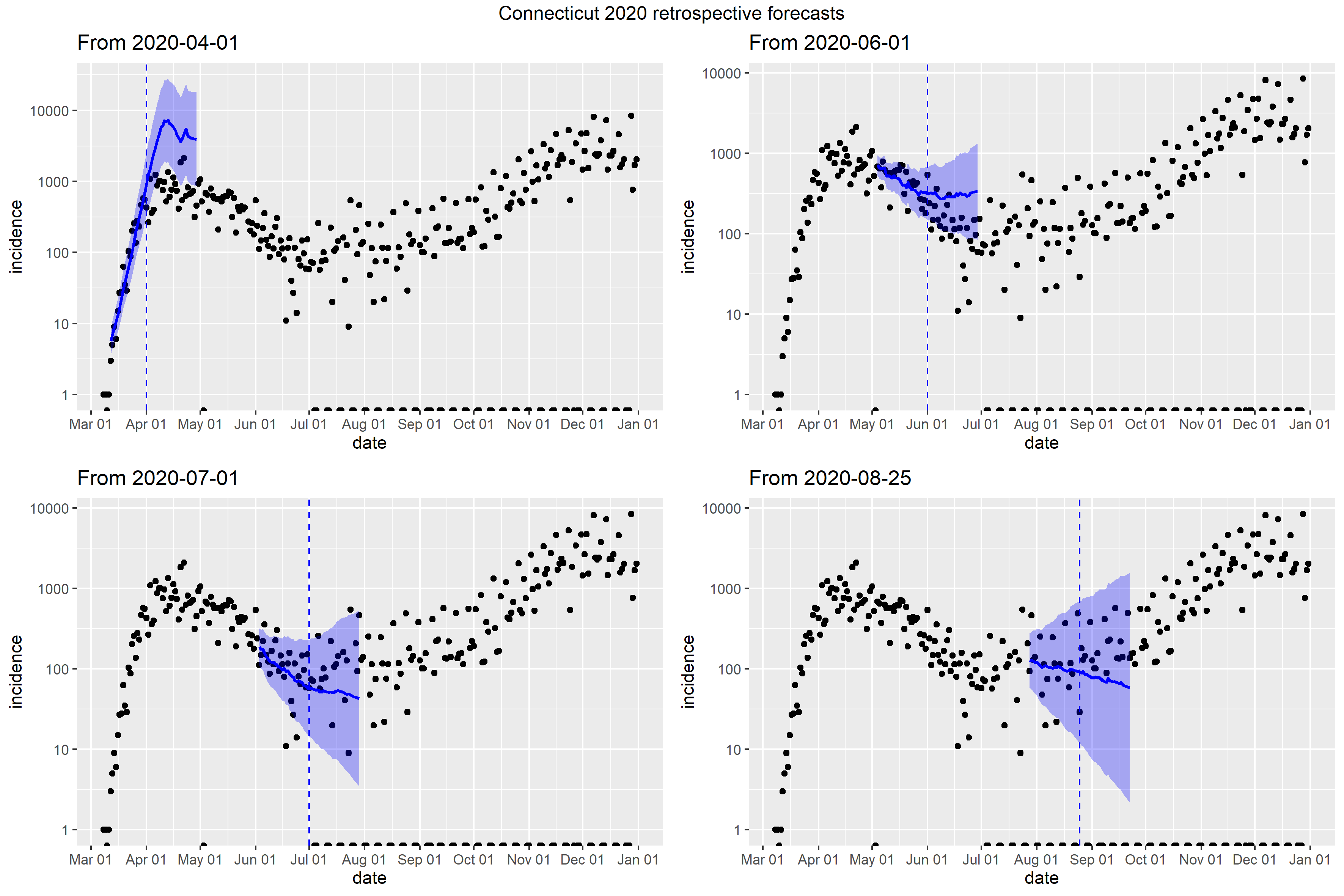

### Delaware_Peff_plot.png

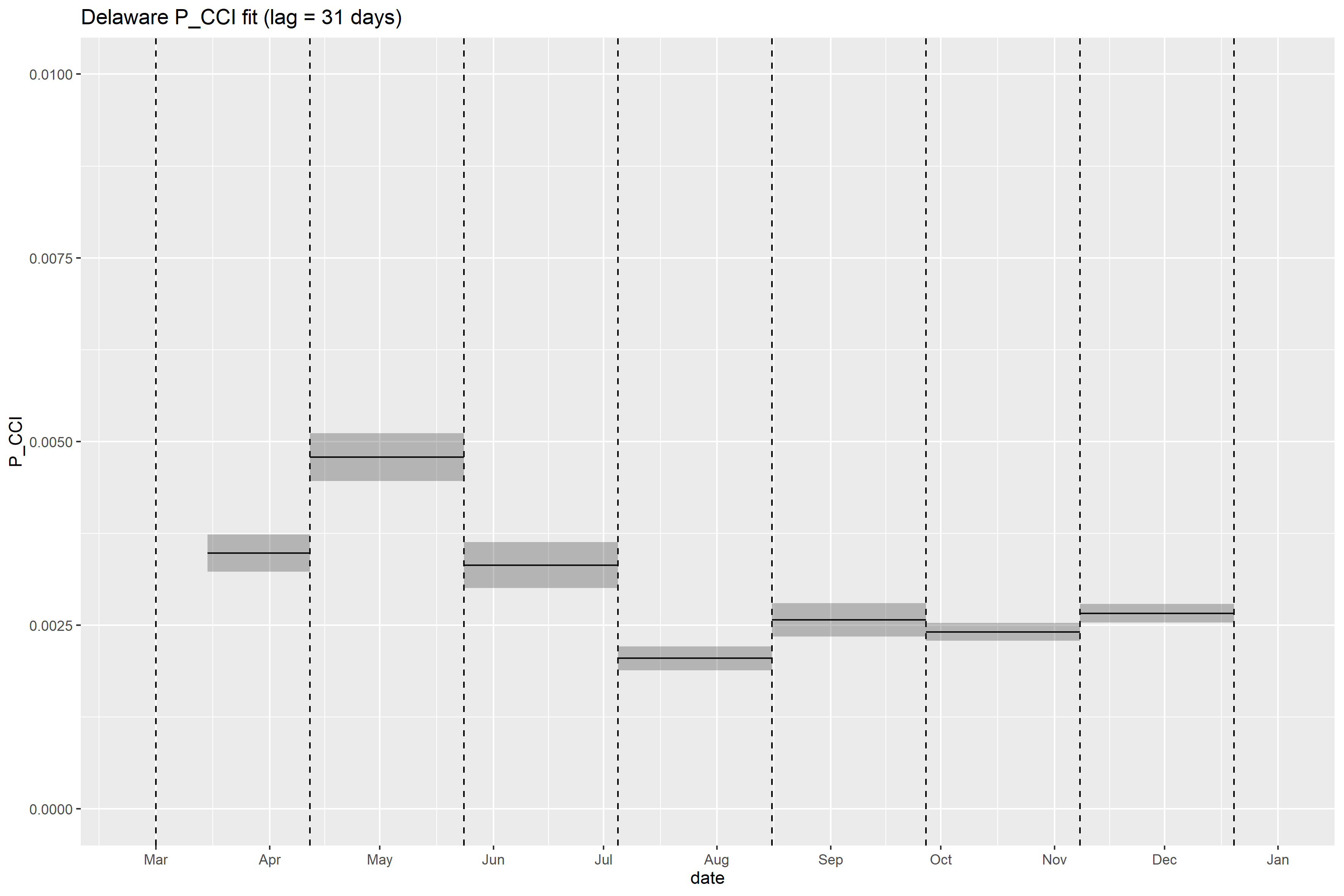

### Delaware_retrospective_forecasts.png

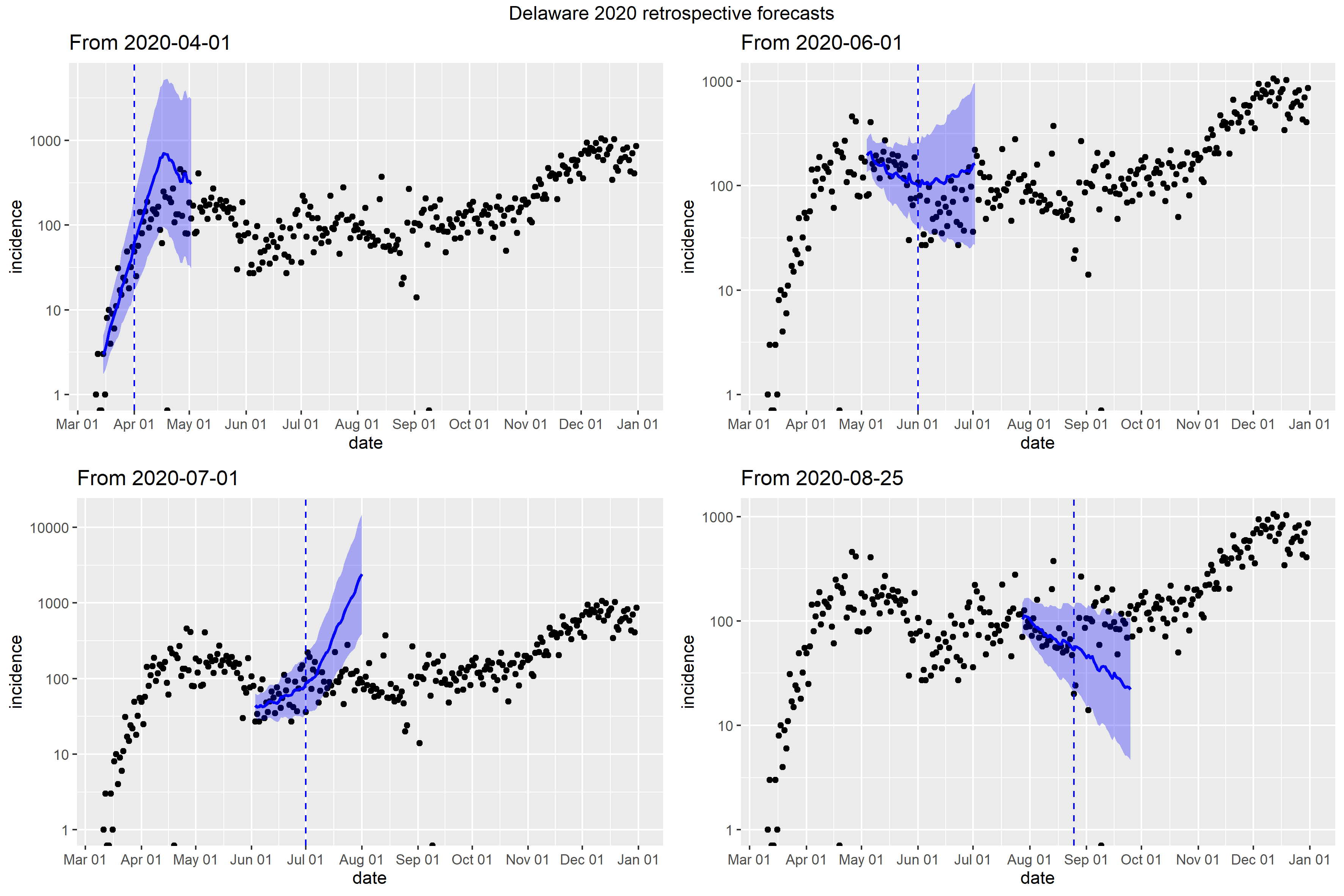

### District of Columbia_retrospective_forecasts.png

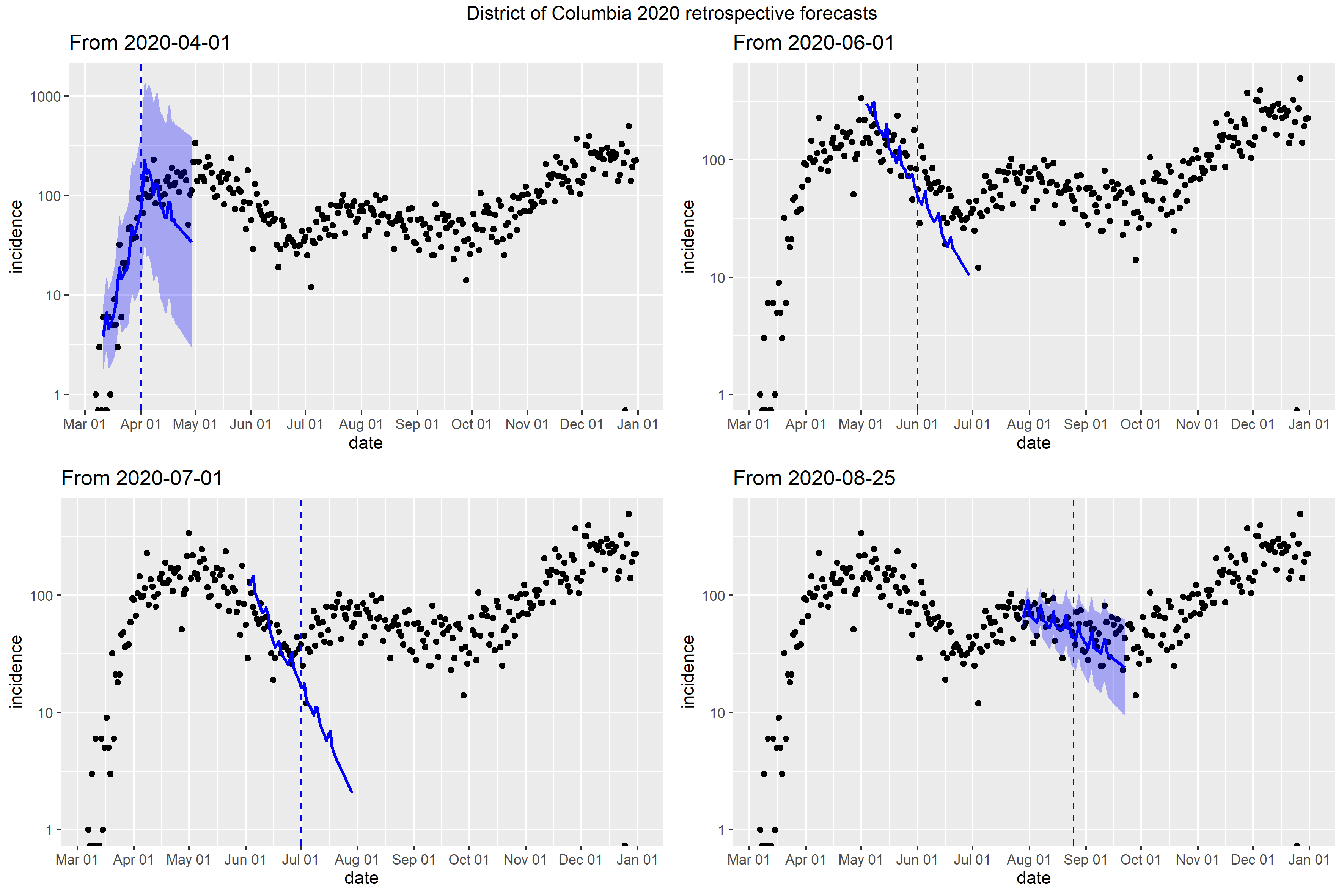

### Florida_Peff_plot.png

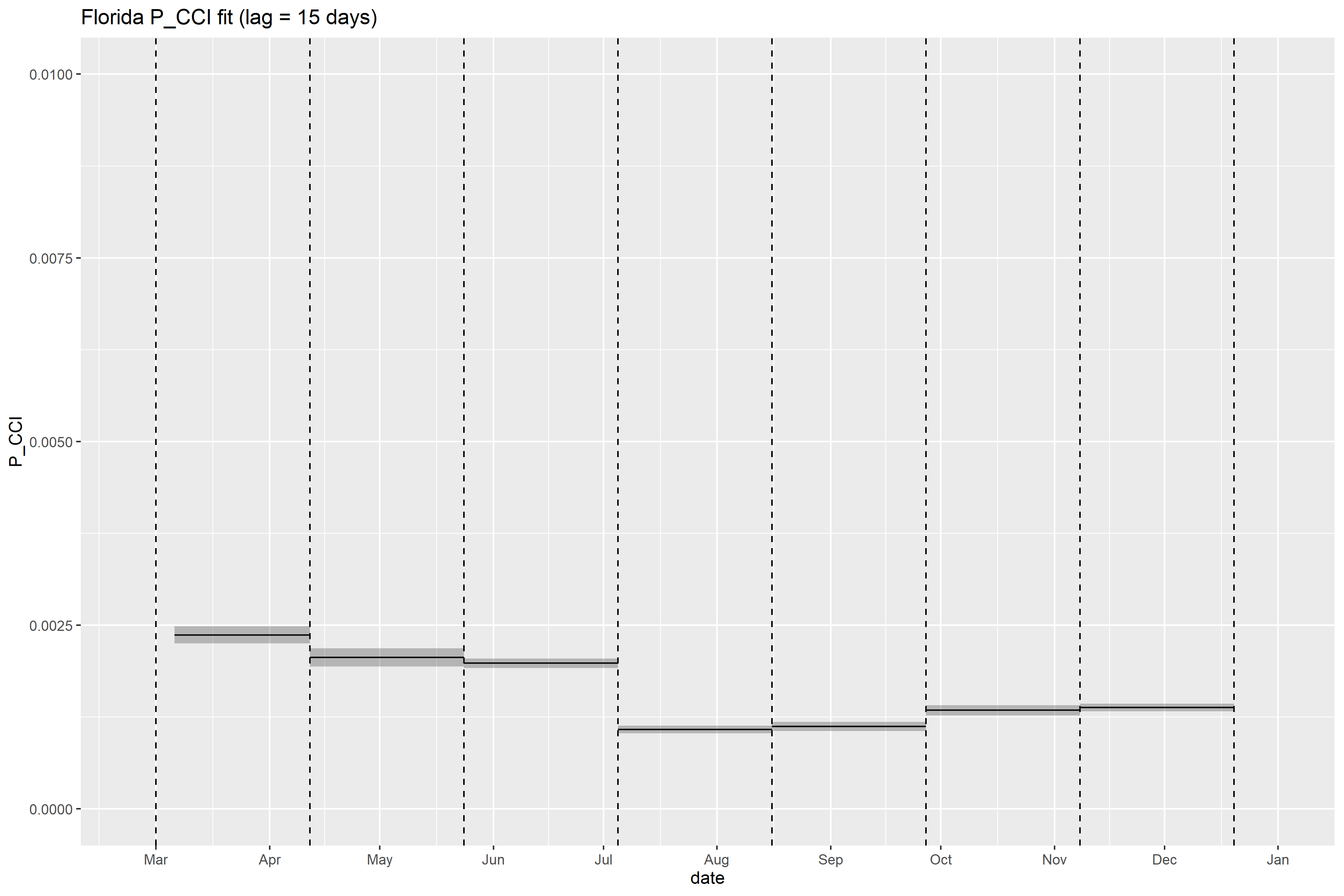

### Florida_retrospective_forecasts.png

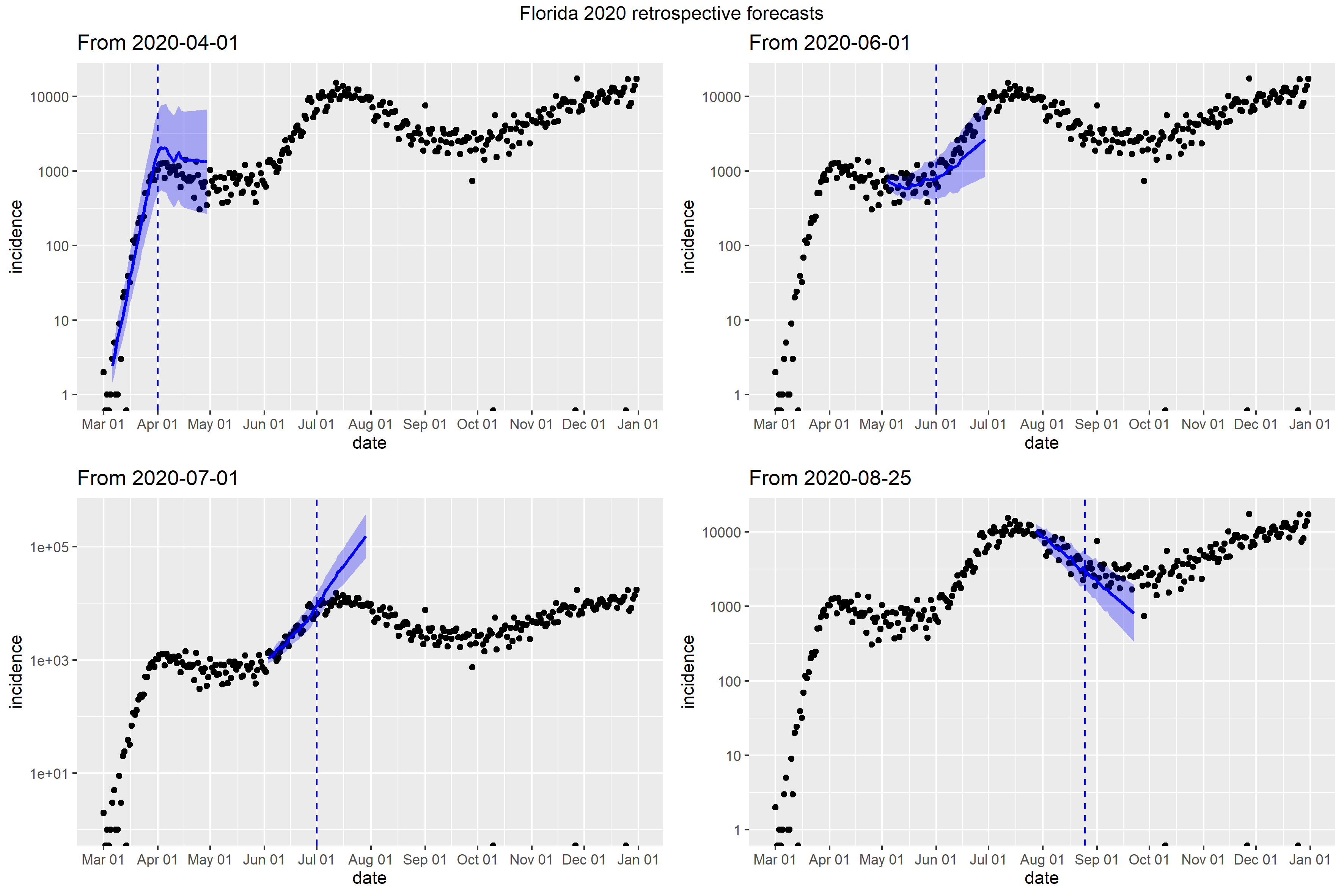

### Georgia_Peff_plot.png

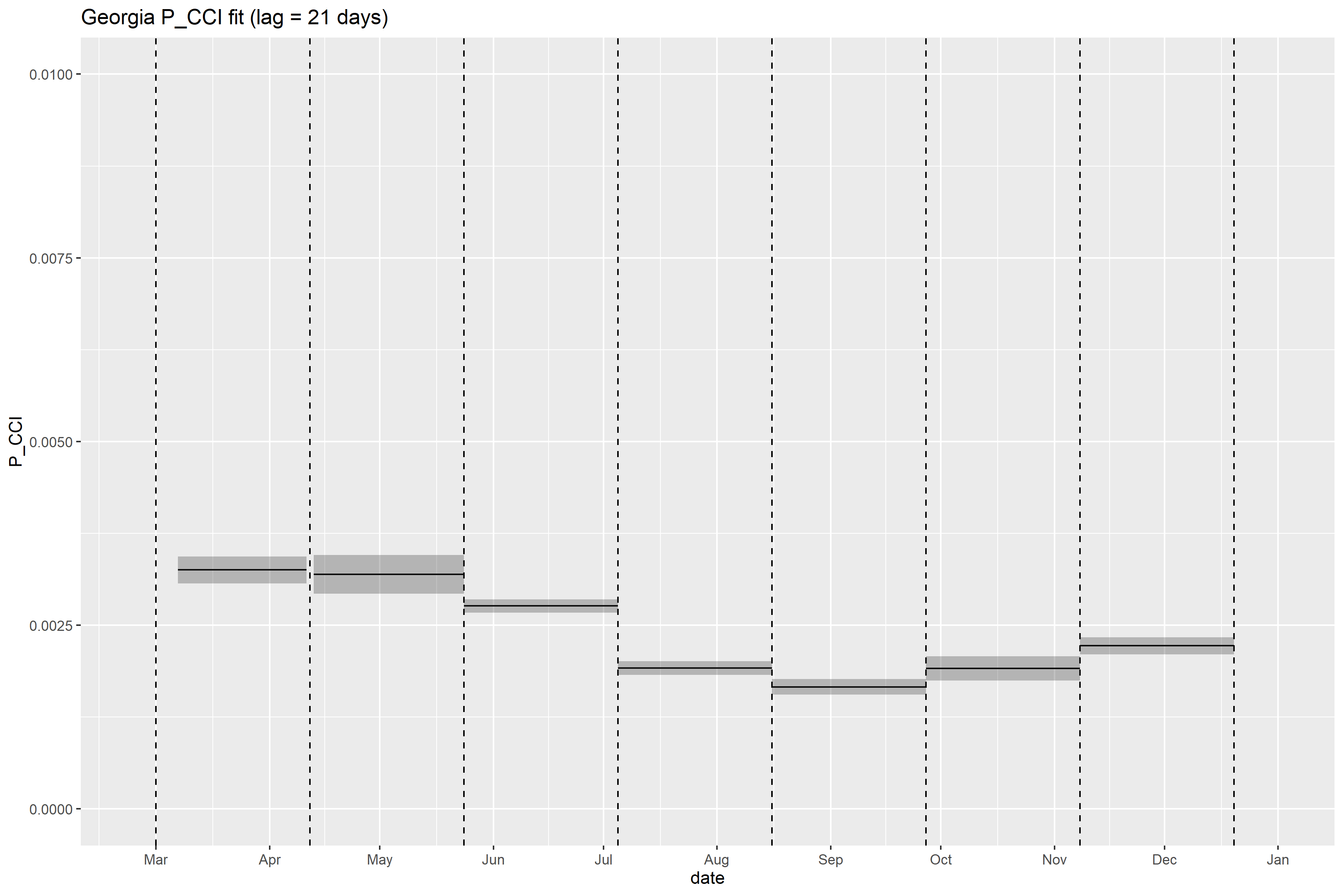

### Georgia_retrospective_forecasts.png

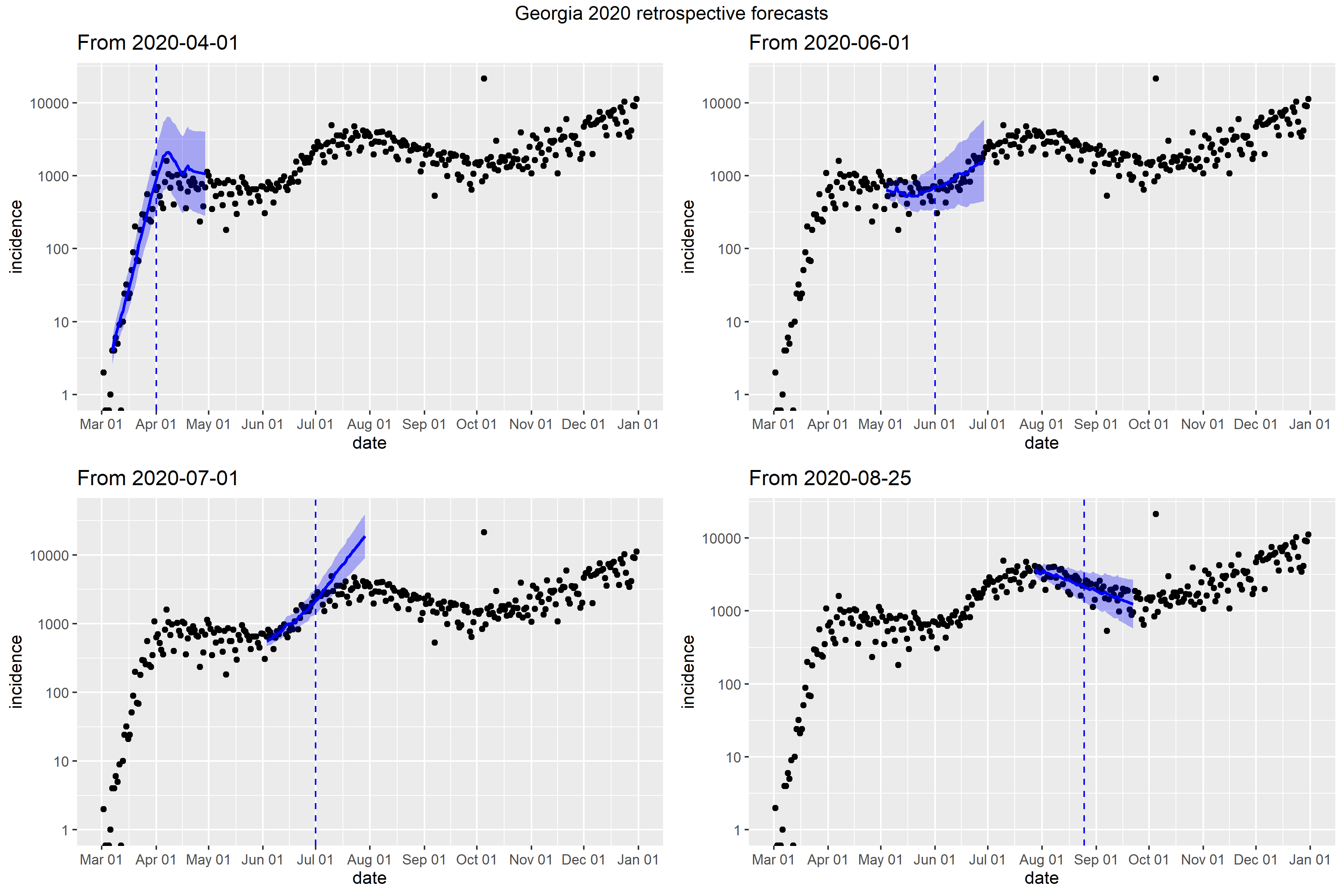

### Hawaii_full_fit_plot.png

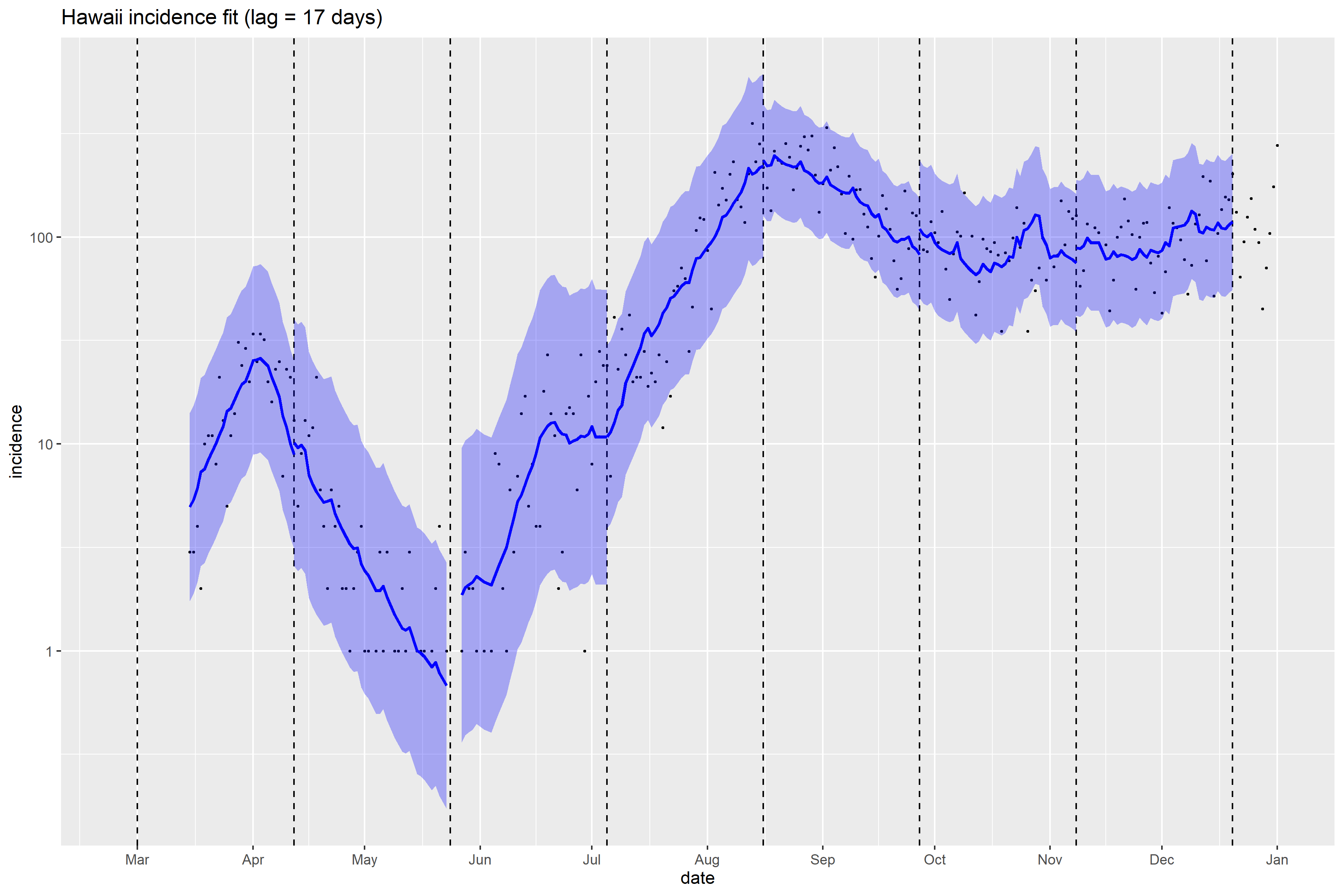

### Hawaii_Peff_plot.png

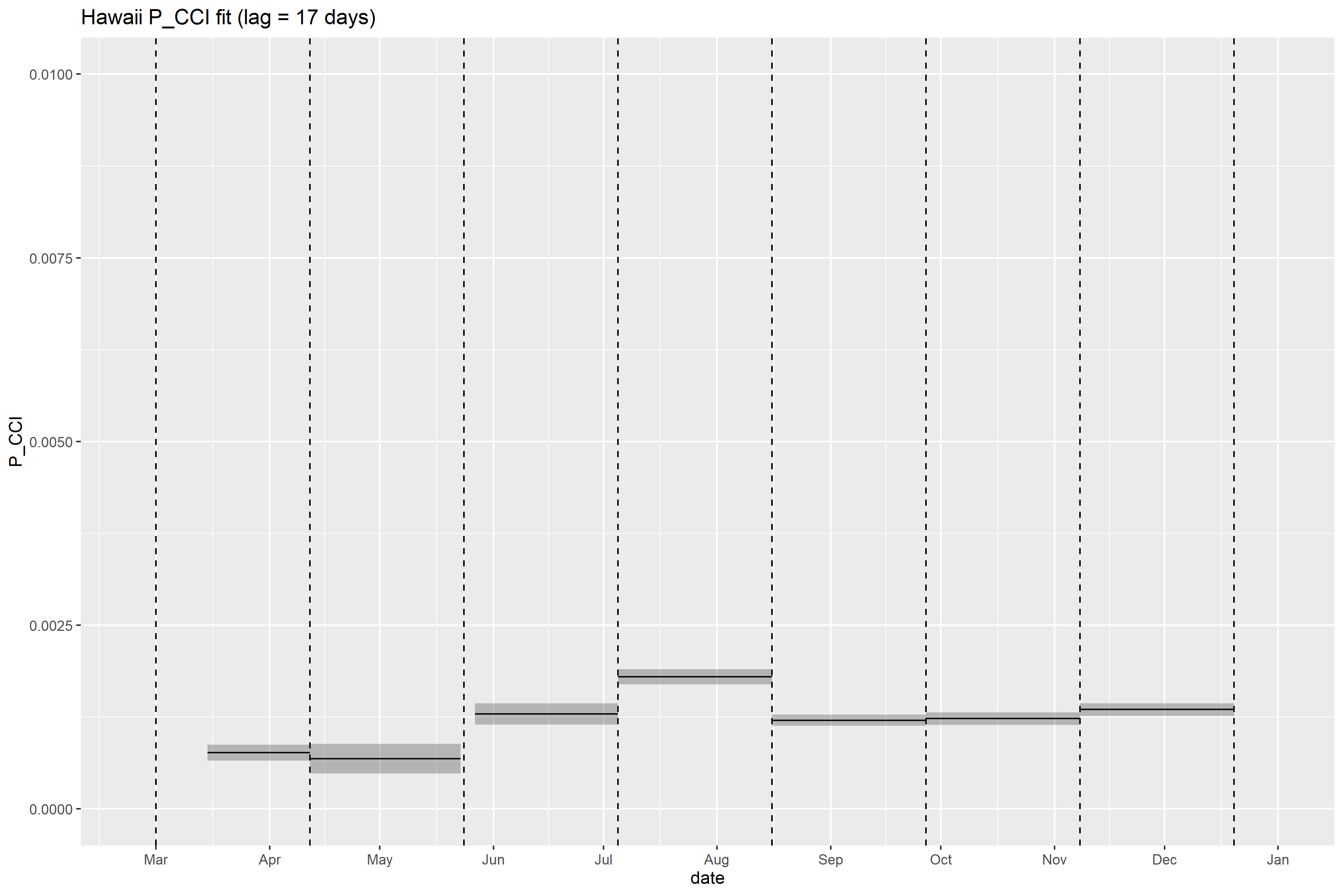

### Hawaii_retrospective_forecasts.png

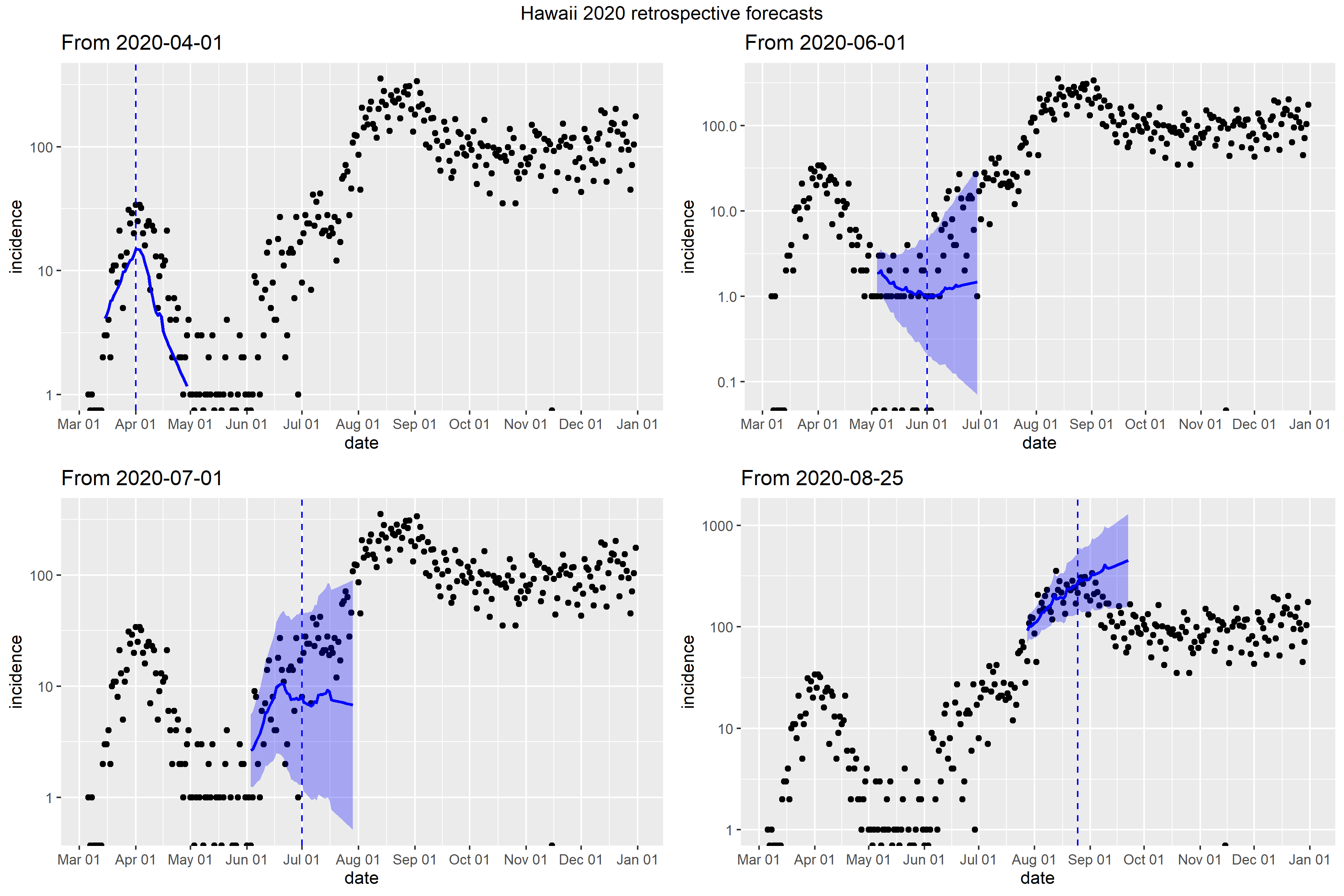

### Idaho_full_fit_plot.png

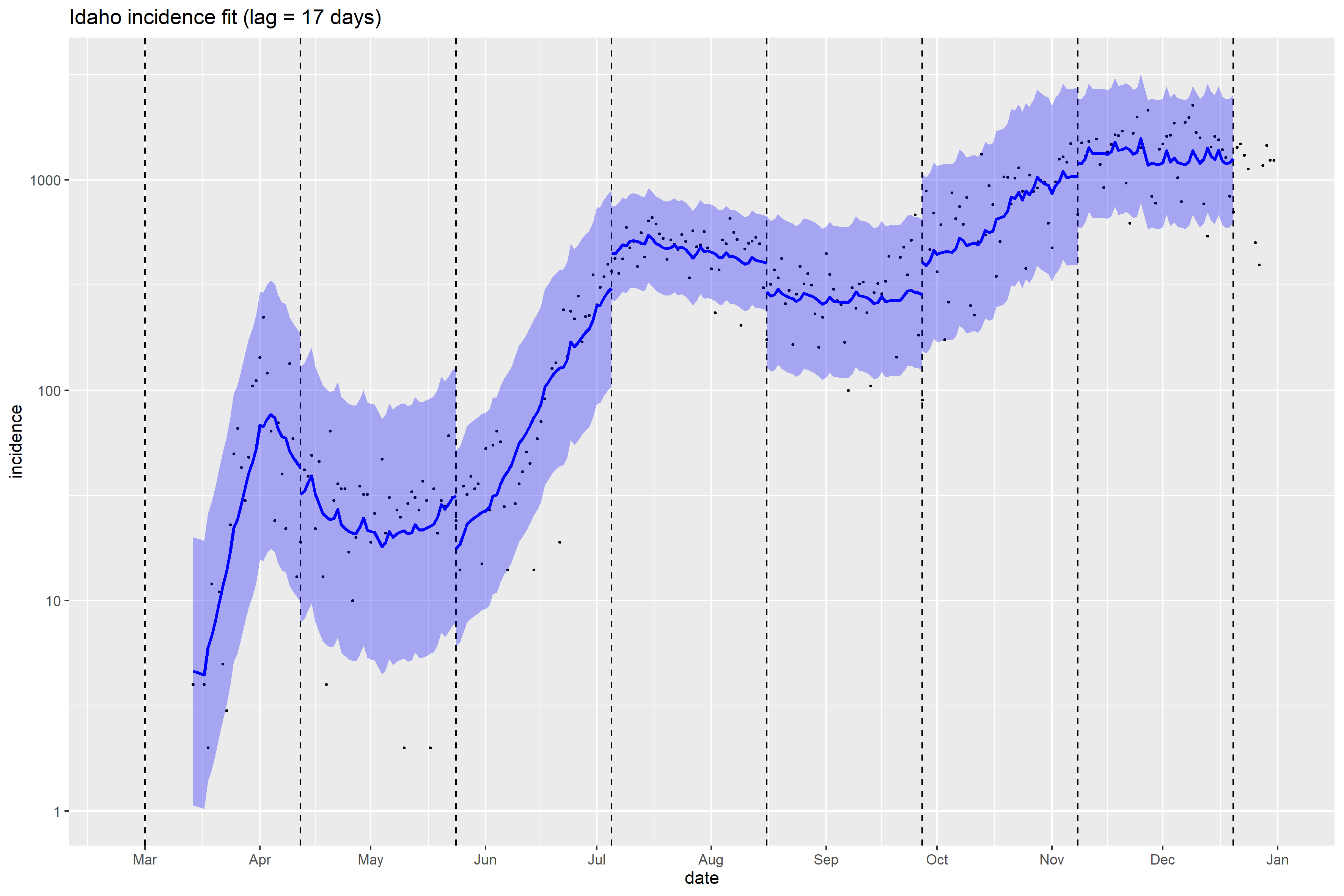

### Idaho_Peff_plot.png

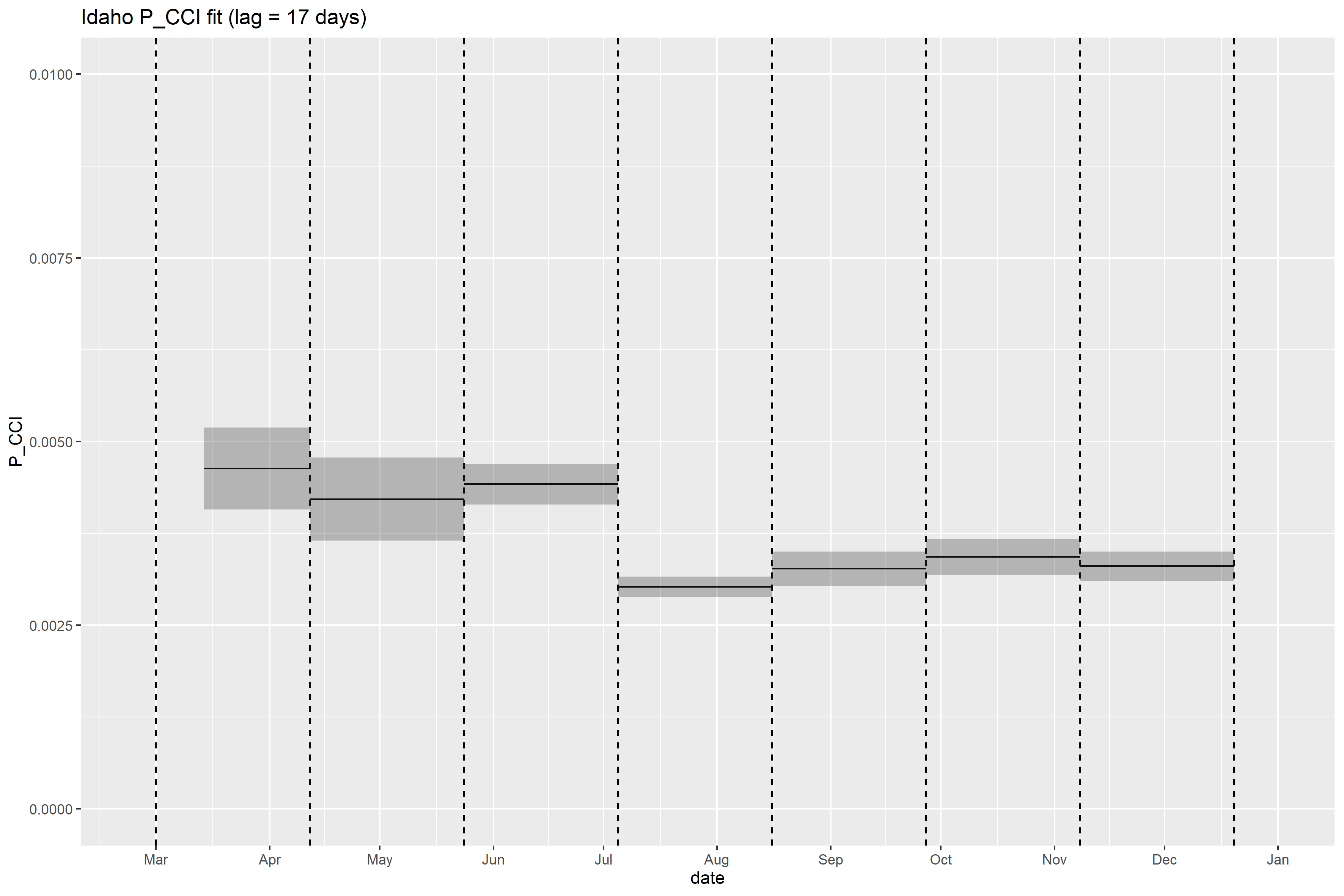

### Idaho_retrospective_forecasts.png

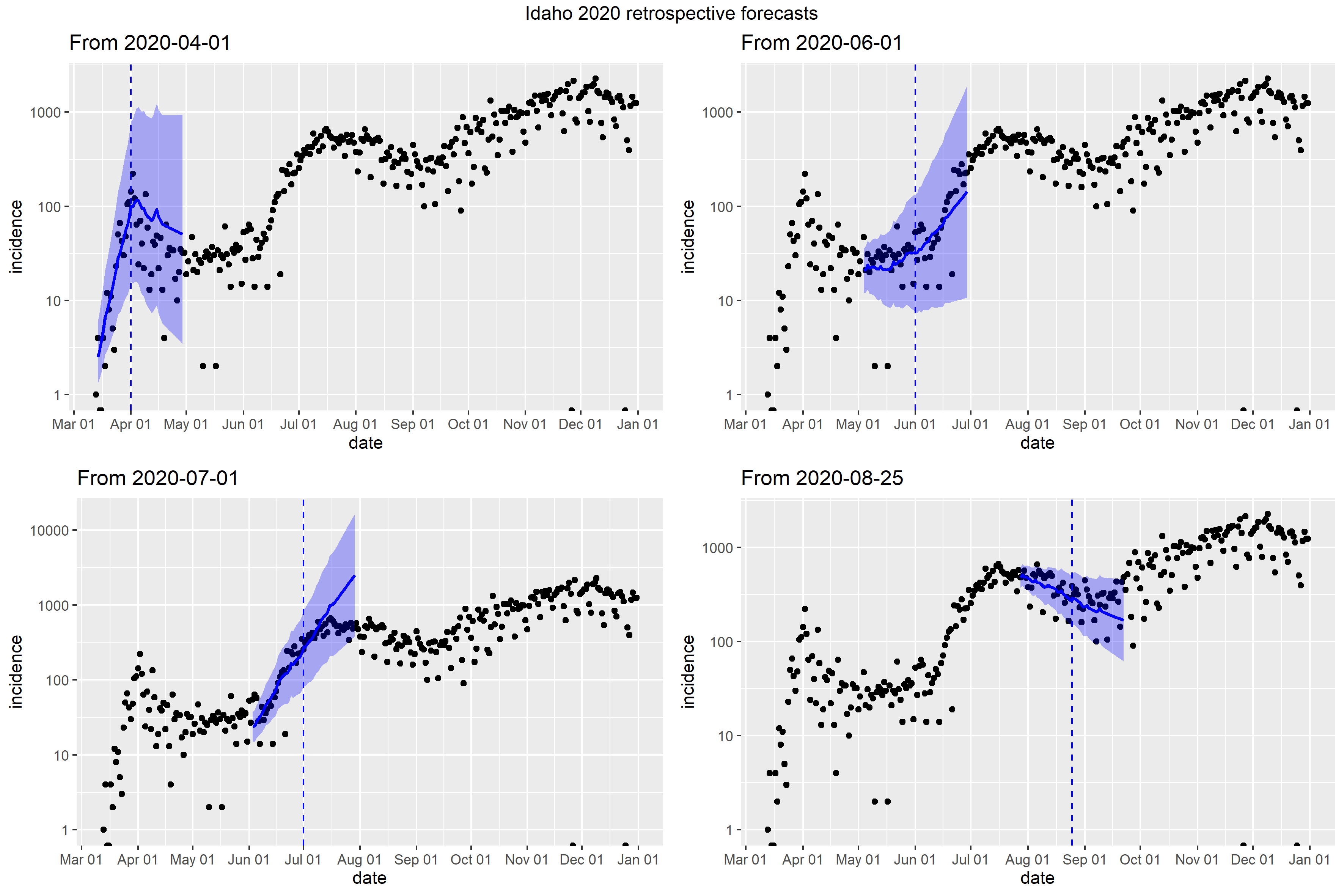

### Illinois_Peff_plot.png

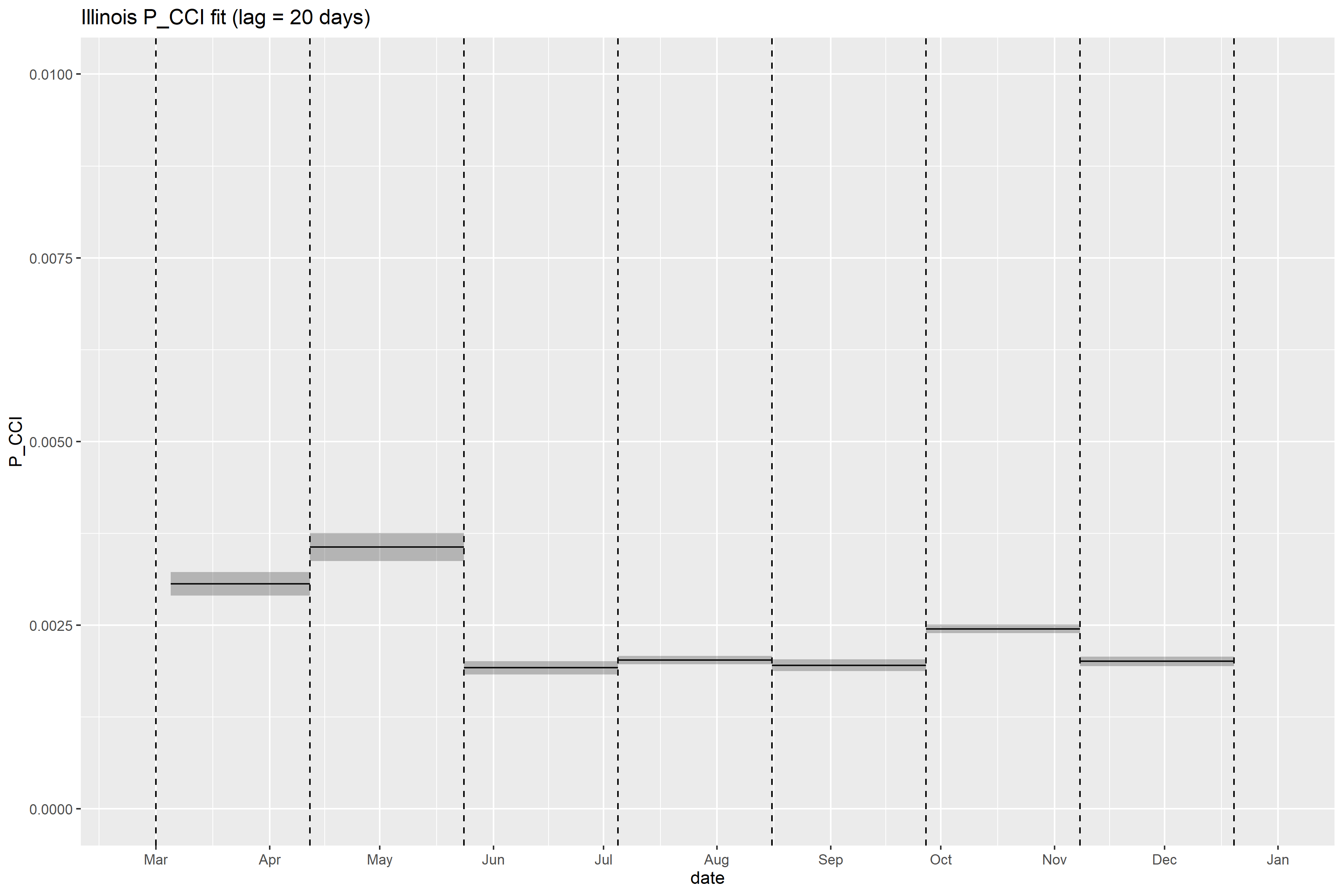

### Illinois_retrospective_forecasts.png

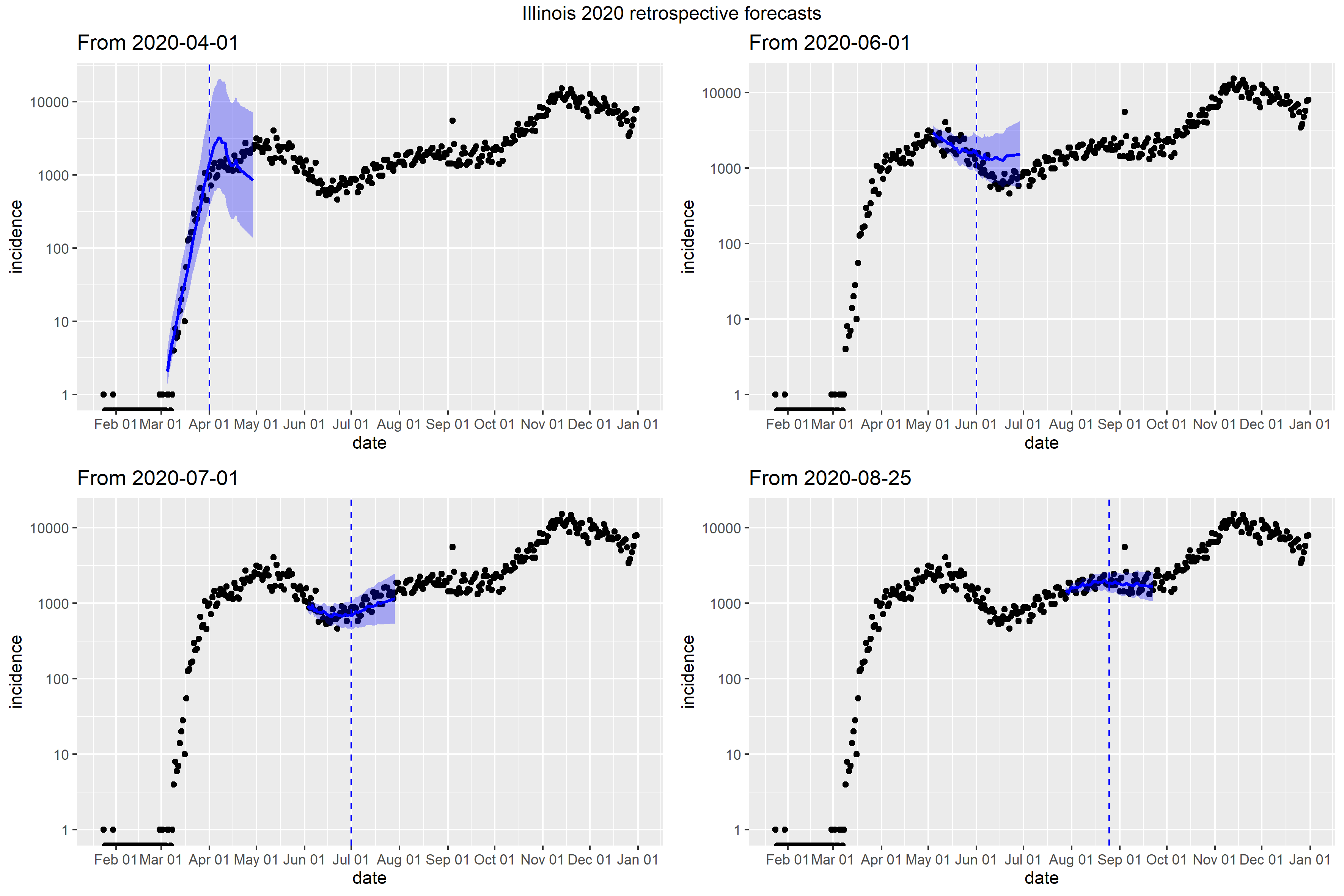

### Indiana_Peff_plot.png

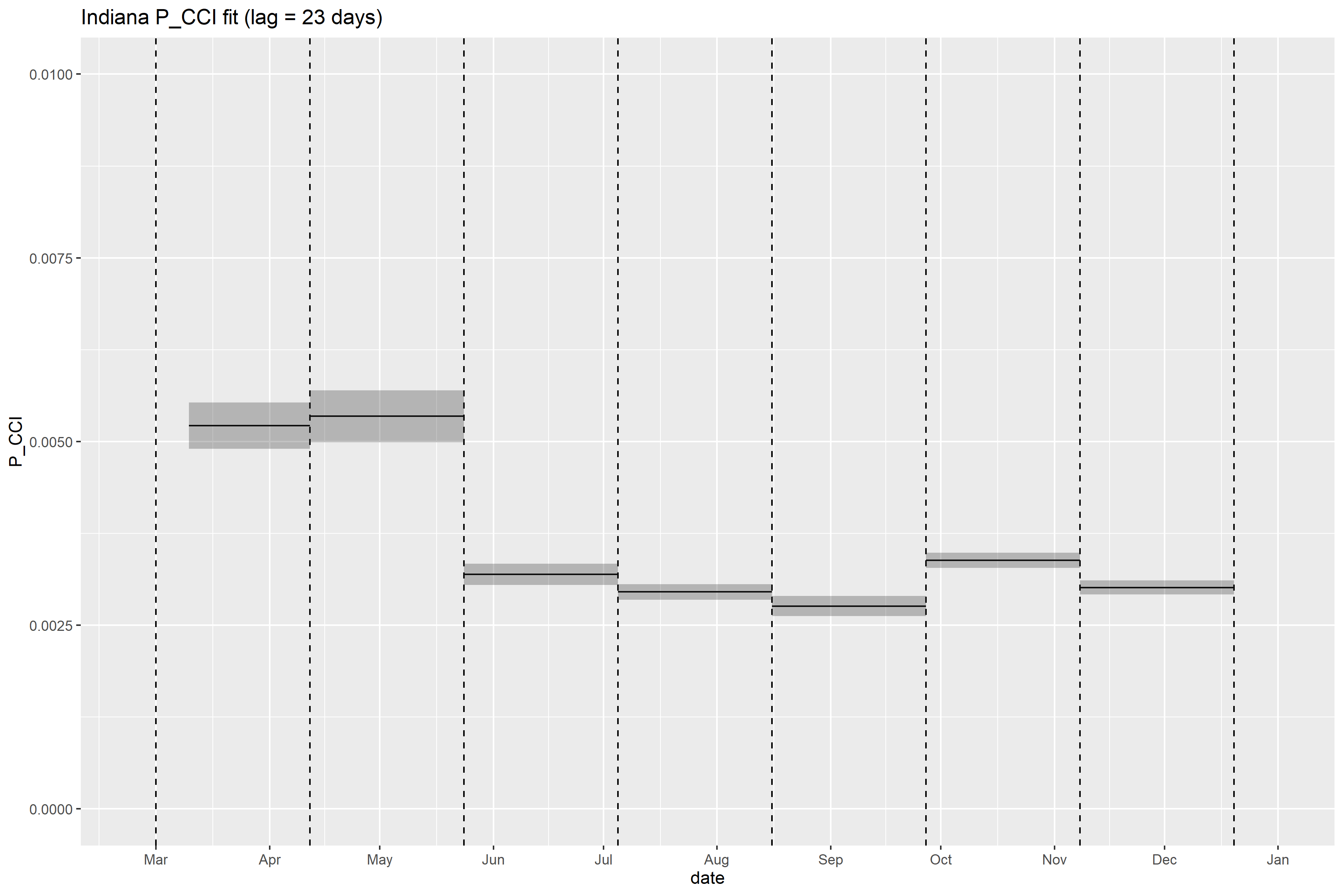

### Indiana_retrospective_forecasts.png

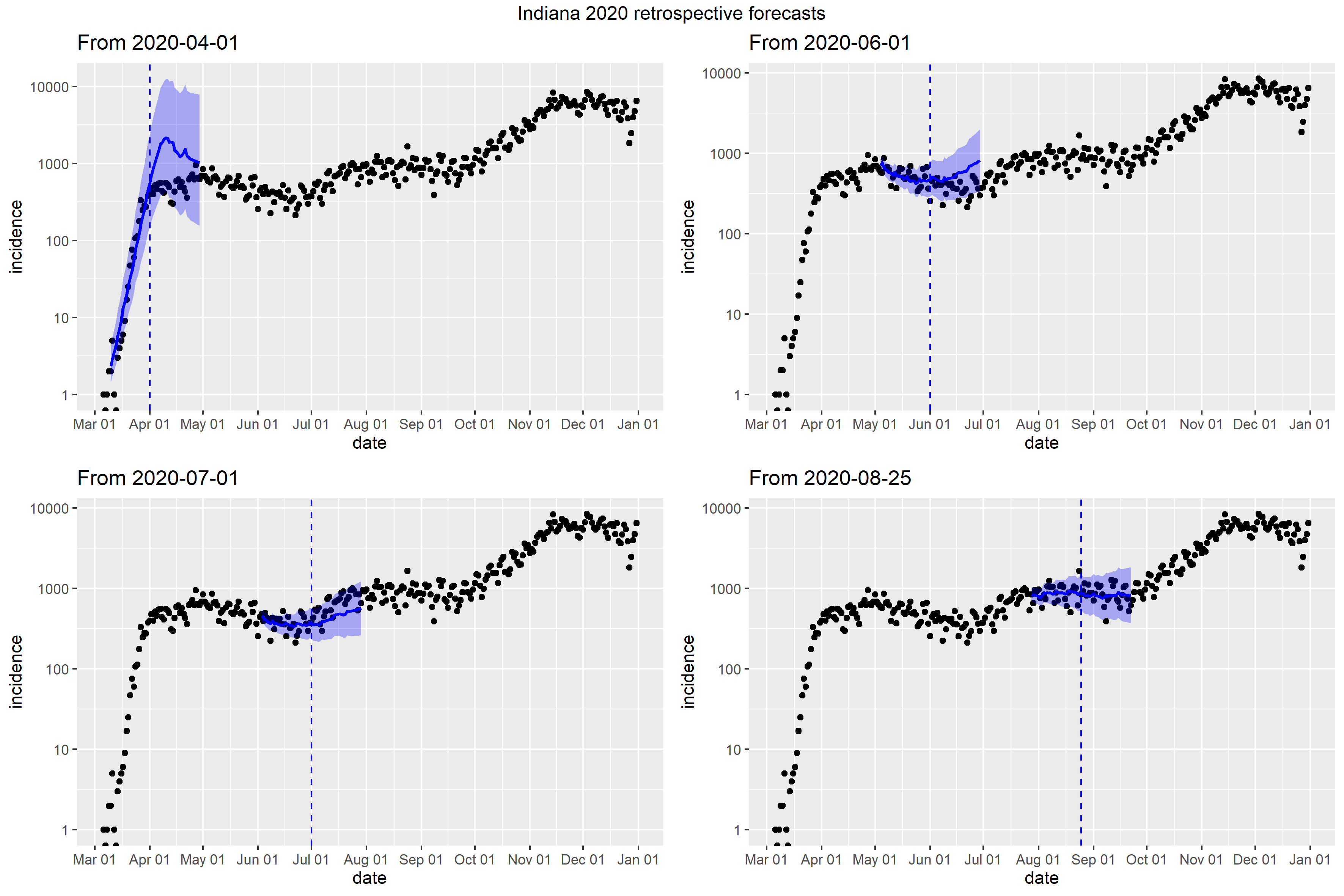

### Iowa_full_fit_plot.png

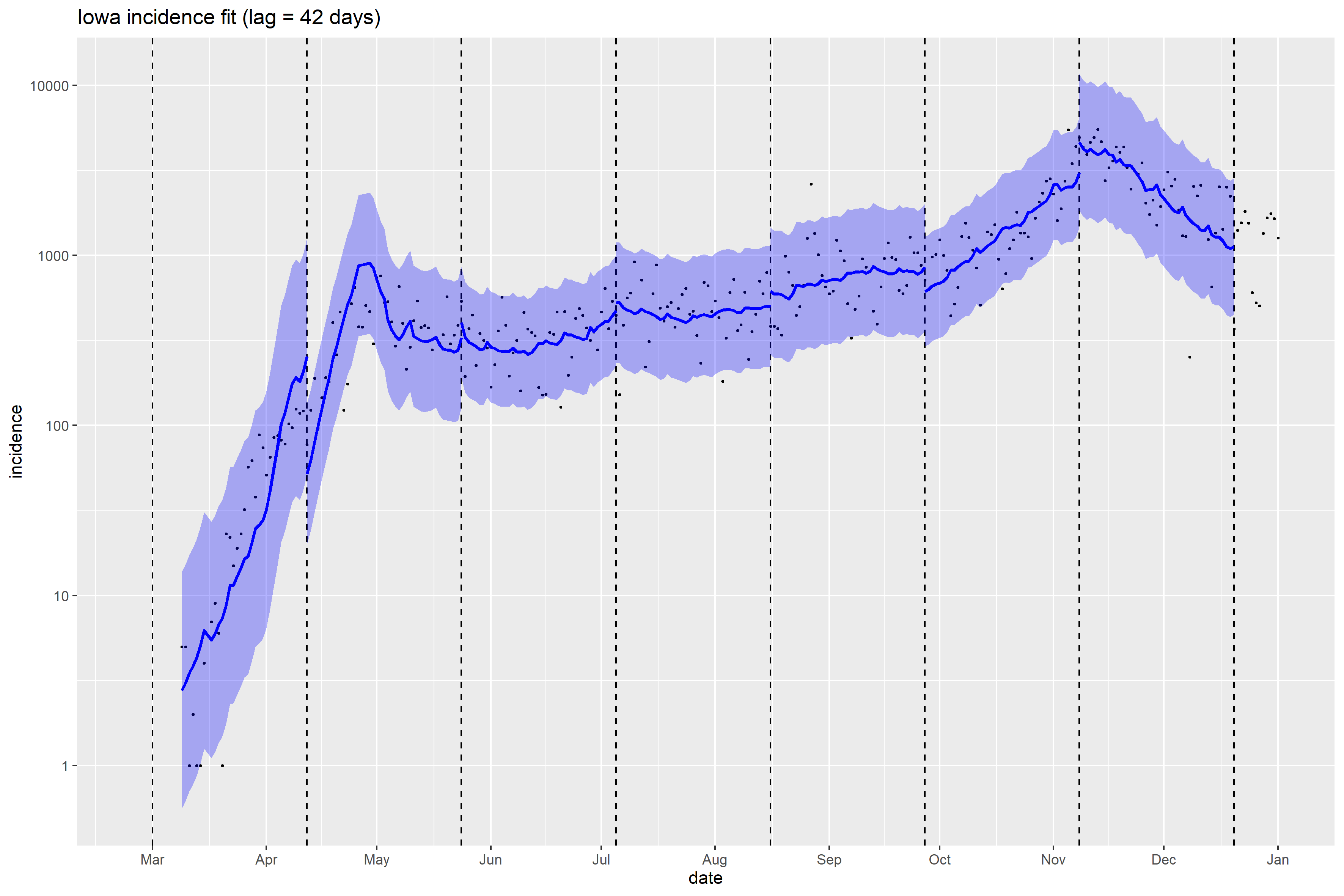

### Iowa_Peff_plot.png

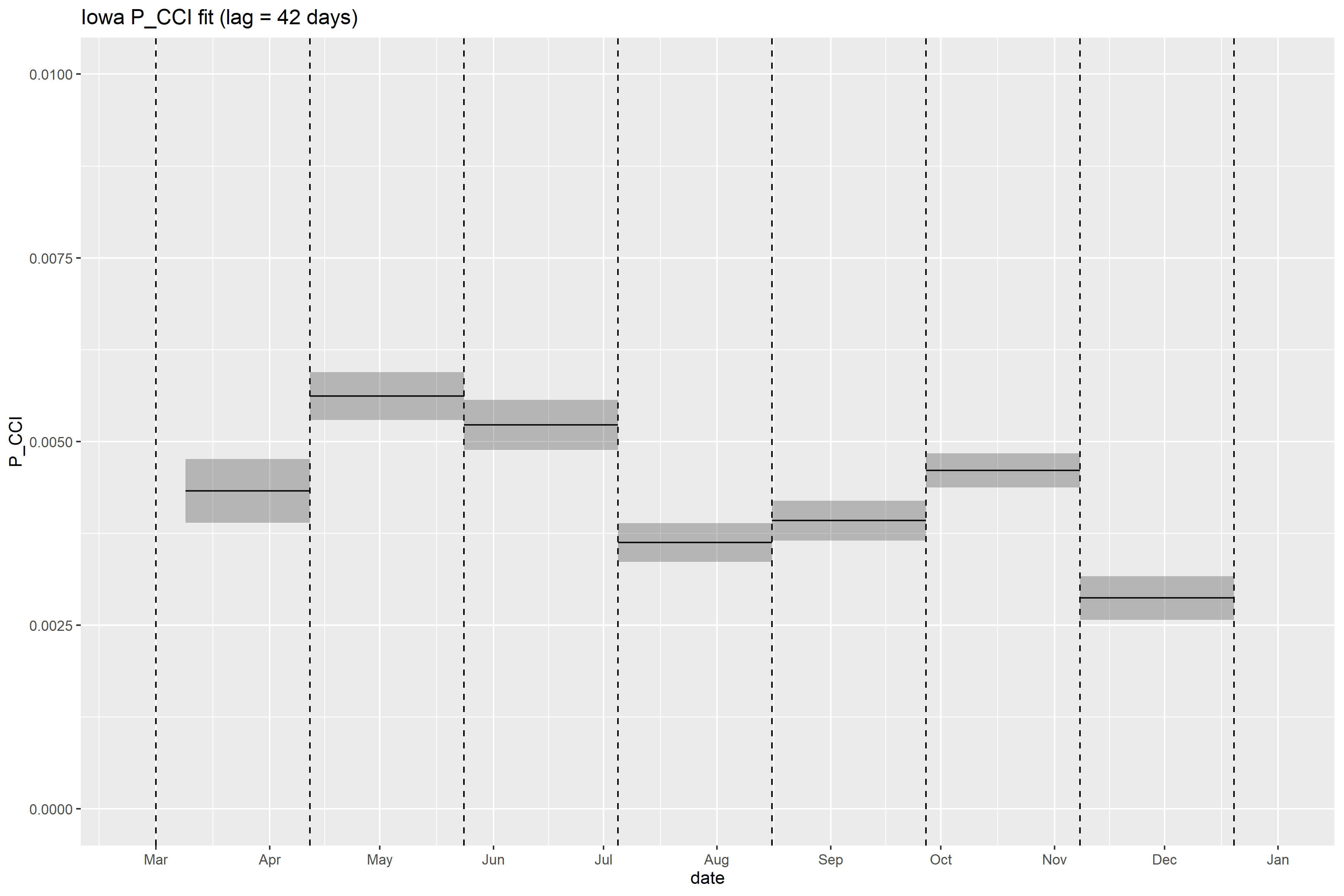

### Iowa_retrospective_forecasts.png

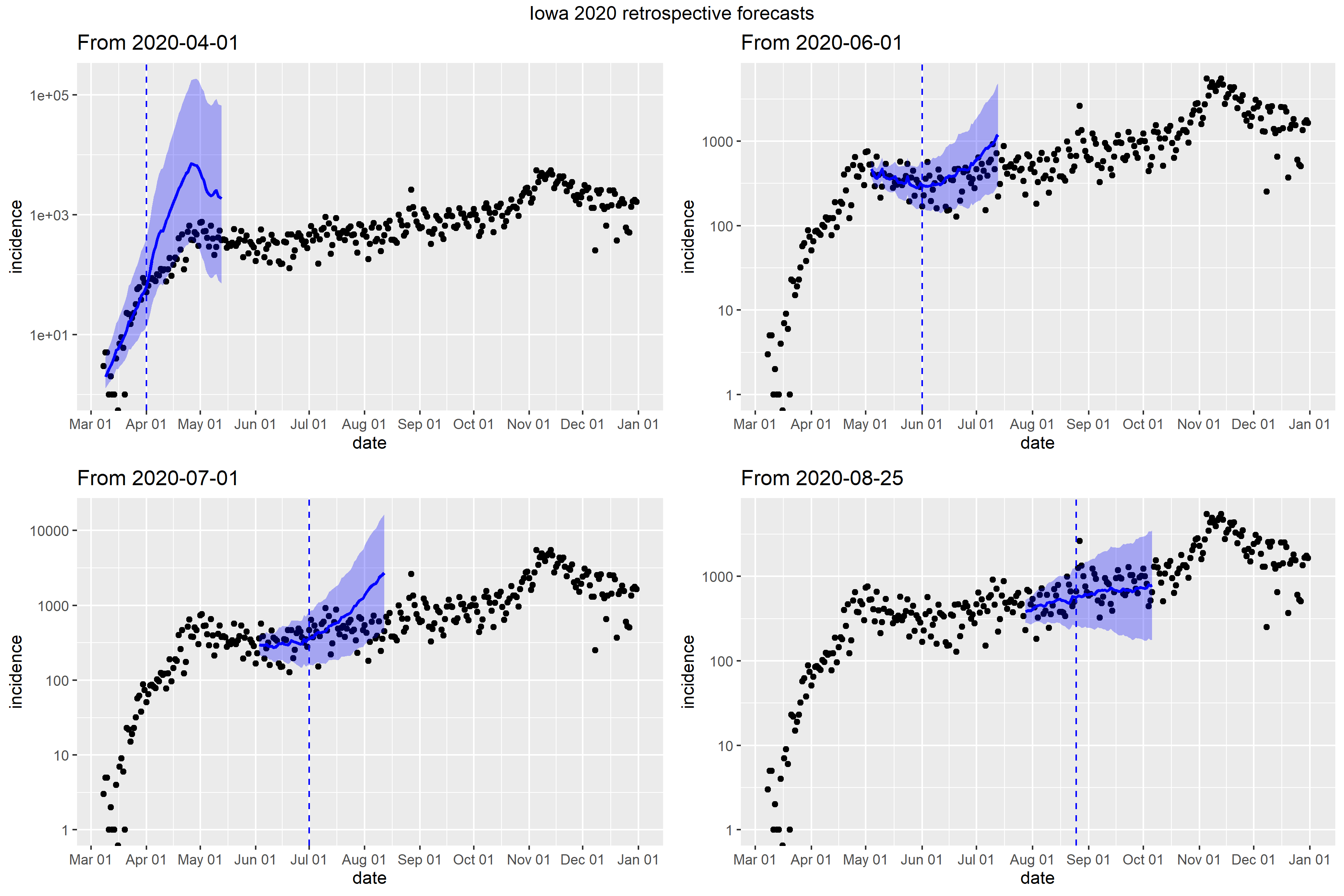

### Kansas_full_fit_plot.png

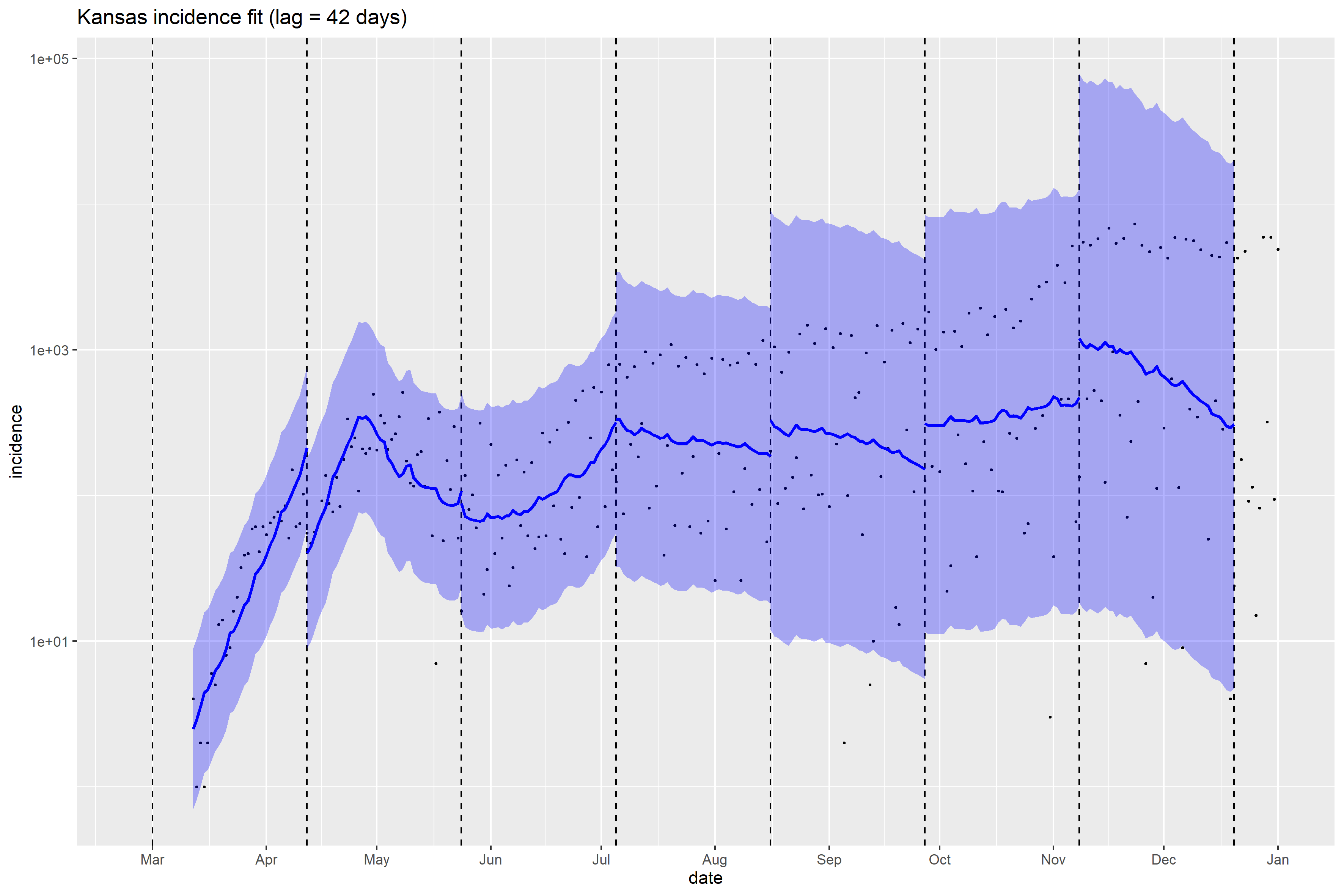

### Kansas_Peff_plot.png

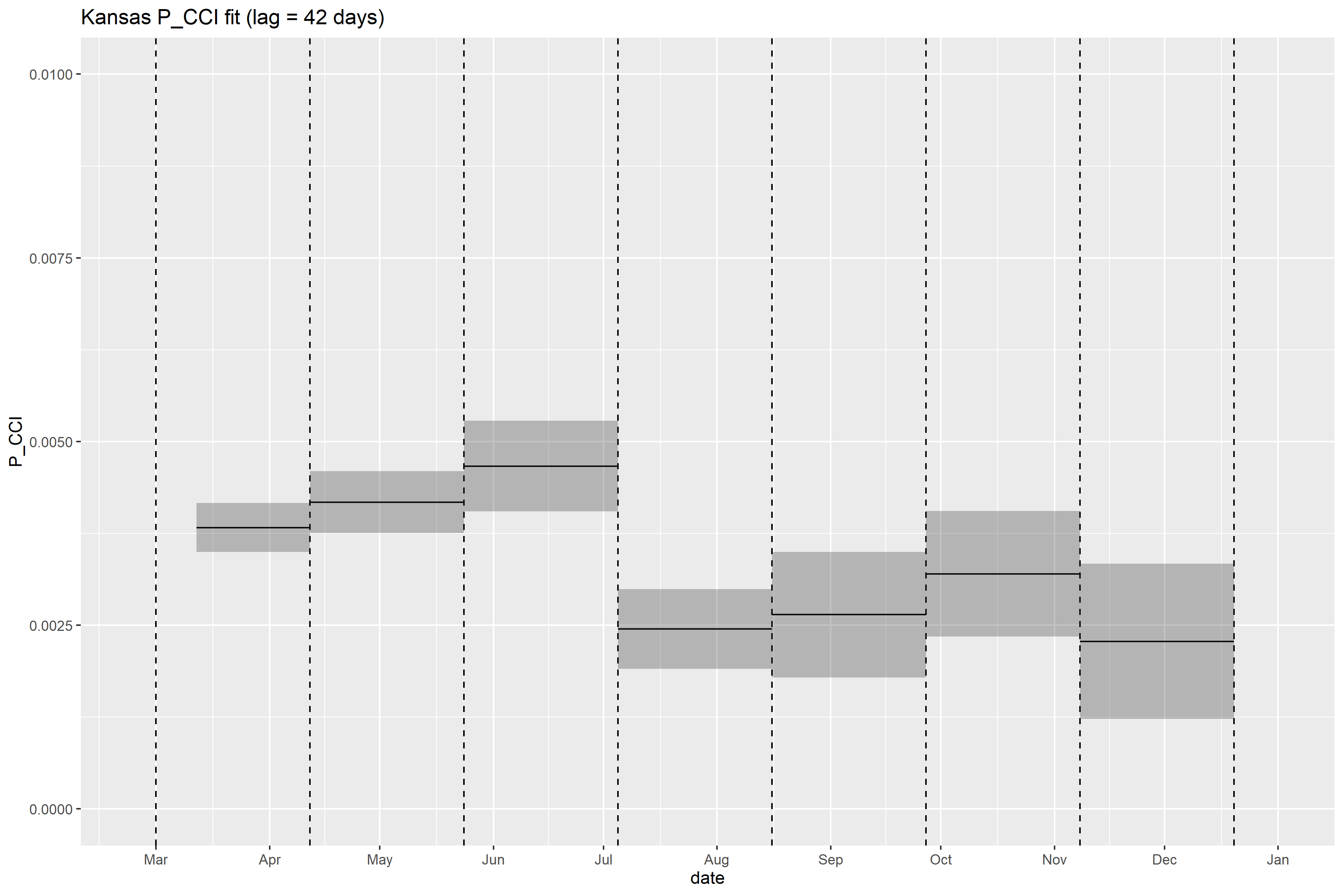

### Kansas_retrospective_forecasts.png

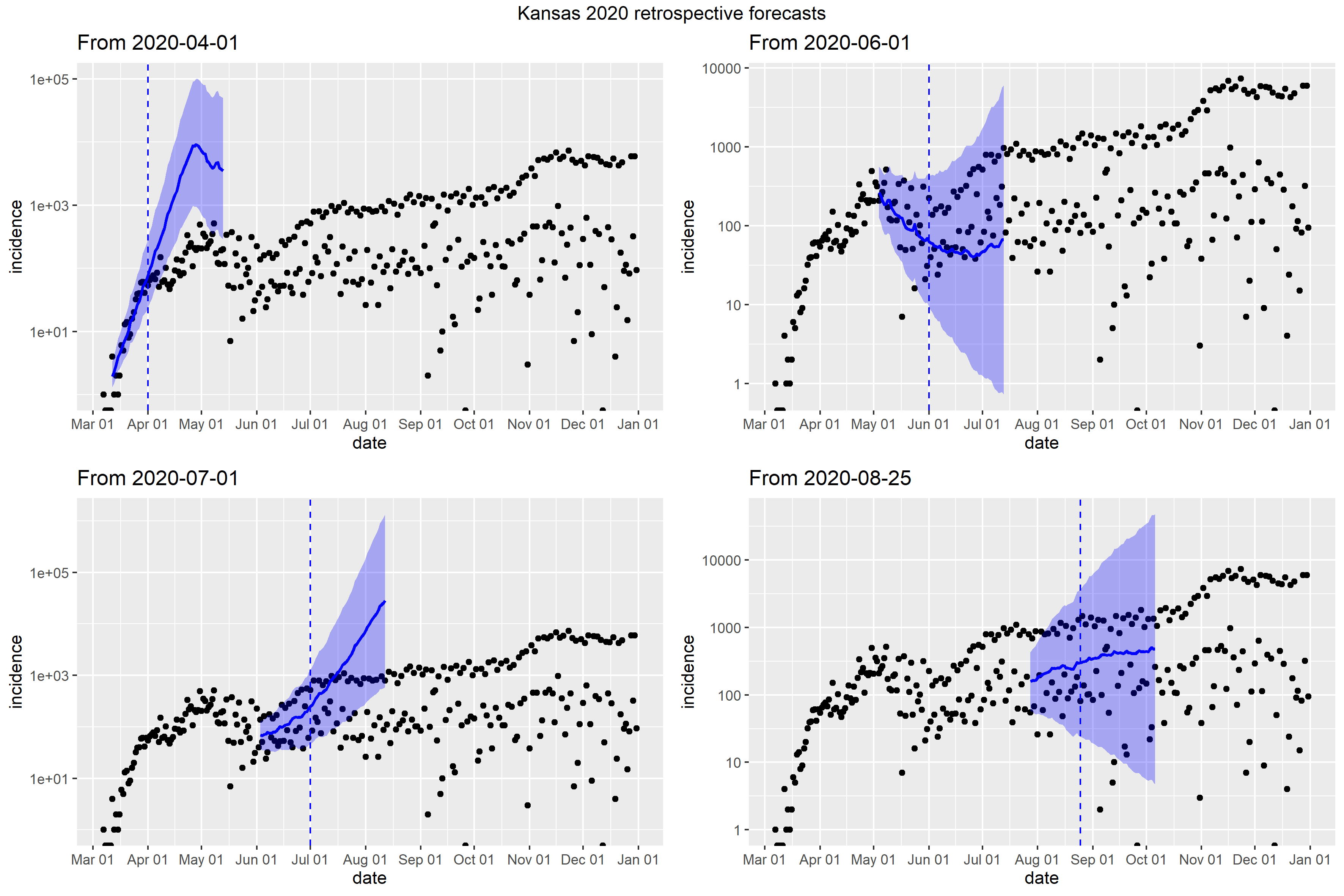

### Kentucky_Peff_plot.png

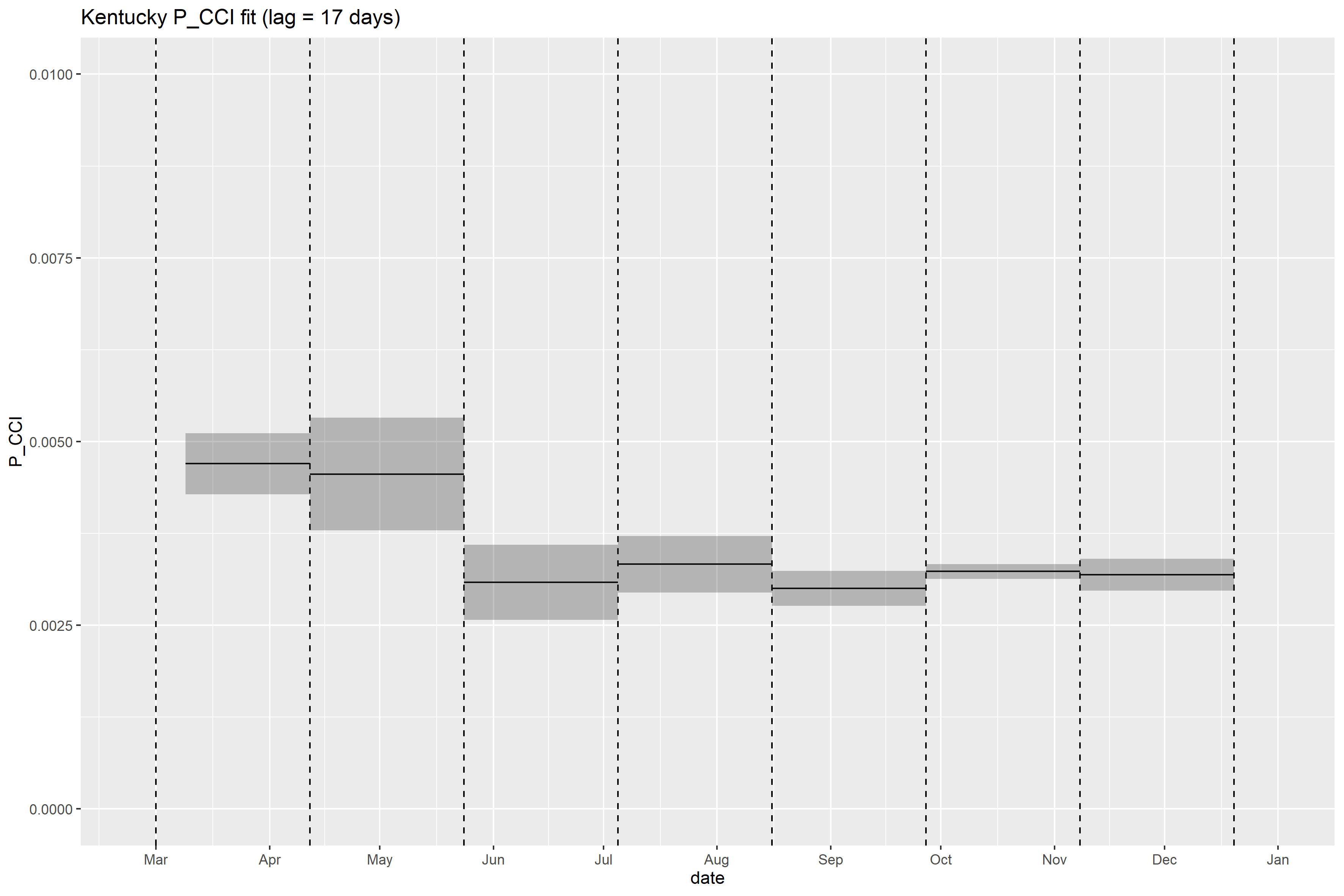

### Kentucky_retrospective_forecasts.png

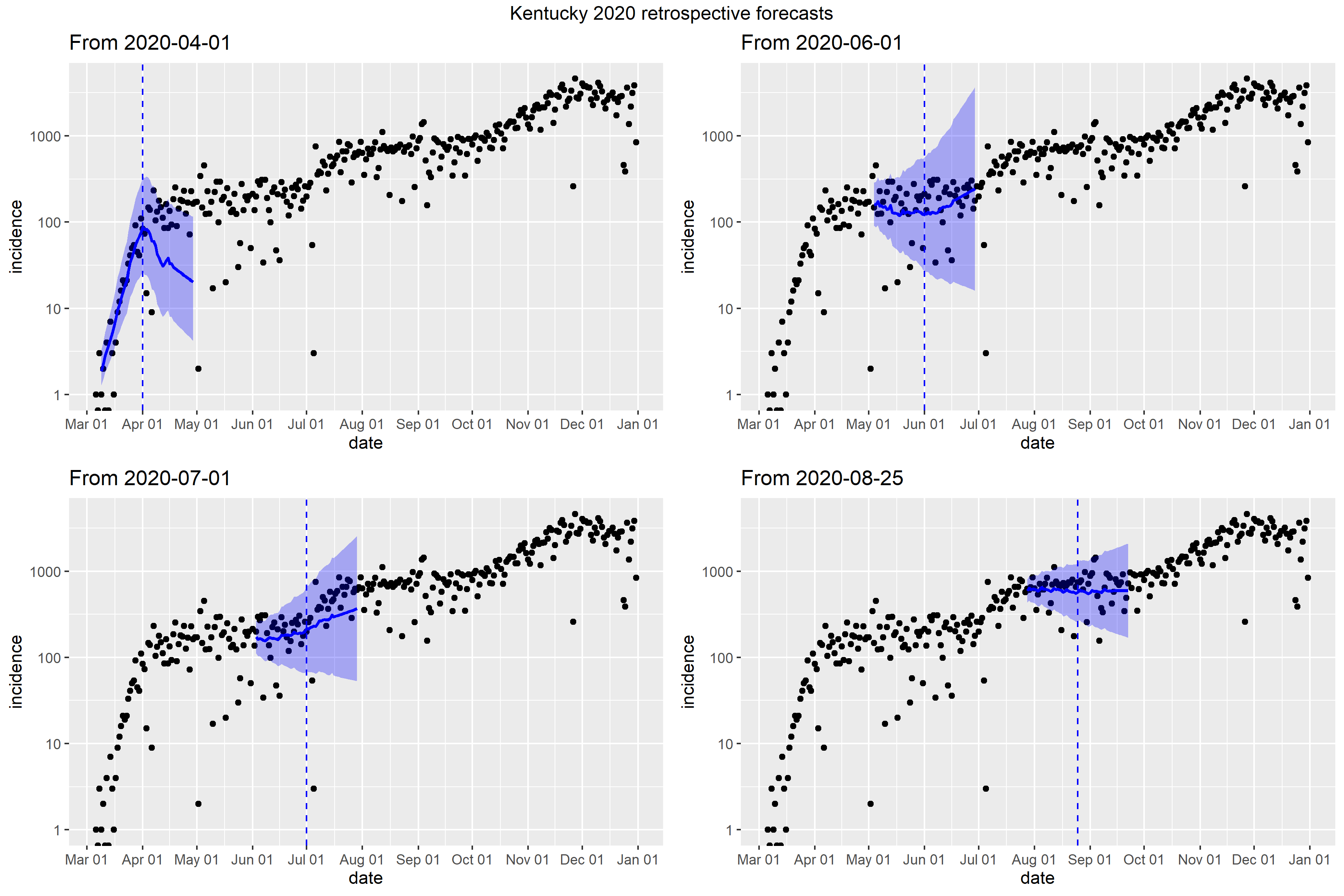

### Louisiana_Peff_plot.png

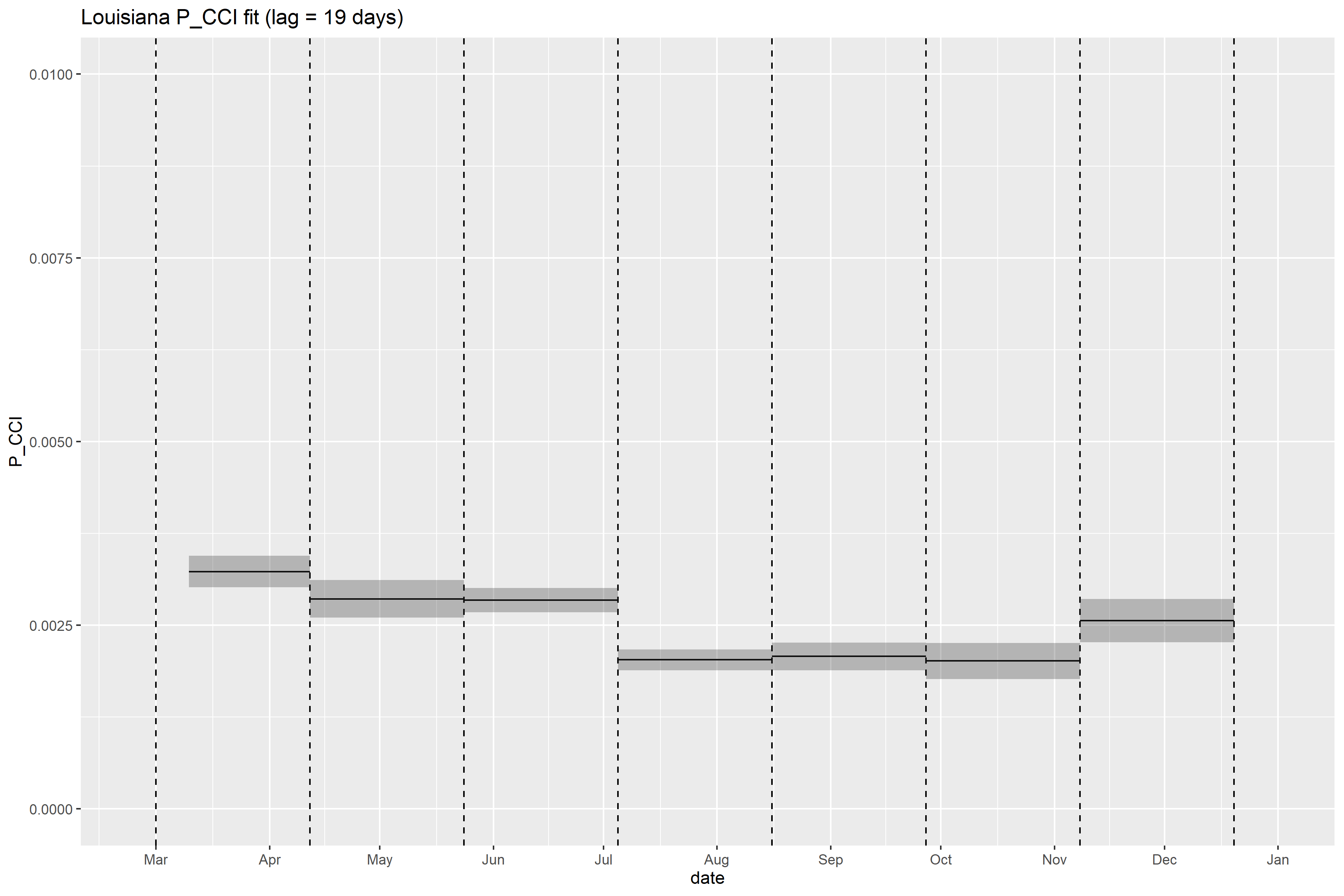

### Louisiana_retrospective_forecasts.png

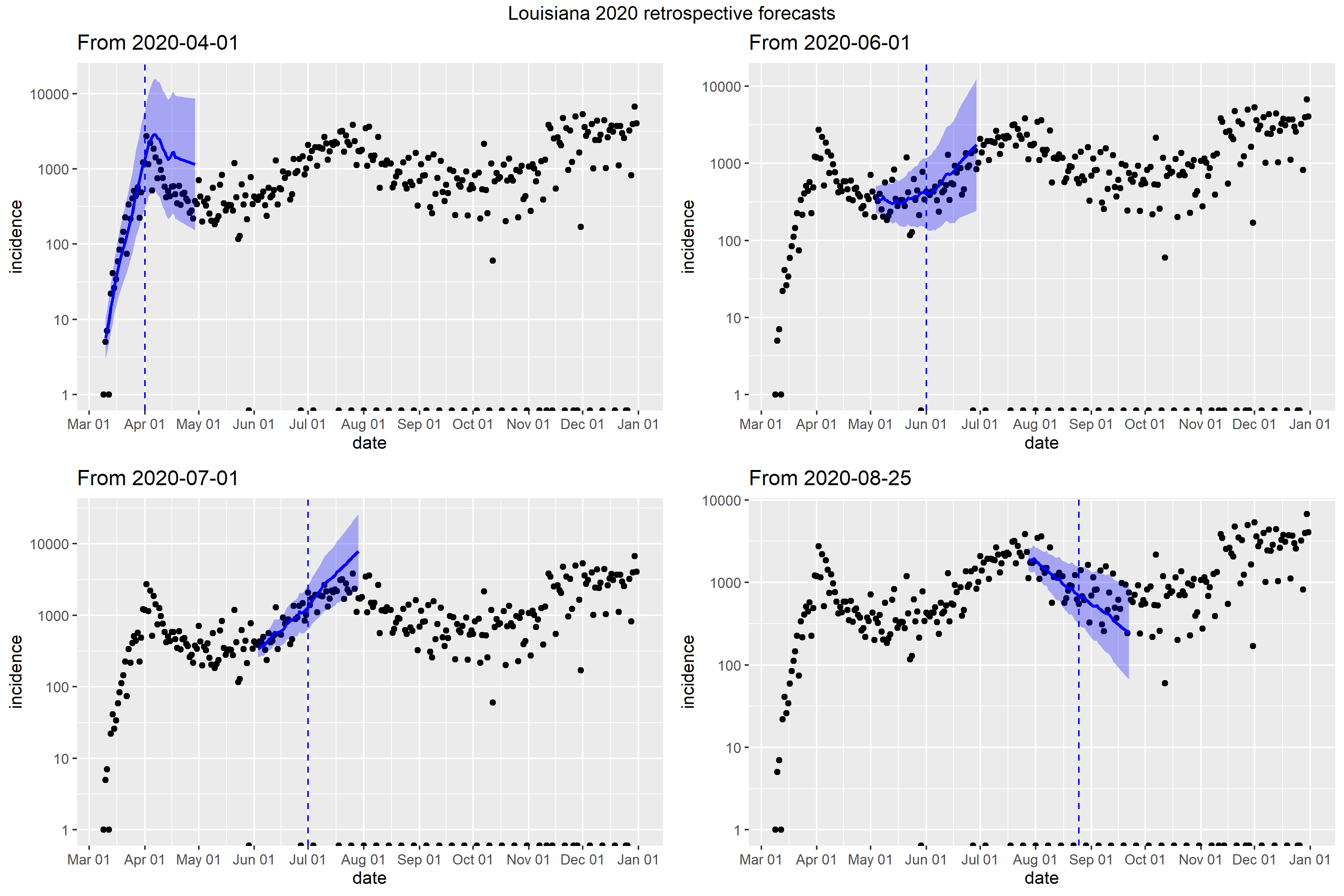

### Maine_full_fit_plot.png

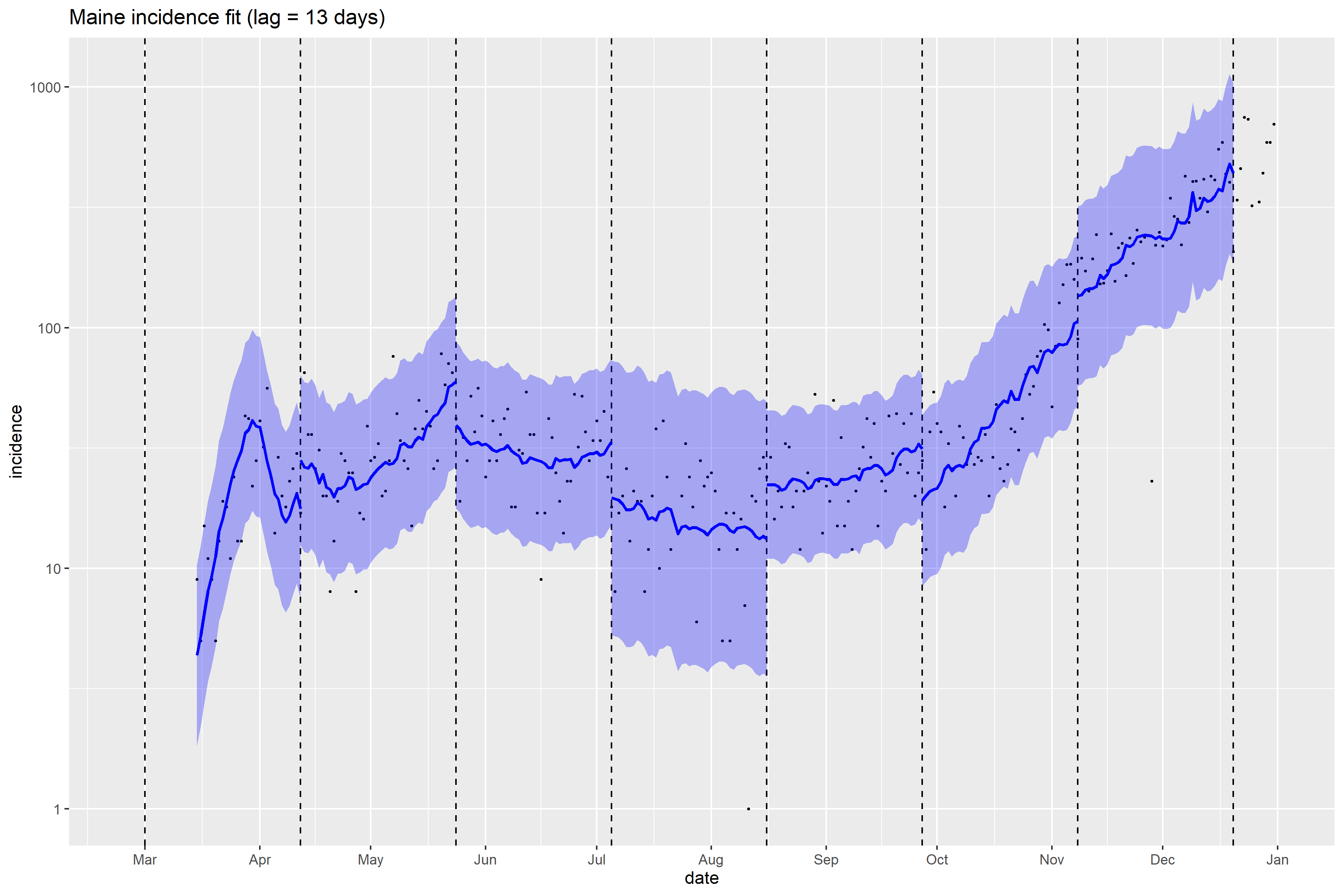

### Maine_Peff_plot.png

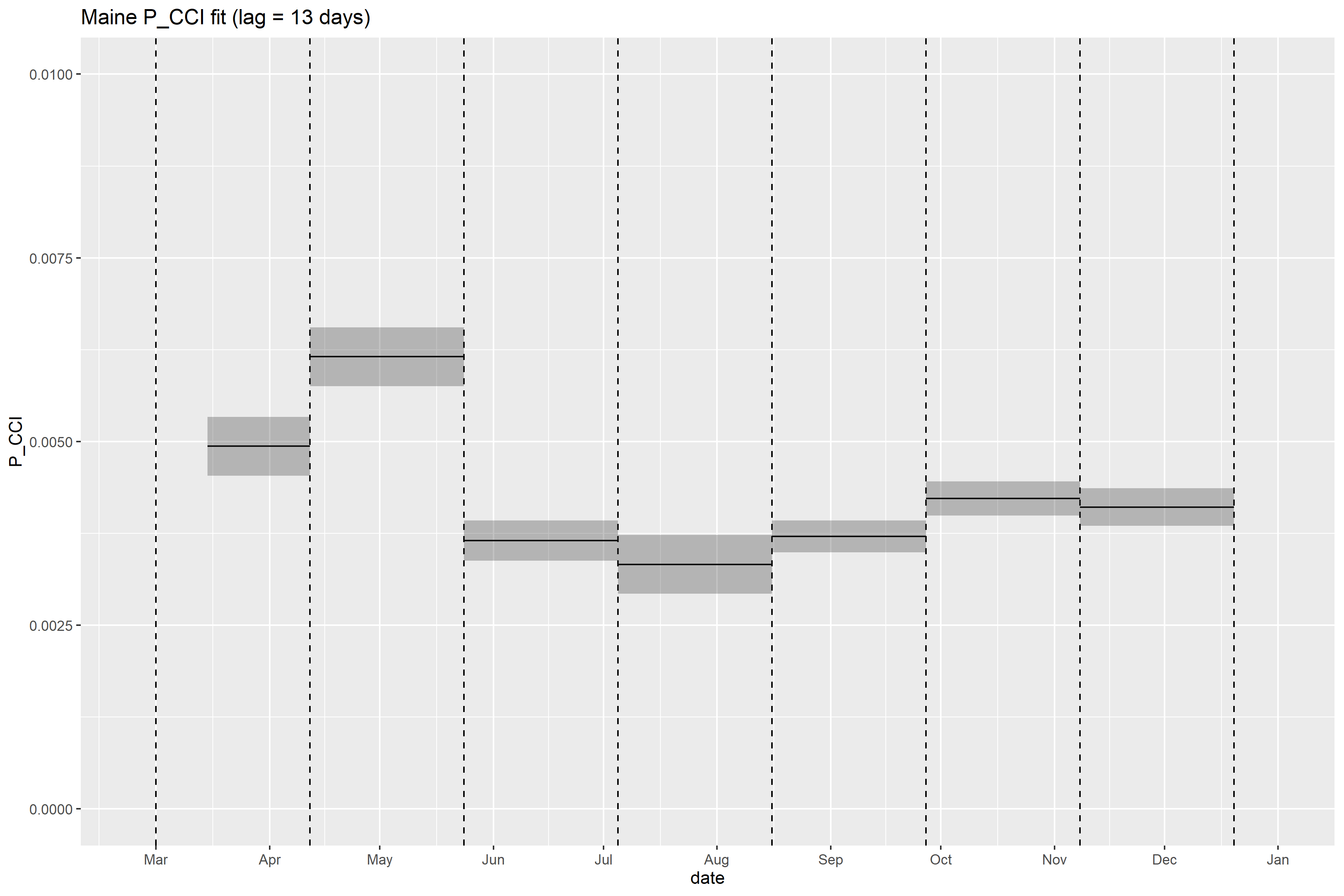

### Maine_retrospective_forecasts.png

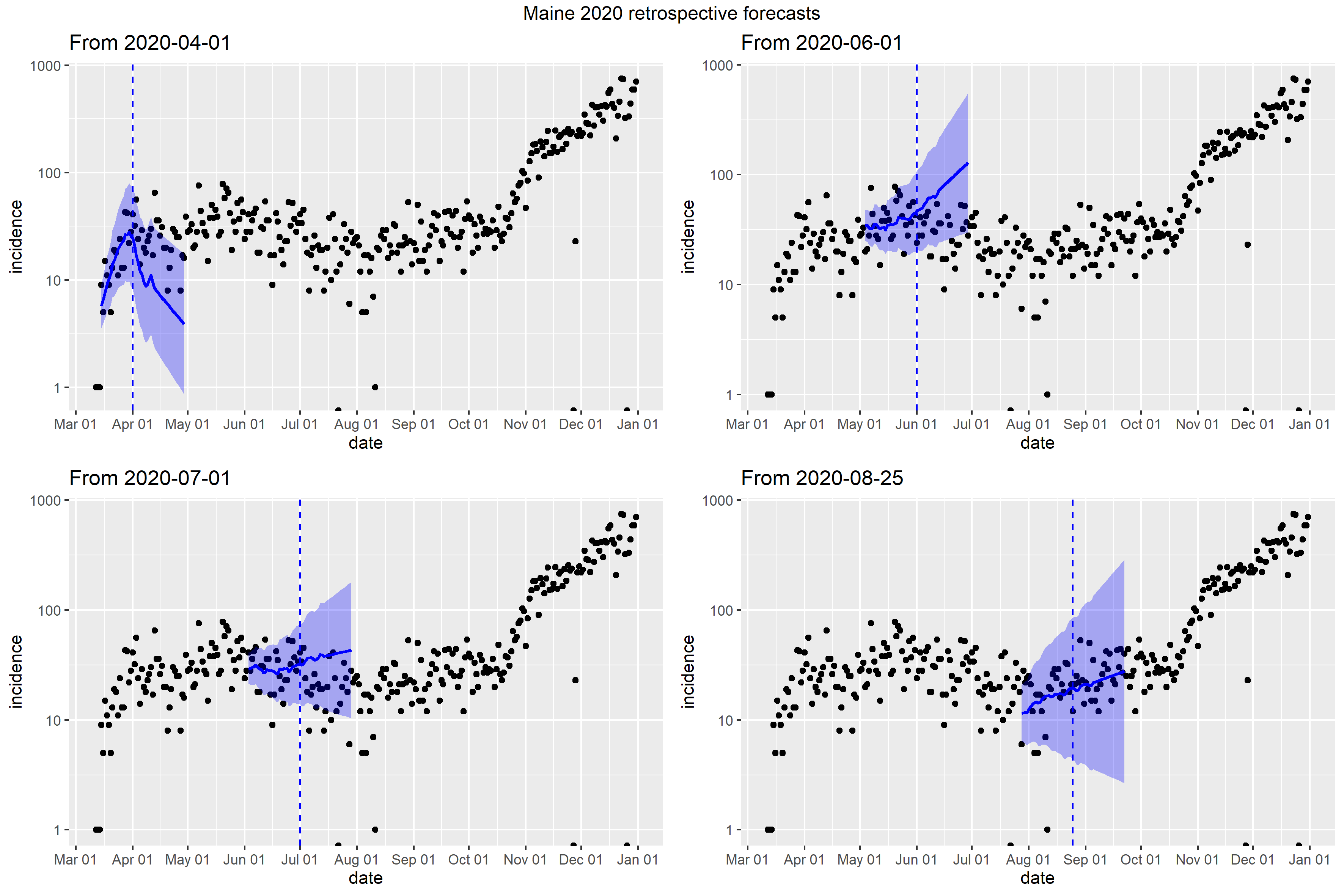

### Maryland_Peff_plot.png

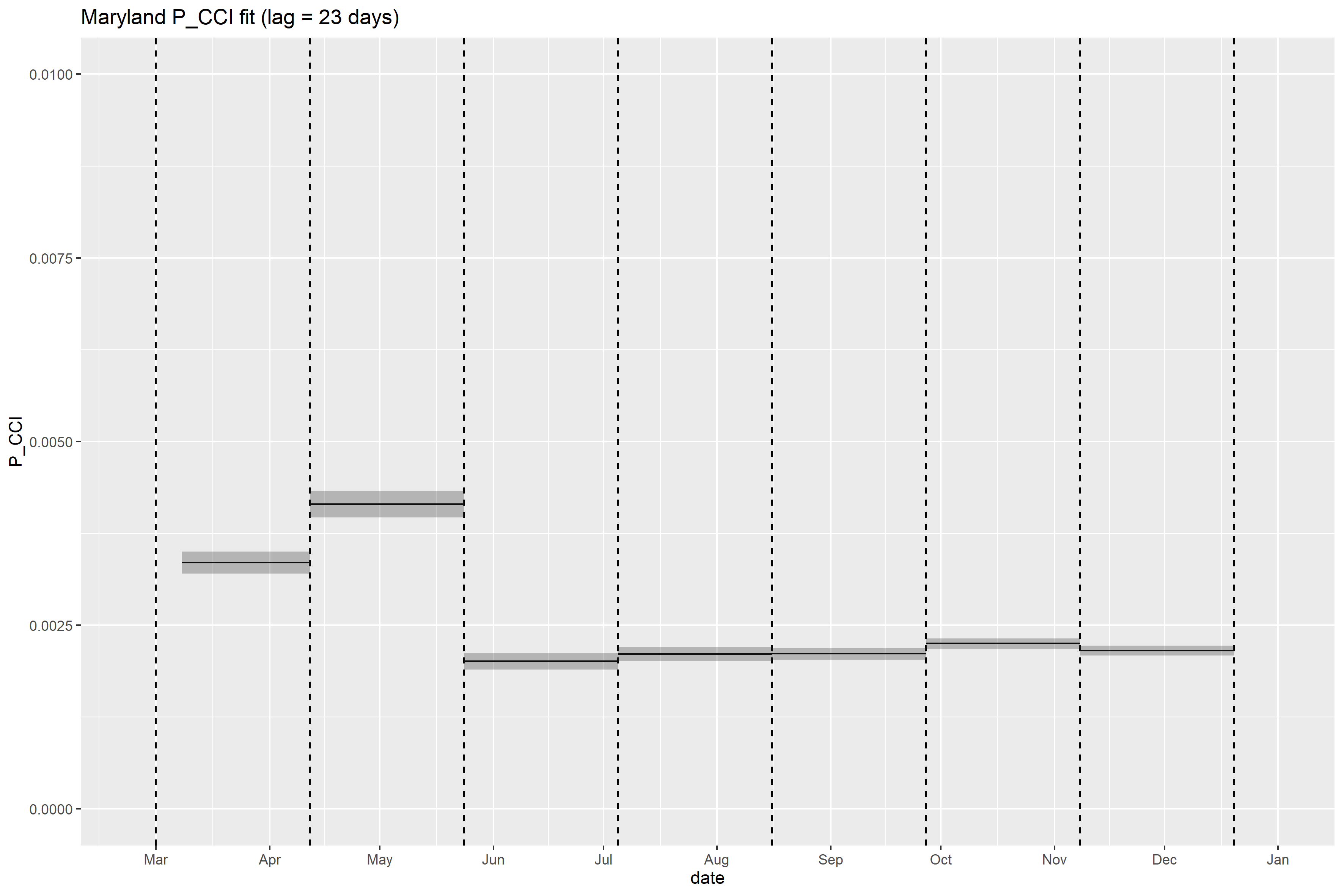

### Maryland_retrospective_forecasts.png

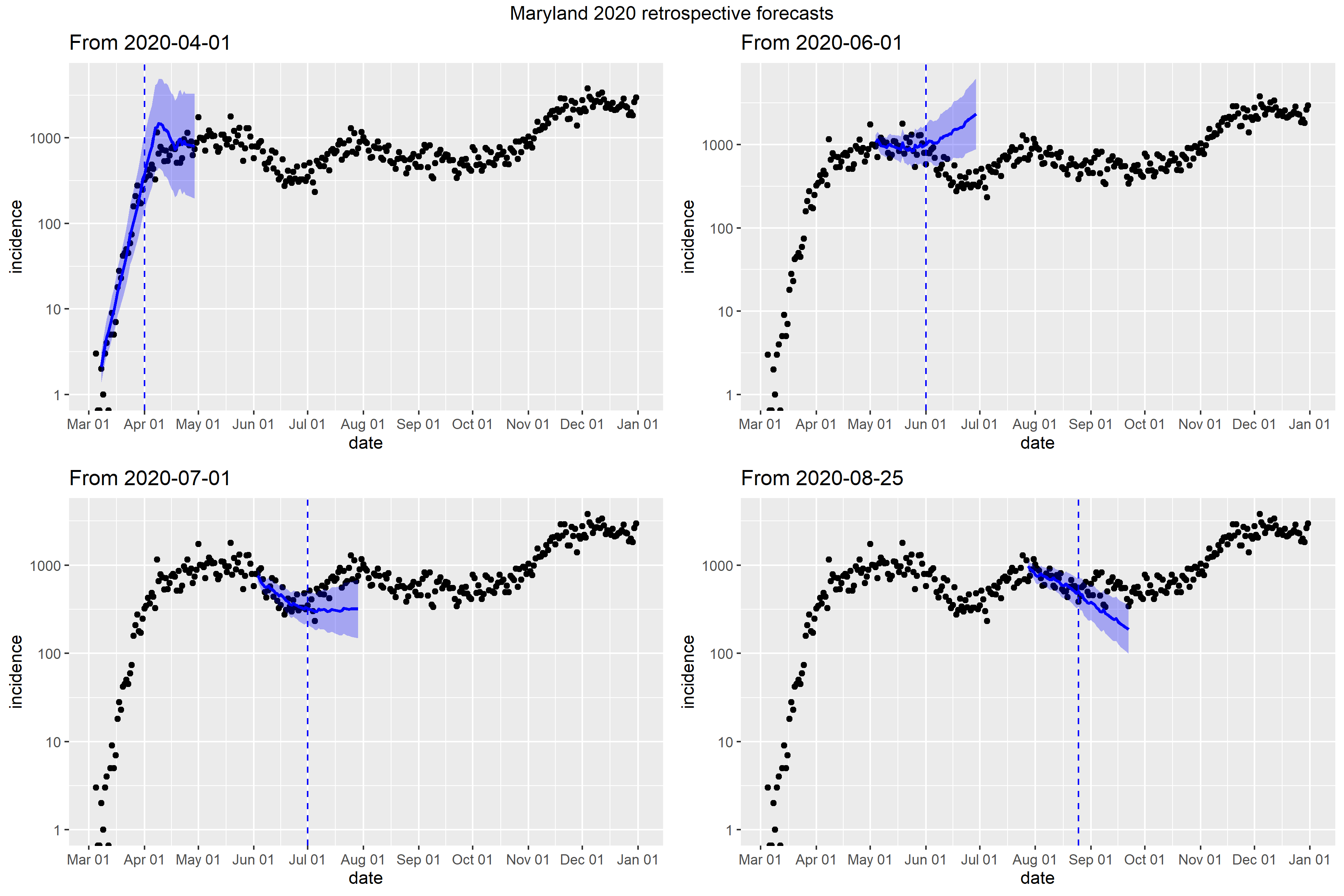

### Massachusetts_retrospective_forecasts.png

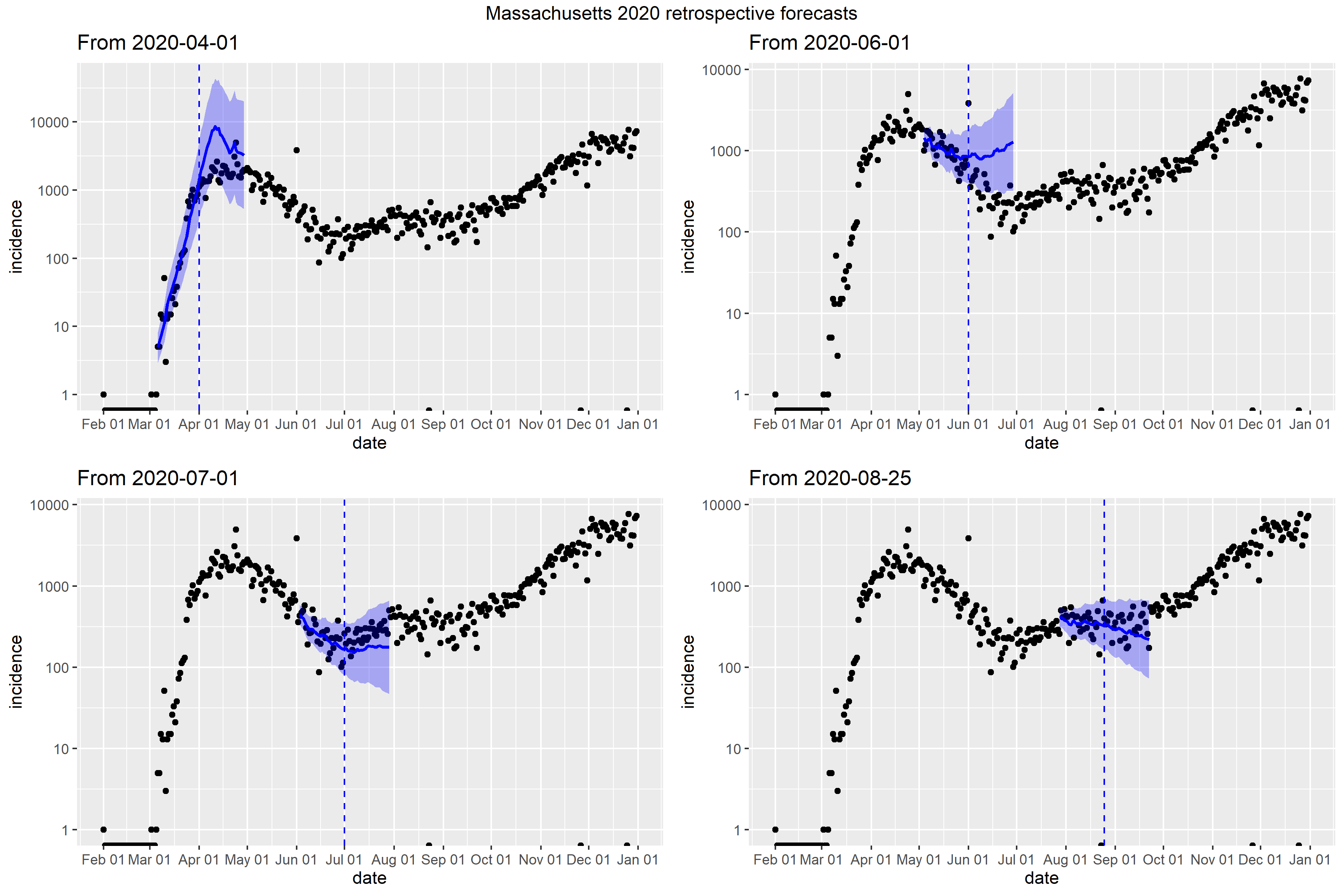

### Michigan_Peff_plot.png

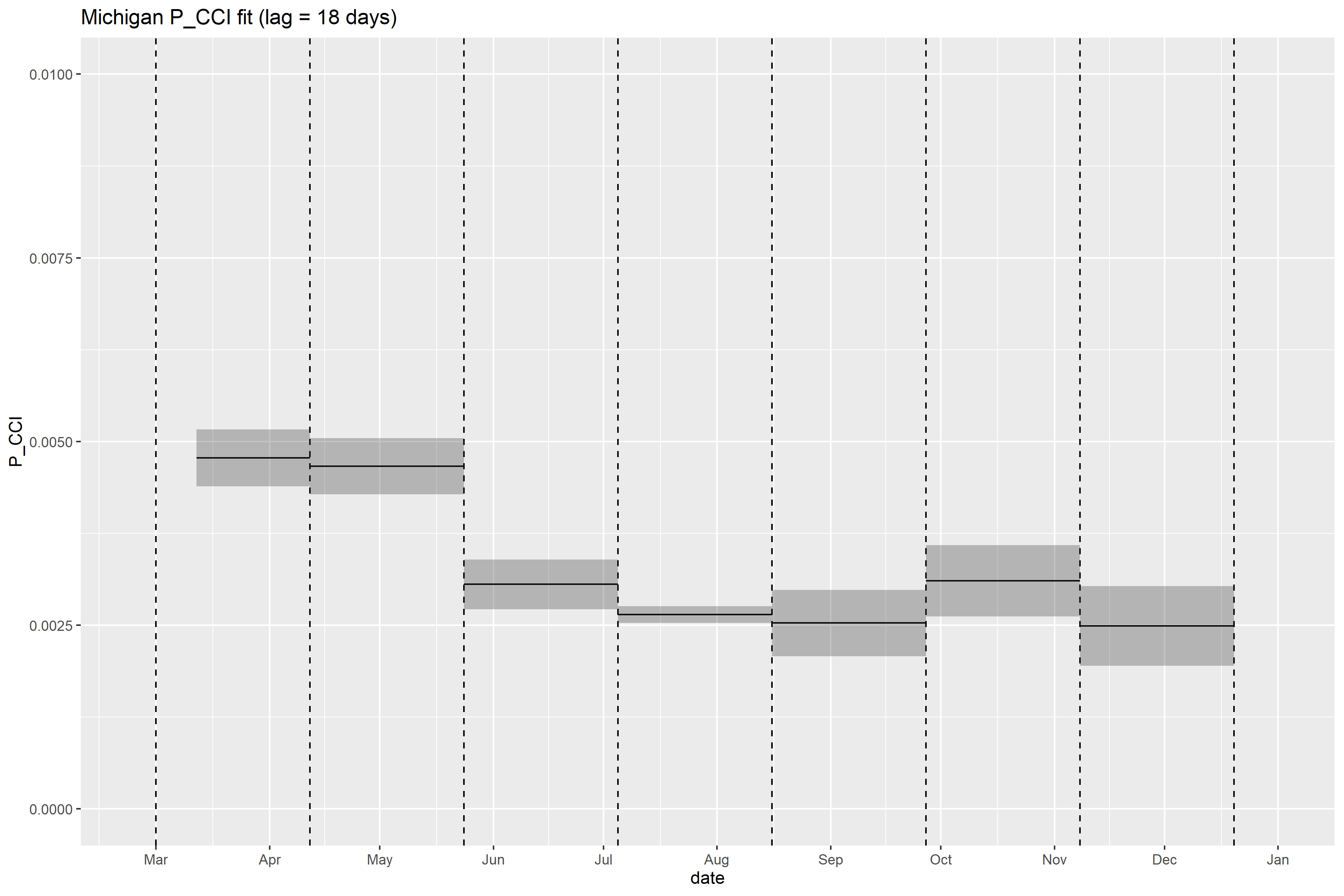

### Michigan_retrospective_forecasts.png

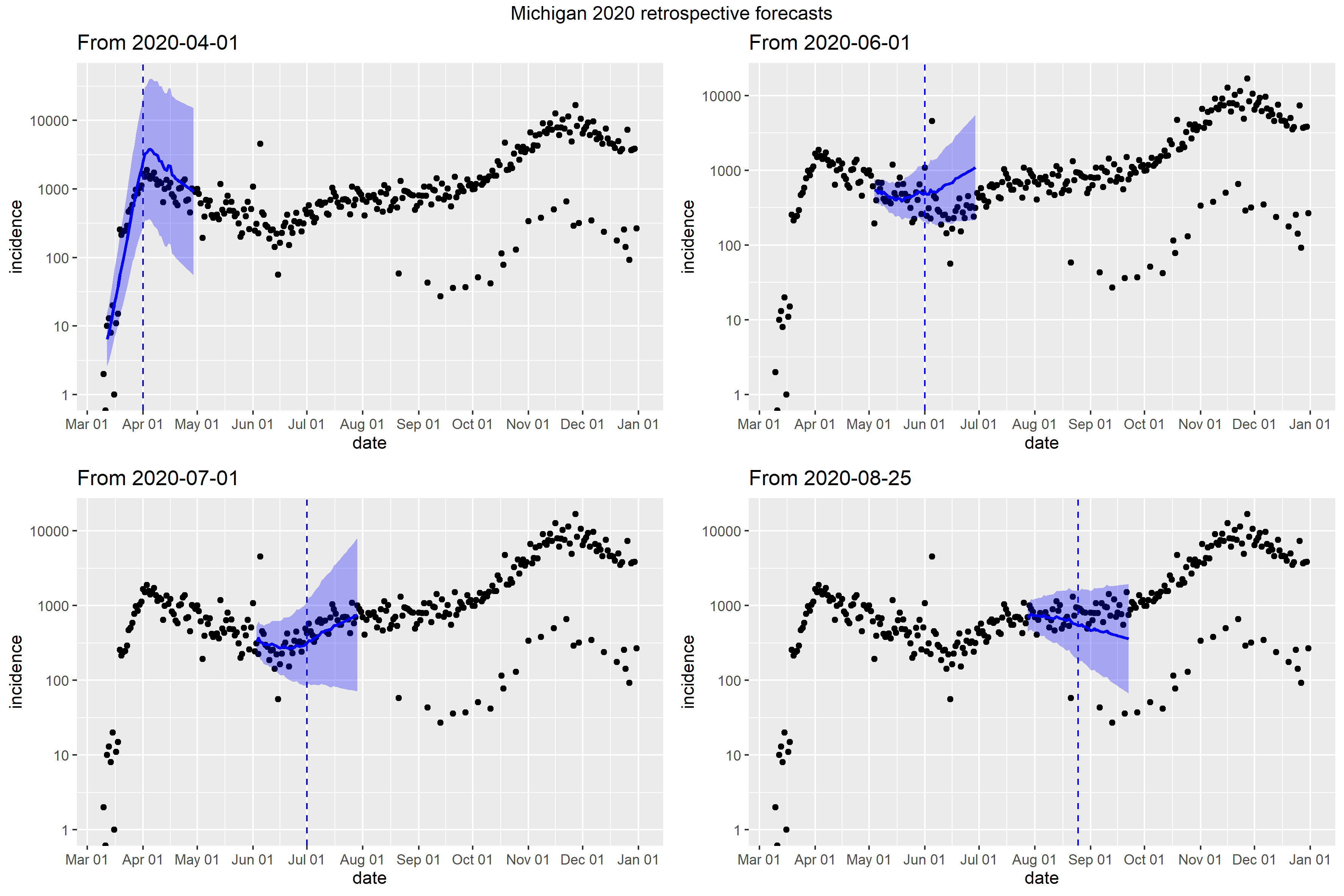

### Minnesota_Peff_plot.png

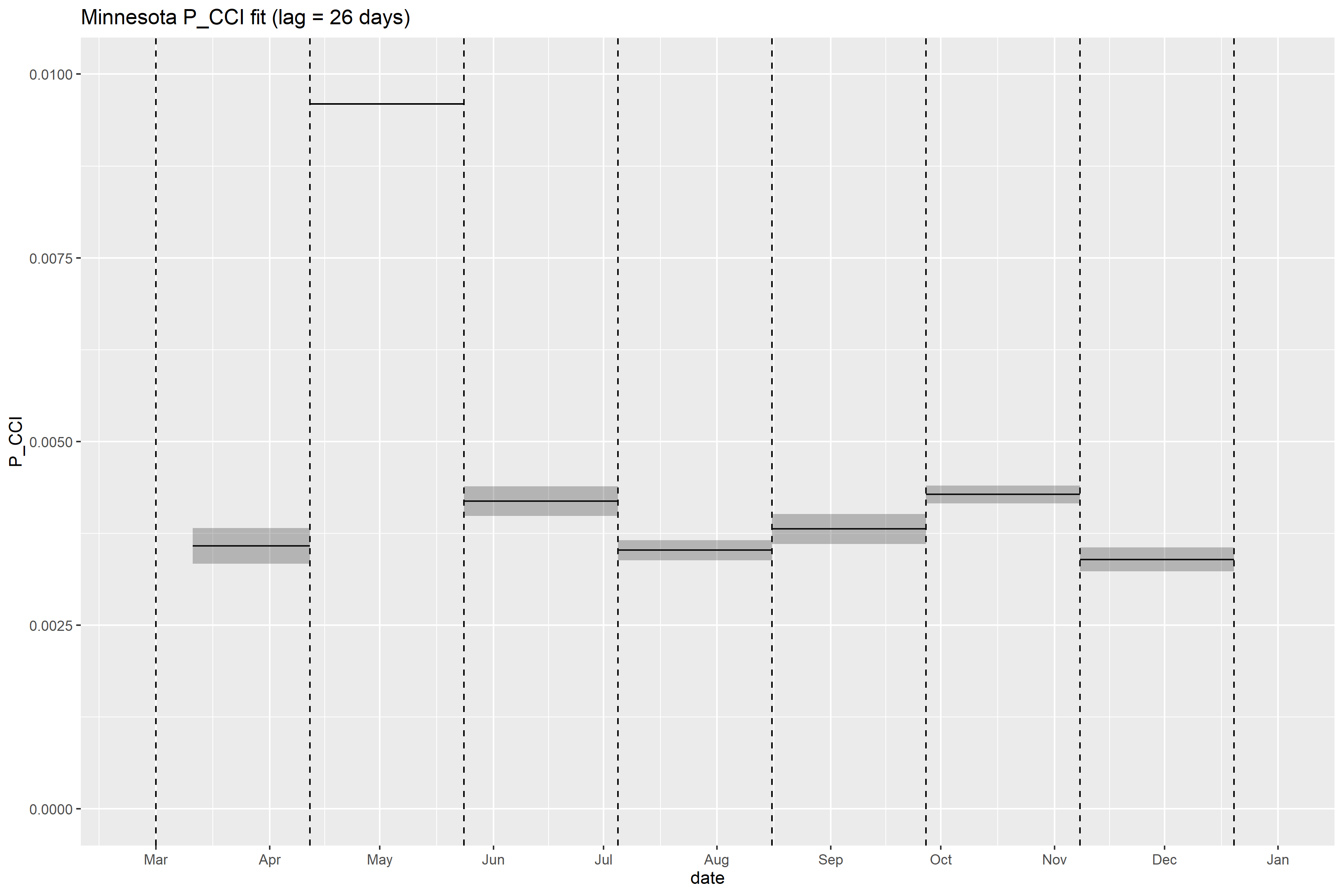

### Minnesota_retrospective_forecasts.png

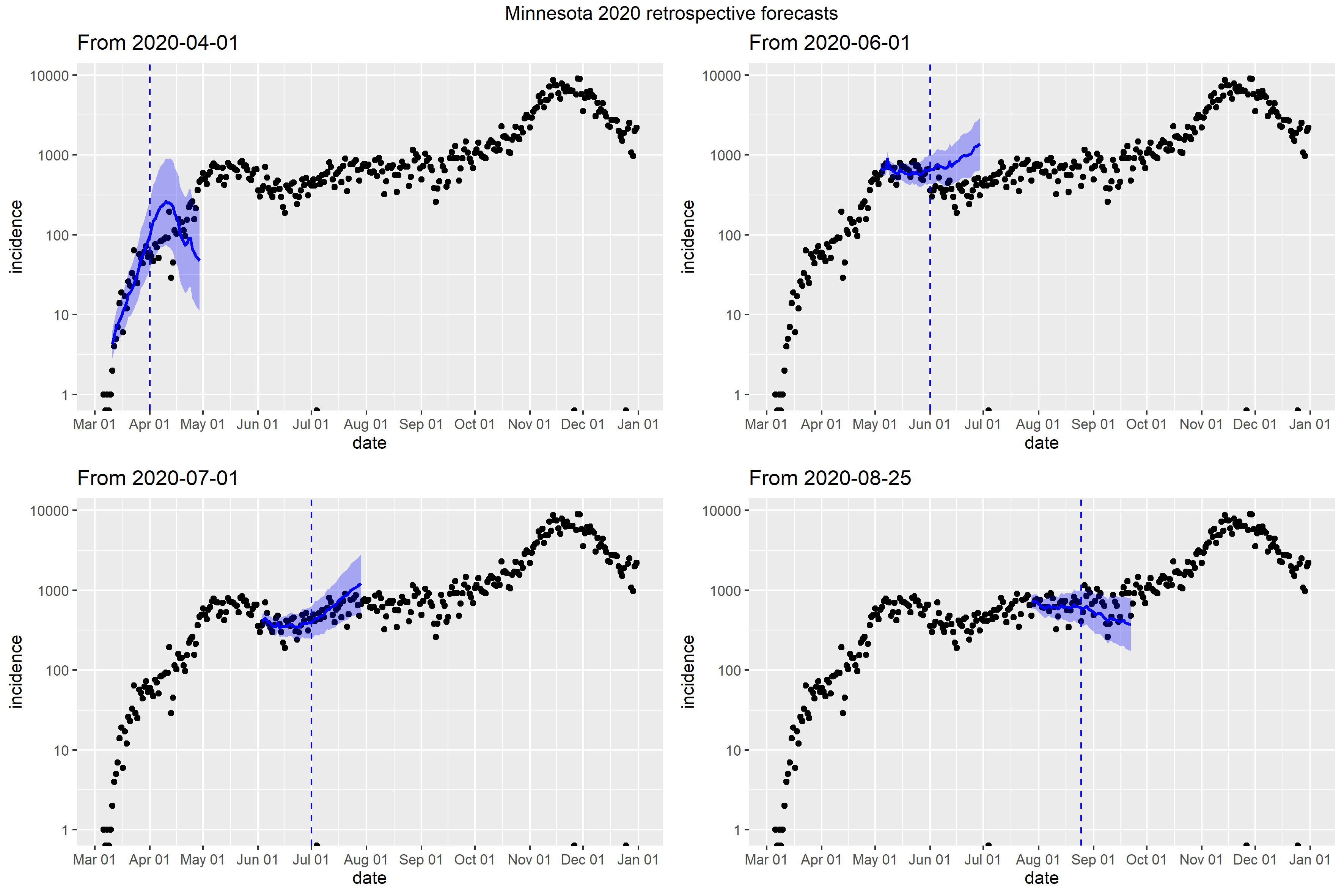

### Mississippi_retrospective_forecasts.png

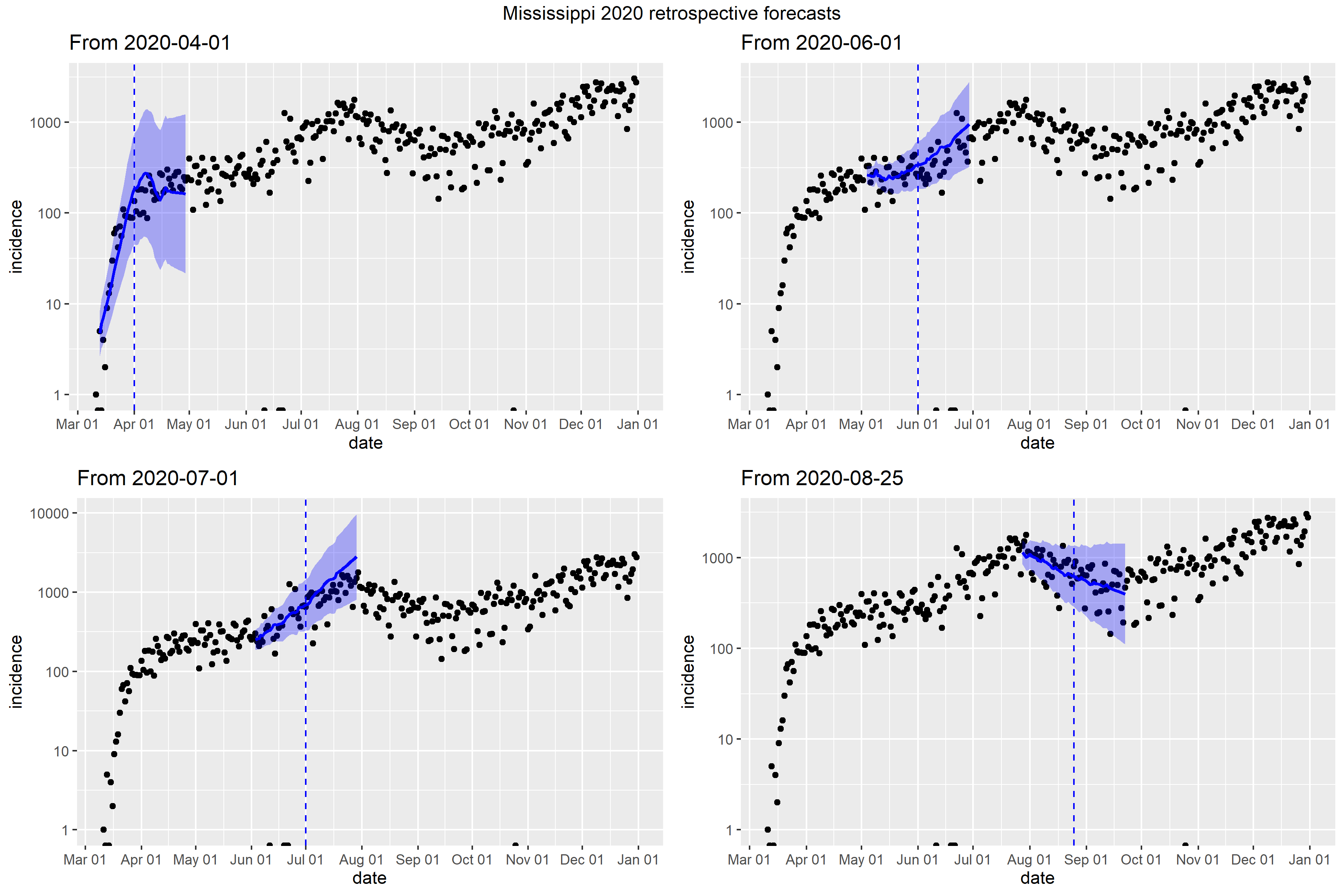

### Missouri_Peff_plot.png

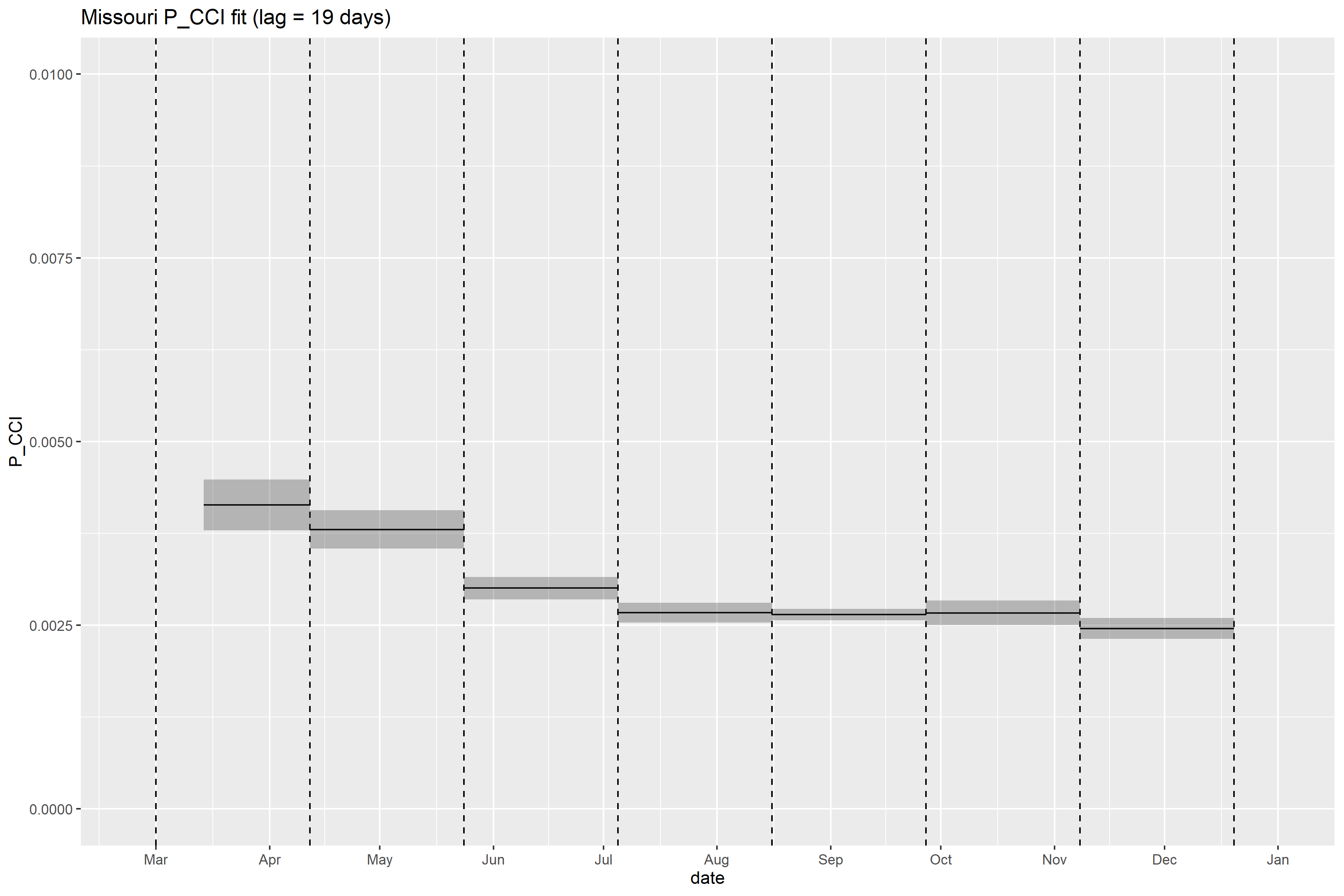

### Missouri_retrospective_forecasts.png

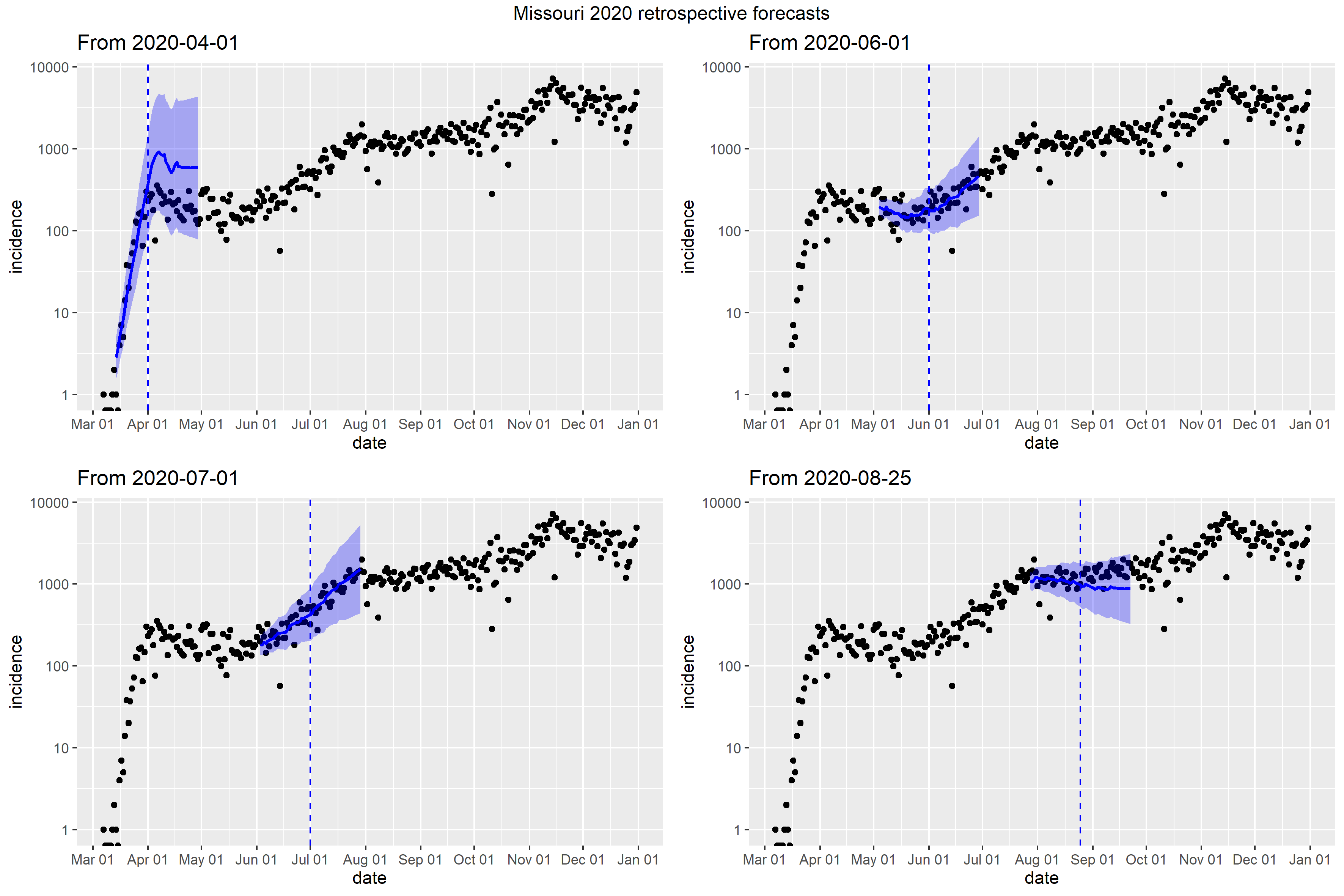

### Montana_Peff_plot.png

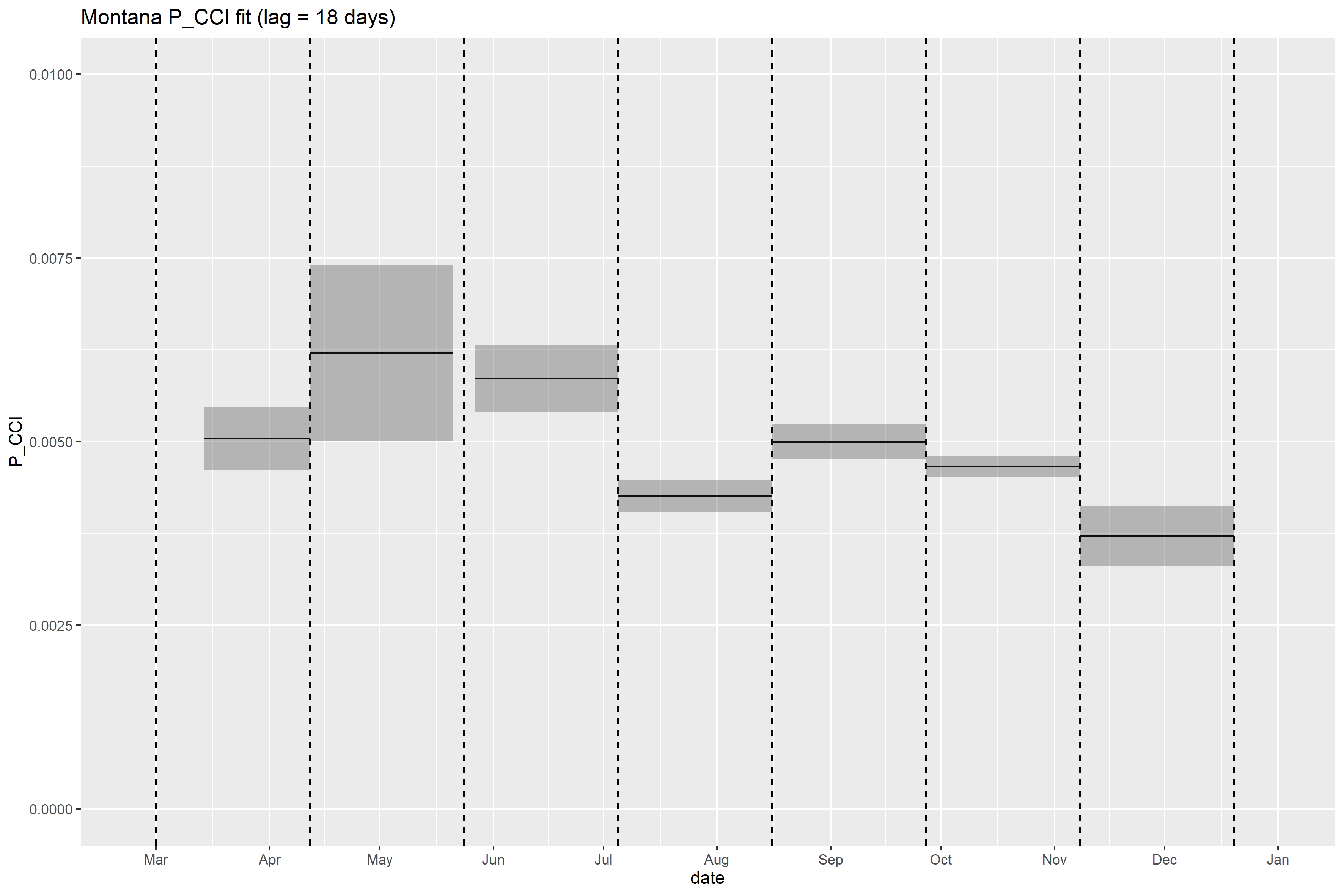

### Montana_retrospective_forecasts.png

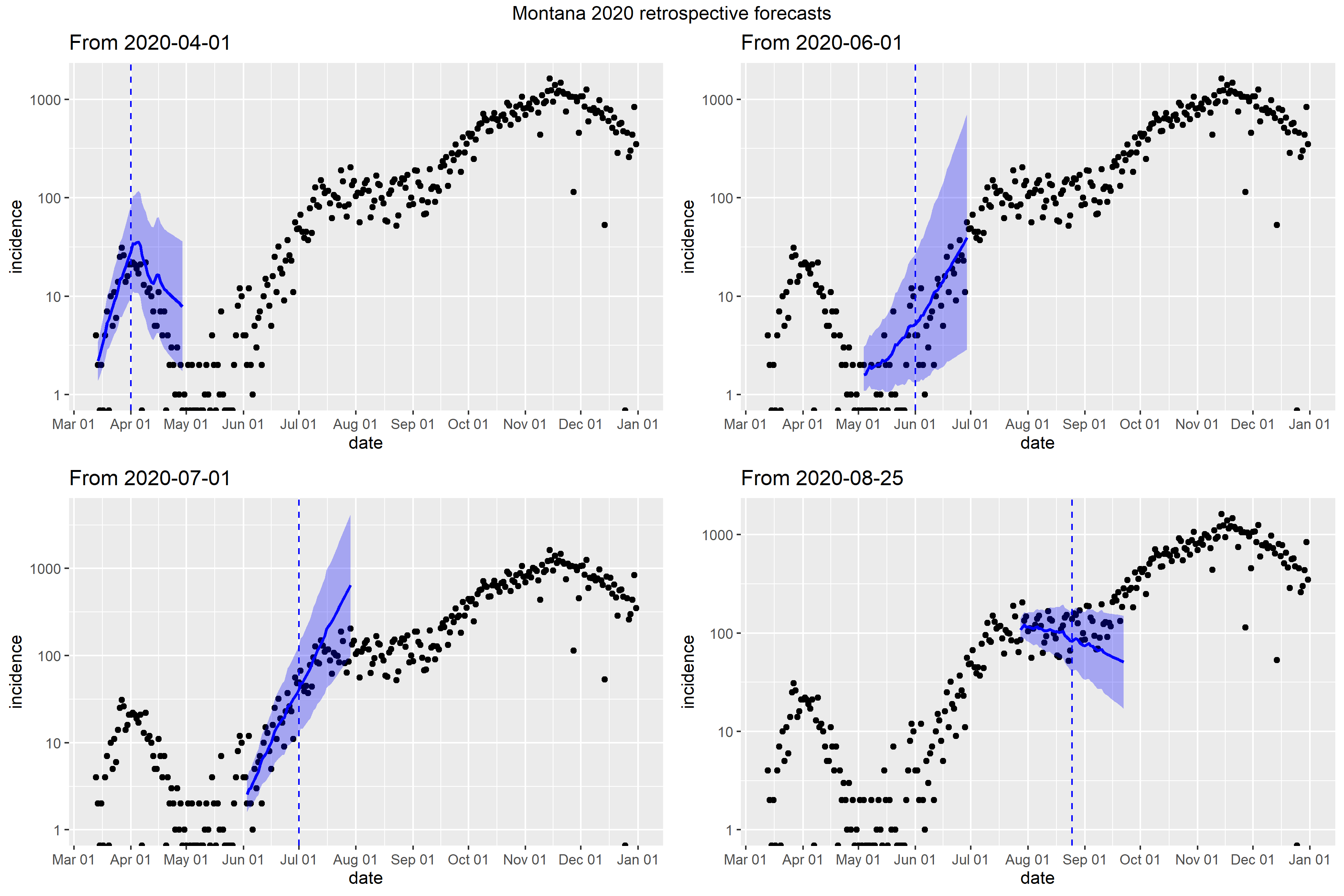

### Nebraska_Peff_plot.png

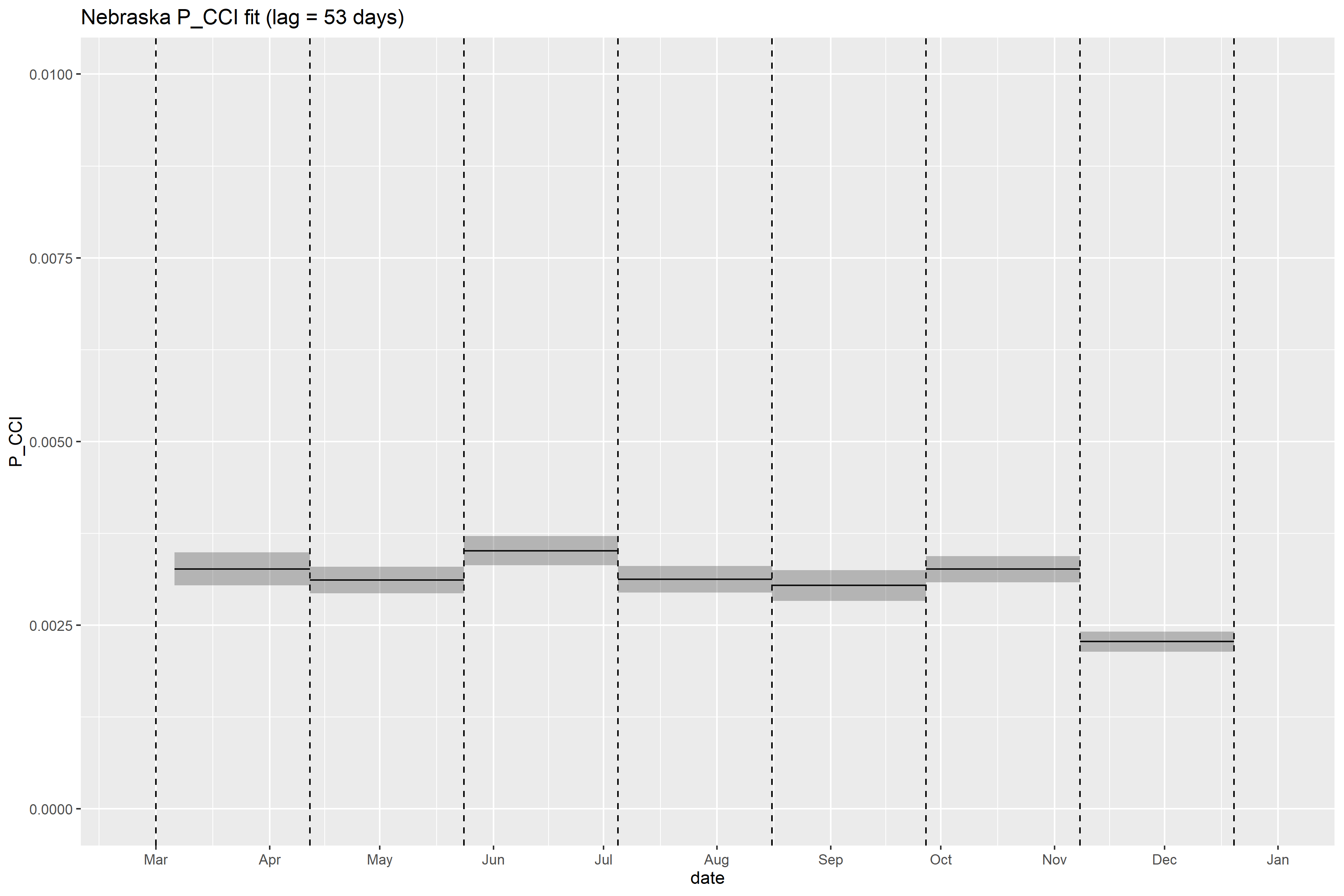

### Nebraska_retrospective_forecasts.png

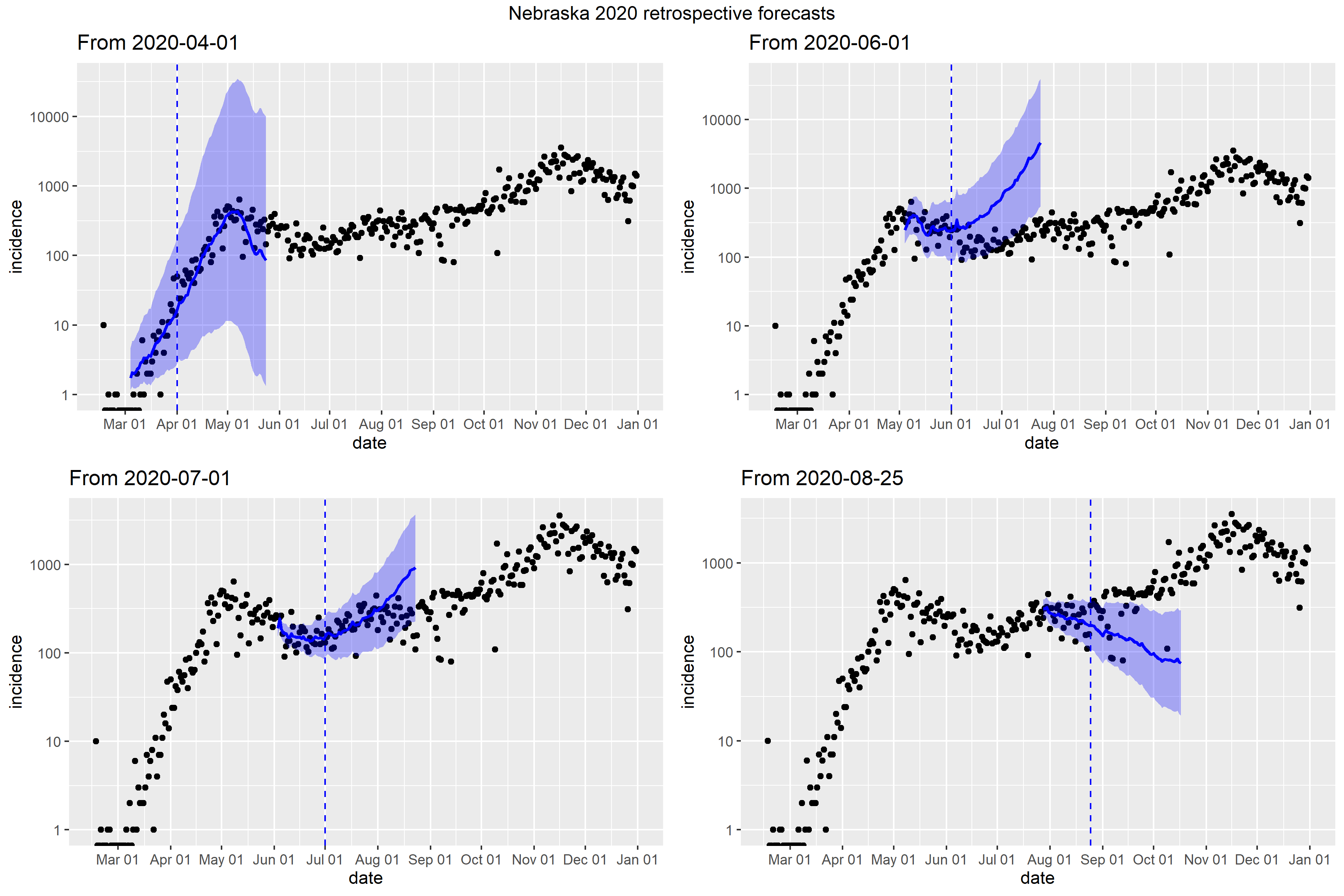

### Nevada_full_fit_plot.png

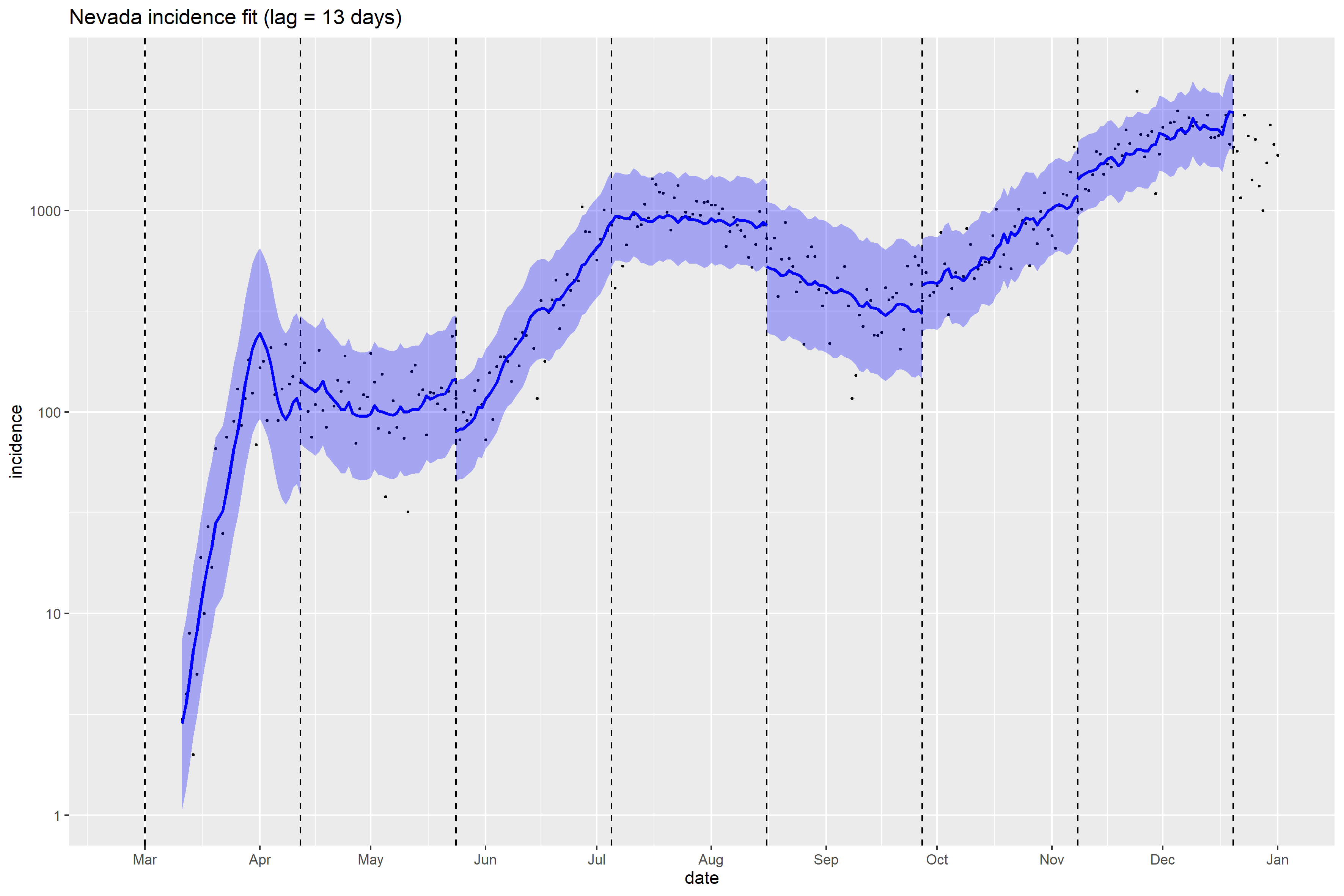

### Nevada_Peff_plot.png

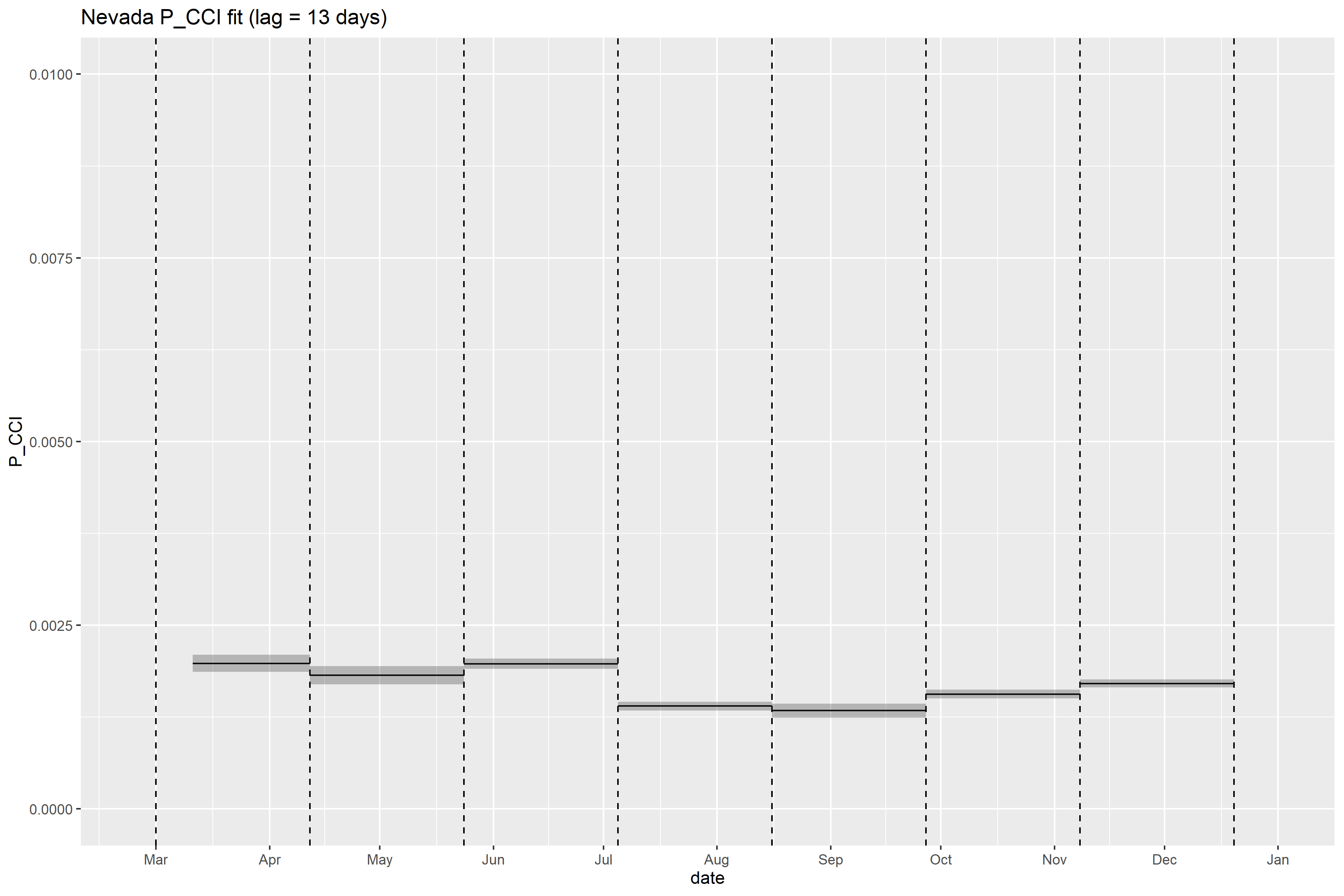

### Nevada_retrospective_forecasts.png

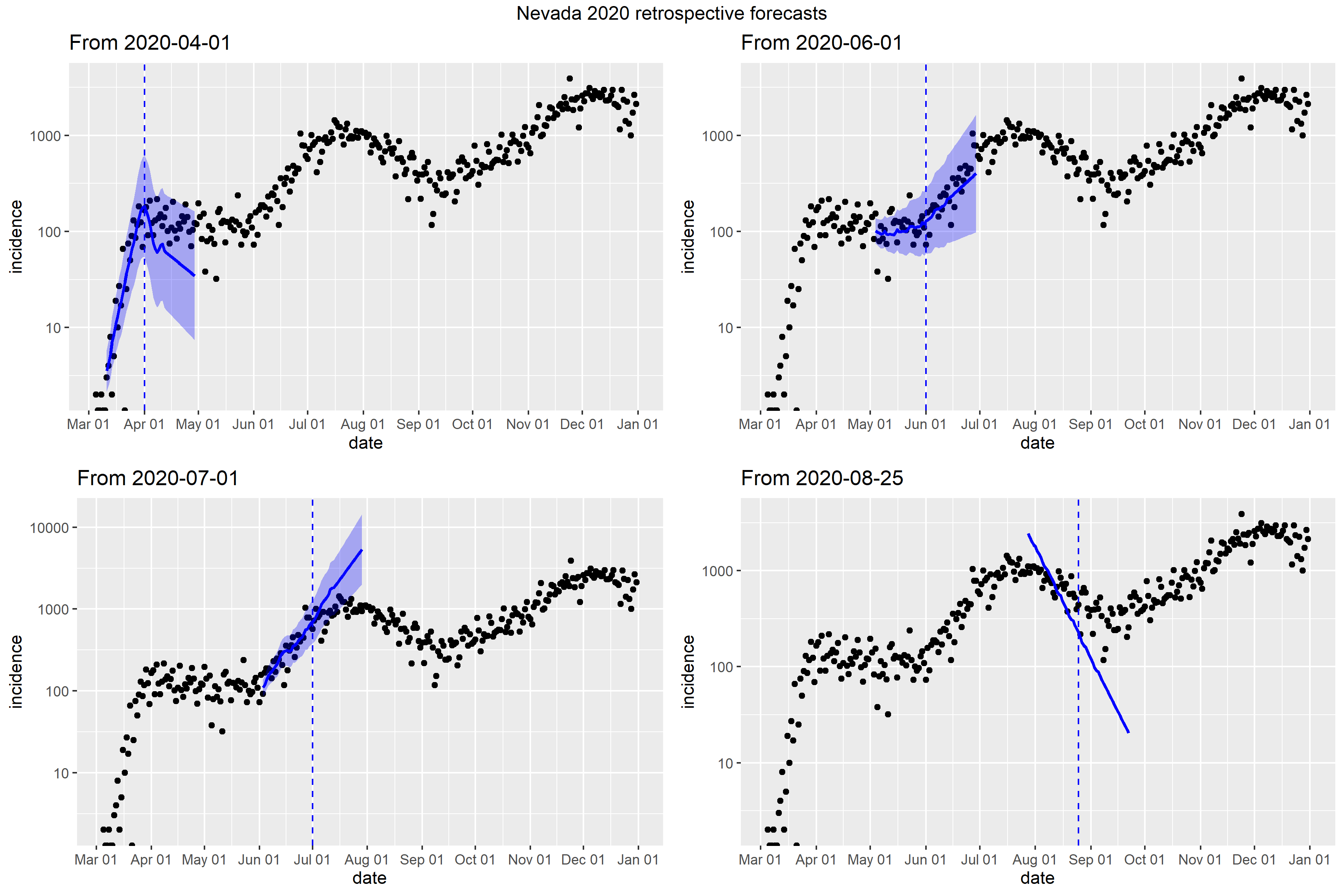

### New Hampshire_retrospective_forecasts.png

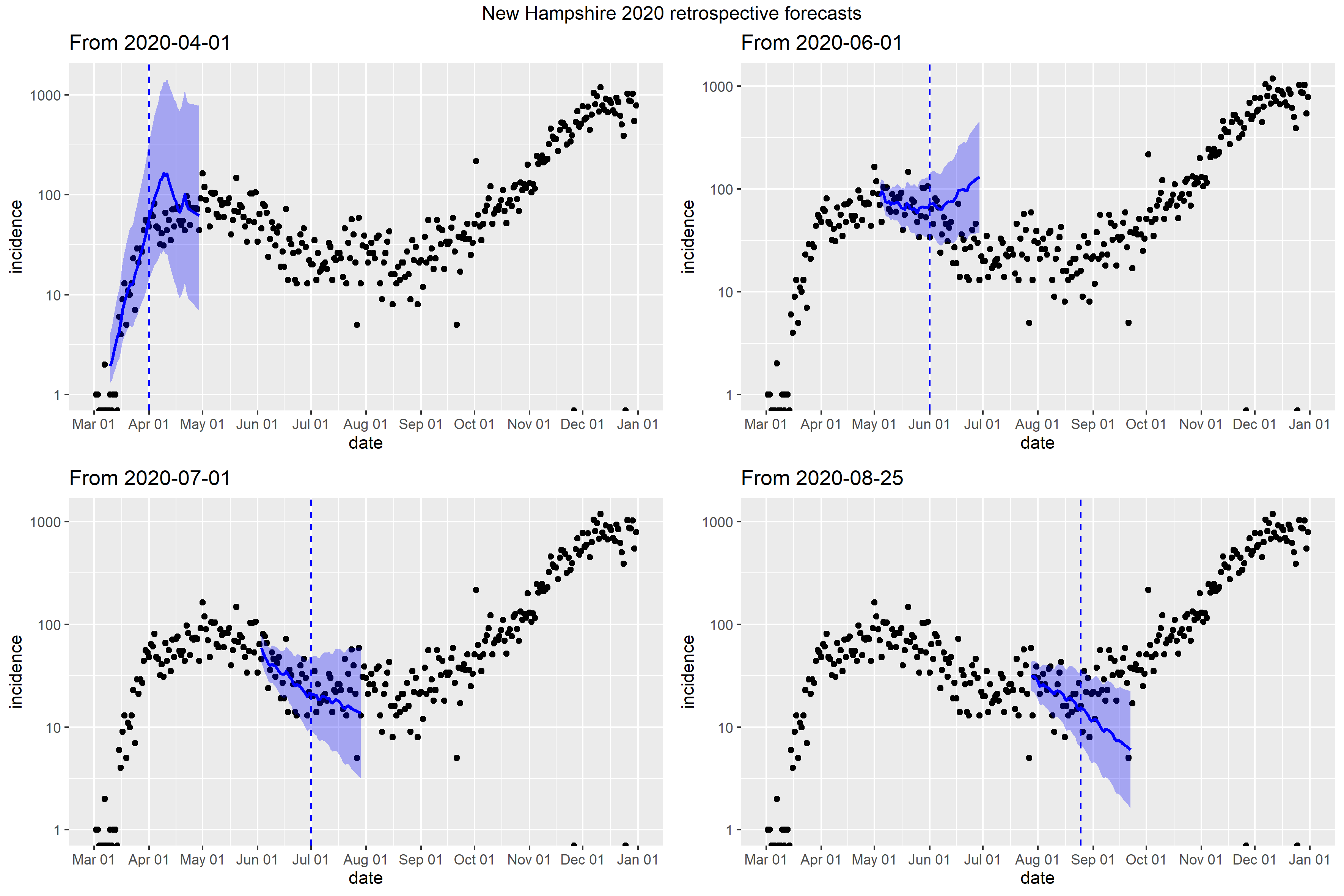

### New Jersey_Peff_plot.png

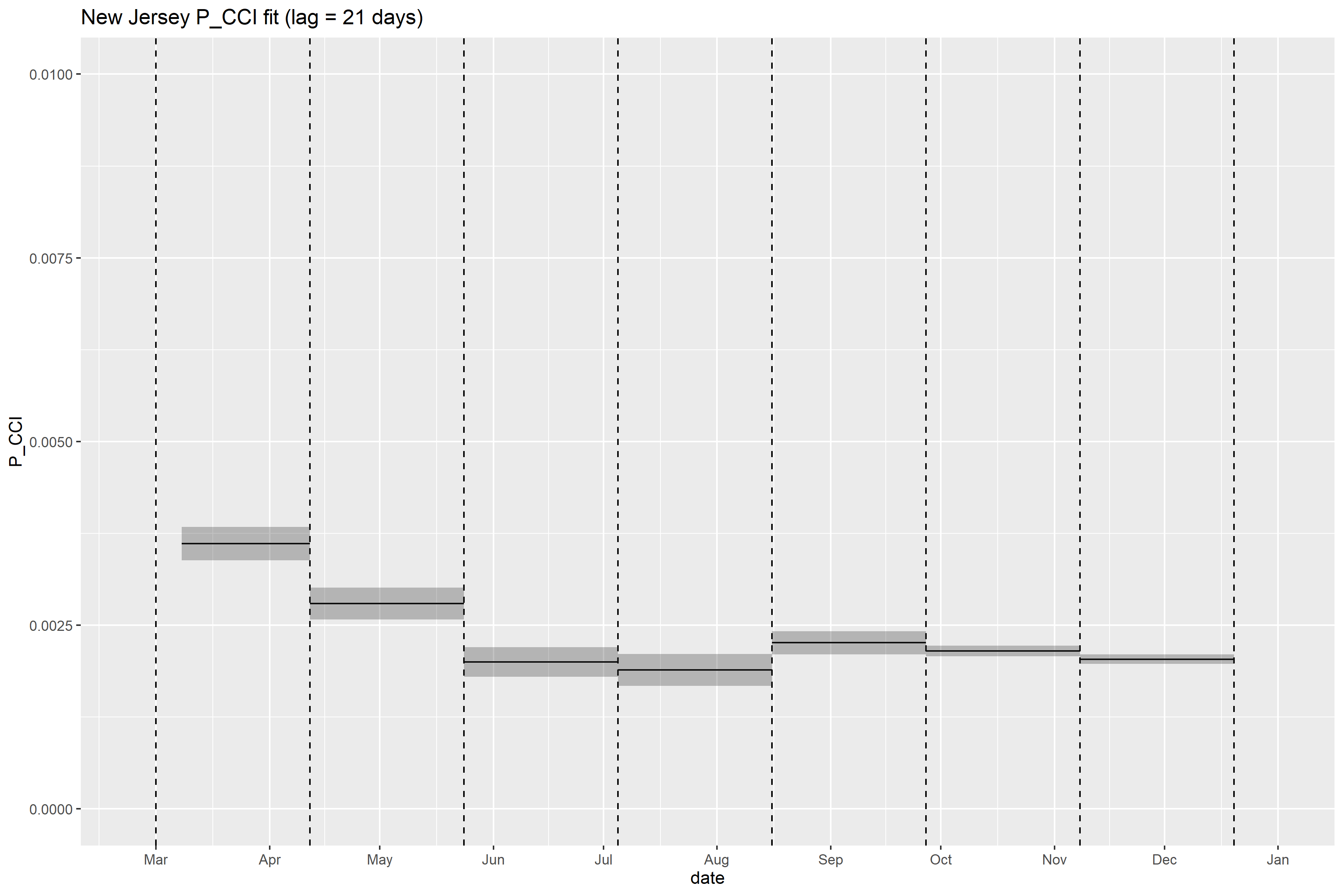

### New Jersey_retrospective_forecasts.png

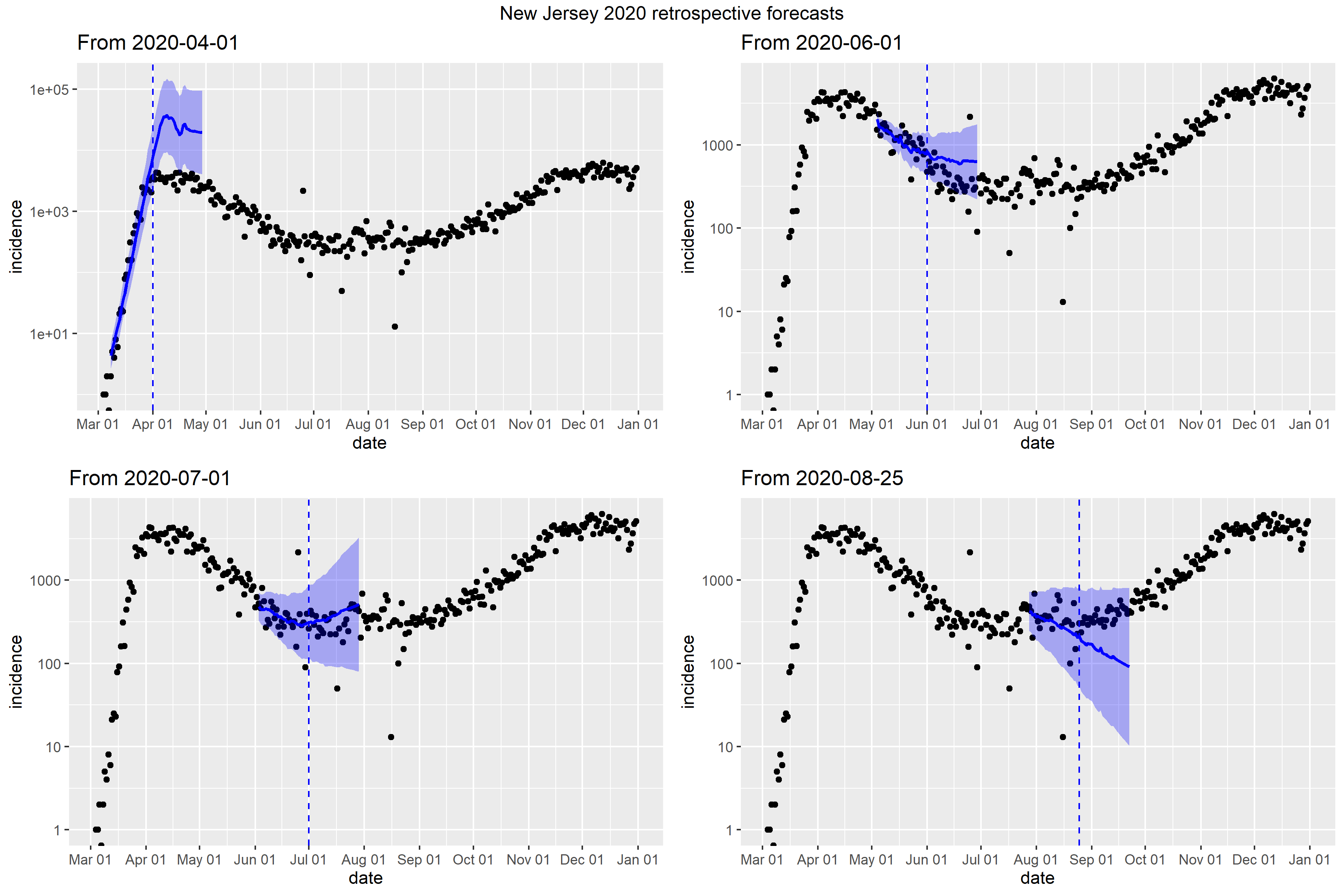

### New Mexico_Peff_plot.png

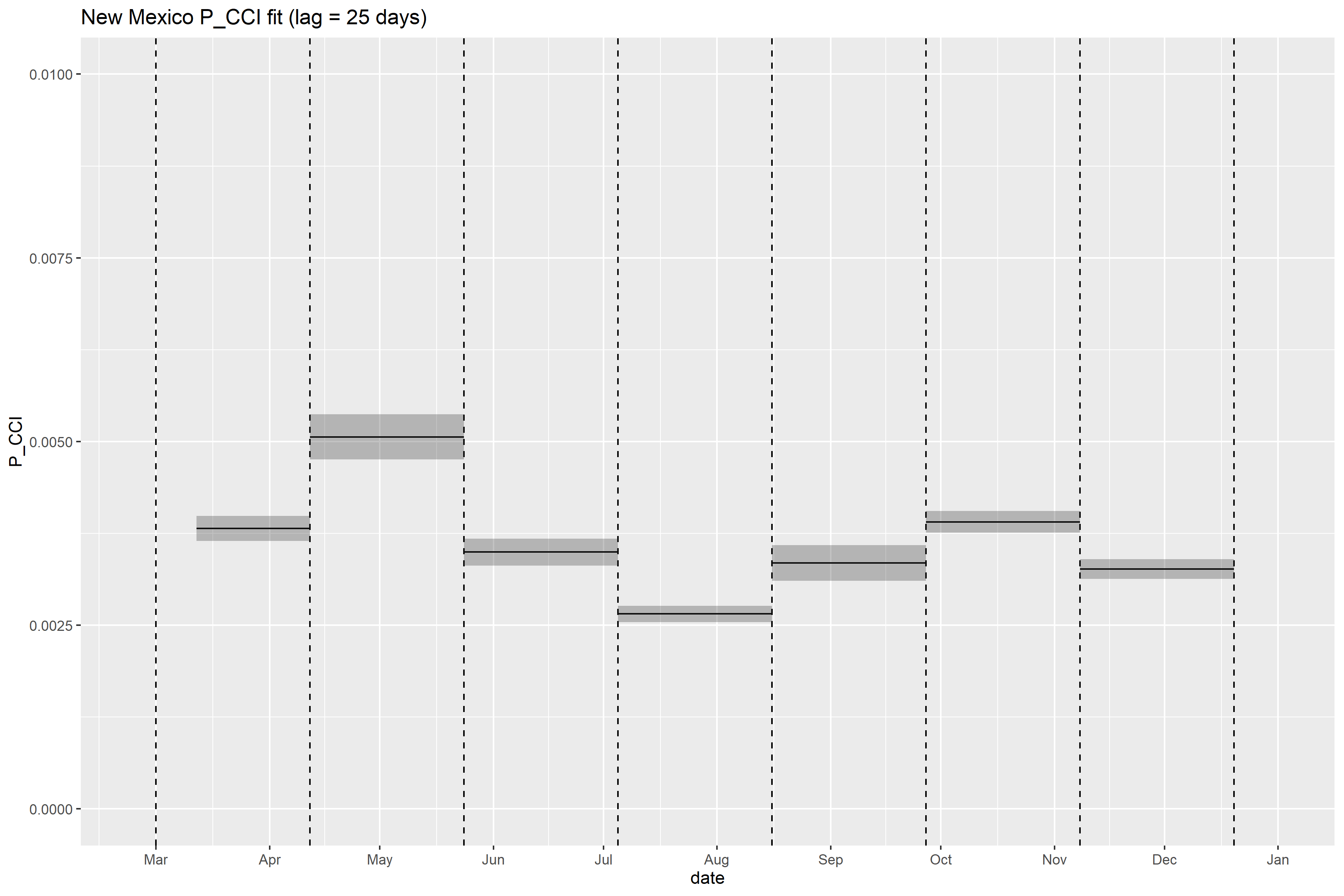

### New Mexico_retrospective_forecasts.png

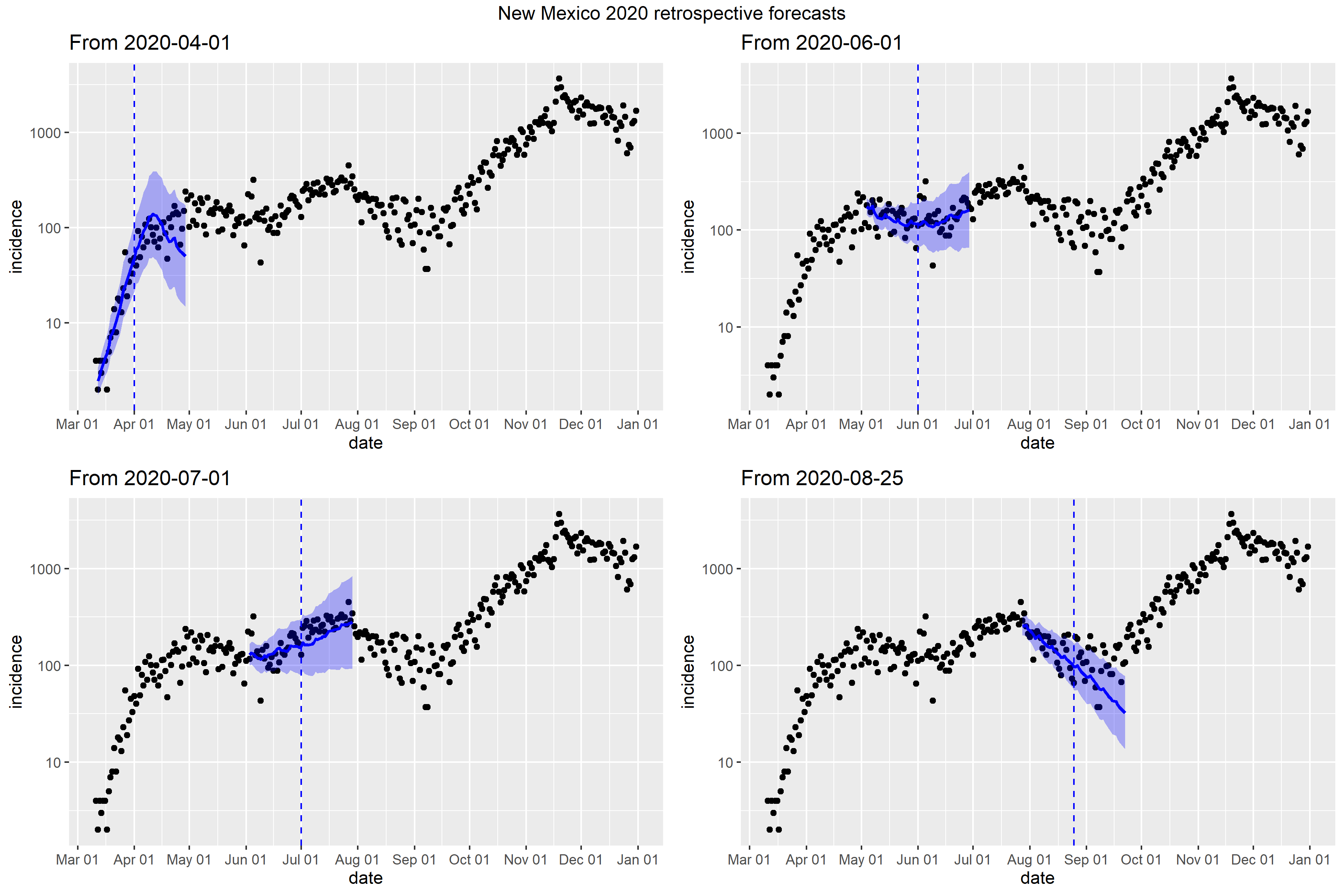

### New York_Peff_plot.png

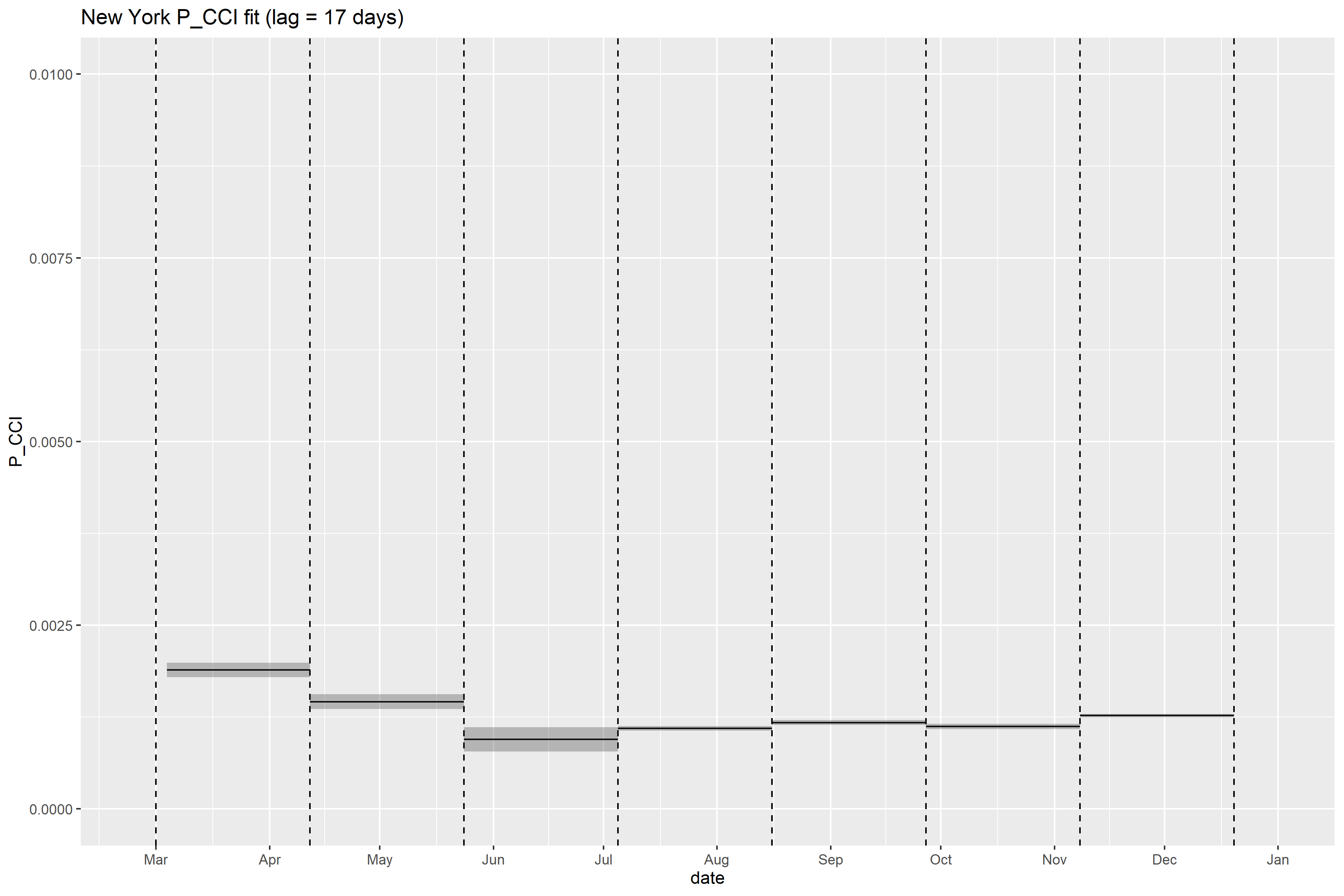

### New York_retrospective_forecasts.png

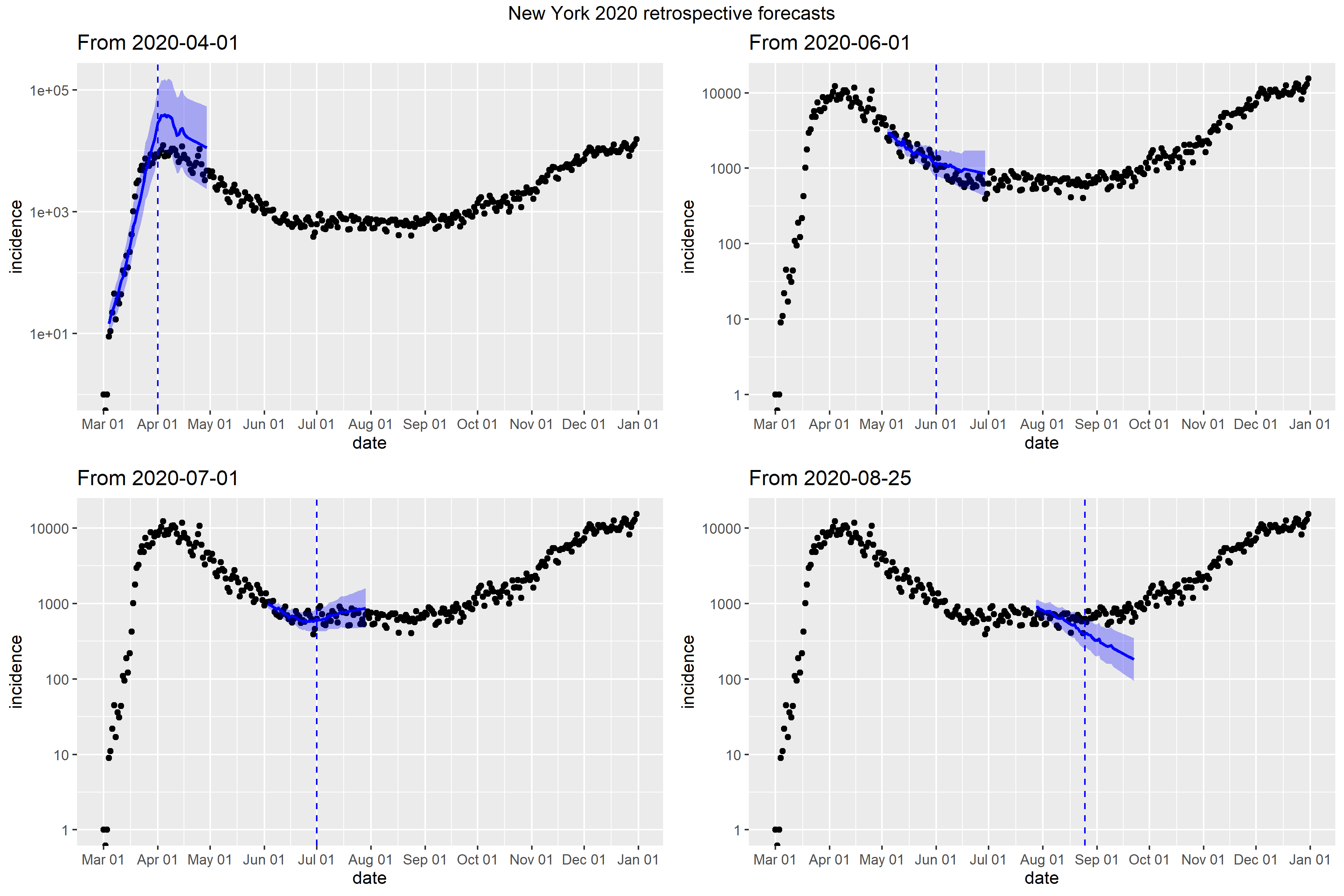

### North Carolina_retrospective_forecasts.png

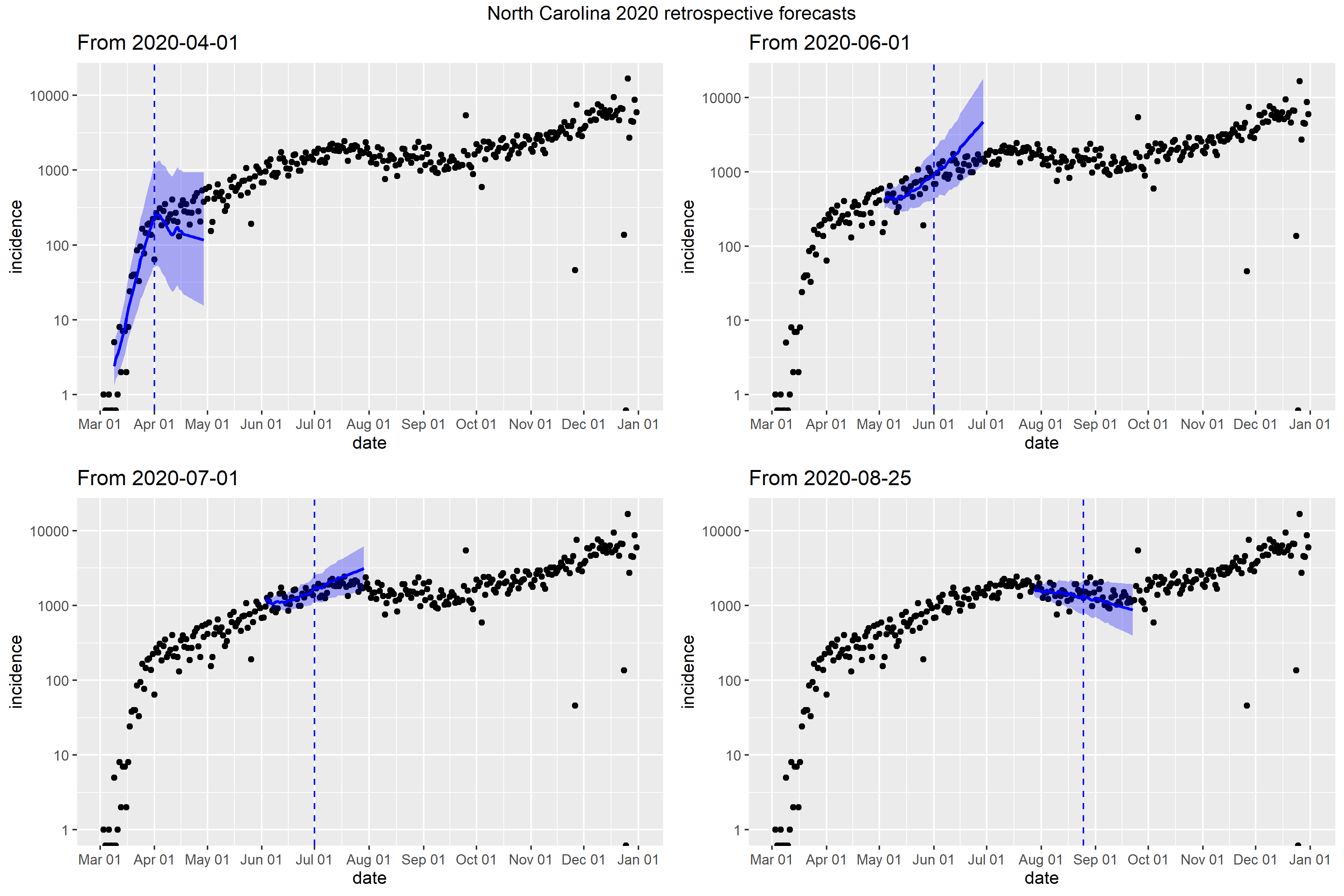

### North Dakota_retrospective_forecasts.png

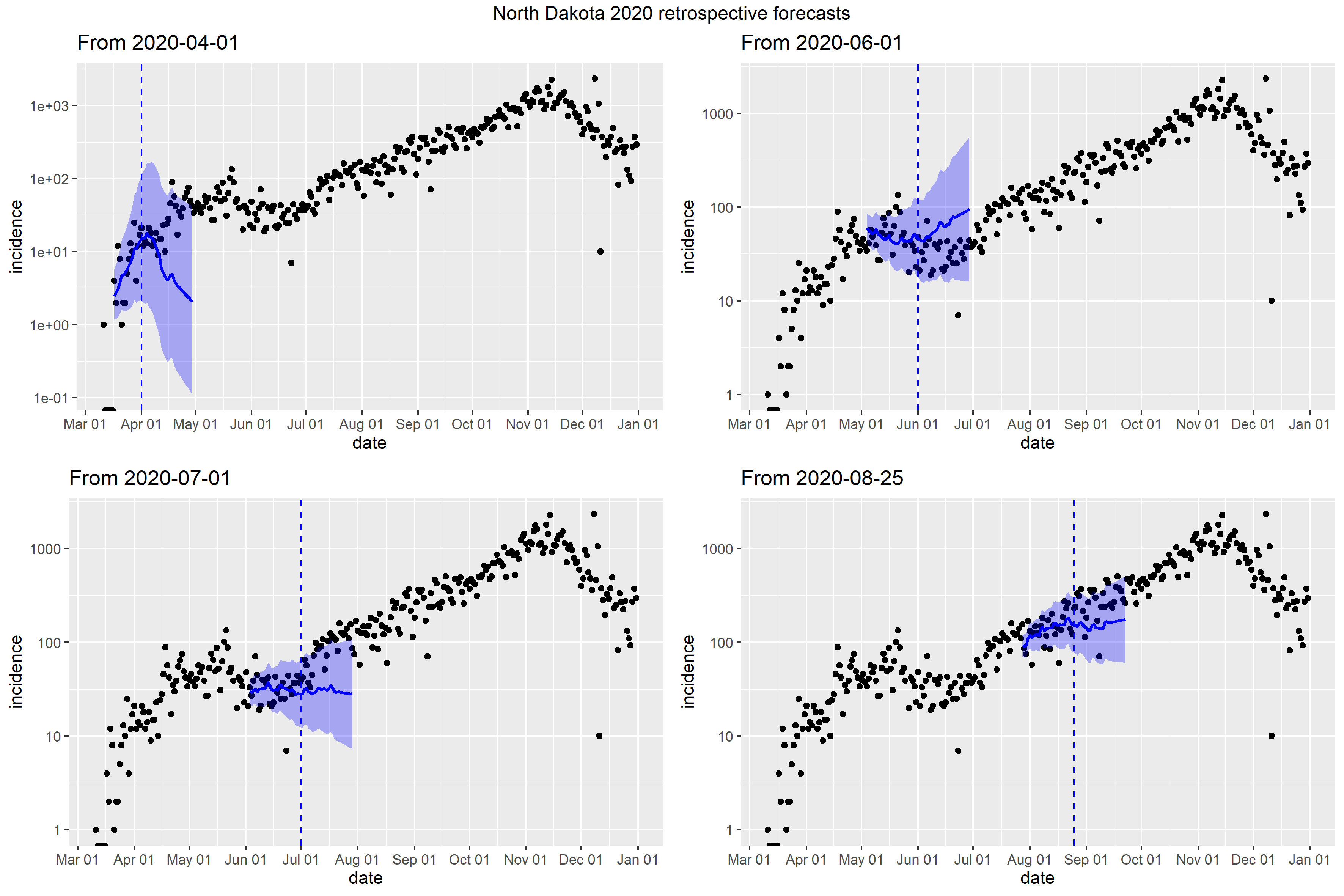

### Ohio_full_fit_plot.png

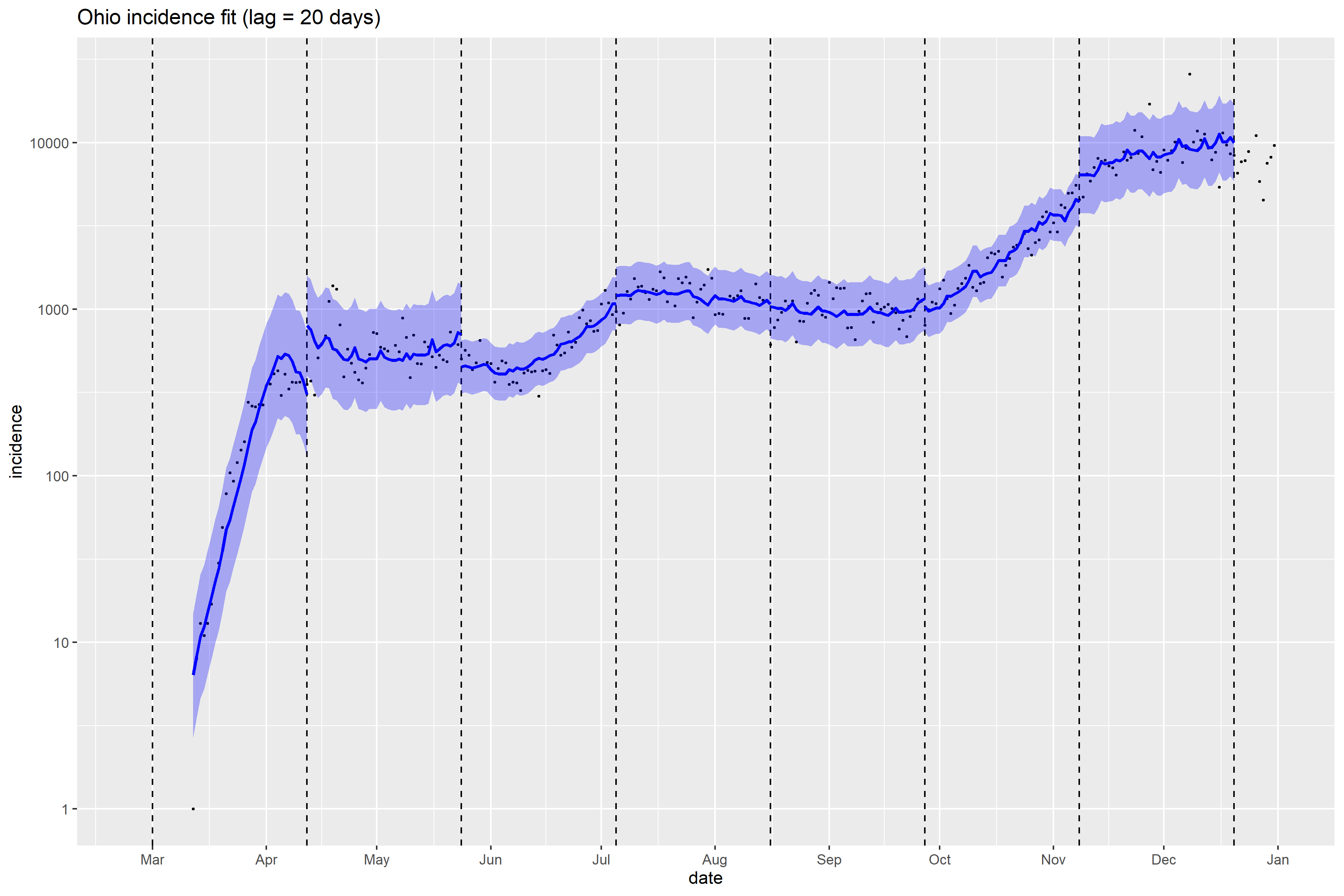

### Ohio_Peff_plot.png

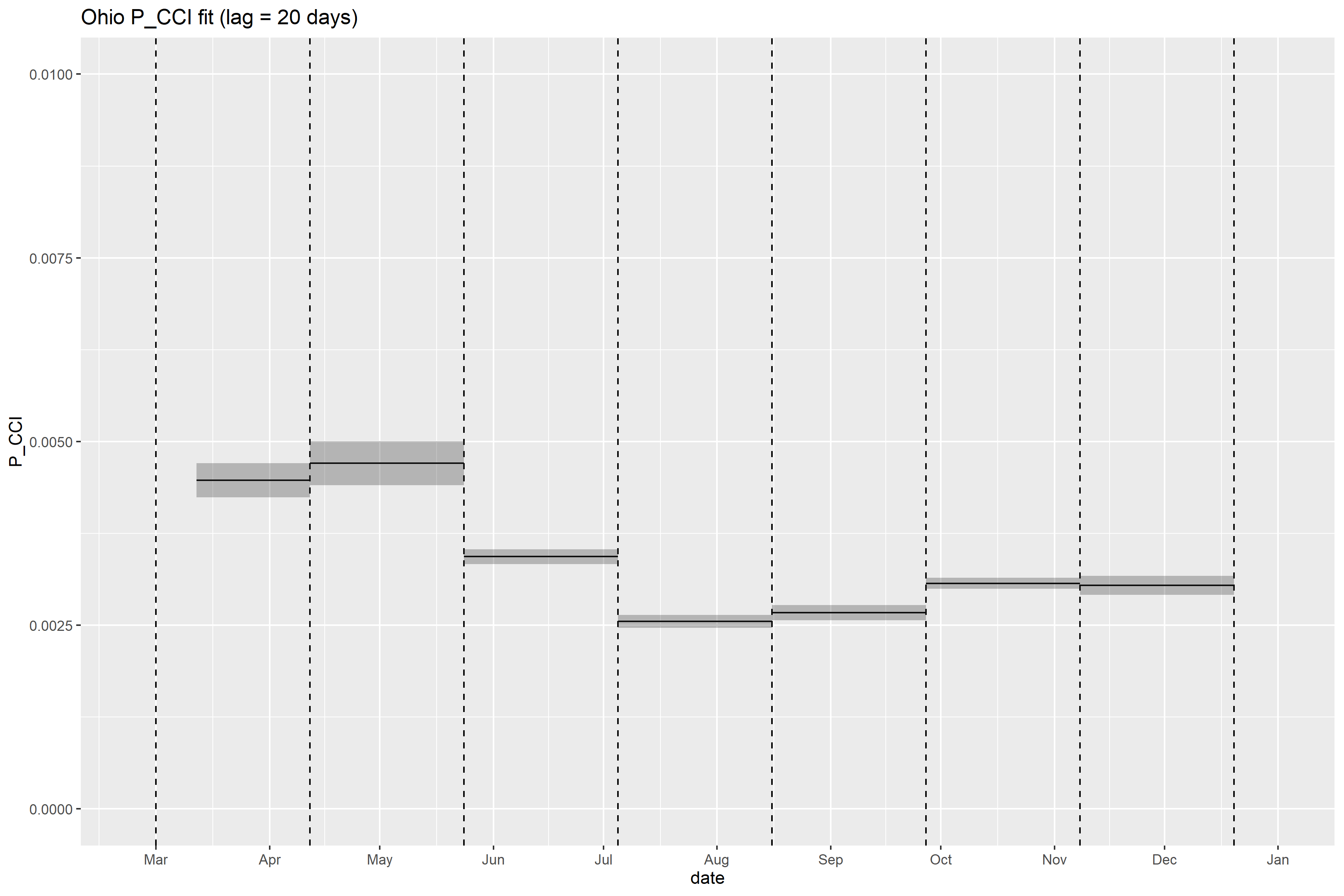

### Ohio_retrospective_forecasts.png

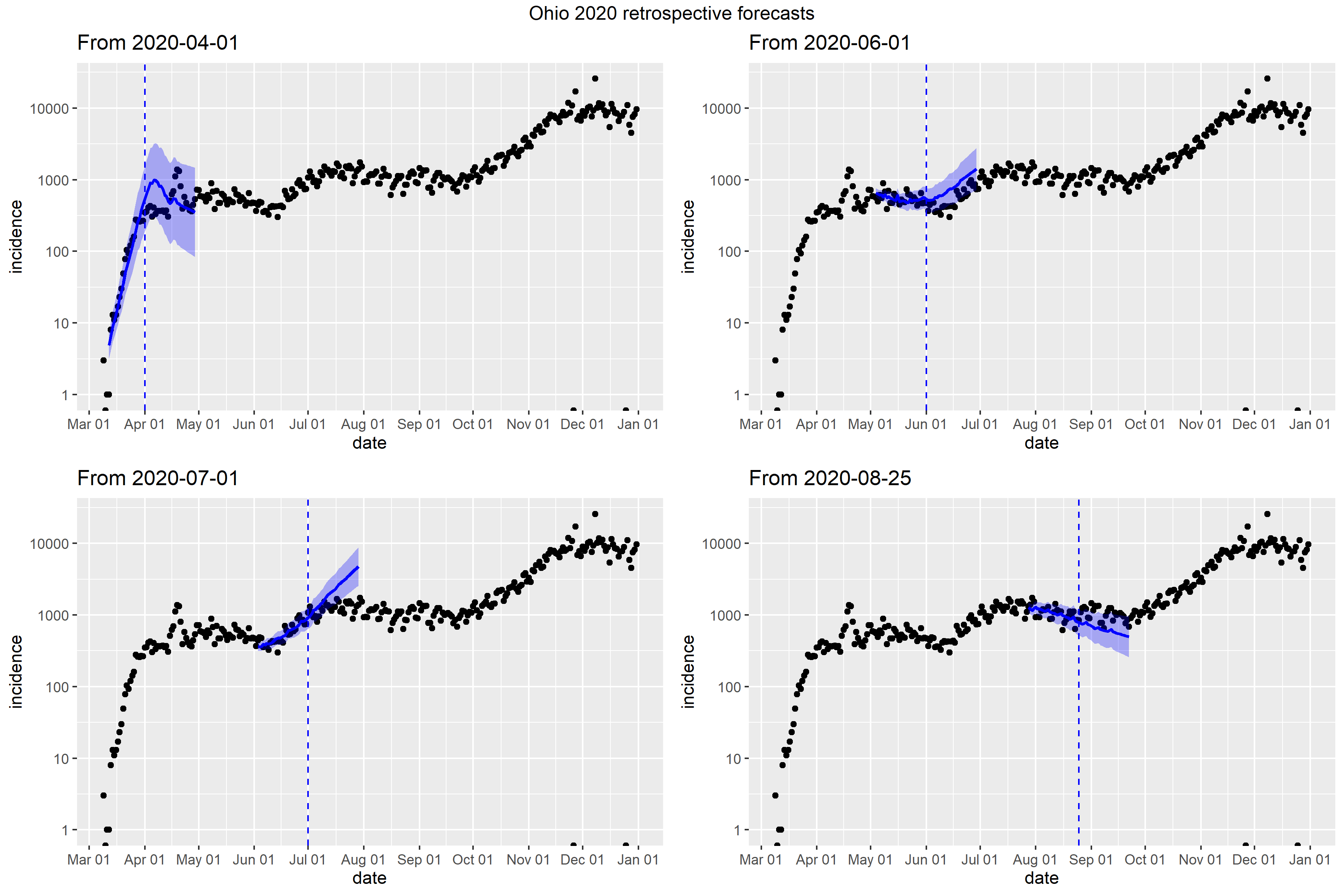

### Oklahoma_Peff_plot.png

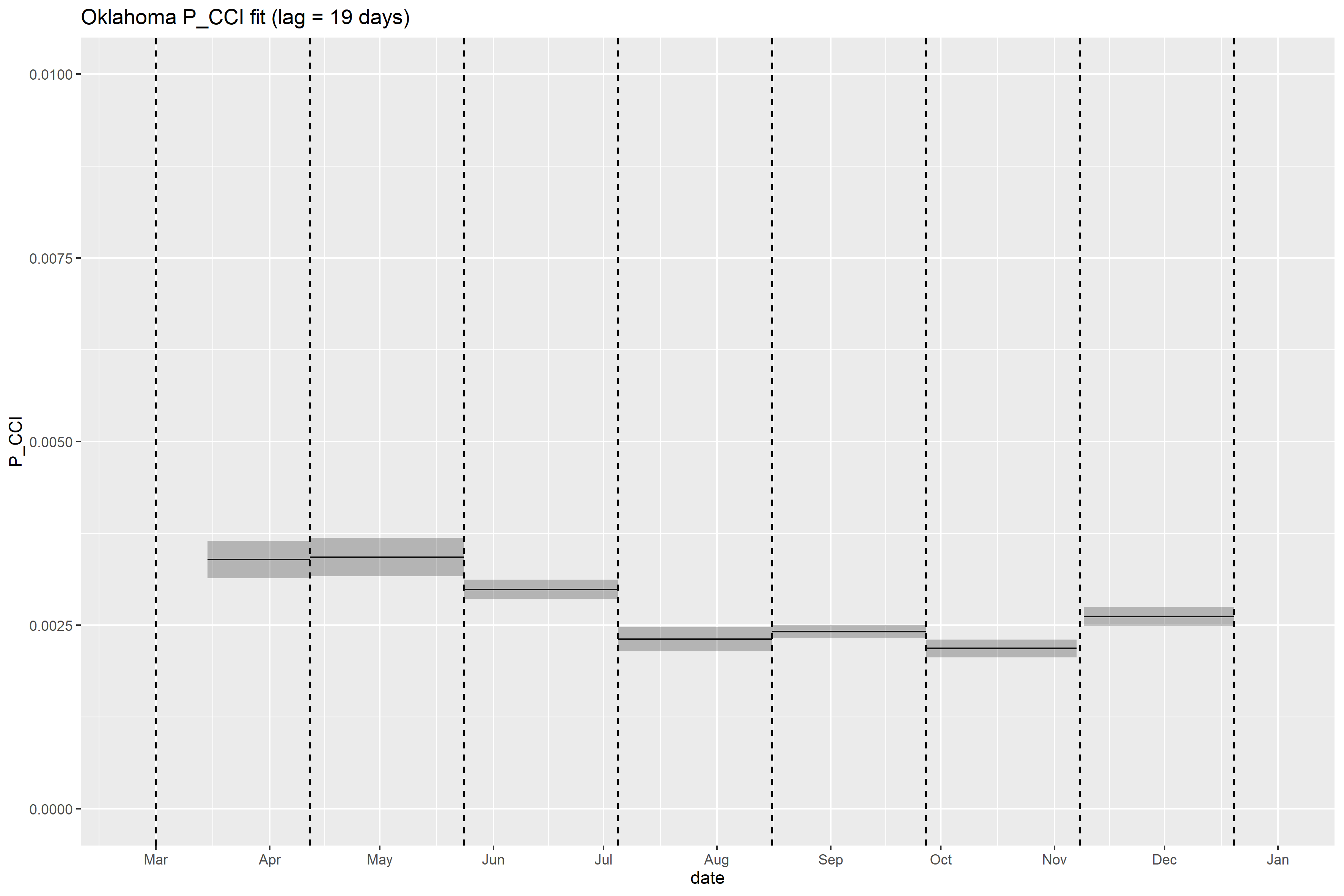

### Oklahoma_retrospective_forecasts.png

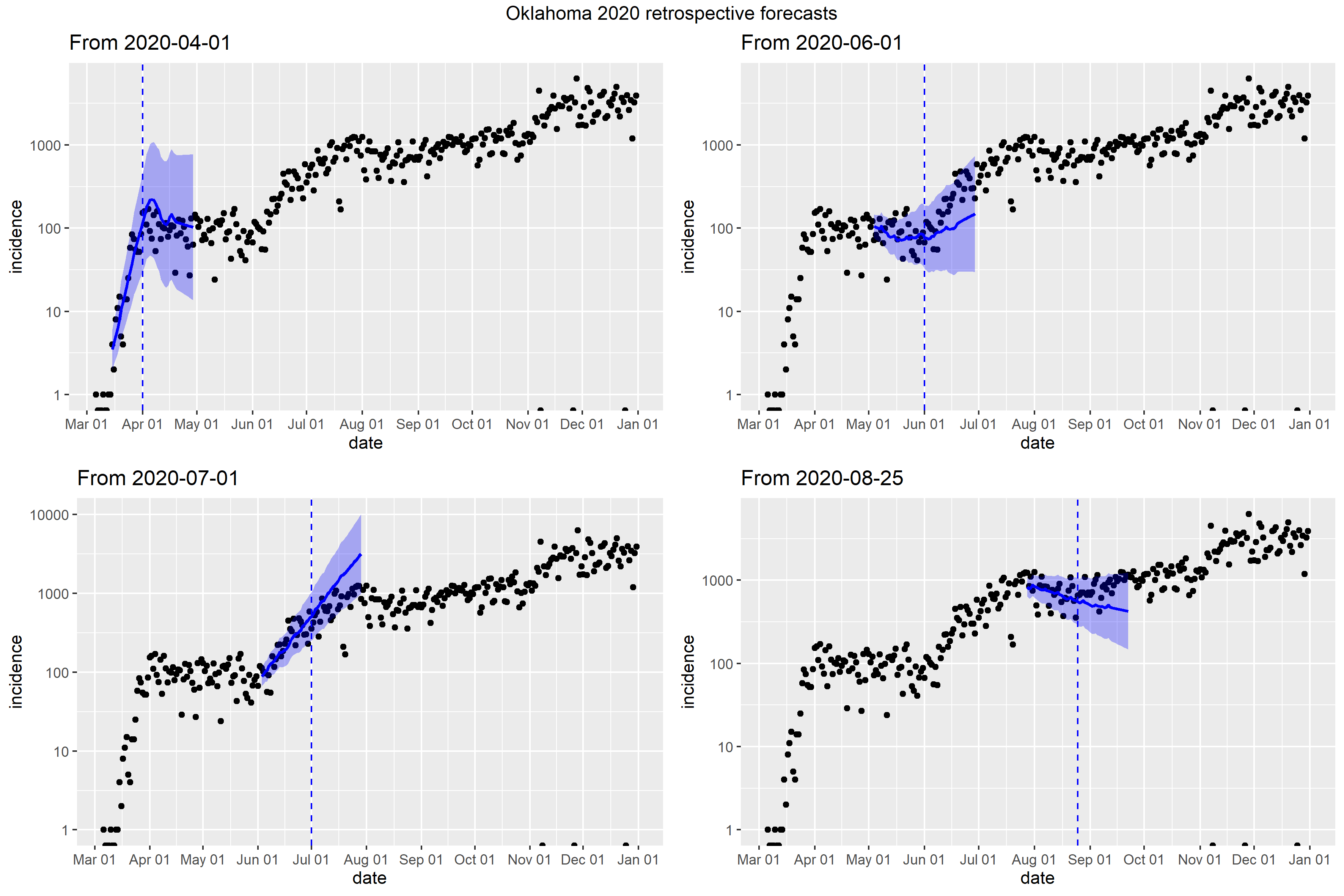

### Oregon_full_fit_plot.png

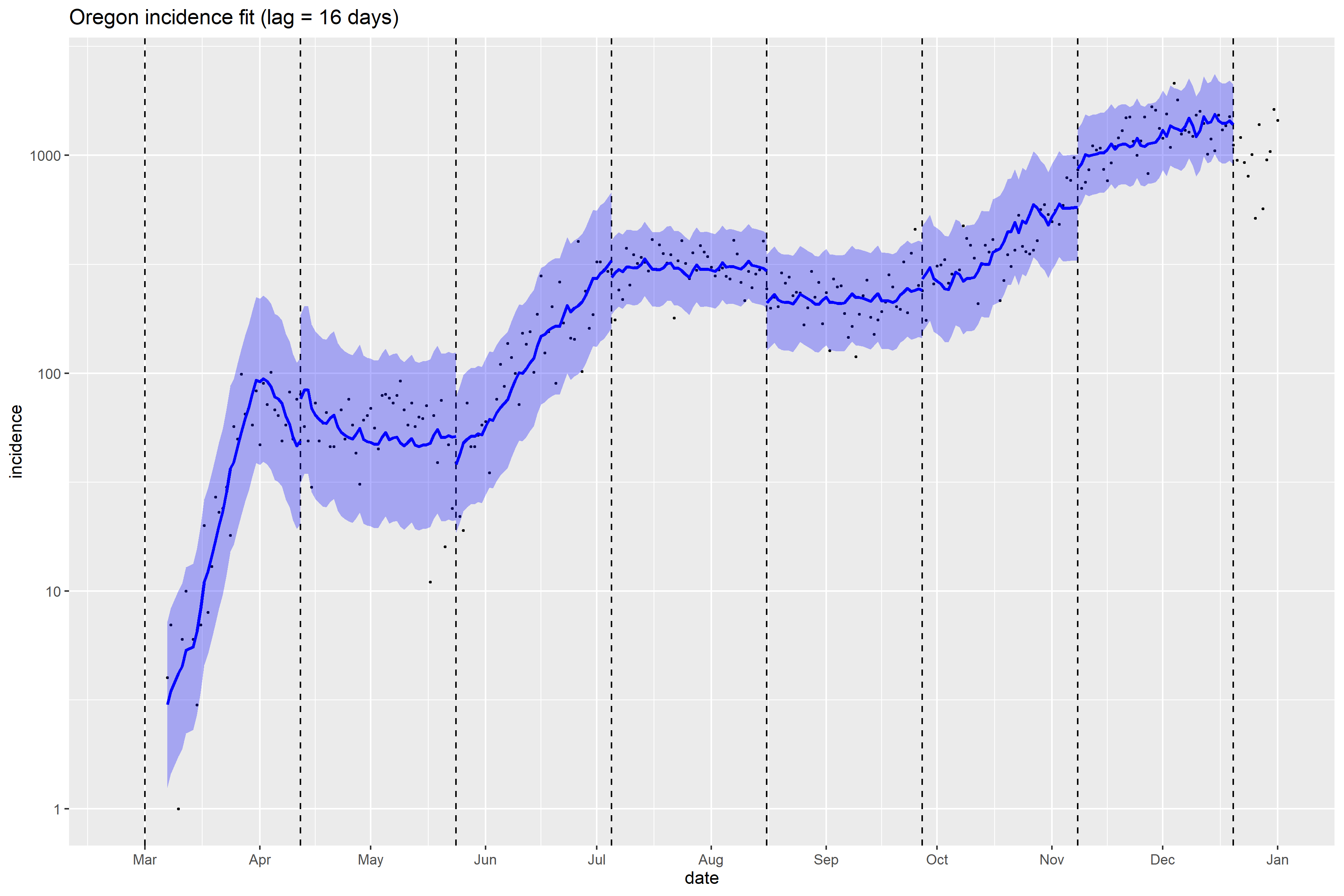

### Oregon_Peff_plot.png

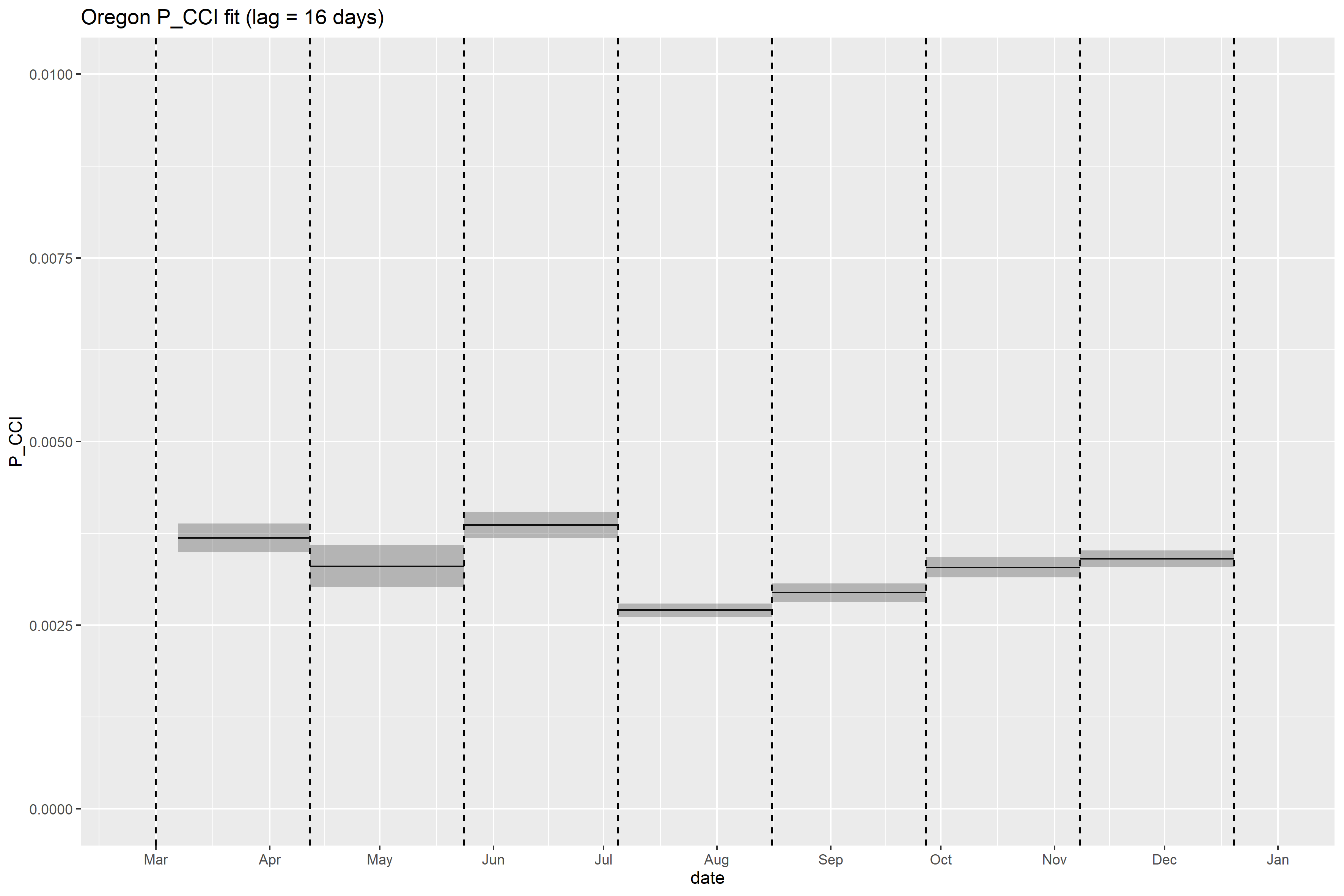

### Oregon_retrospective_forecasts.png

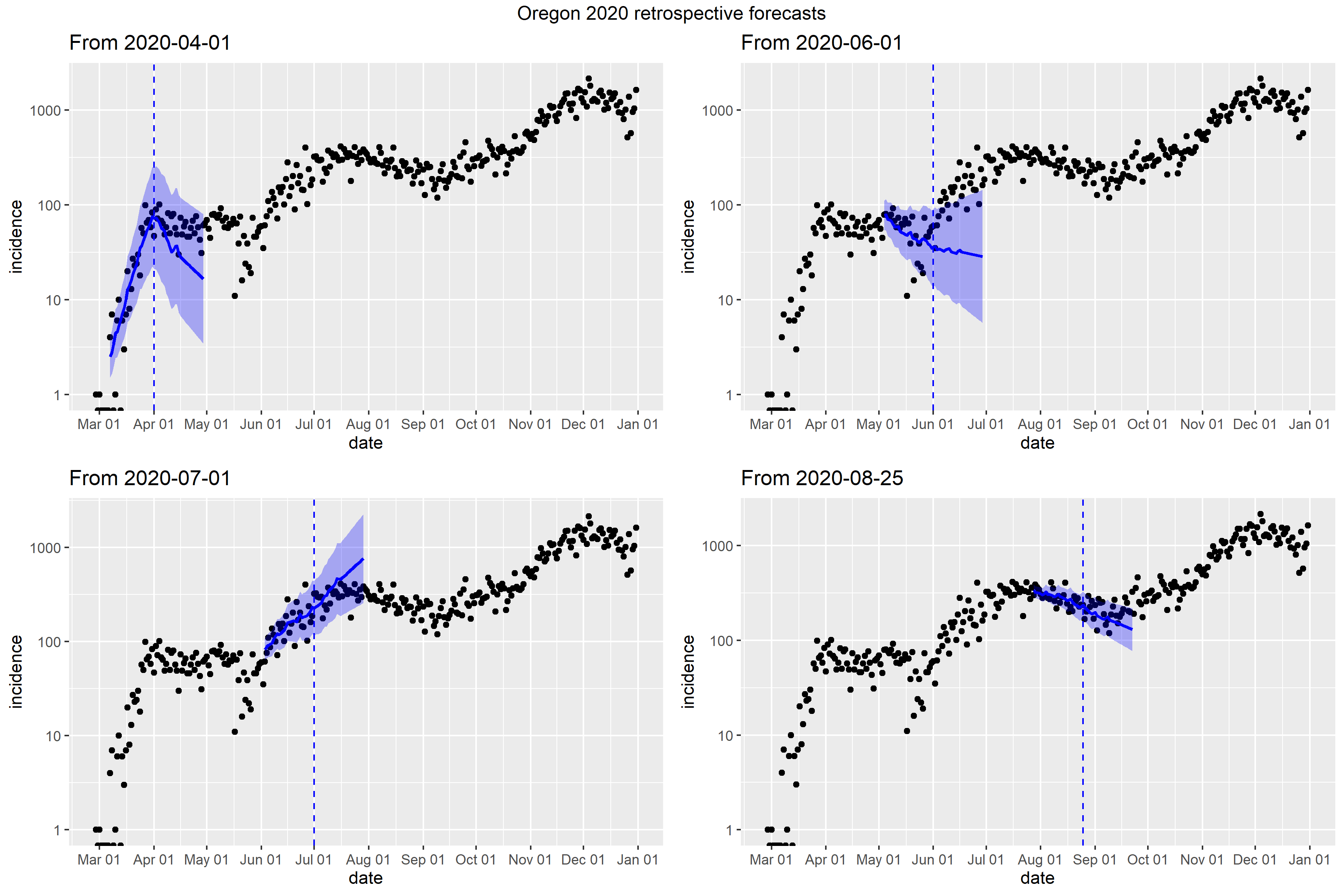

### Pennsylvania_retrospective_forecasts.png

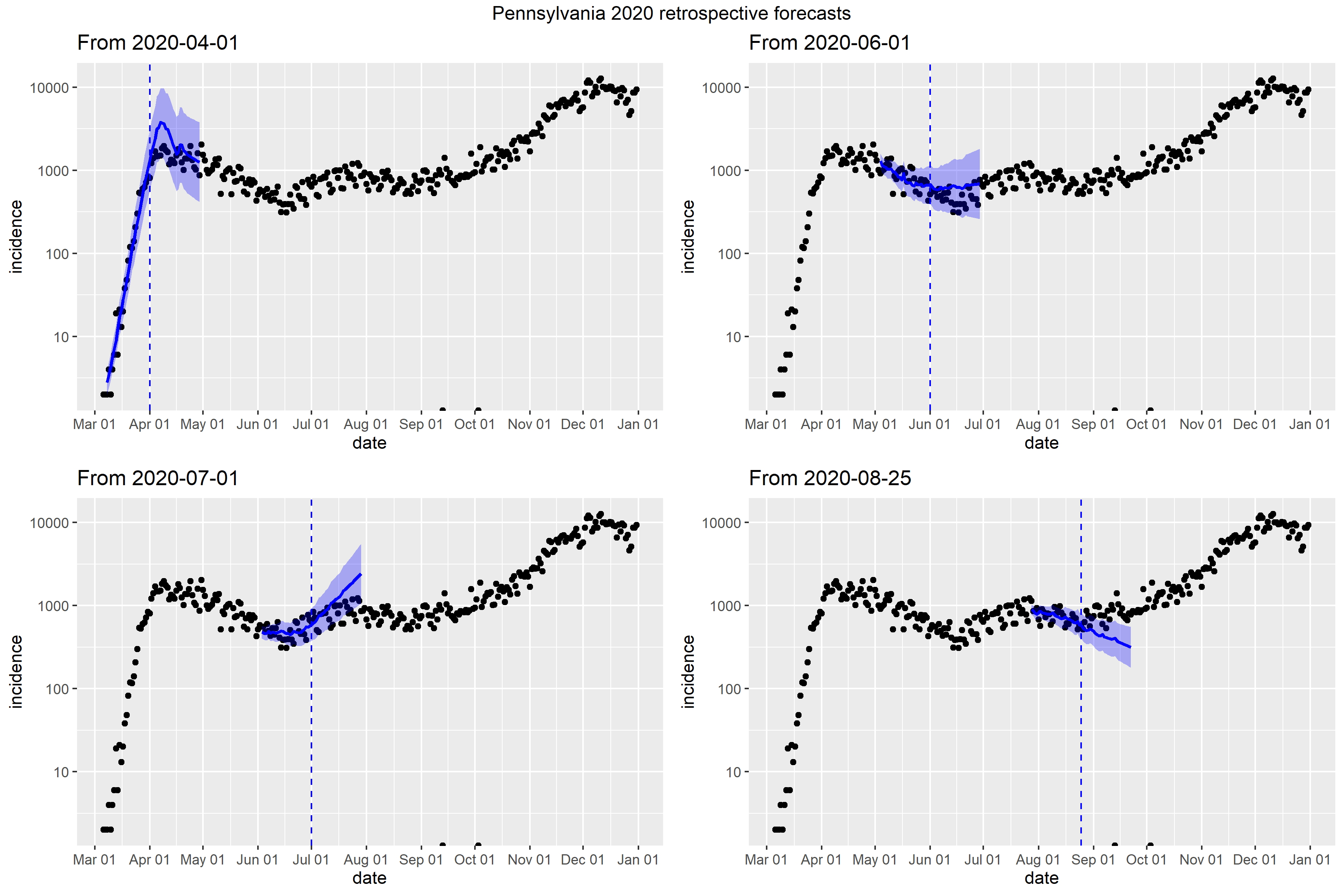

### Rhode Island_retrospective_forecasts.png

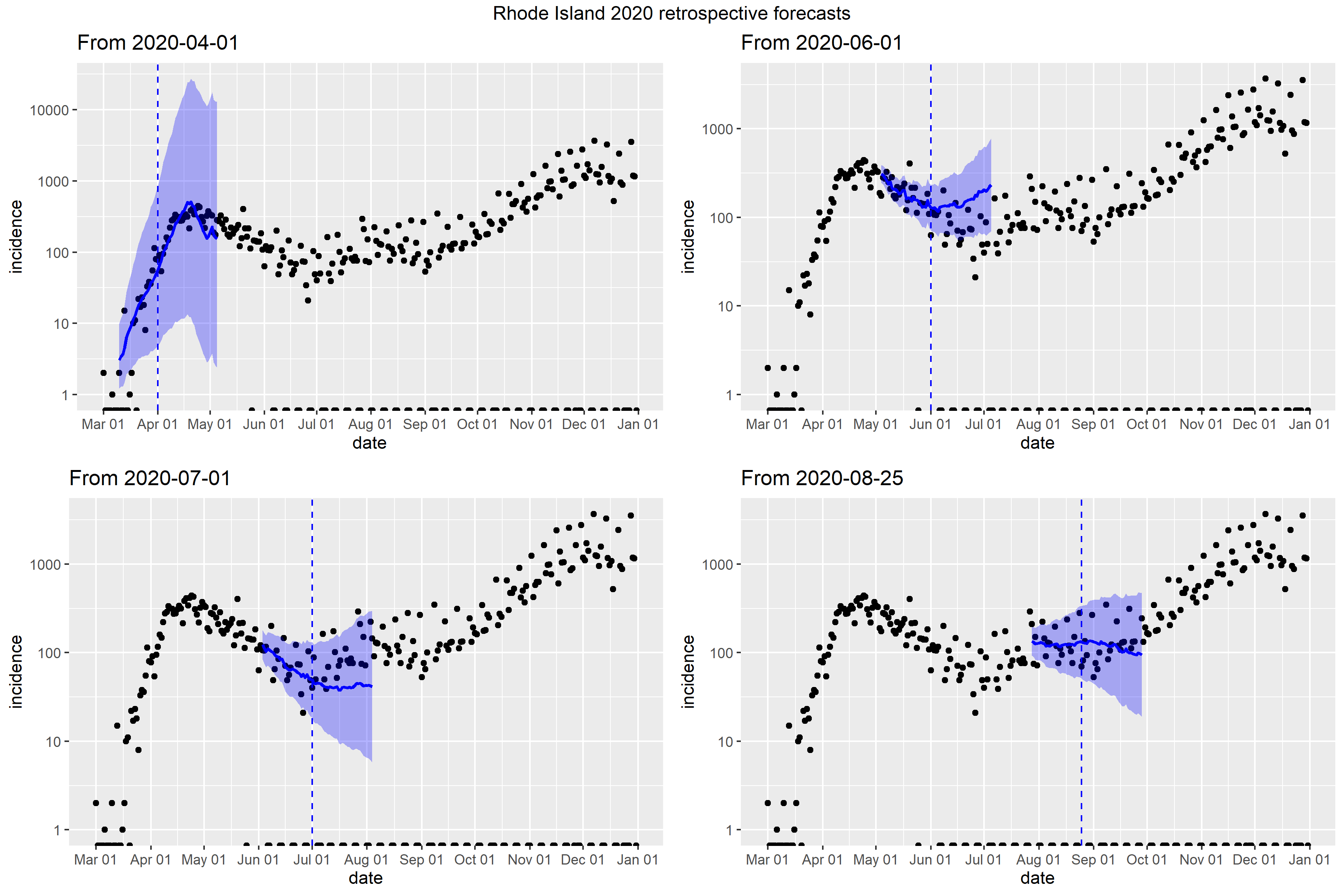

### South Carolina_retrospective_forecasts.png

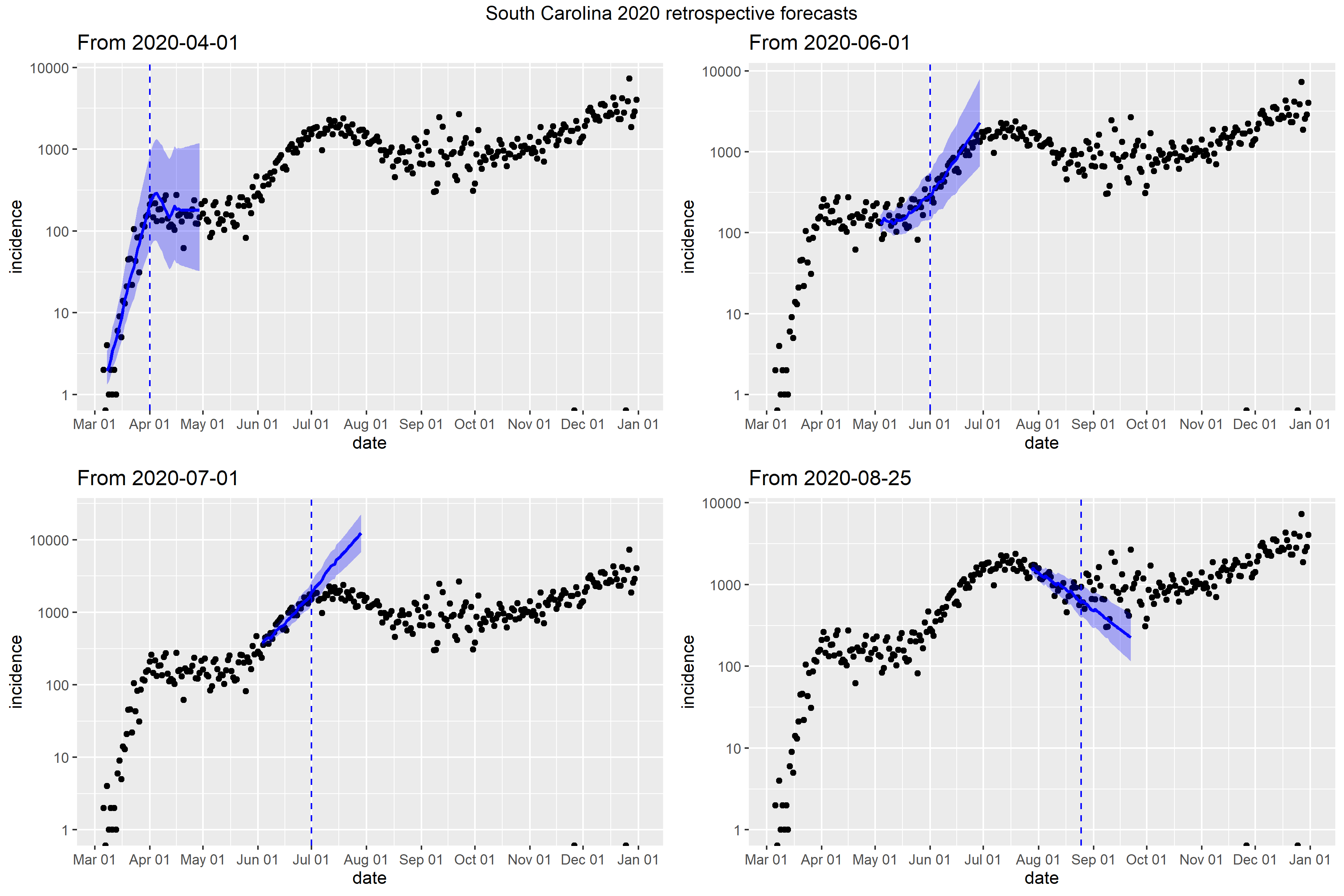

### South Dakota_retrospective_forecasts.png

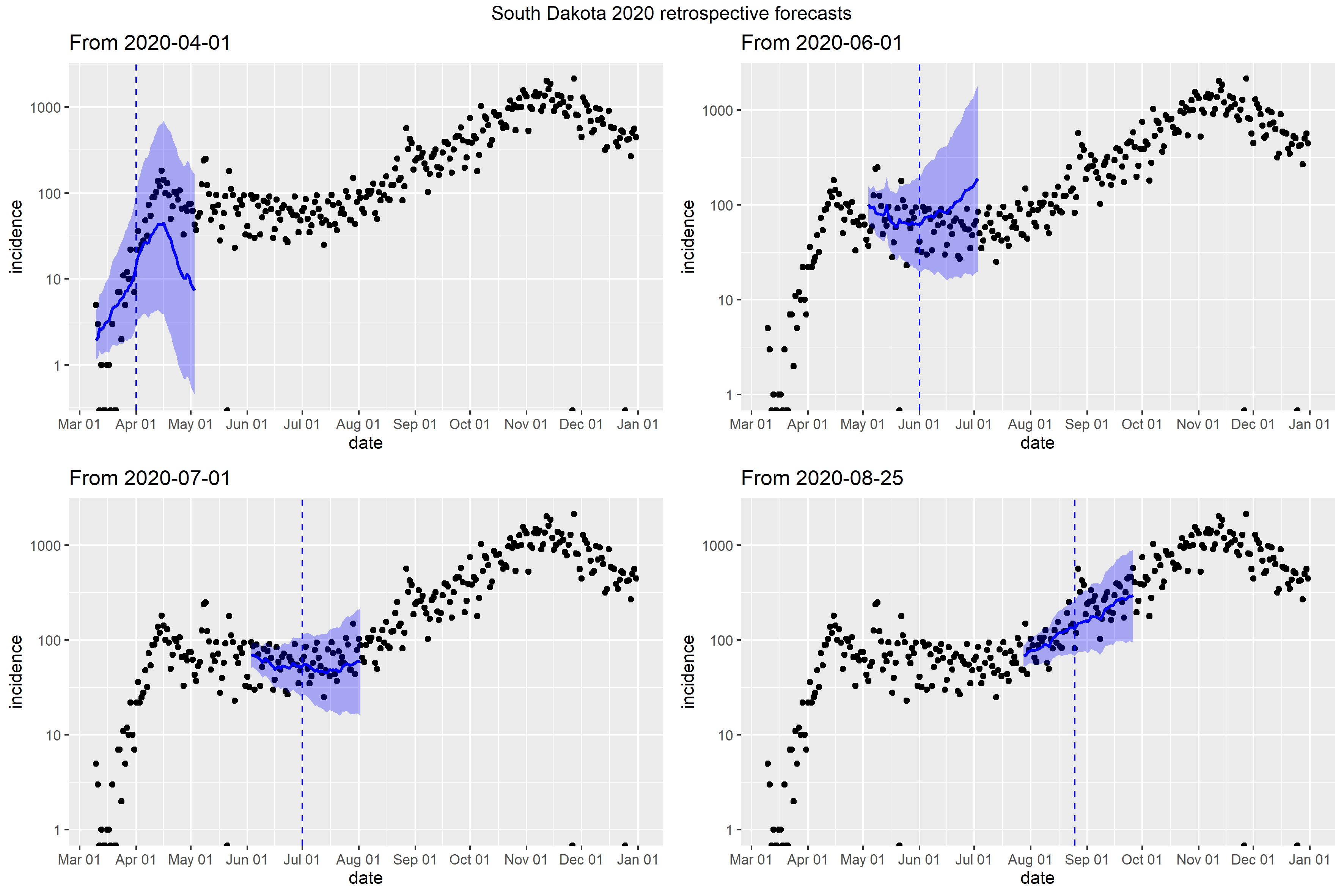

### Tennessee_Peff_plot.png

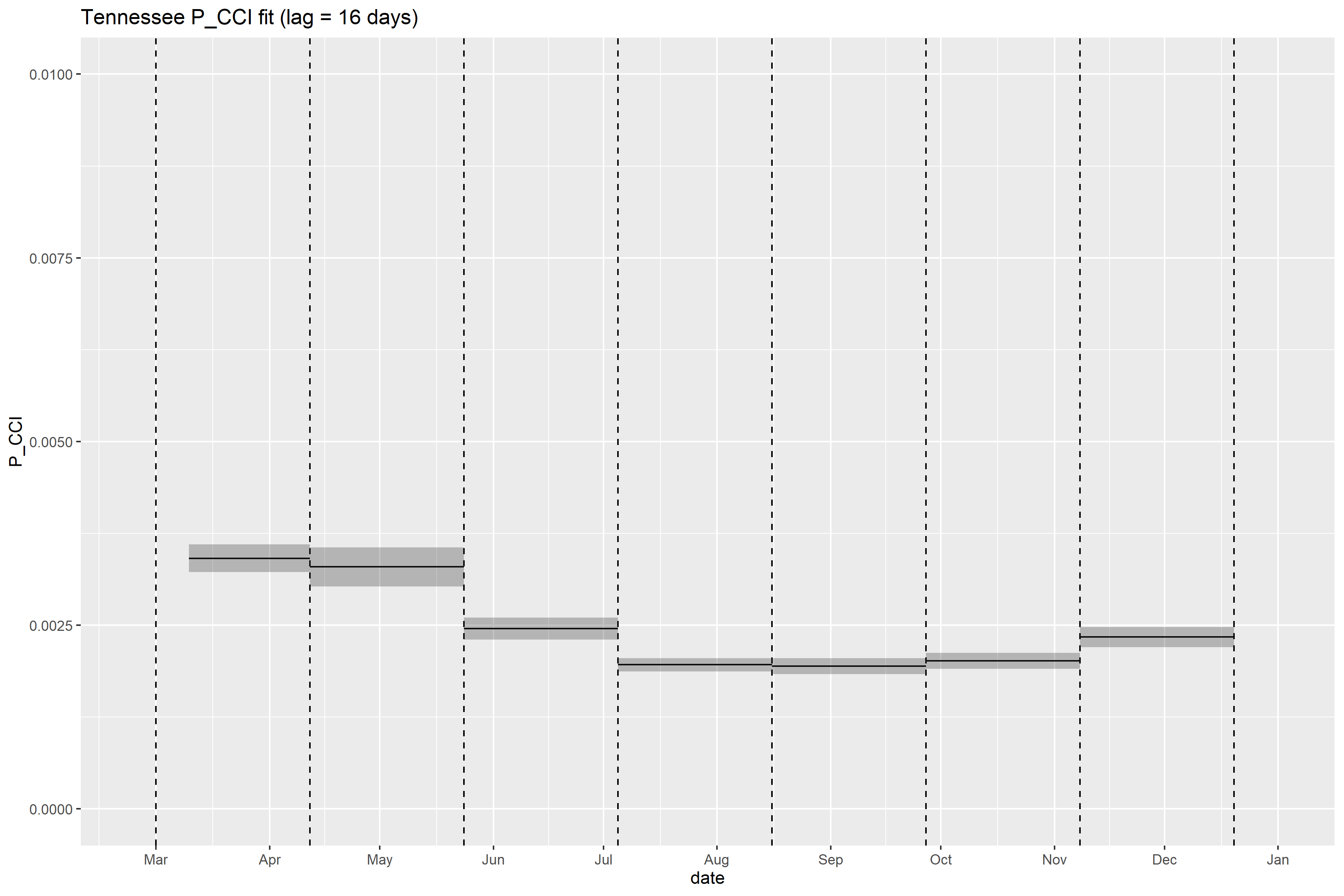

### Tennessee_retrospective_forecasts.png

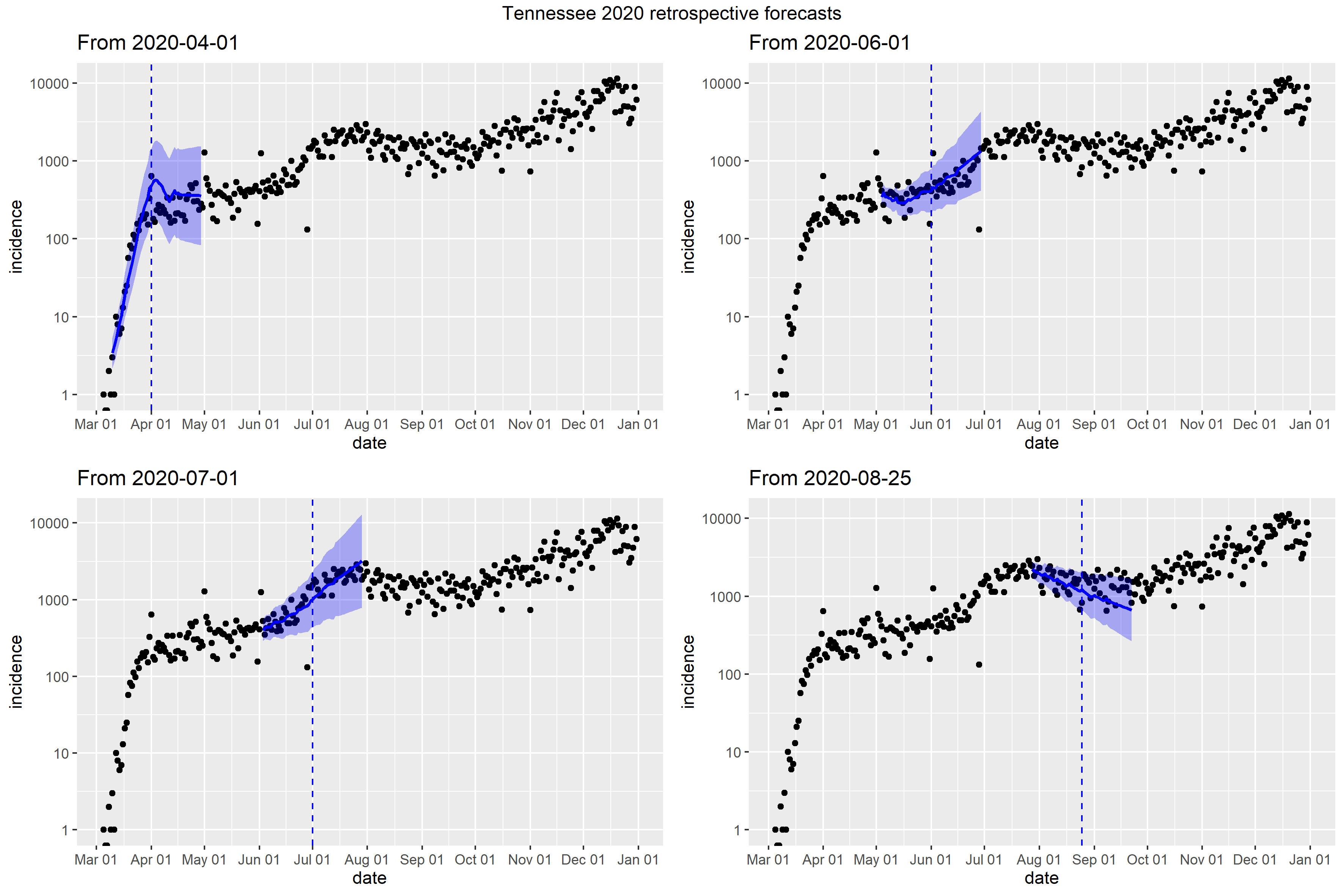

### Texas_full_fit_plot.png

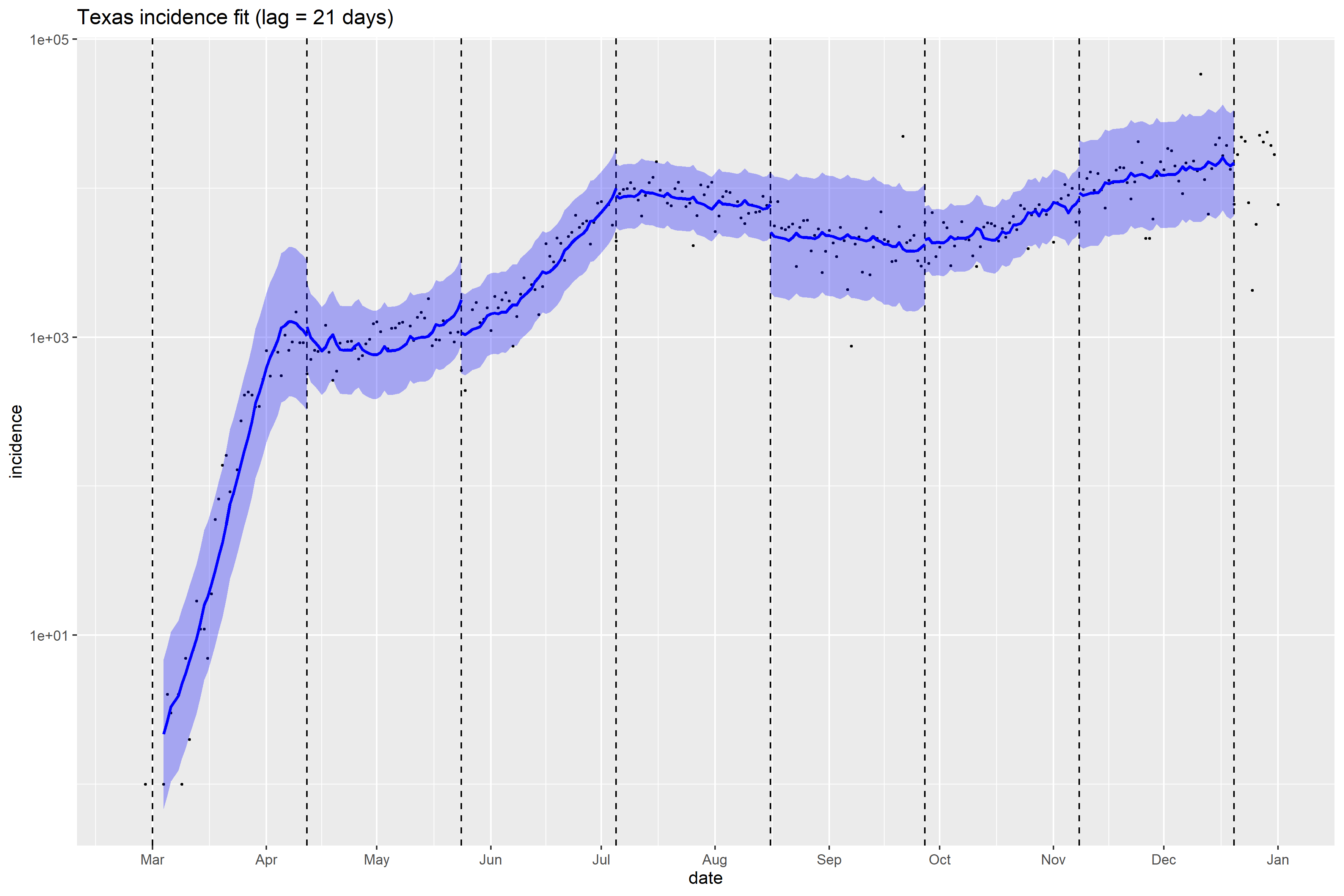

### Texas_Peff_plot.png

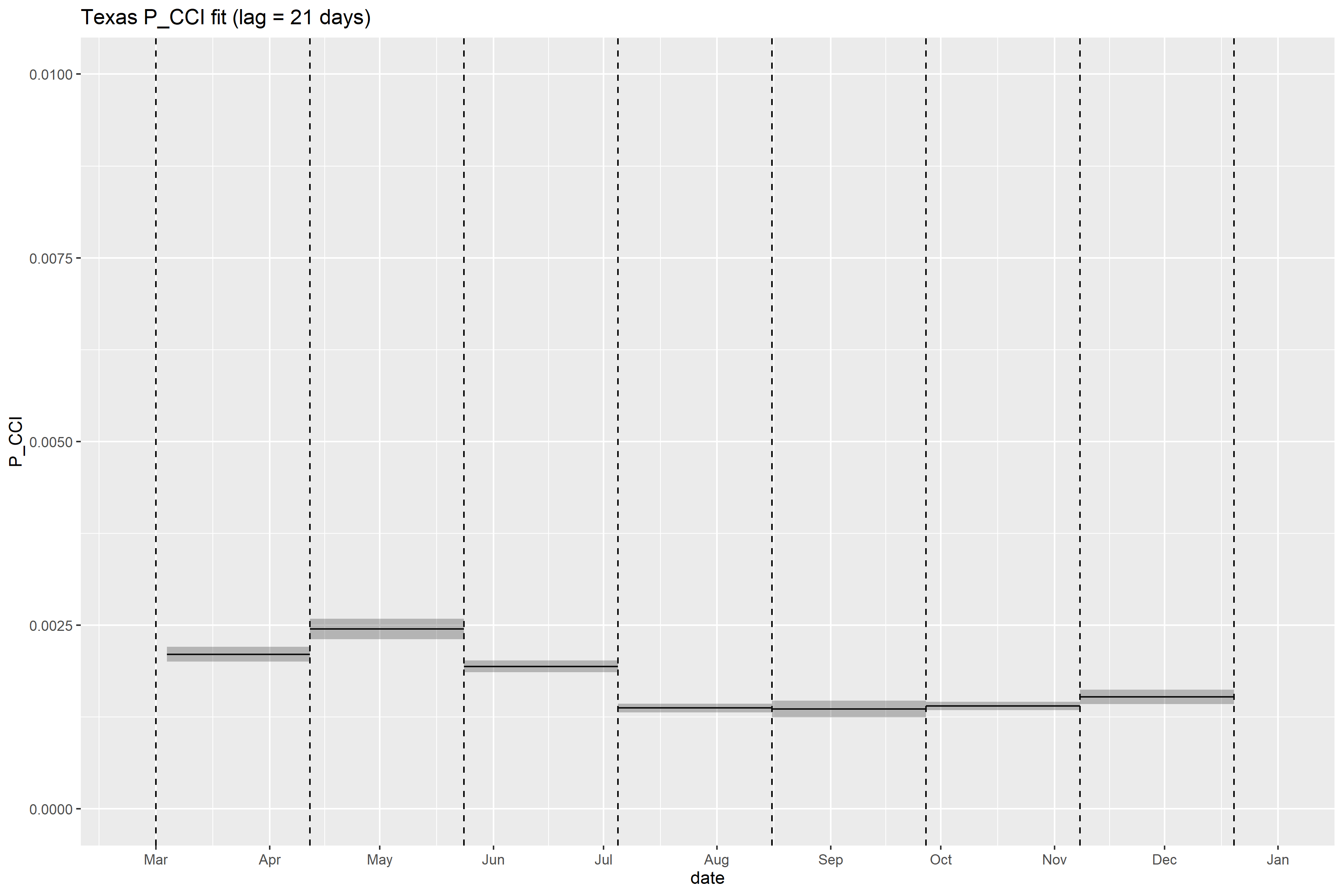

### Texas_retrospective_forecasts.png

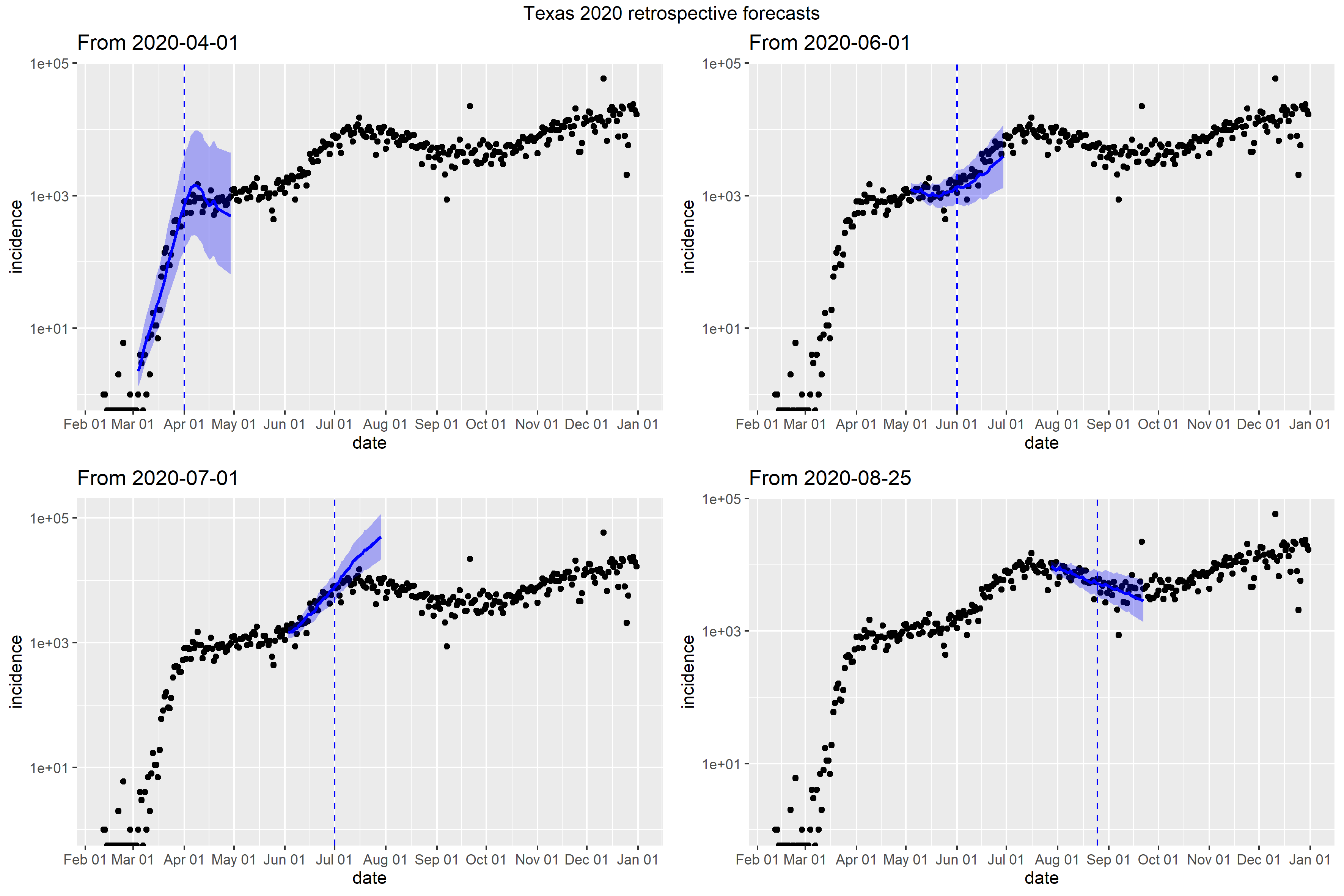

### Utah_full_fit_plot.png

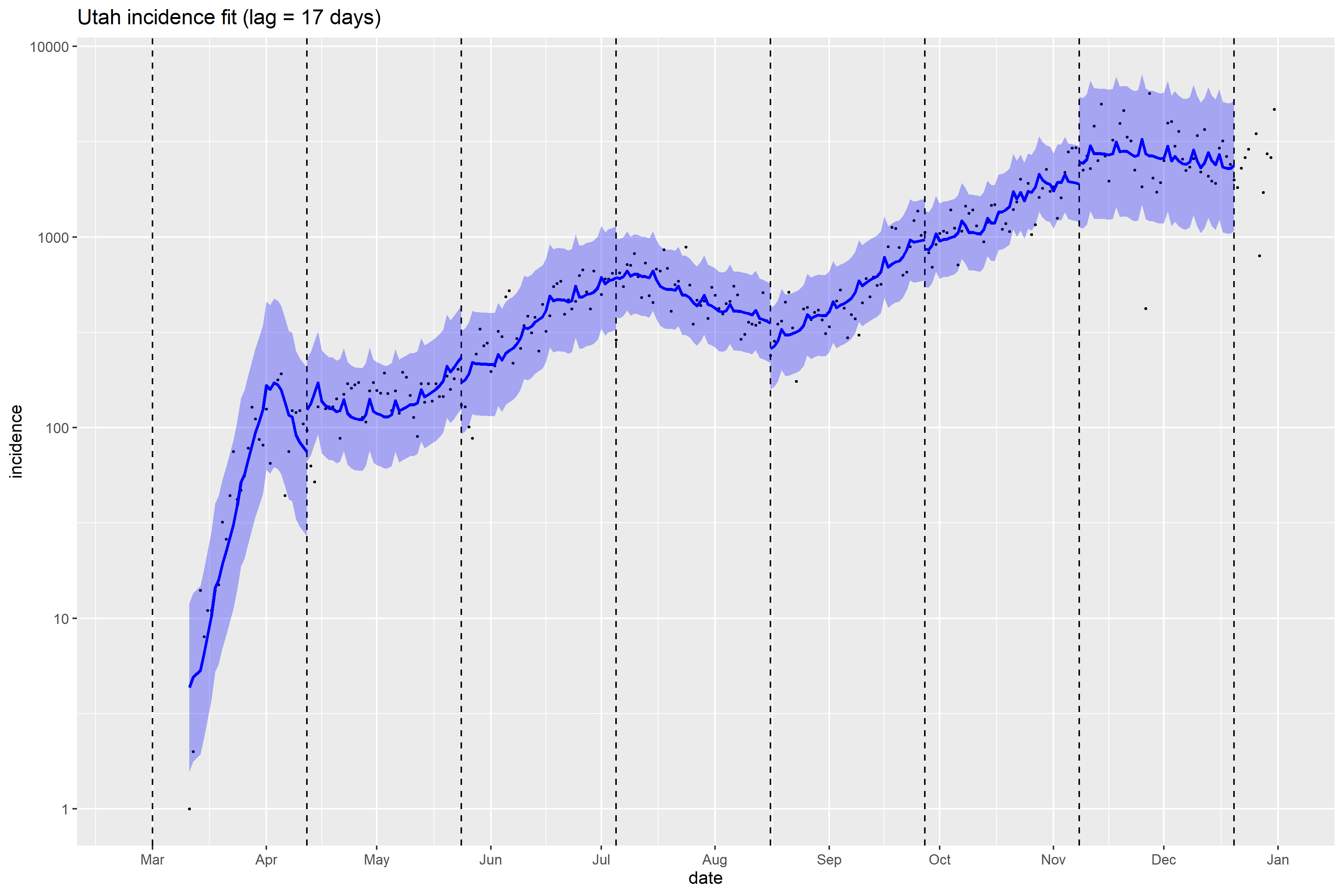

### Utah_Peff_plot.png

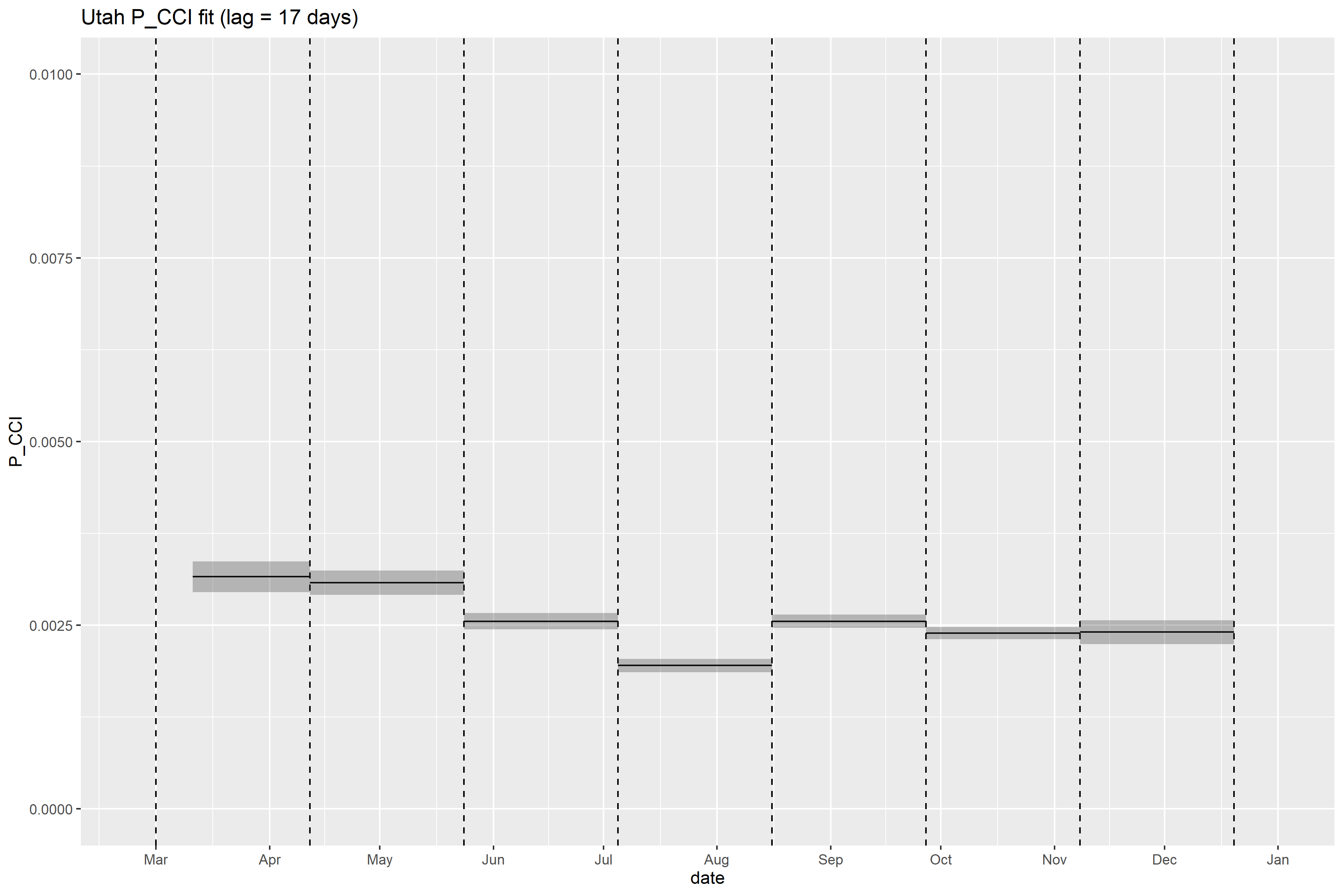

### Utah_retrospective_forecasts.png

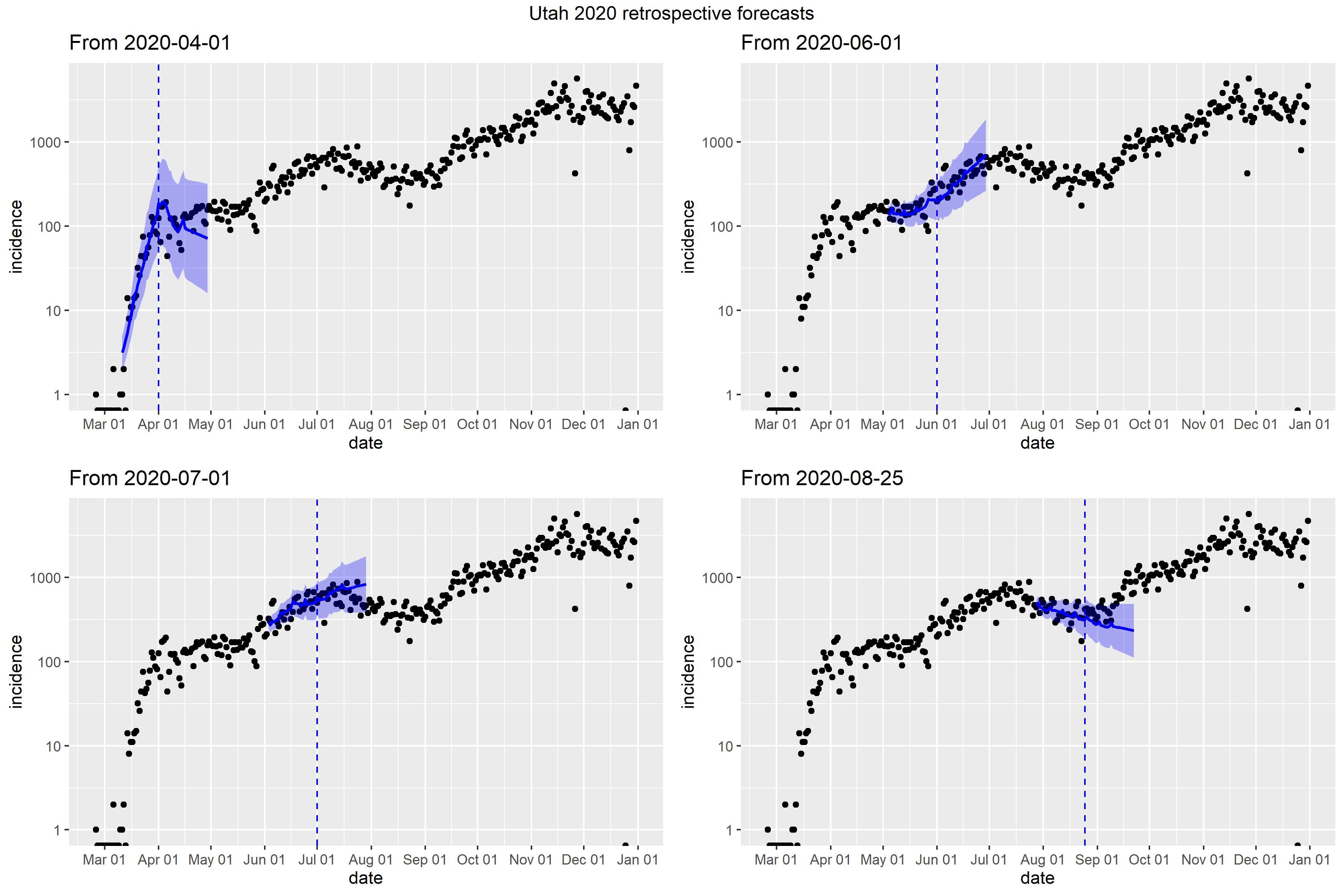

### Vermont_Peff_plot.png

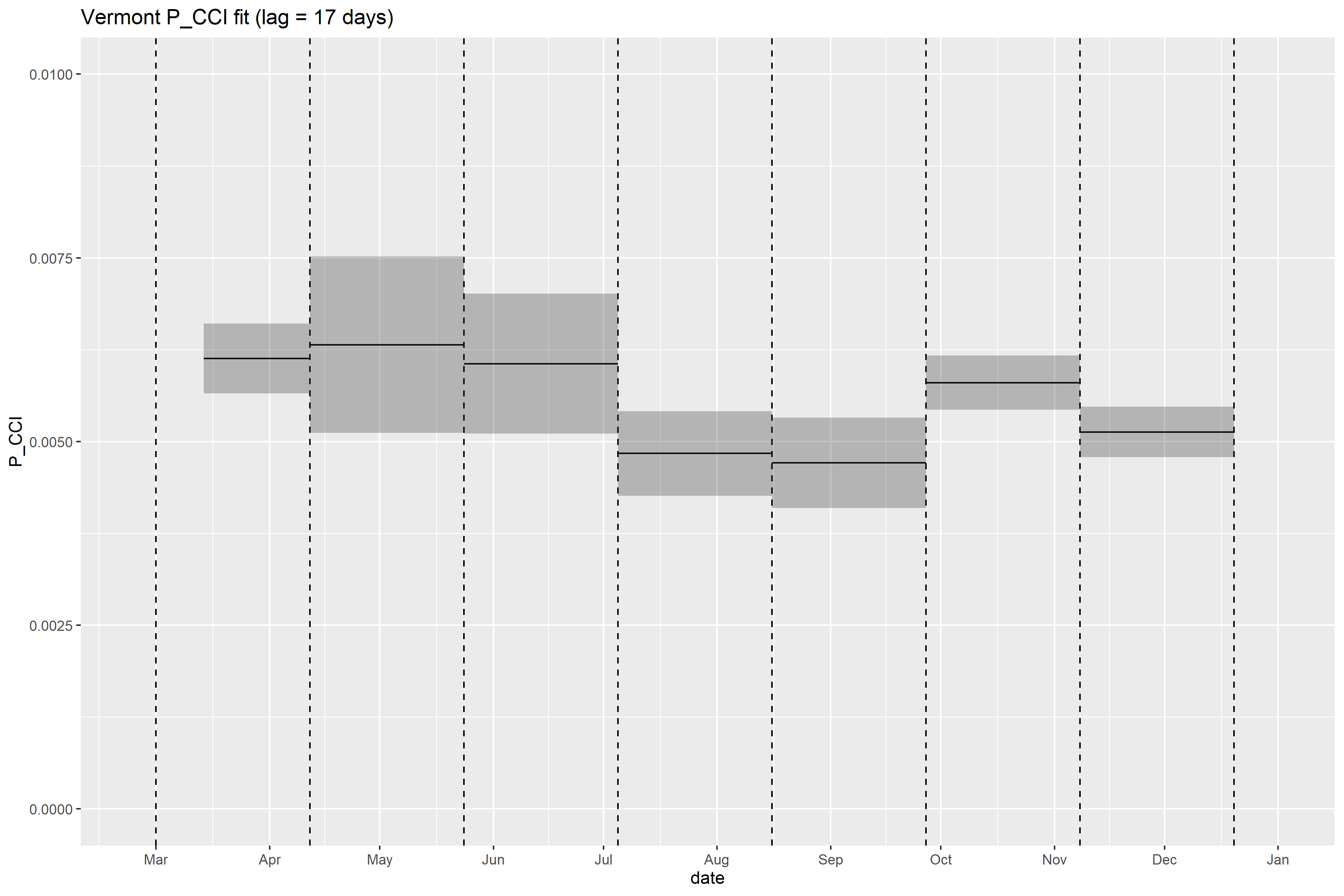

### Vermont_retrospective_forecasts.png

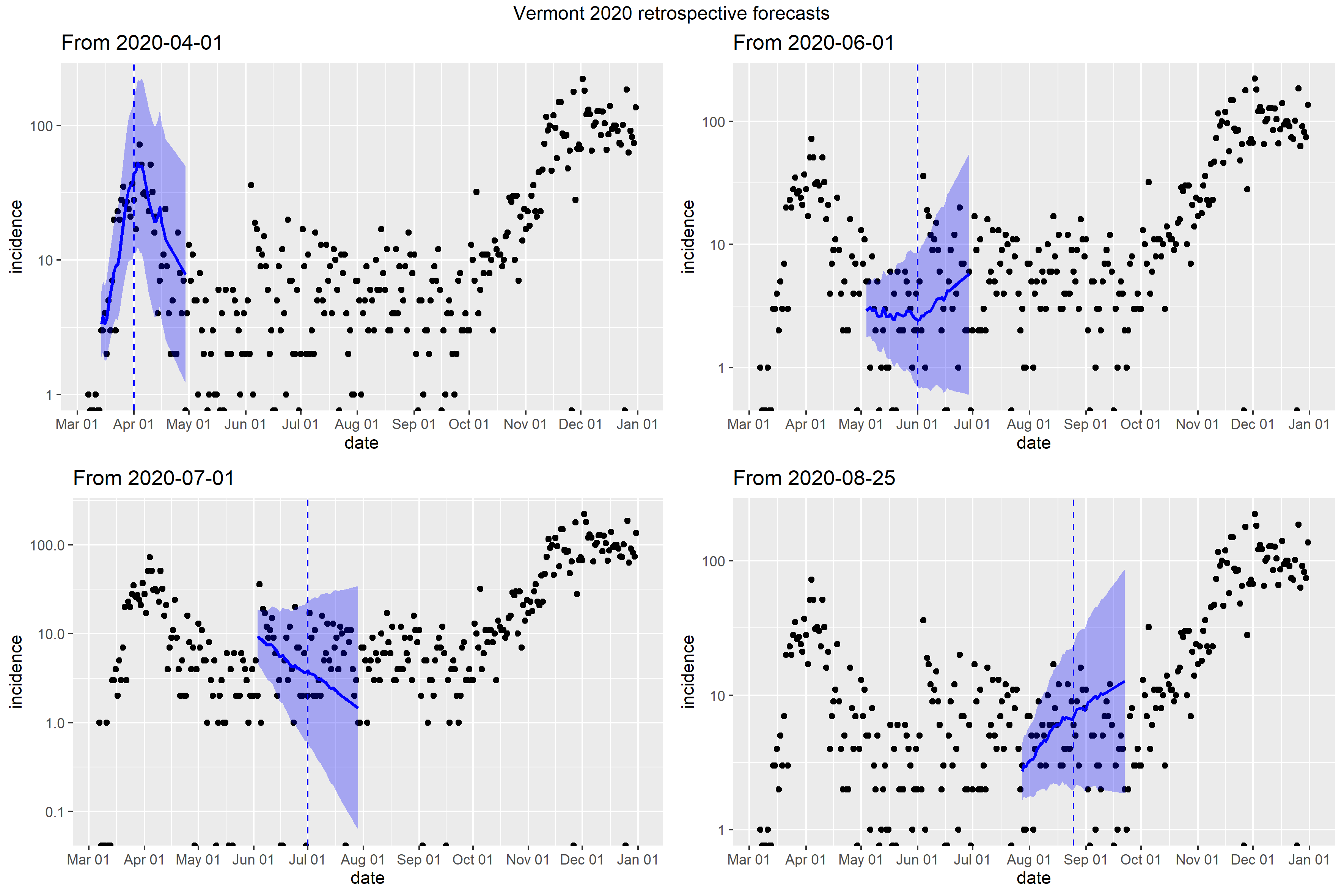

### Virginia_Peff_plot.png

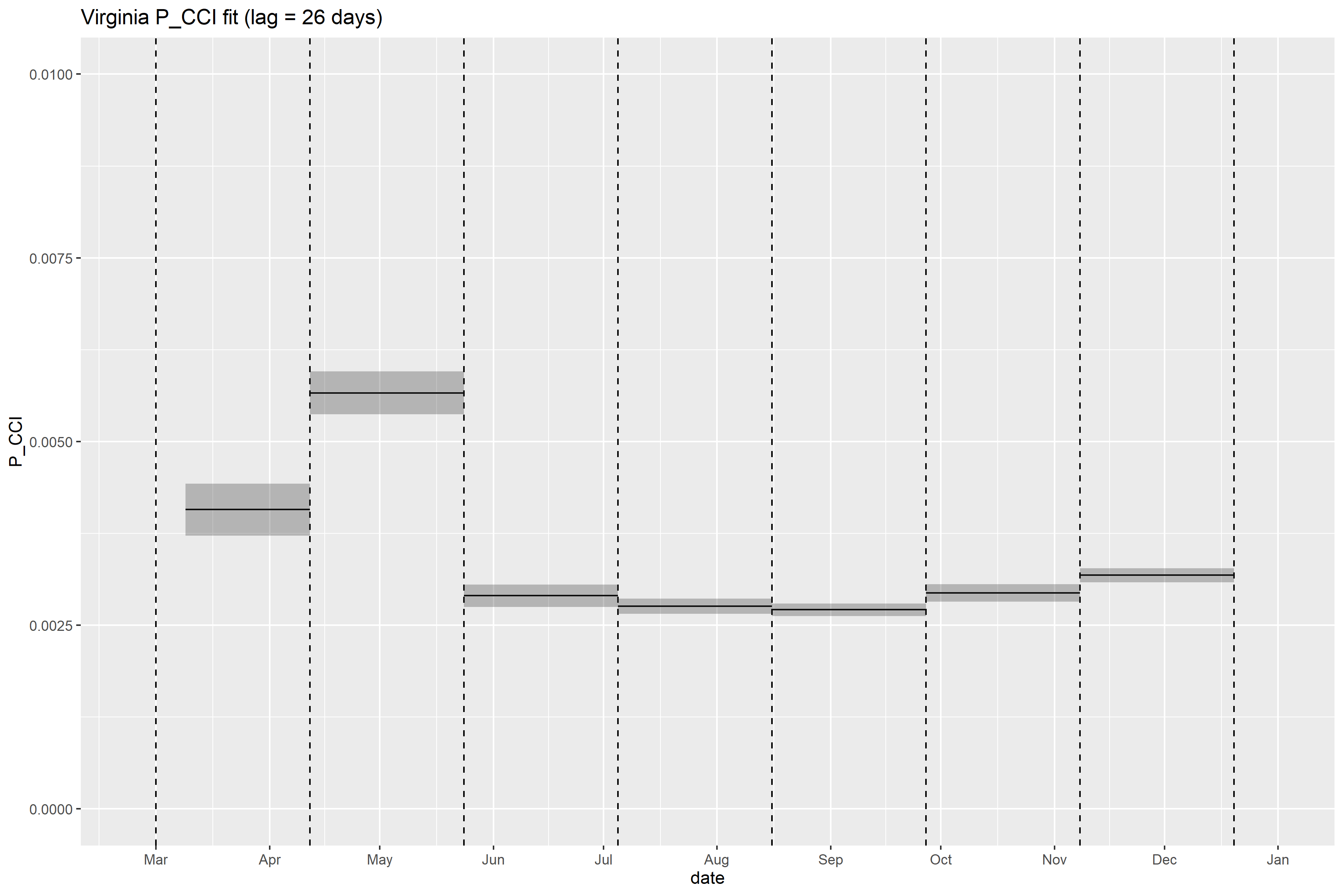

### Virginia_retrospective_forecasts.png

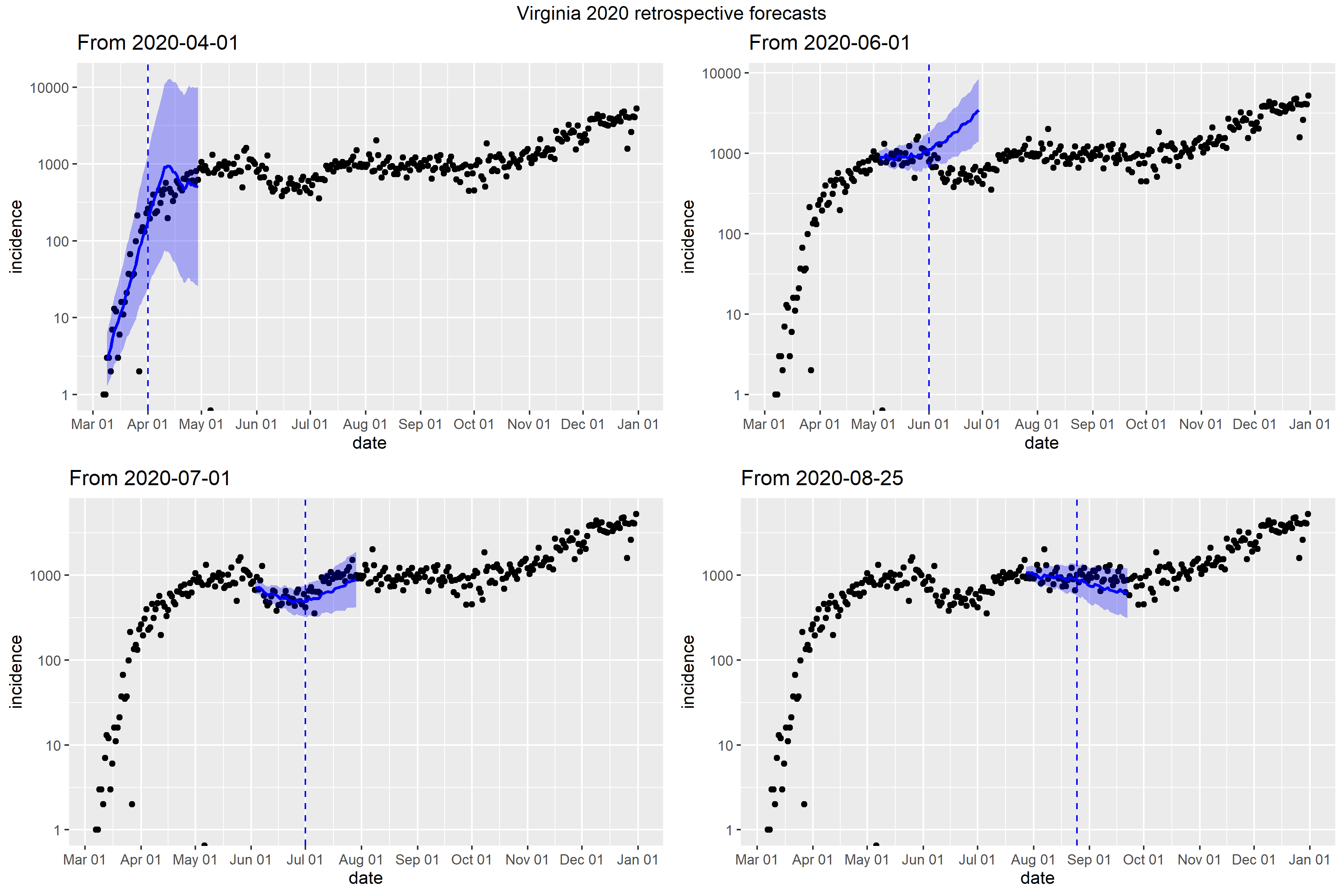

### Washington_retrospective_forecasts.png

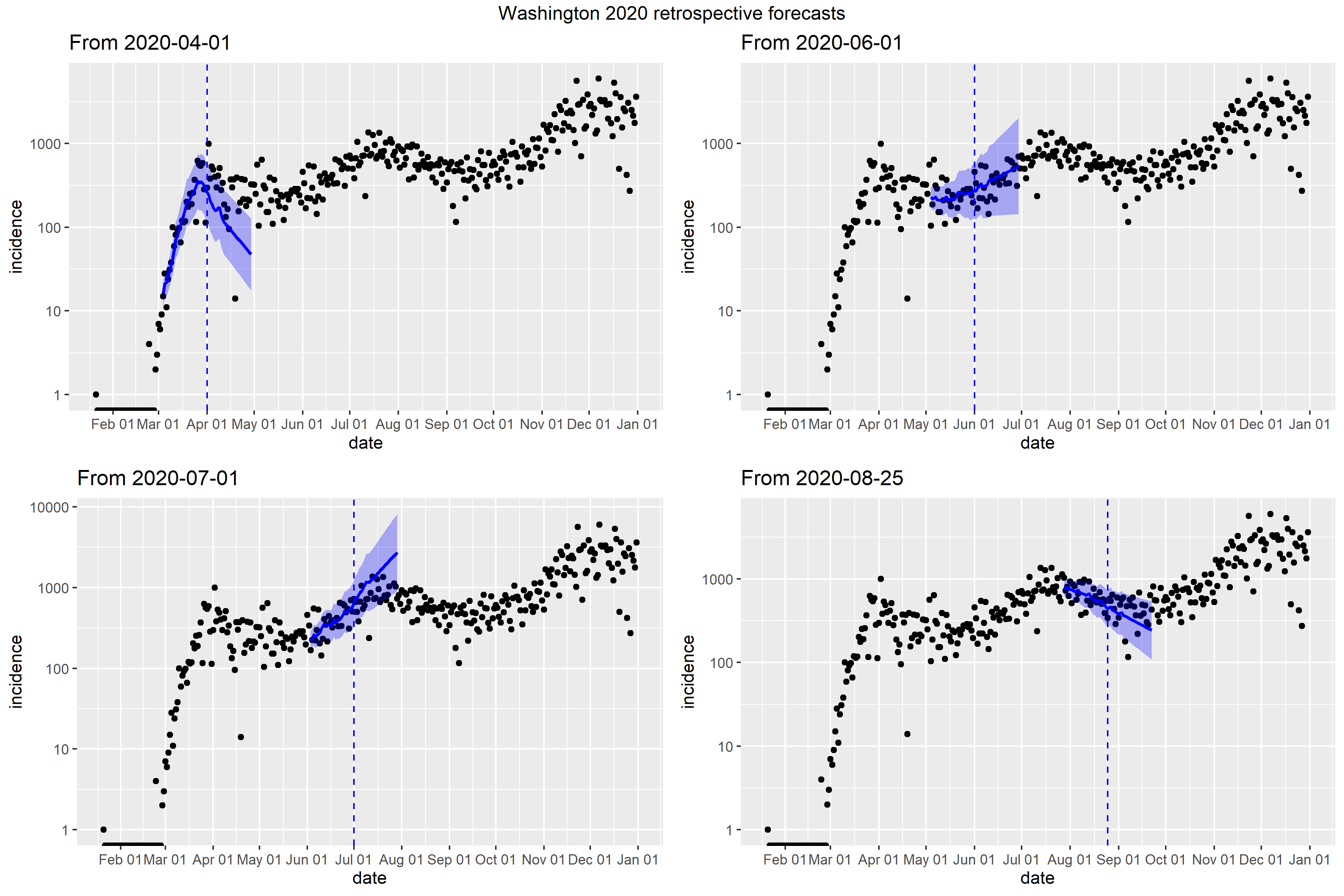

### West Virginia_retrospective_forecasts.png

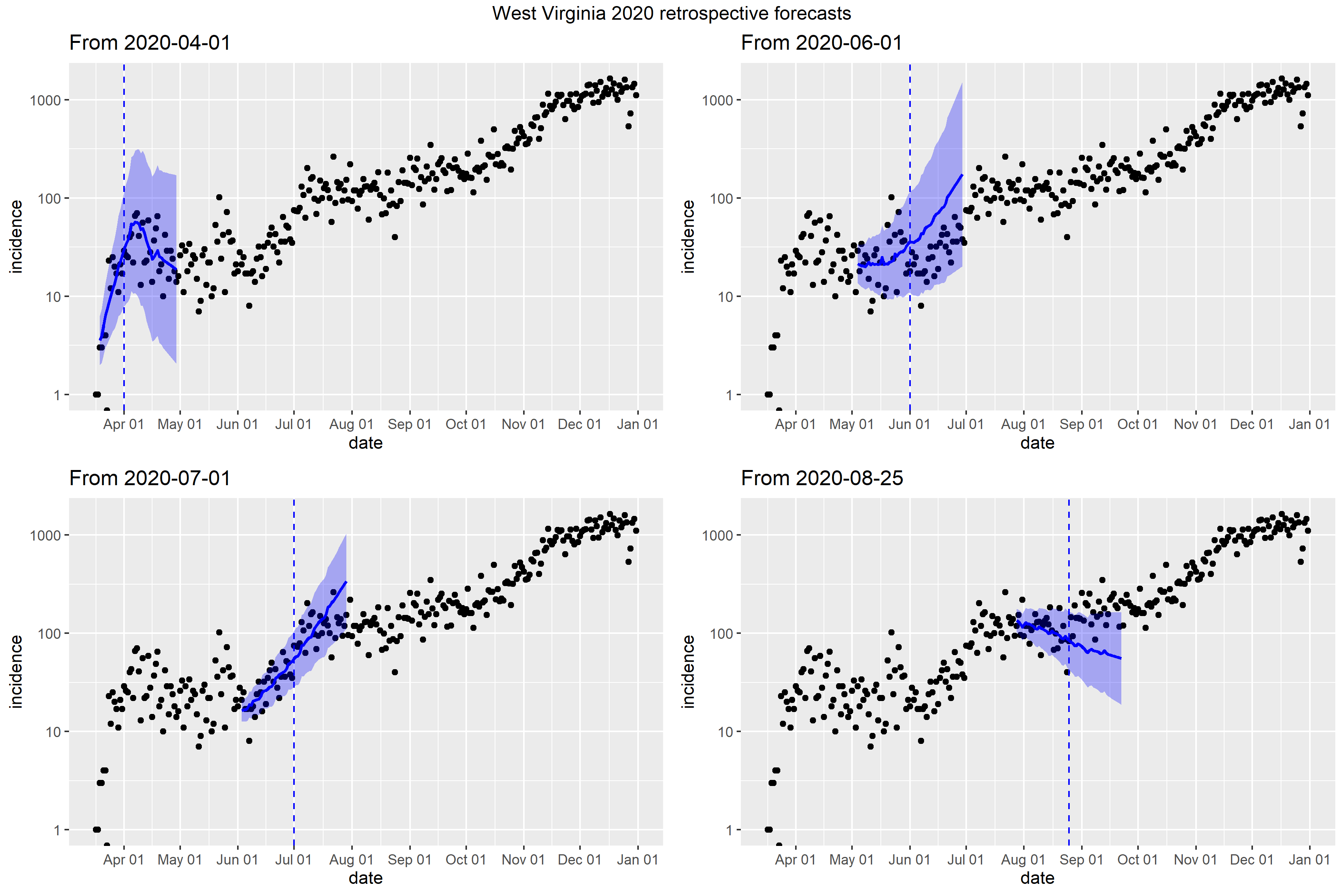

### Wisconsin_Peff_plot.png

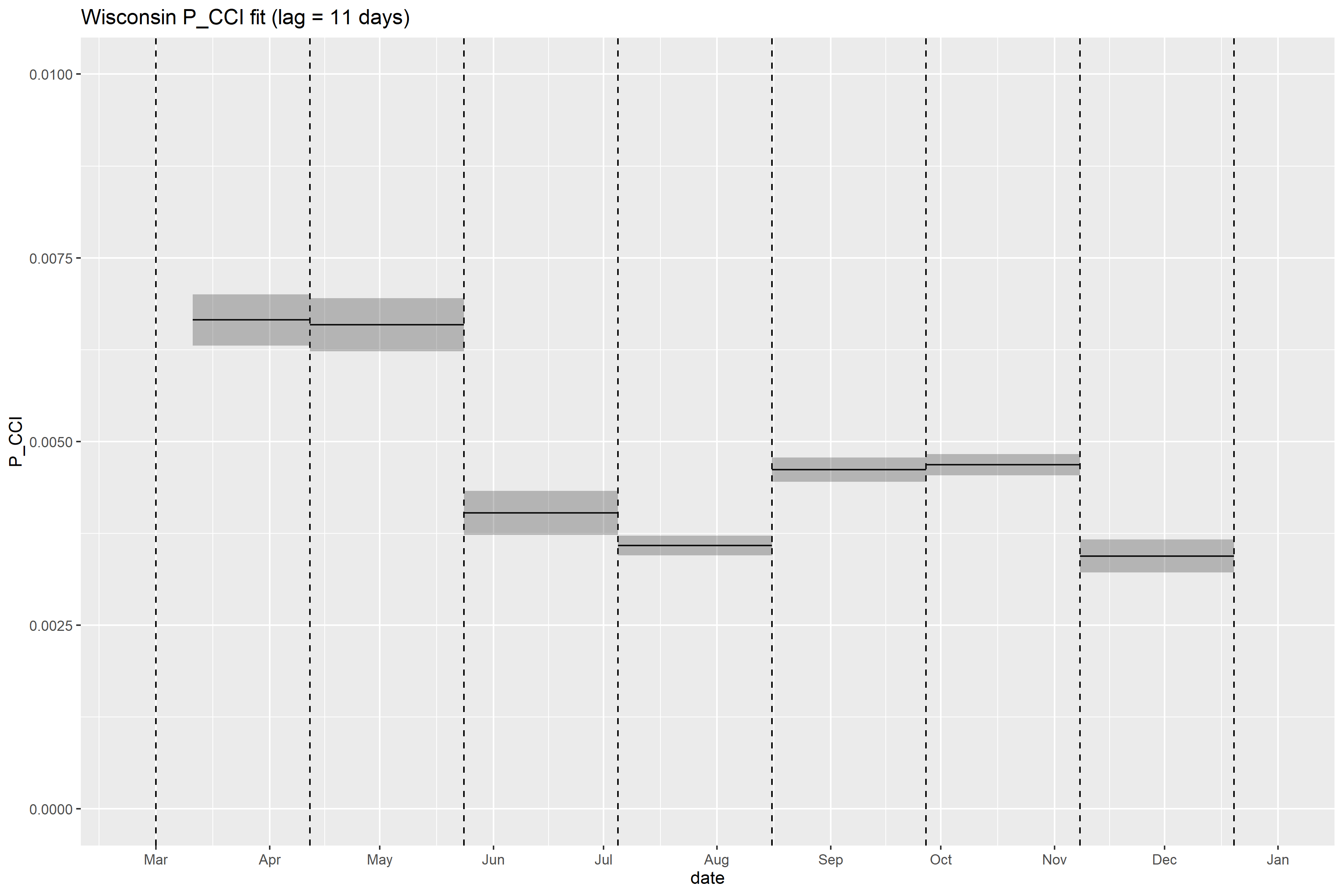

### Wisconsin_retrospective_forecasts.png

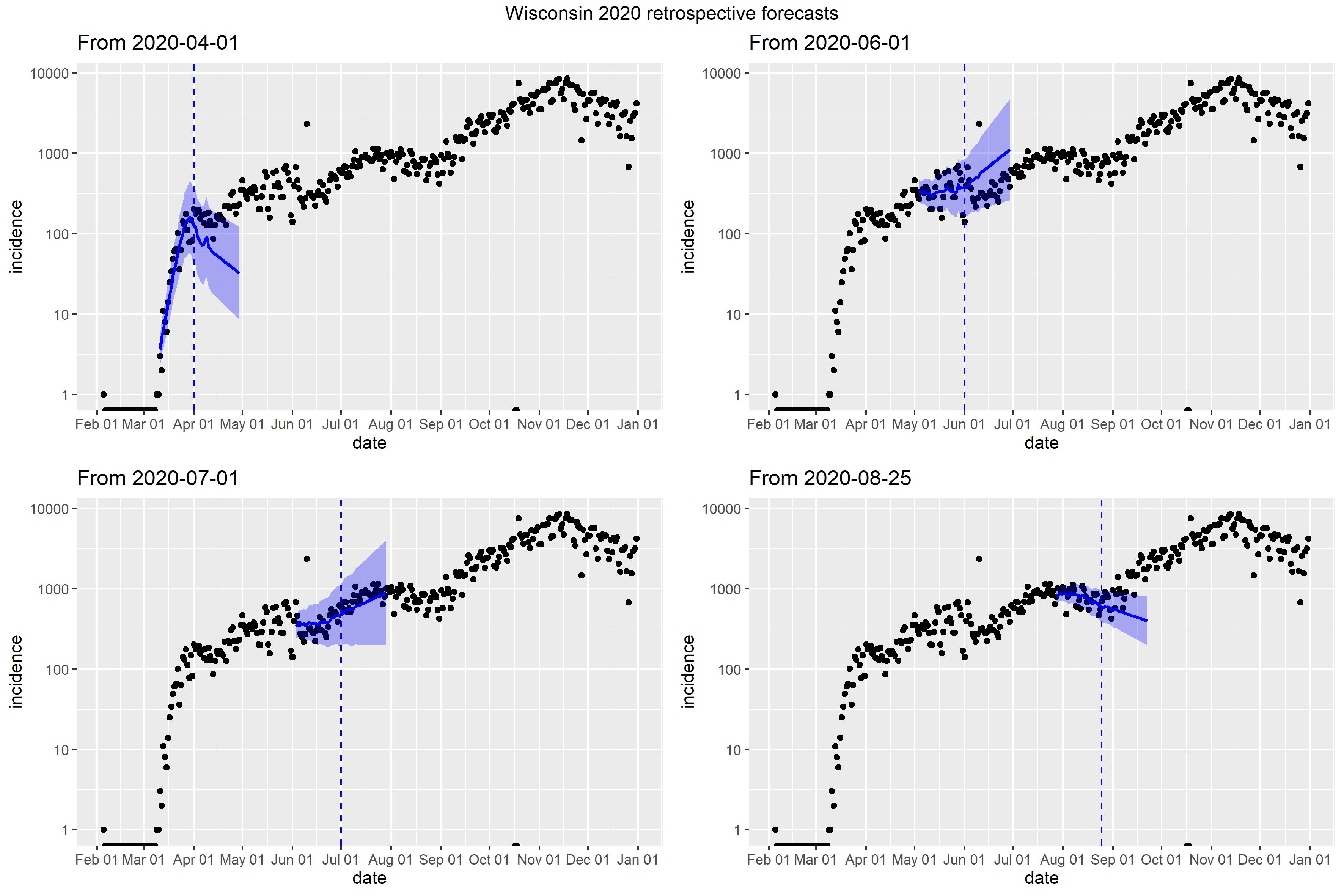

### Wyoming_Peff_plot.png

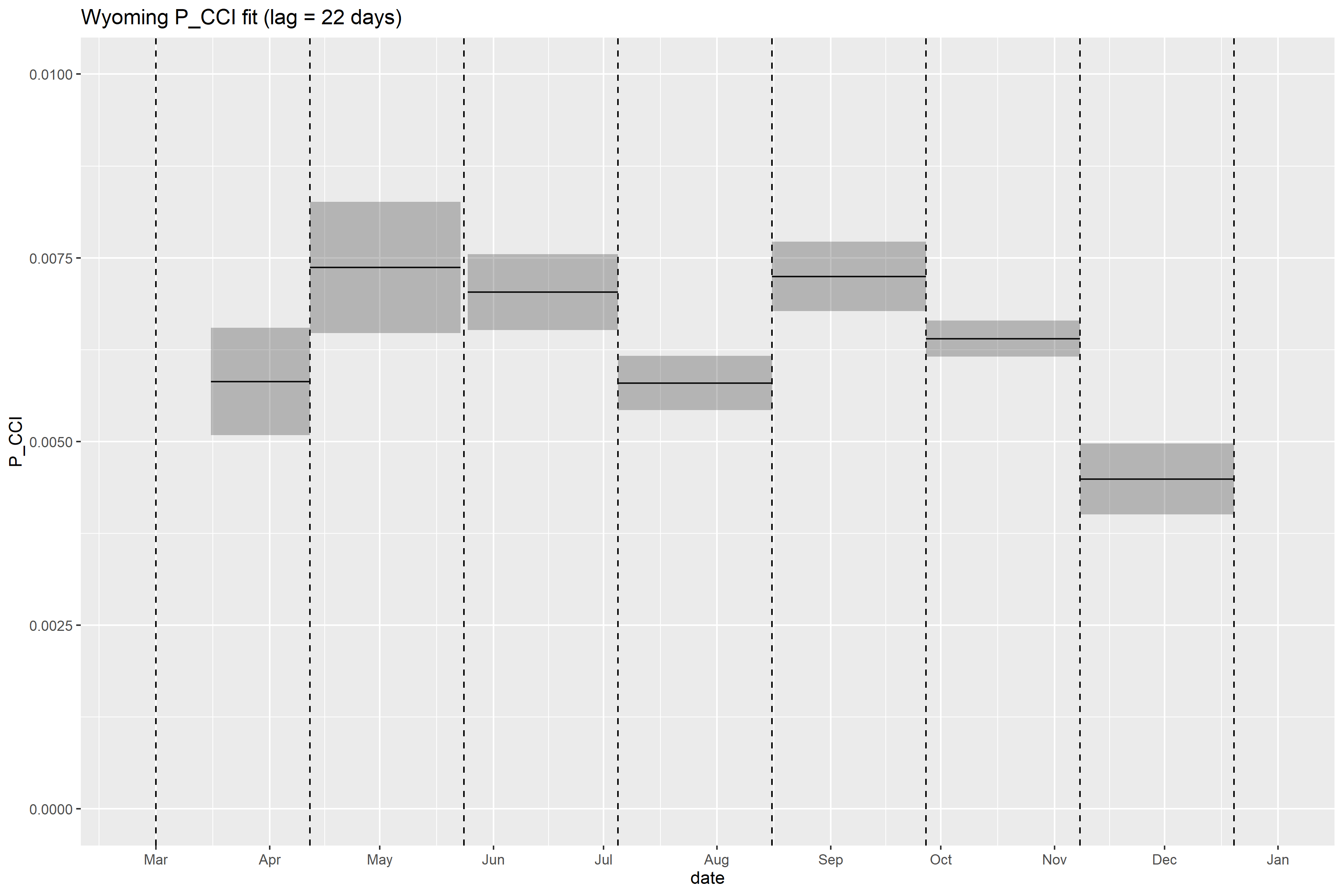

### Wyoming_retrospective_forecasts.png

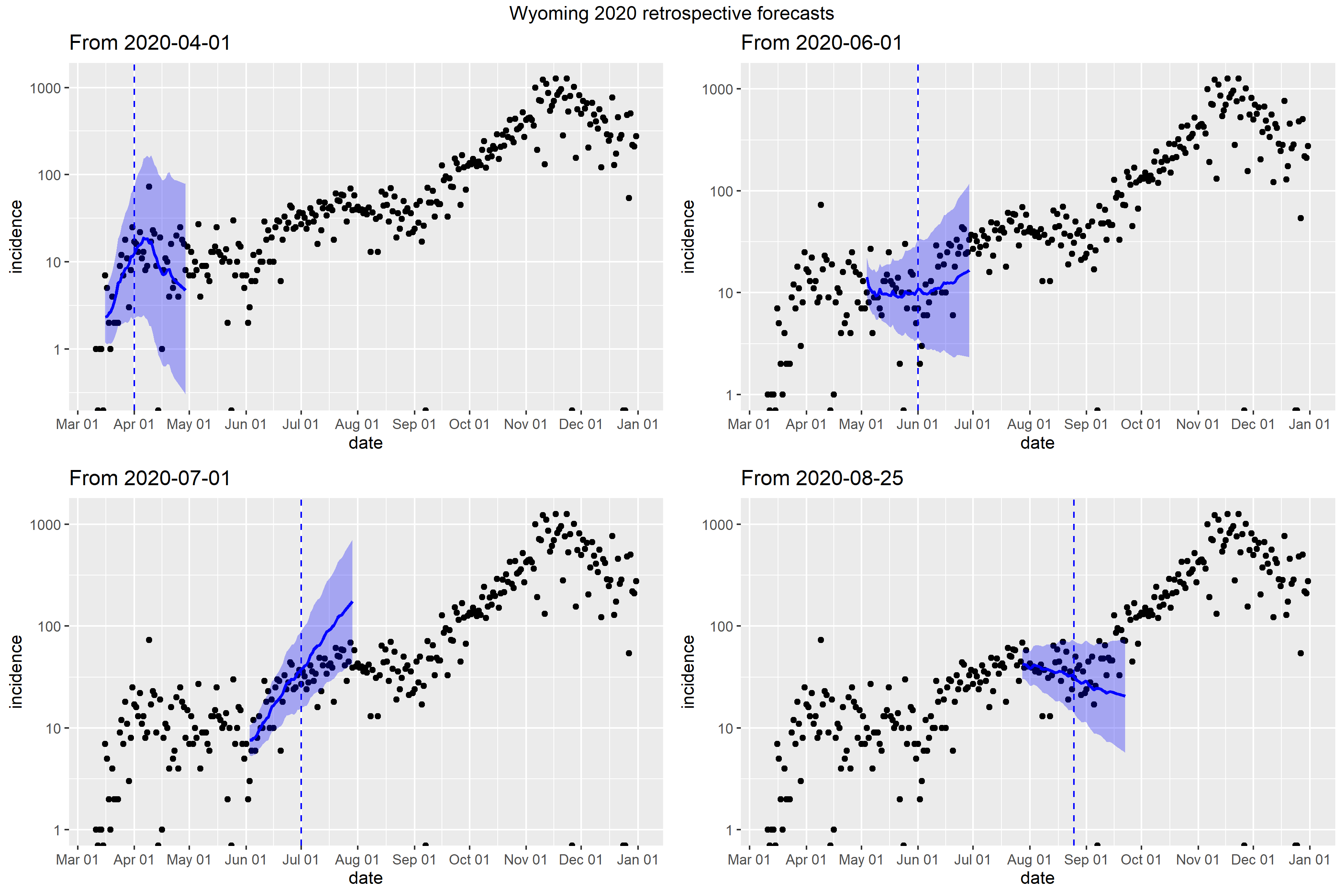
