## Supplementary figures and images for "U.S. state-level COVID-19 transmission insights from a mechanistic mobility-incidence model"

### Alabama_full_fit_plot.png

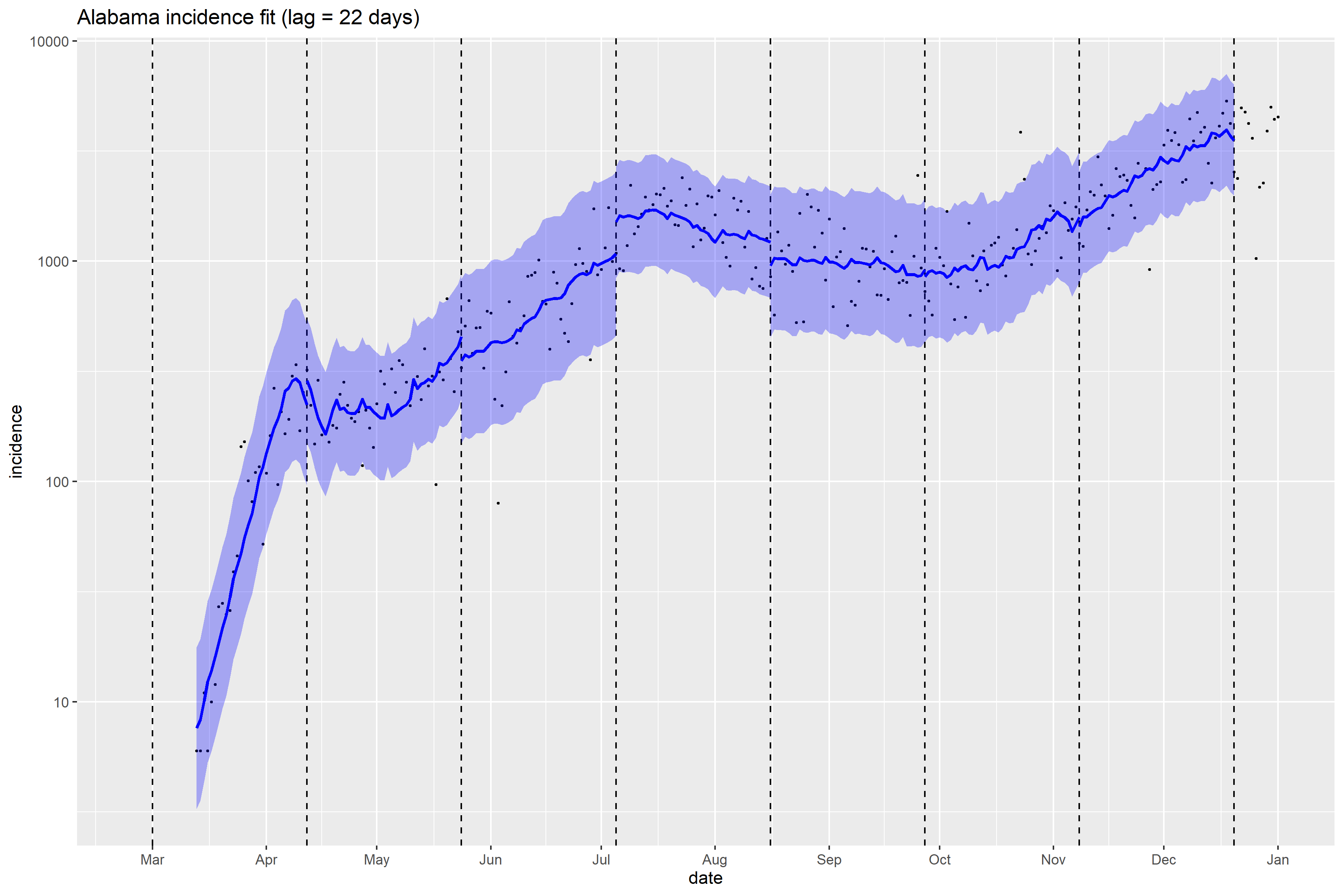

### Arizona_full_fit_plot.png

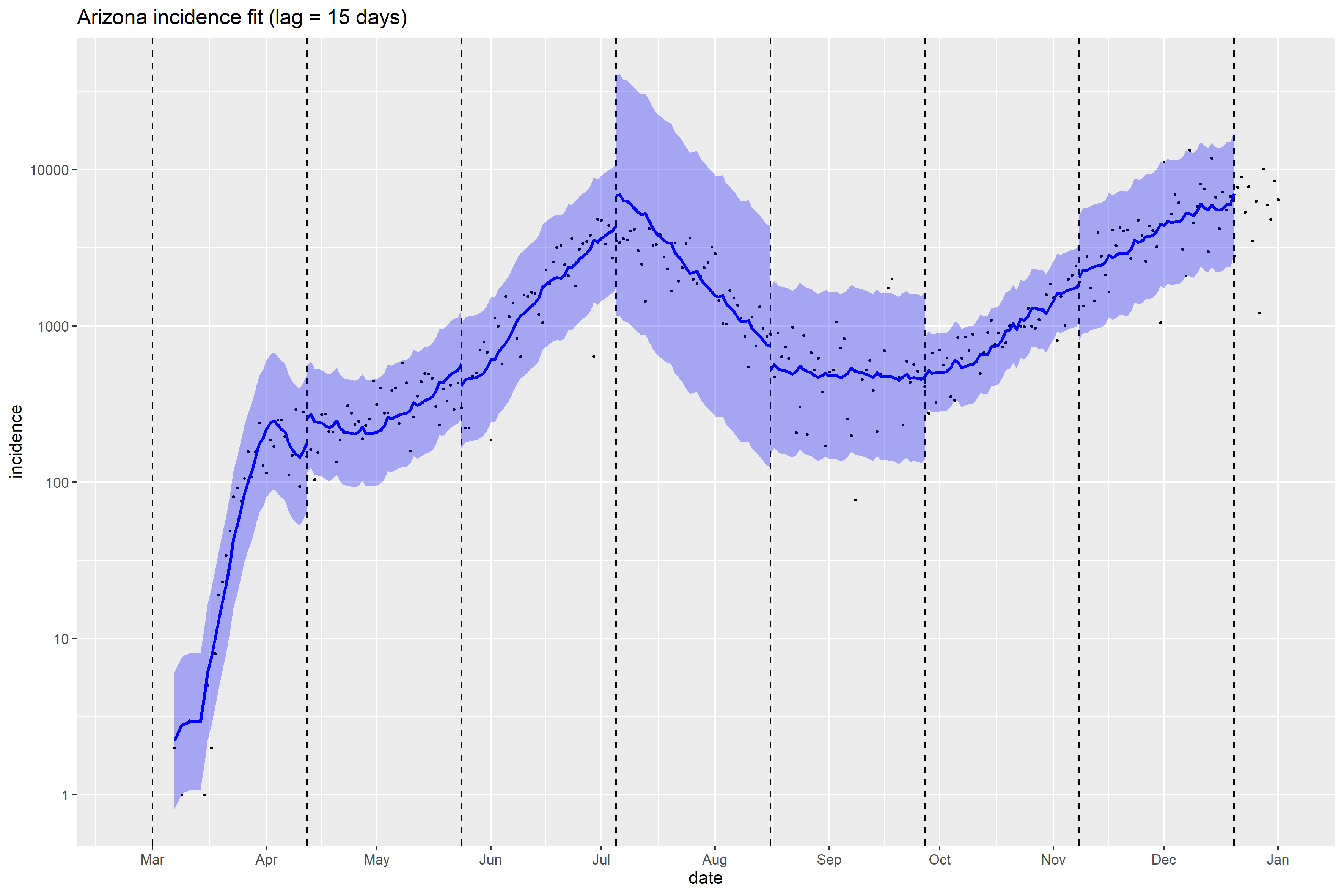

### Arkansas_full_fit_plot.png

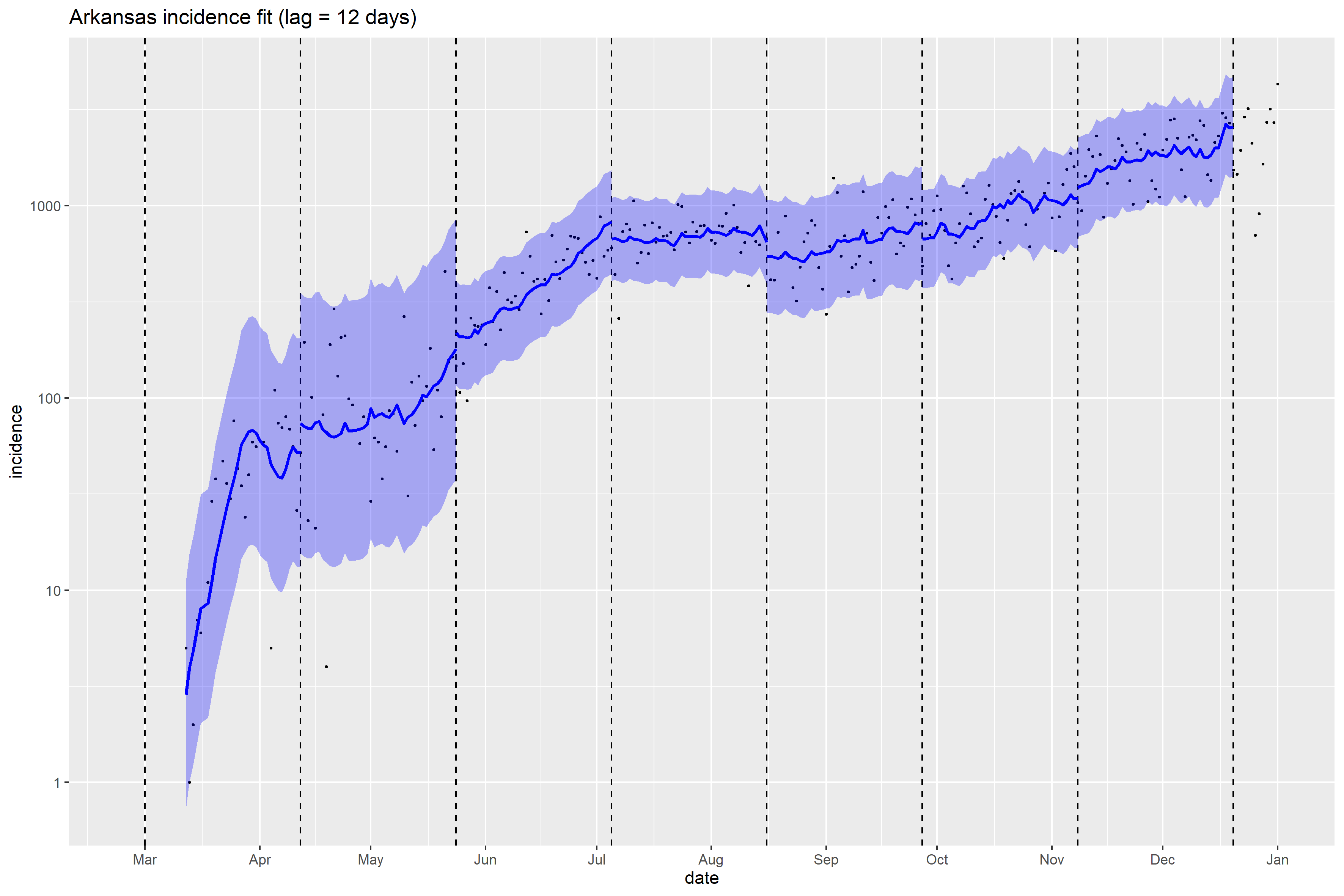

### California_full_fit_plot.png

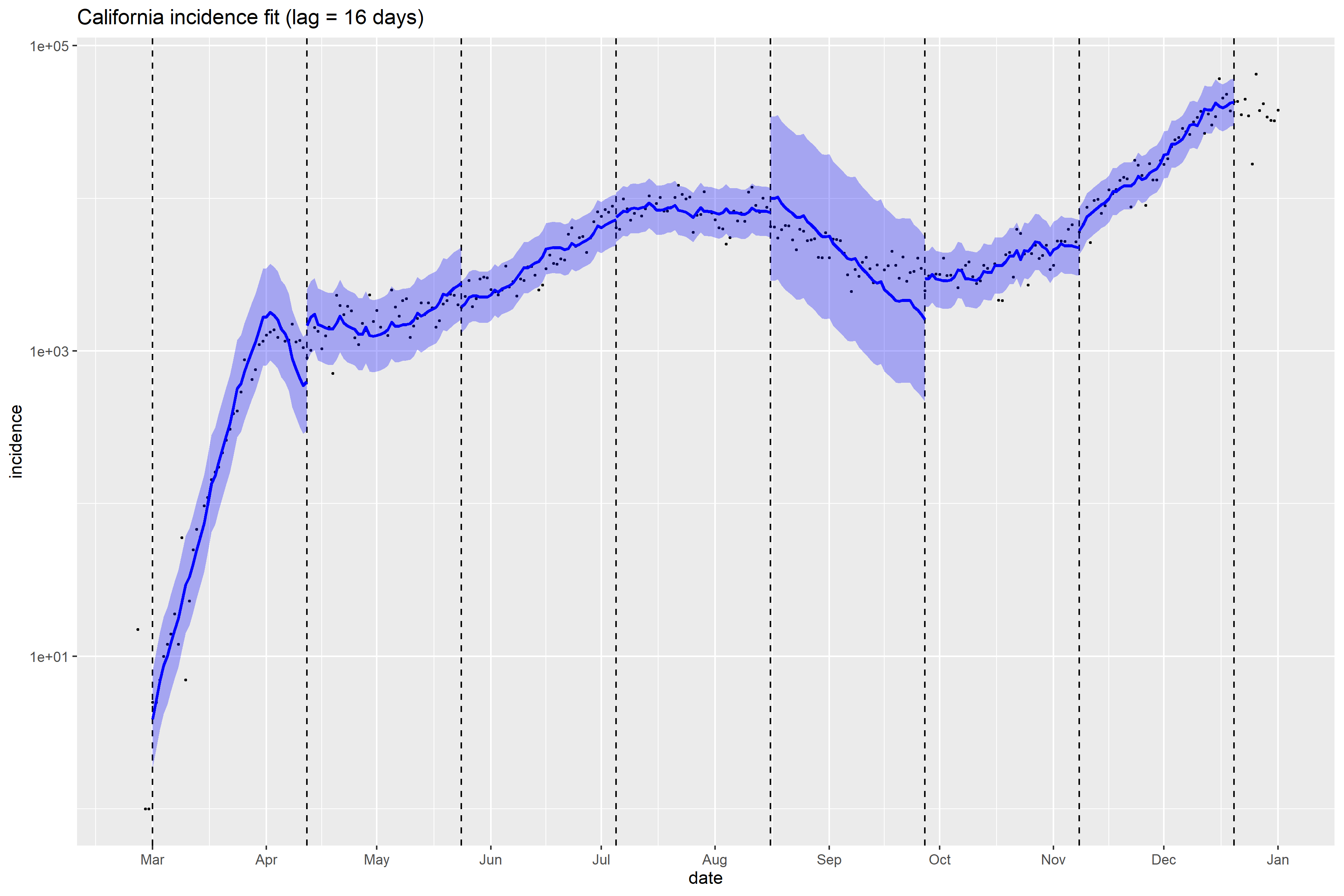

### Colorado_full_fit_plot.png

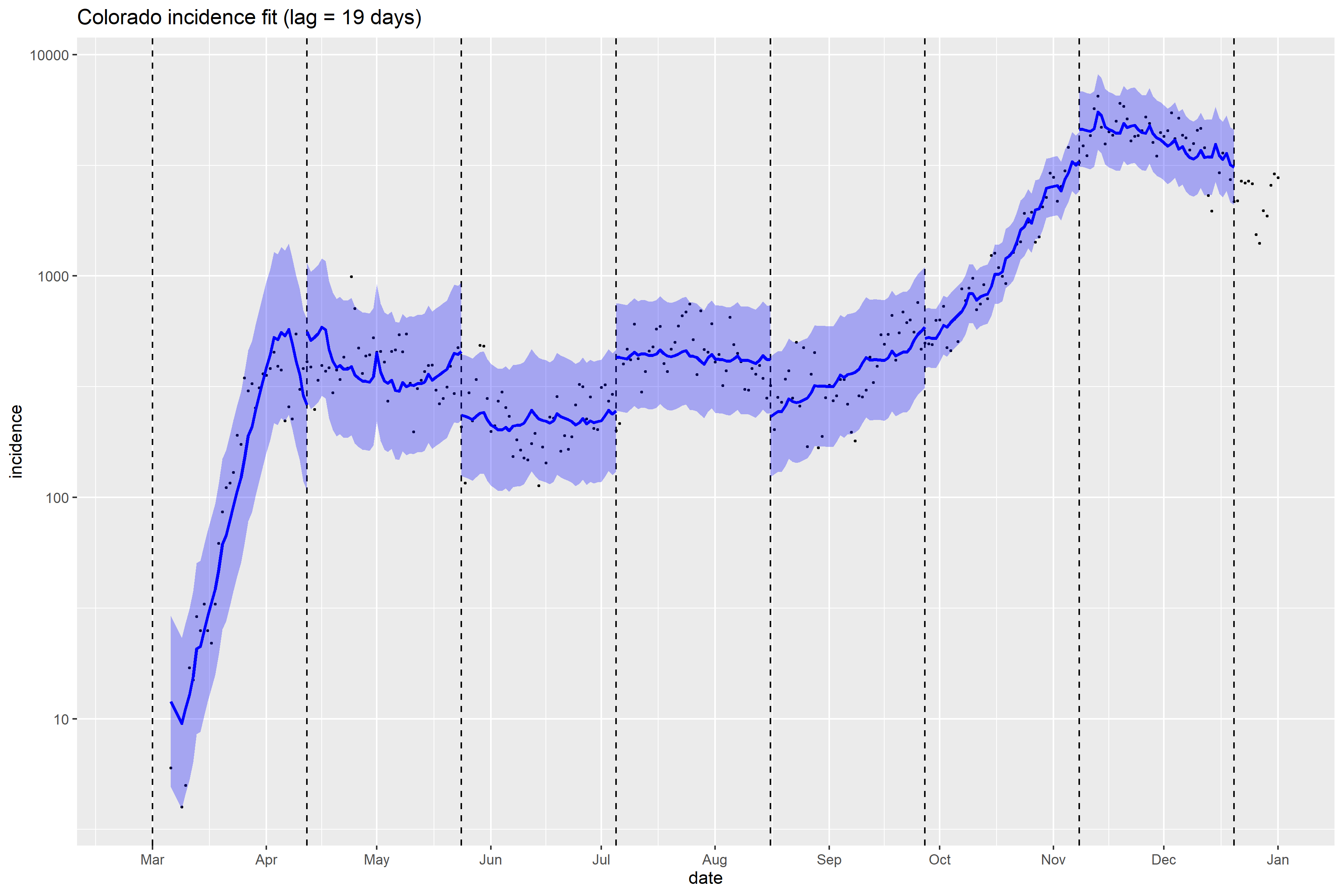

### Connecticut_full_fit_plot.png

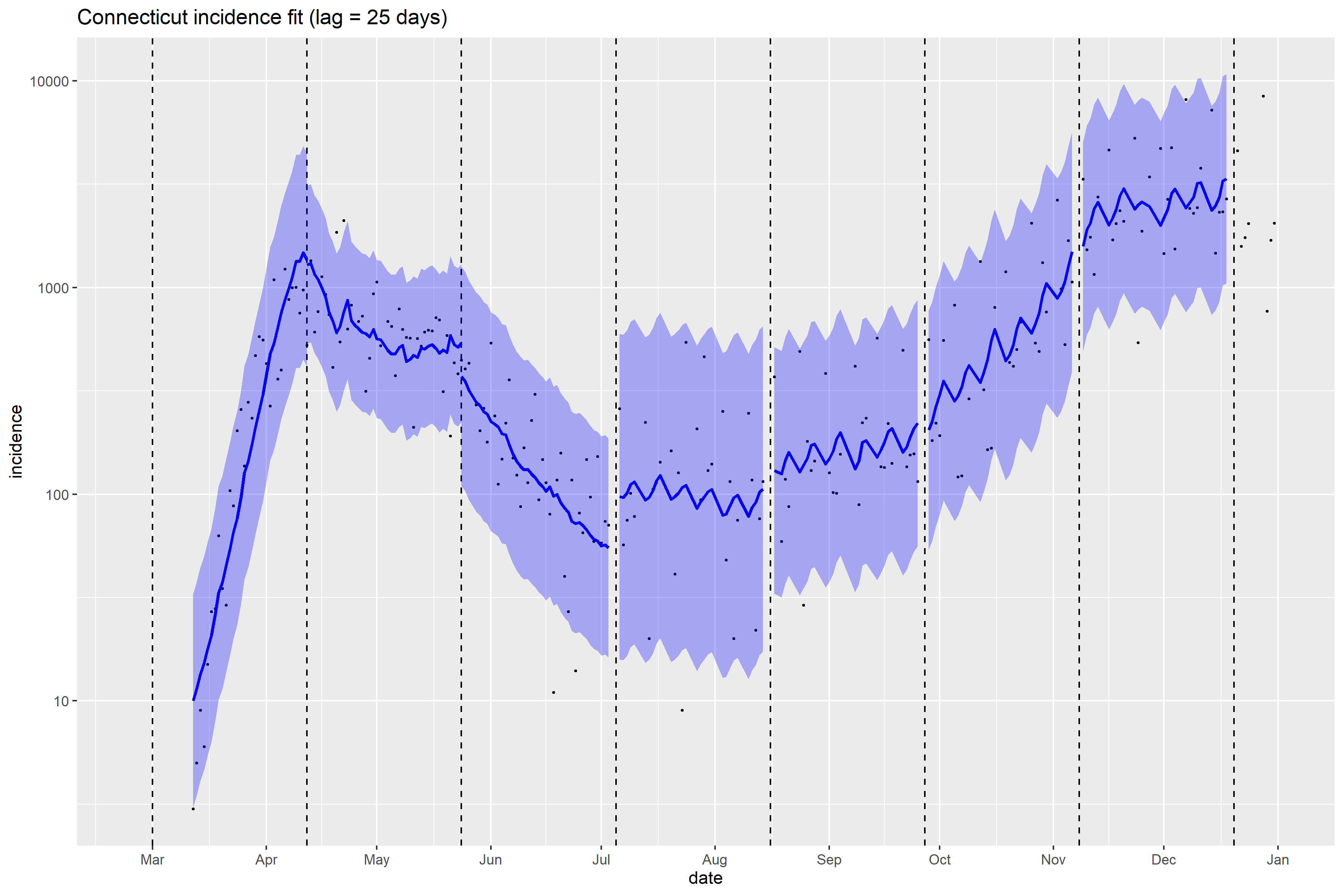

### Connecticut_Peff_plot.png

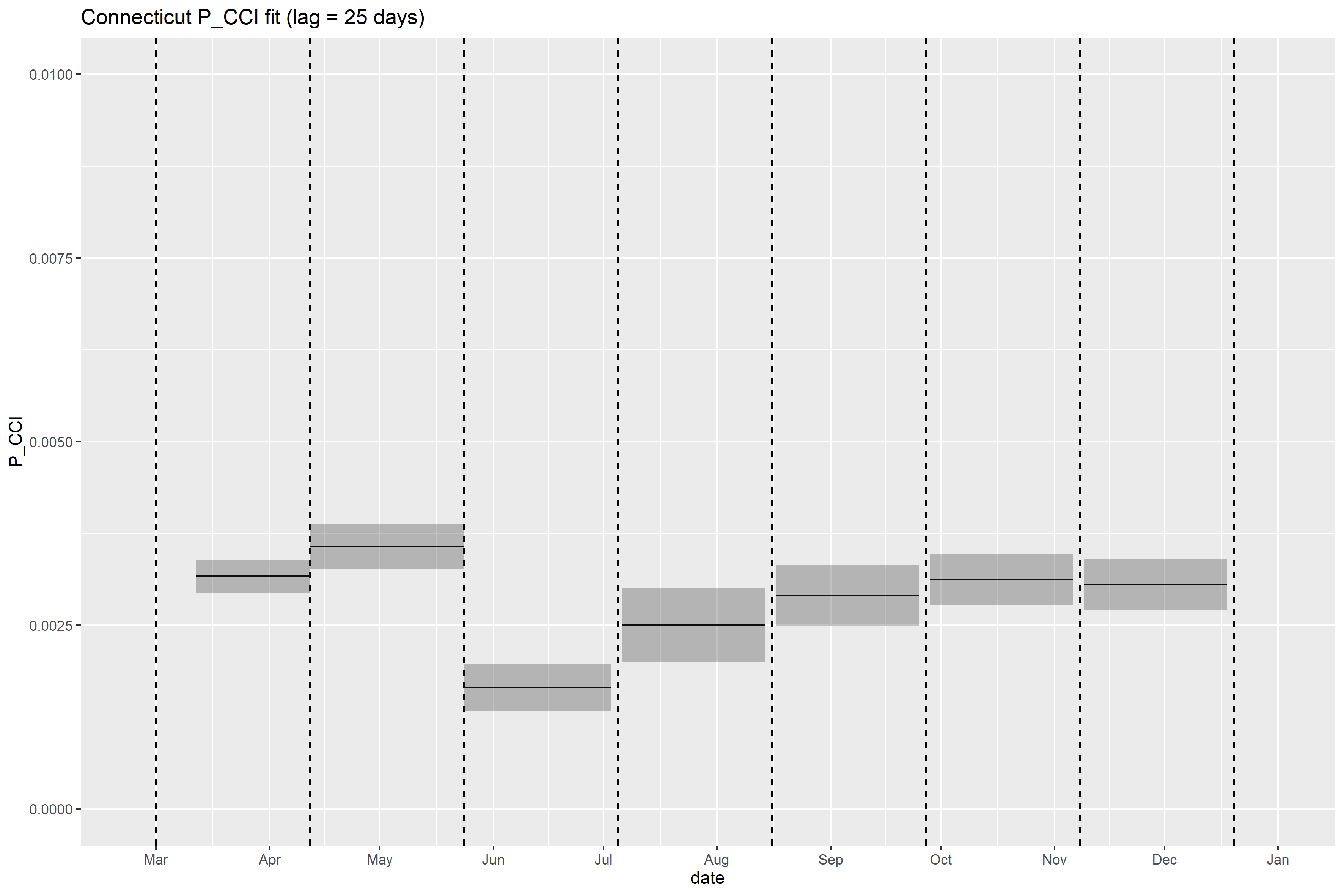

### Delaware_full_fit_plot.png

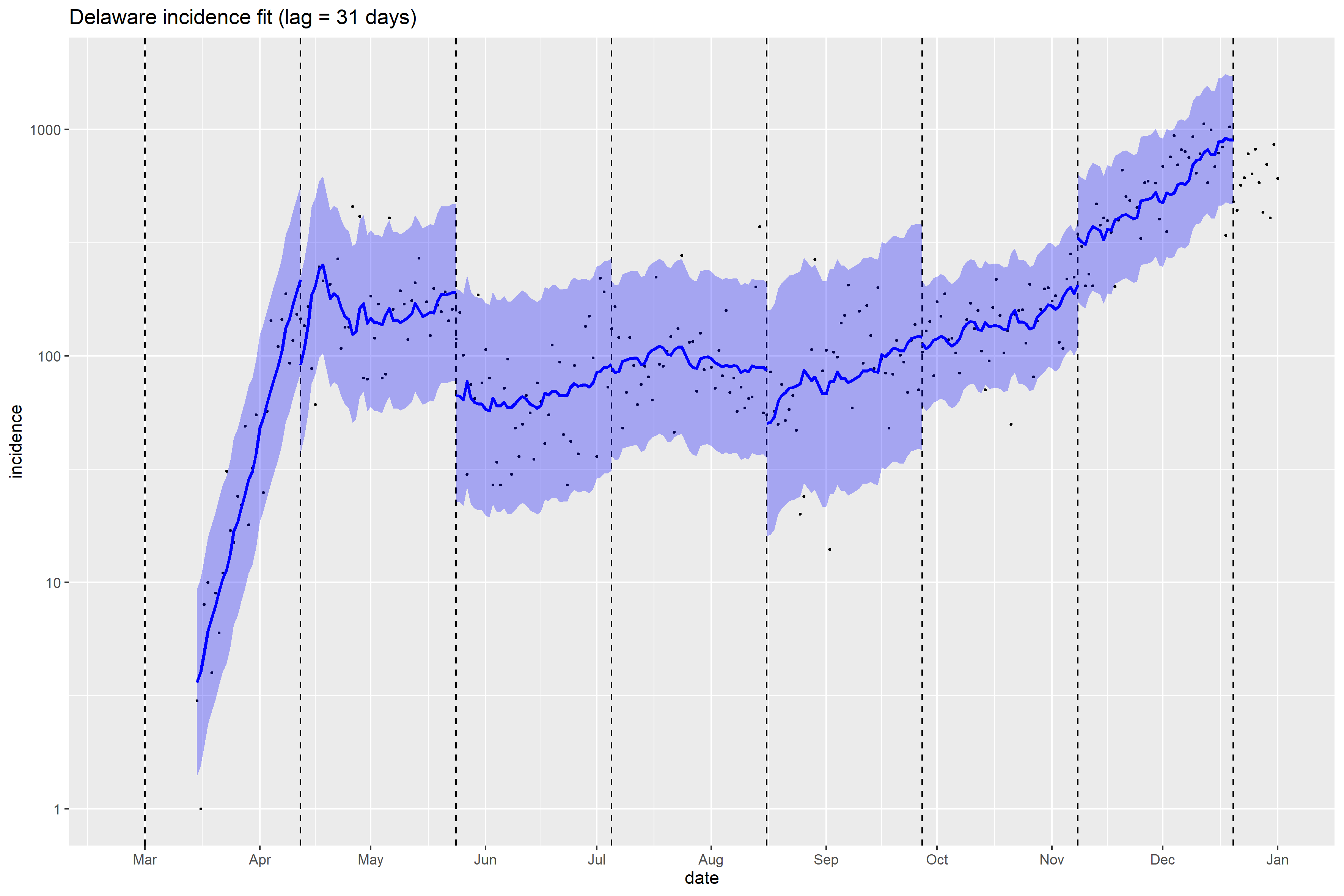

### District of Columbia_full_fit_plot.png

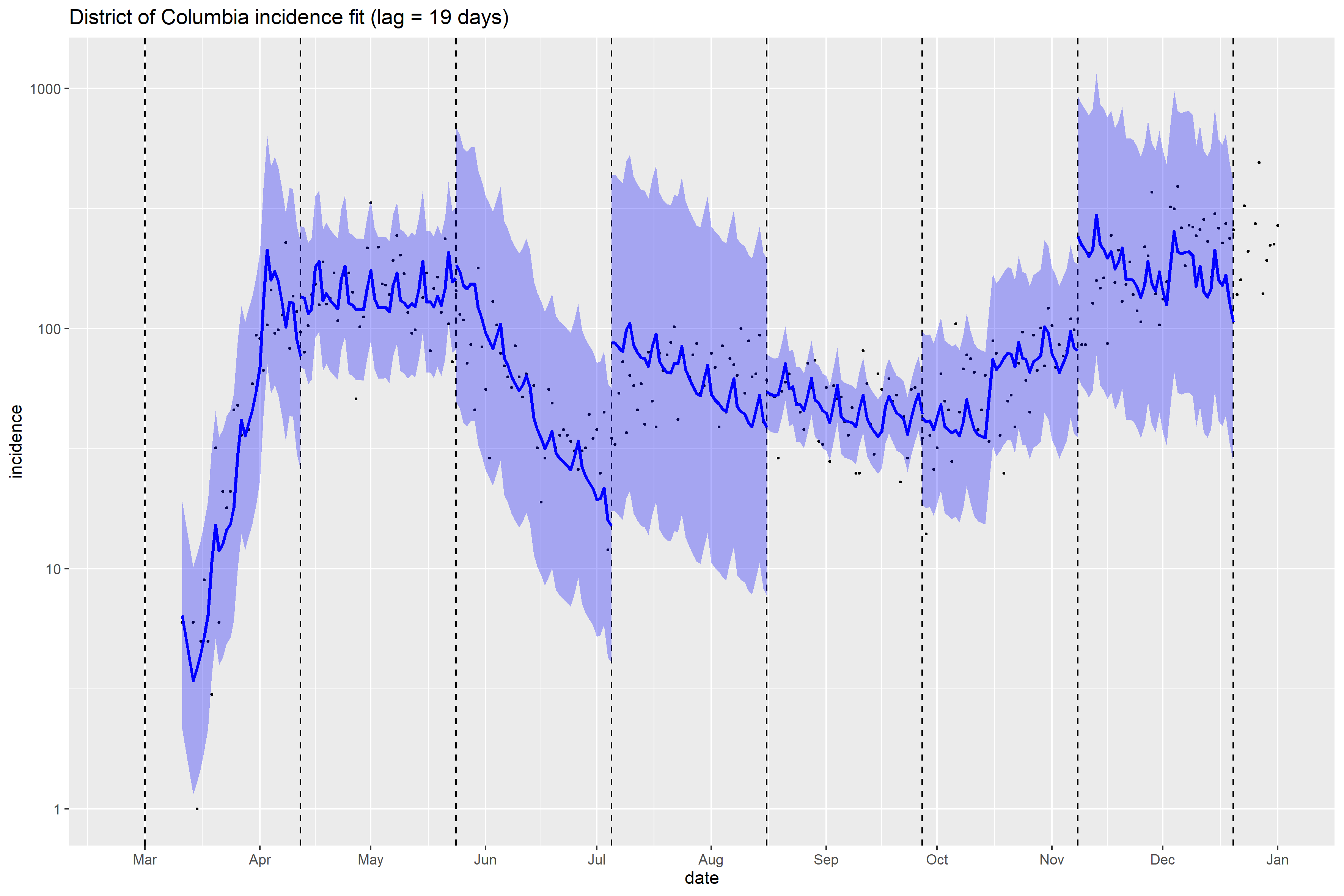

### District of Columbia_Peff_plot.png

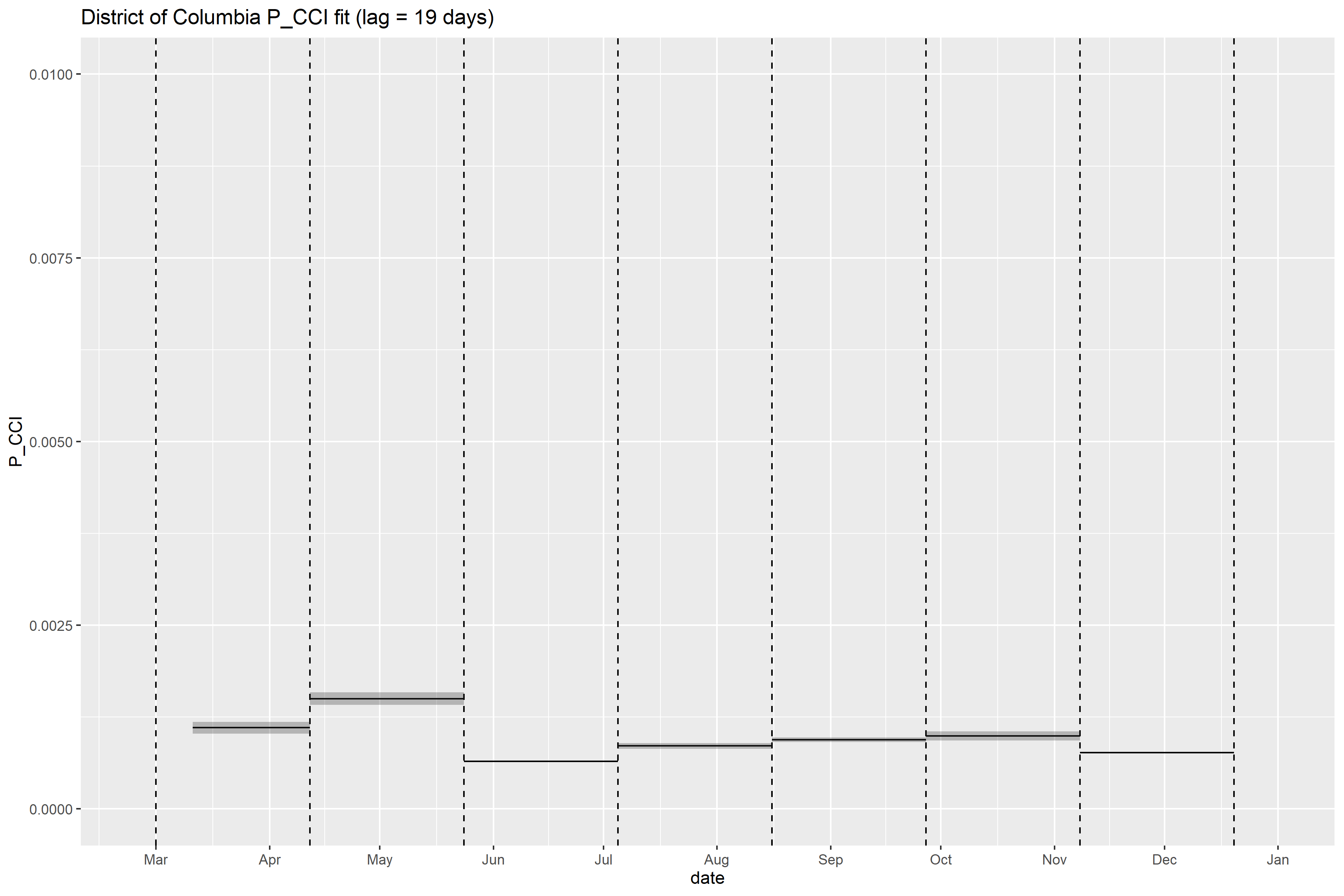

### Florida_full_fit_plot.png

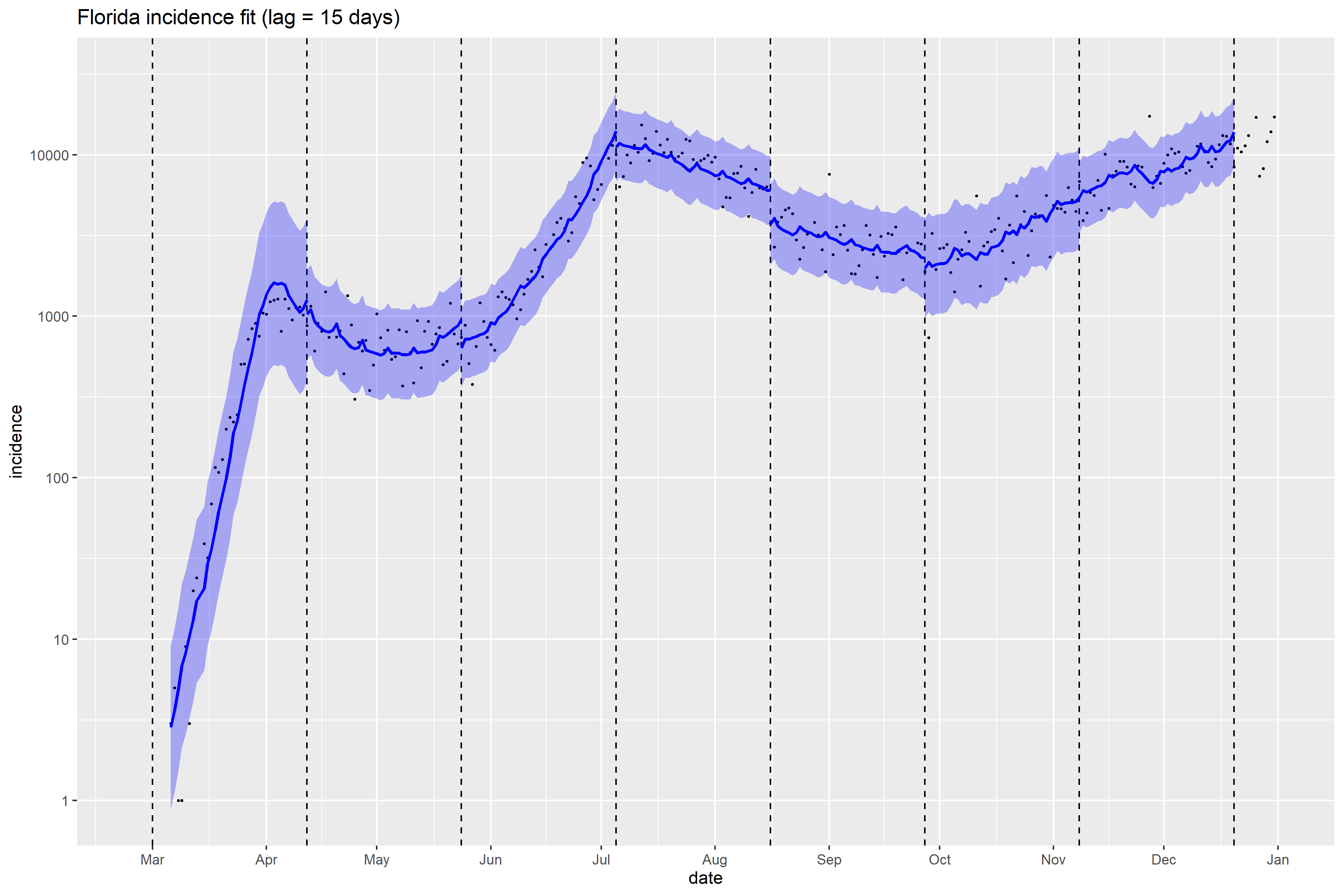

### Georgia_full_fit_plot.png

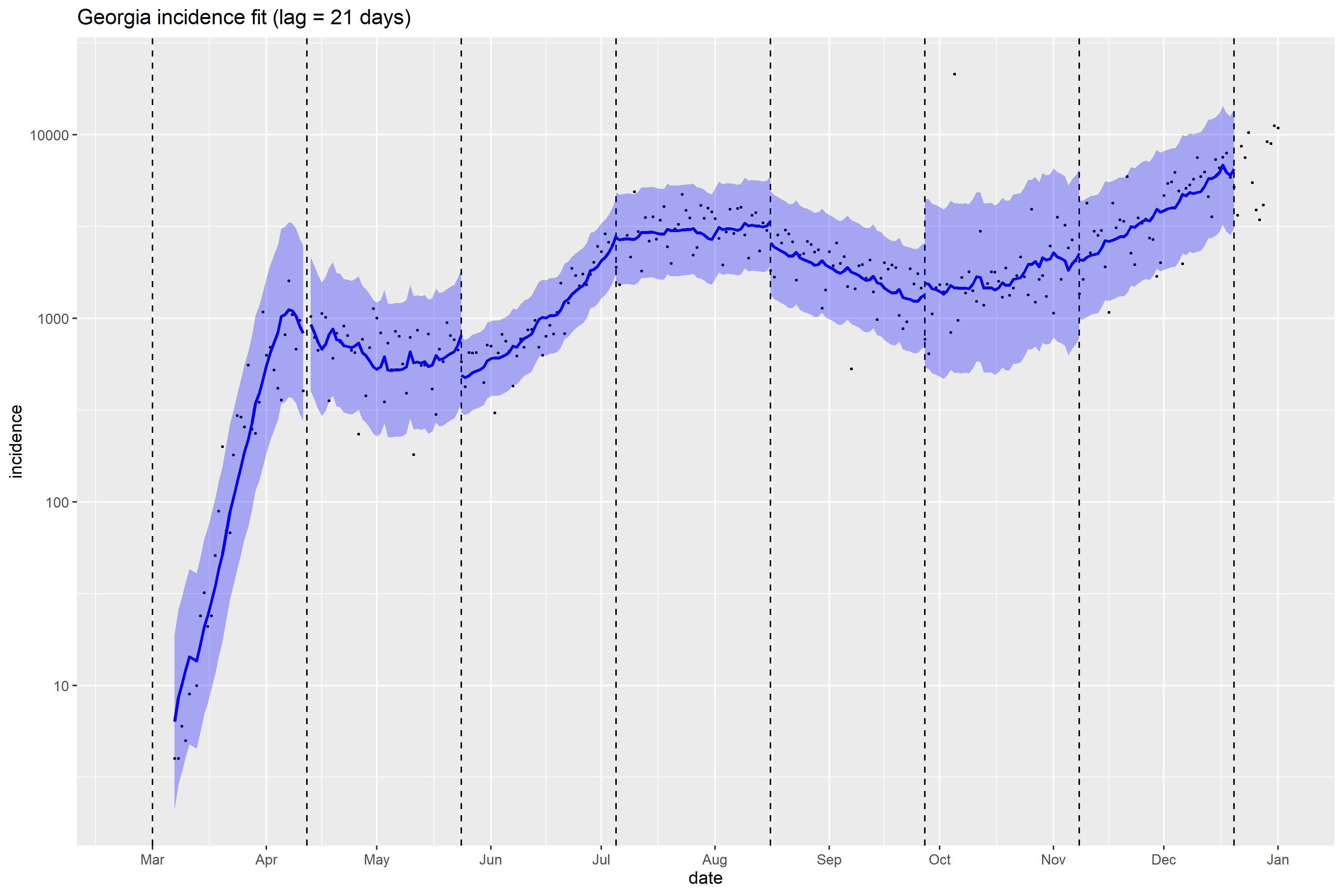

### Illinois_full_fit_plot.png

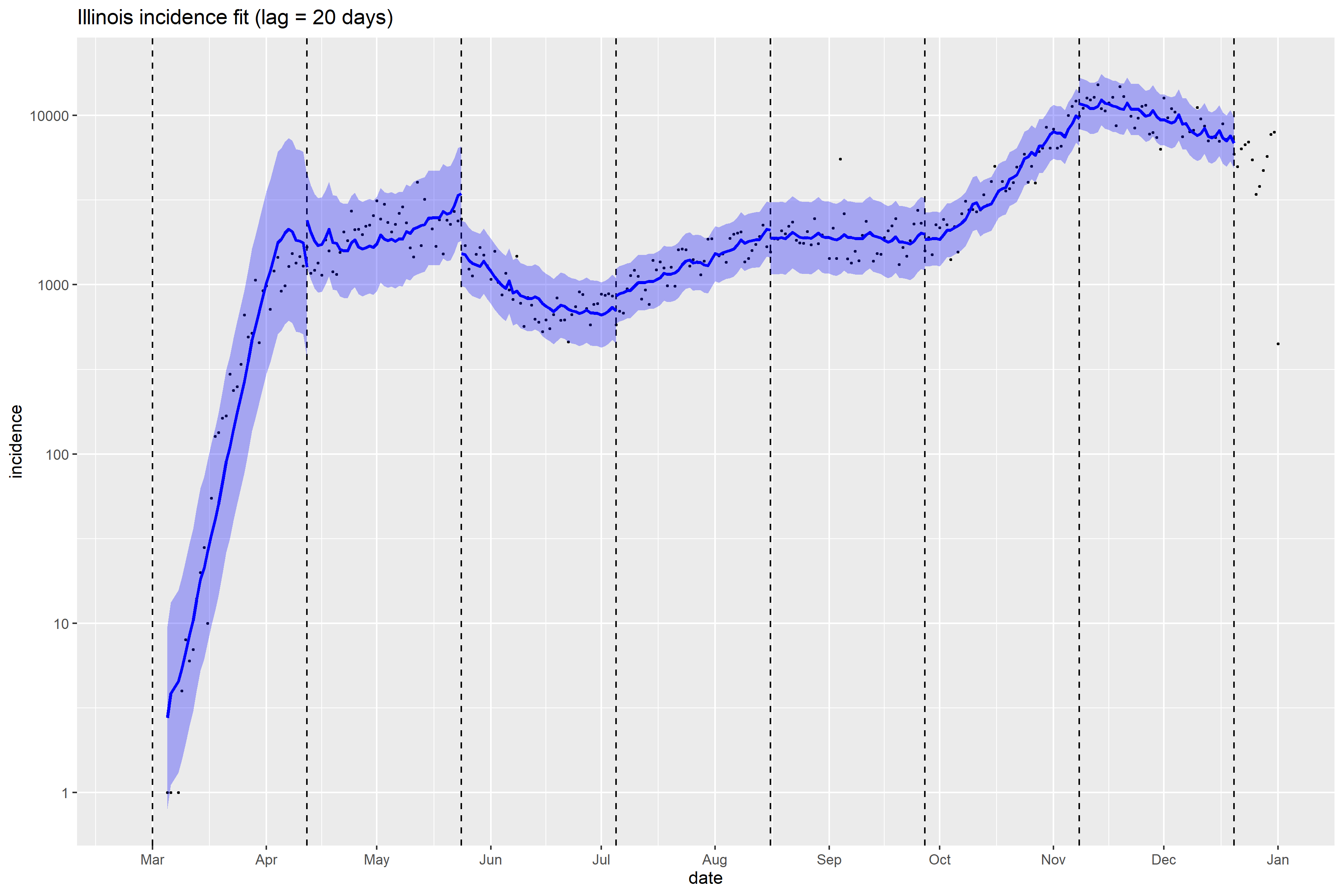

### Indiana_full_fit_plot.png

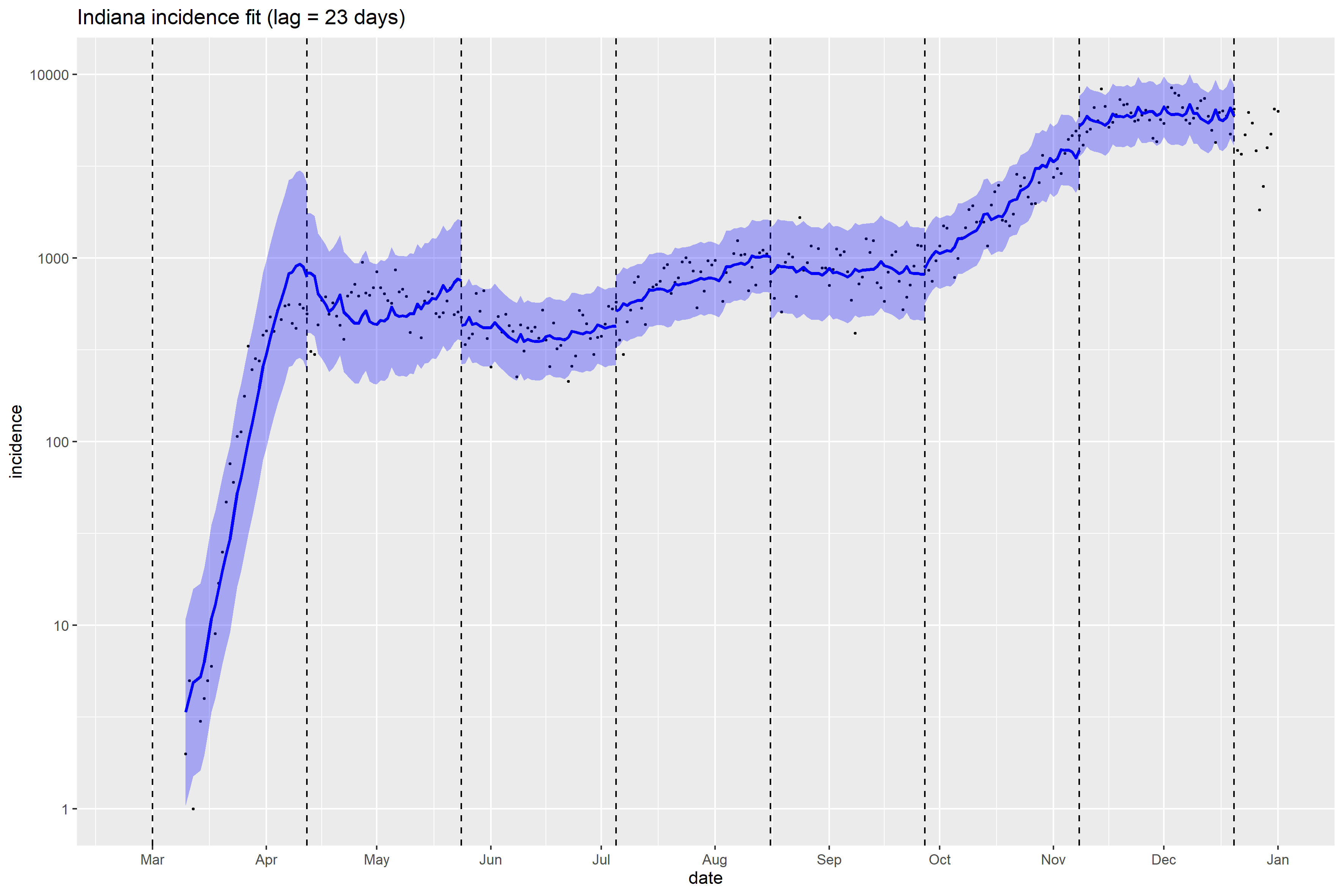

### Kentucky_full_fit_plot.png

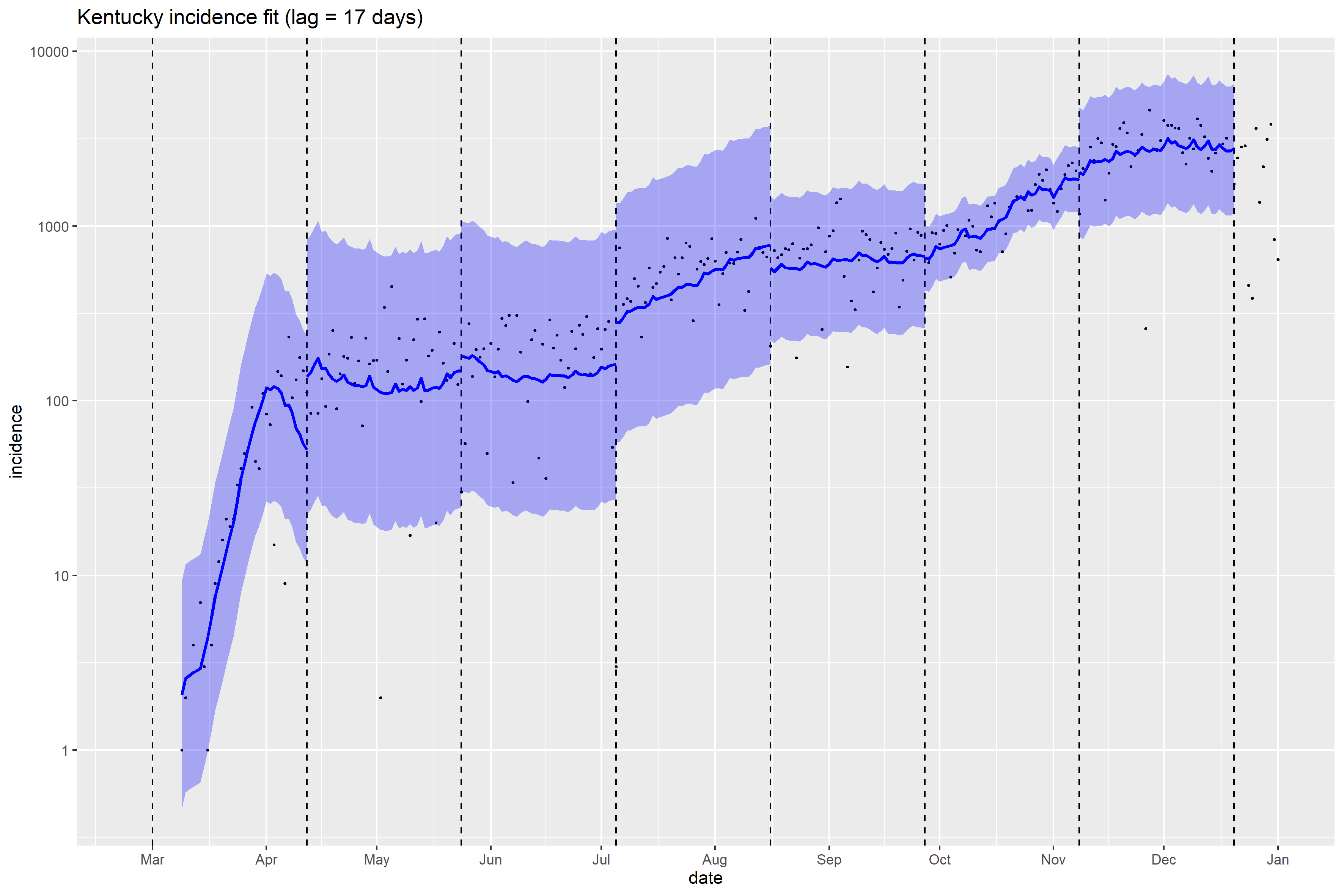

### Louisiana_full_fit_plot.png

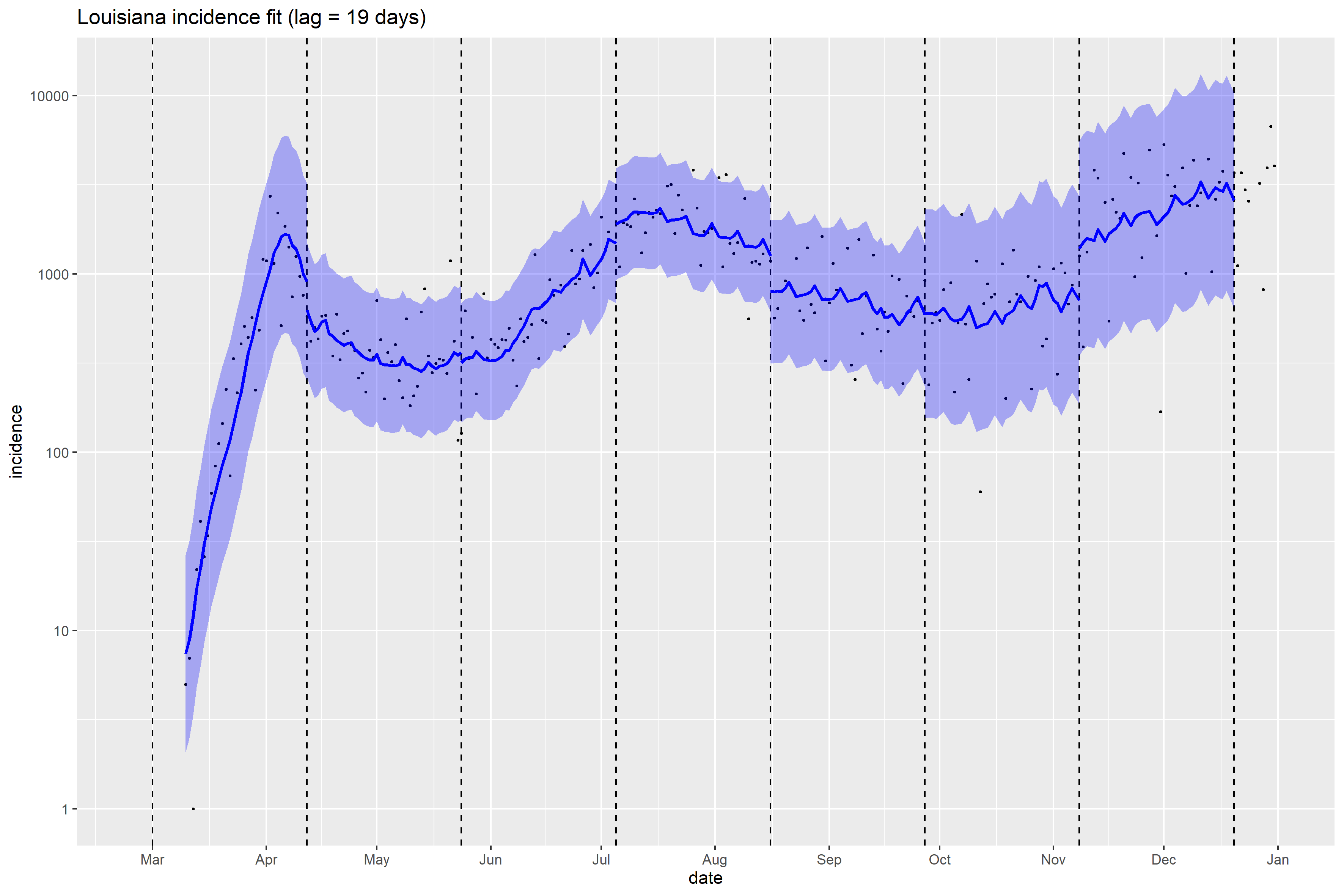

### Maryland_full_fit_plot.png

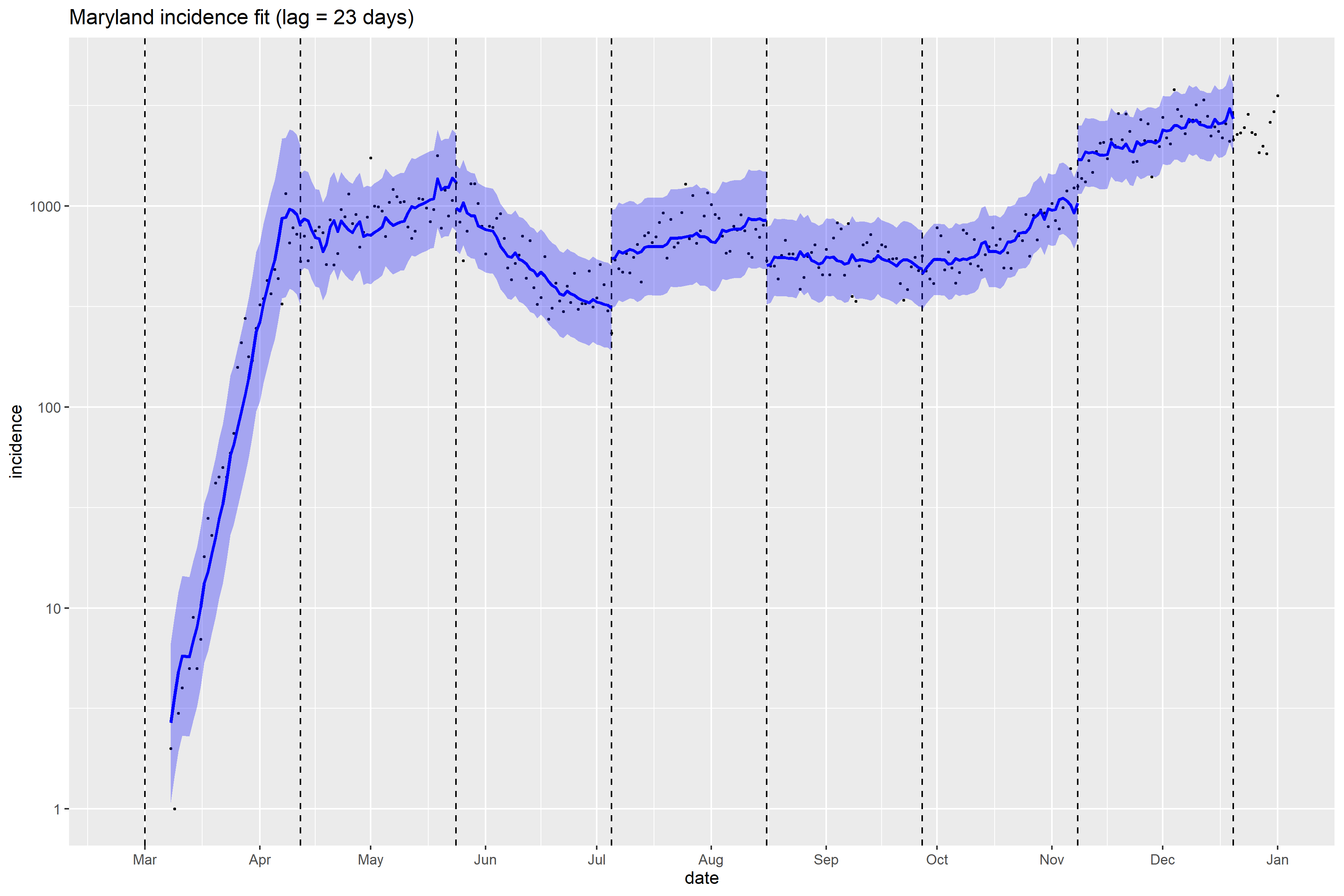

### Massachusetts_full_fit_plot.png

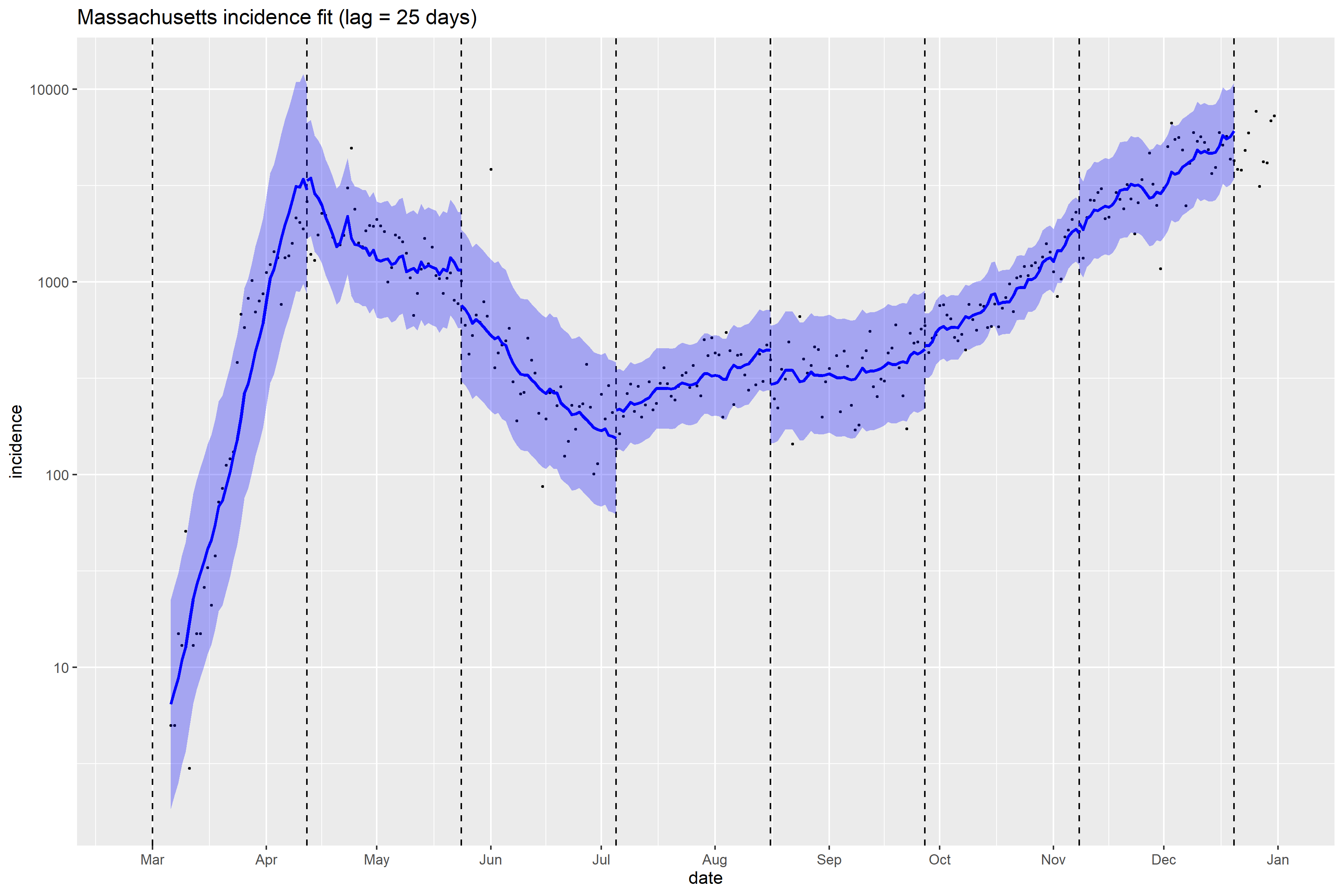

### Massachusetts_Peff_plot.png

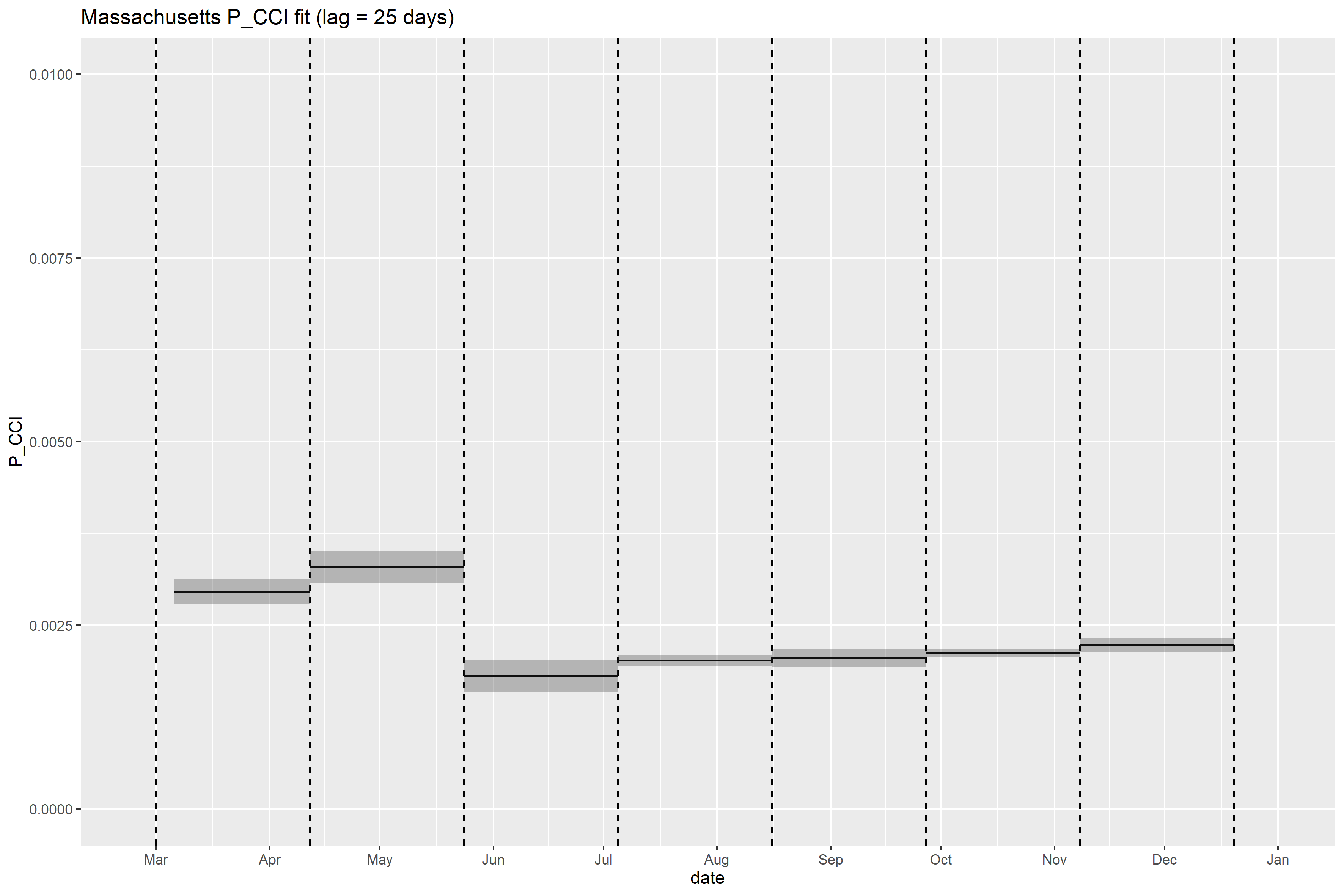

### Michigan_full_fit_plot.png

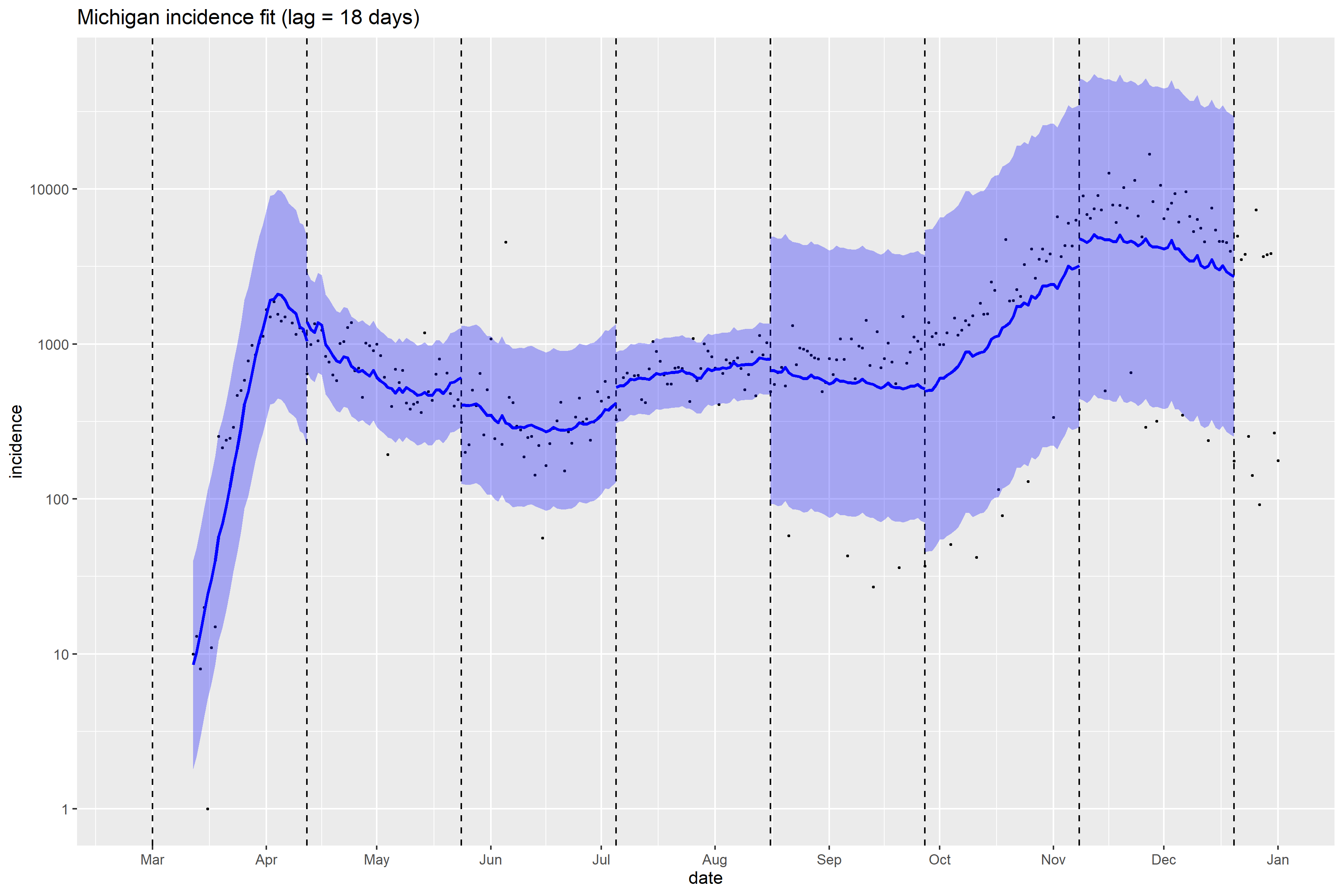

### Minnesota_full_fit_plot.png

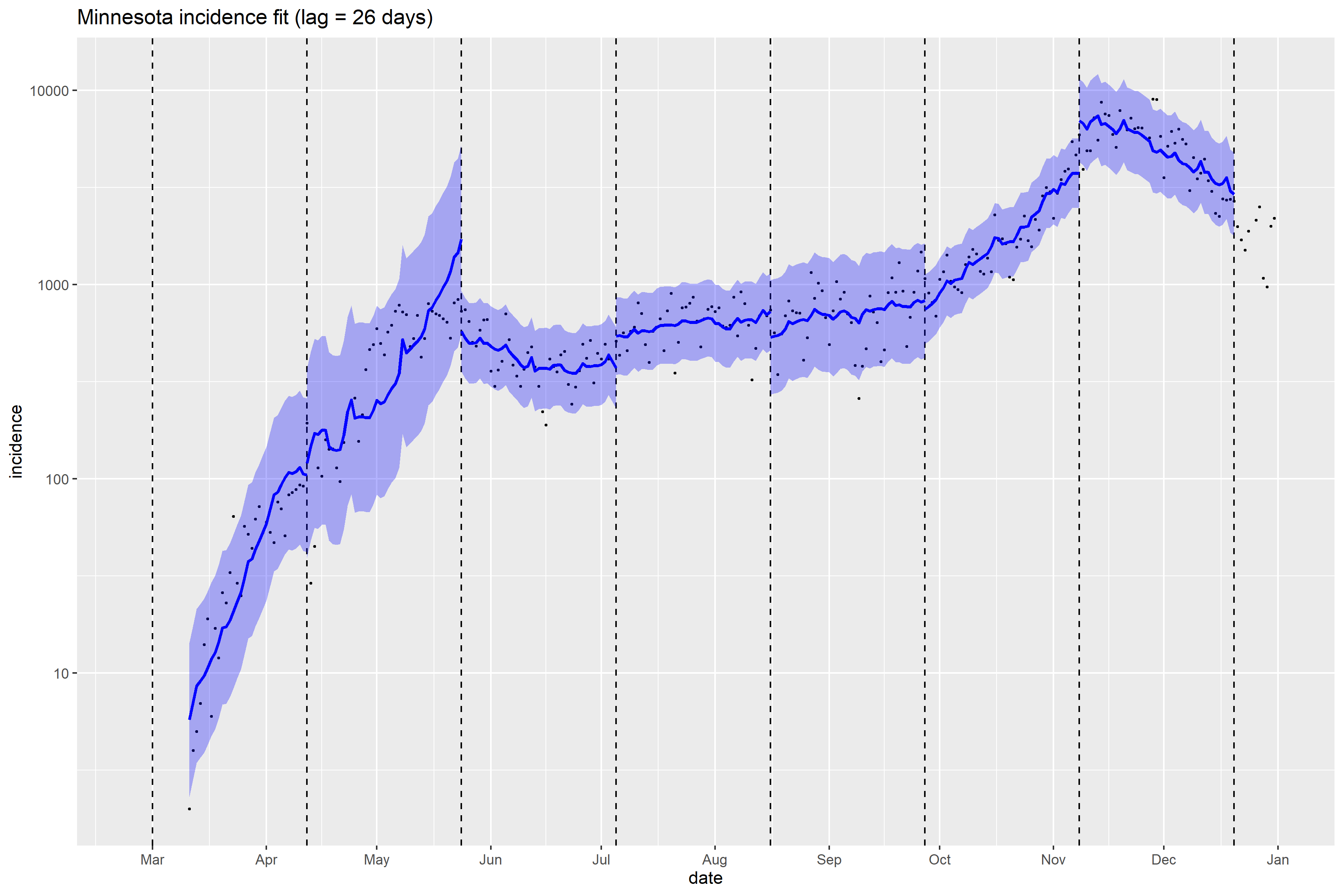

### Mississippi_full_fit_plot.png

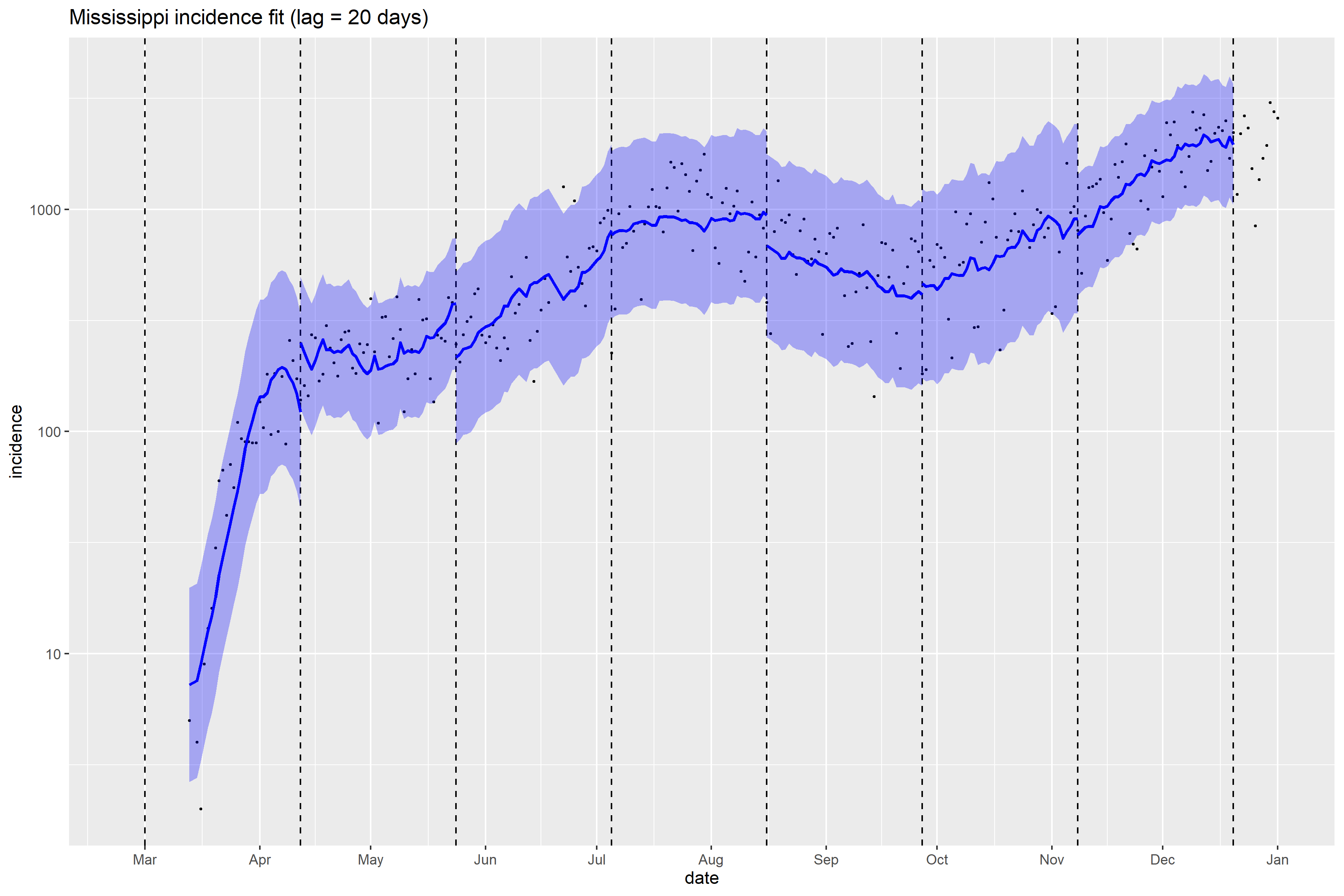

### Mississippi_Peff_plot.png

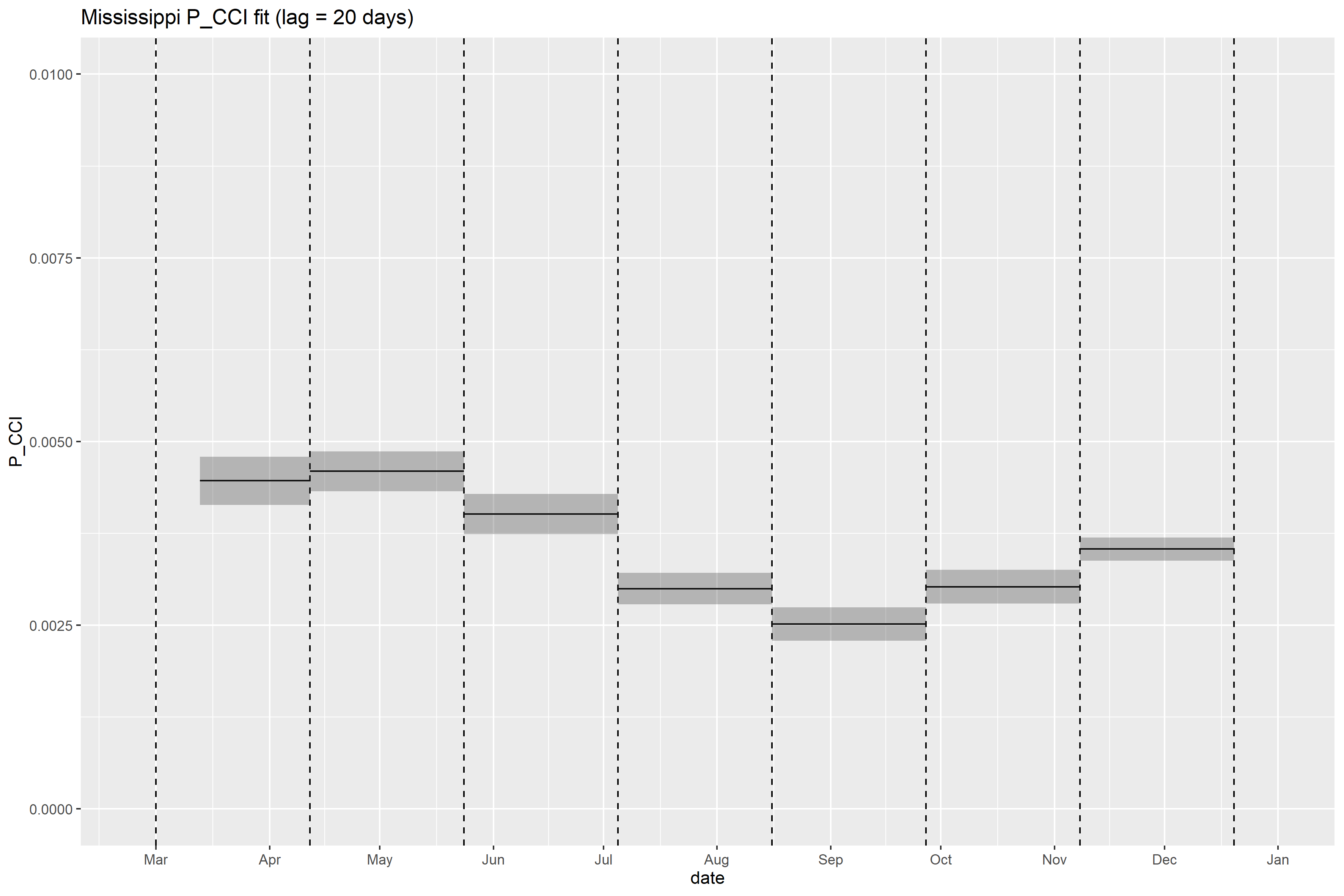

### Missouri_full_fit_plot.png

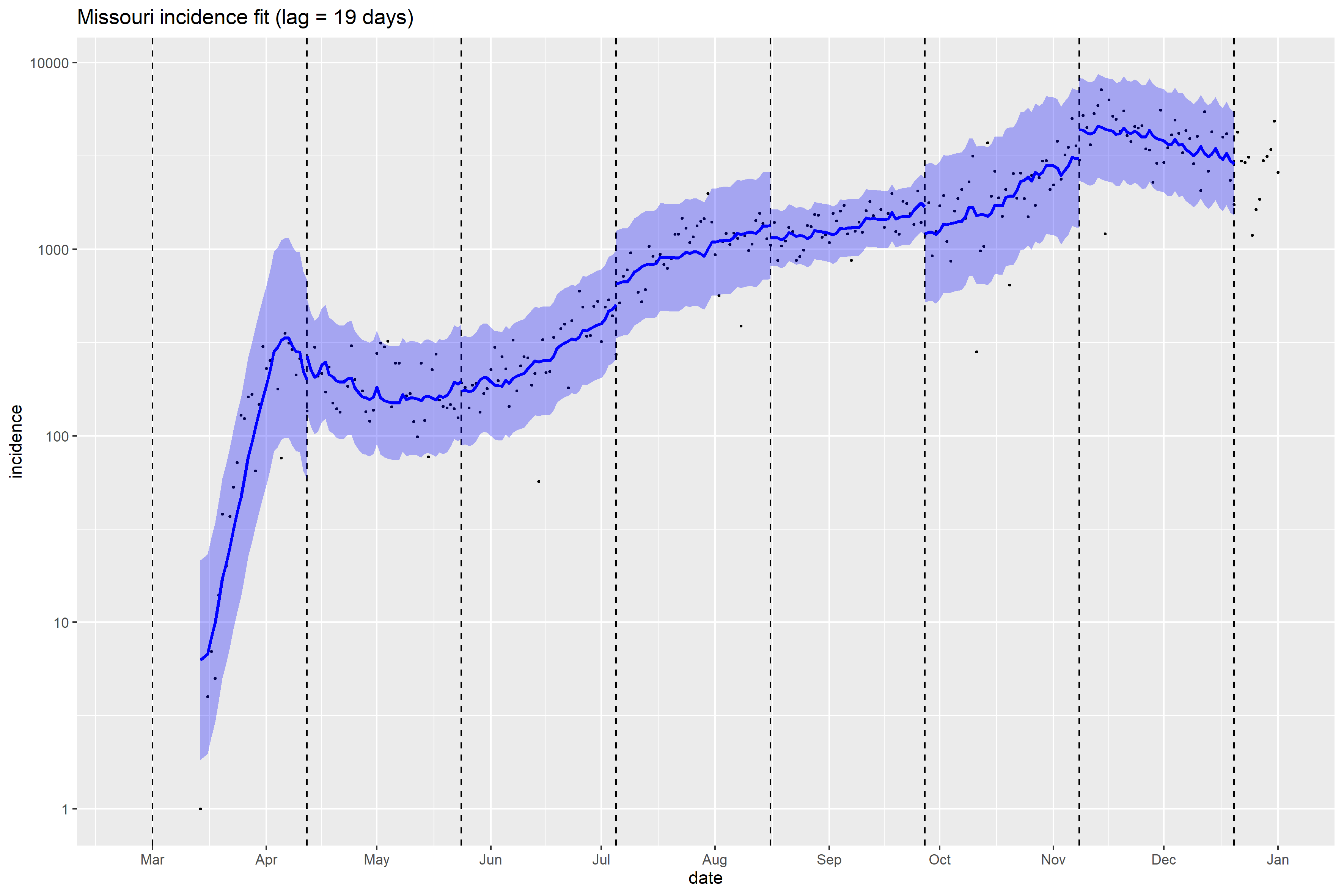

### Montana_full_fit_plot.png

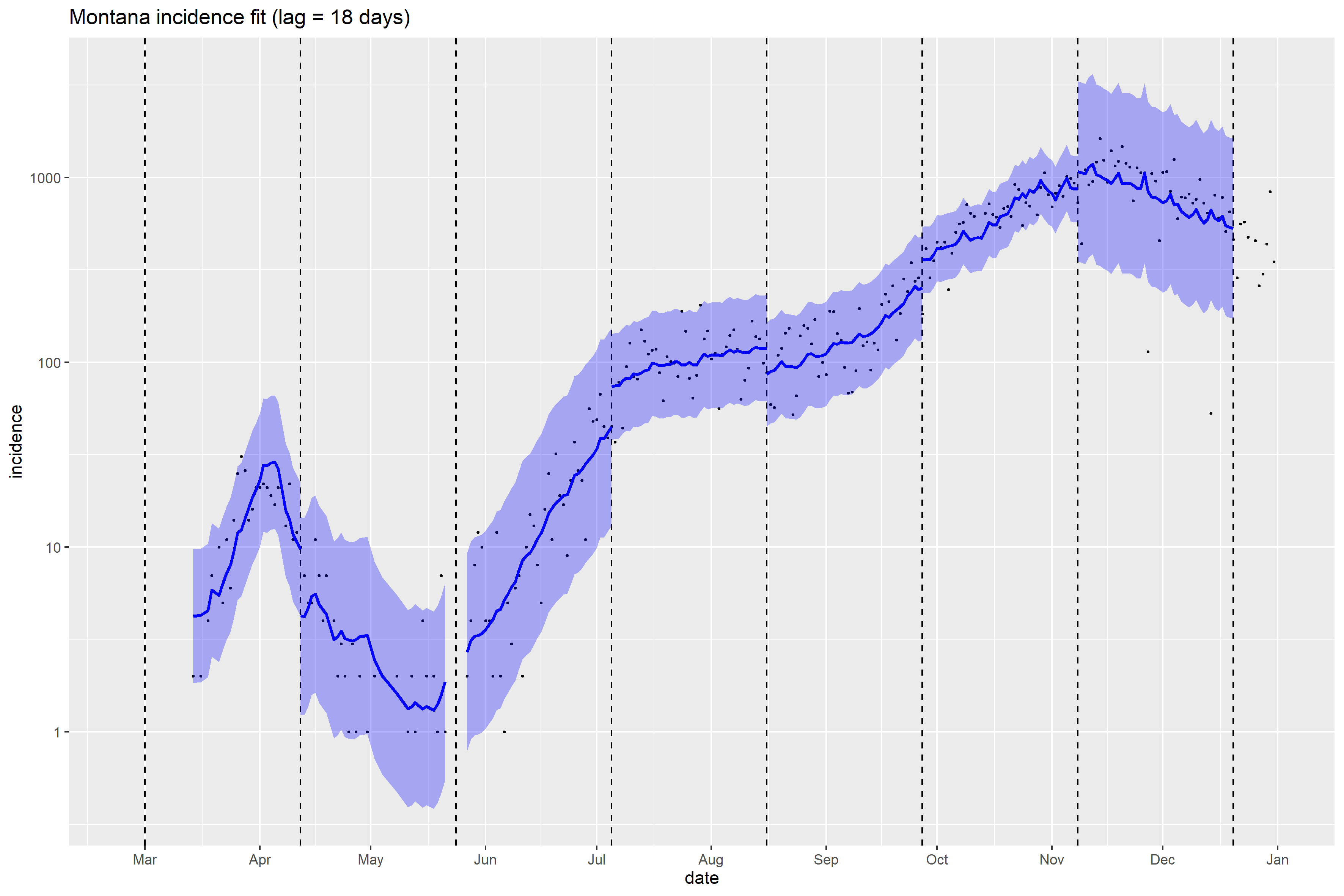

### Nebraska_full_fit_plot.png

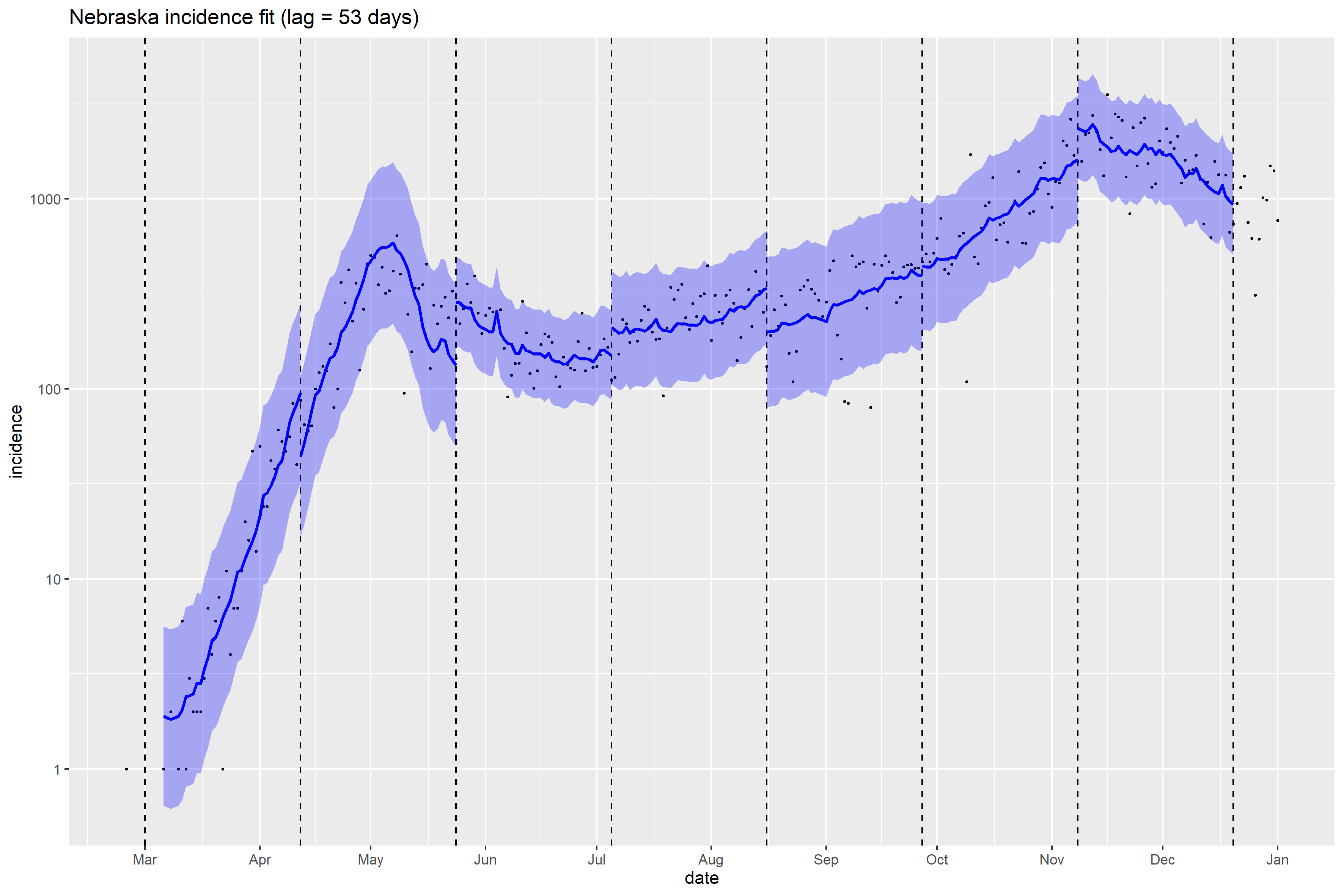

### New Hampshire_full_fit_plot.png

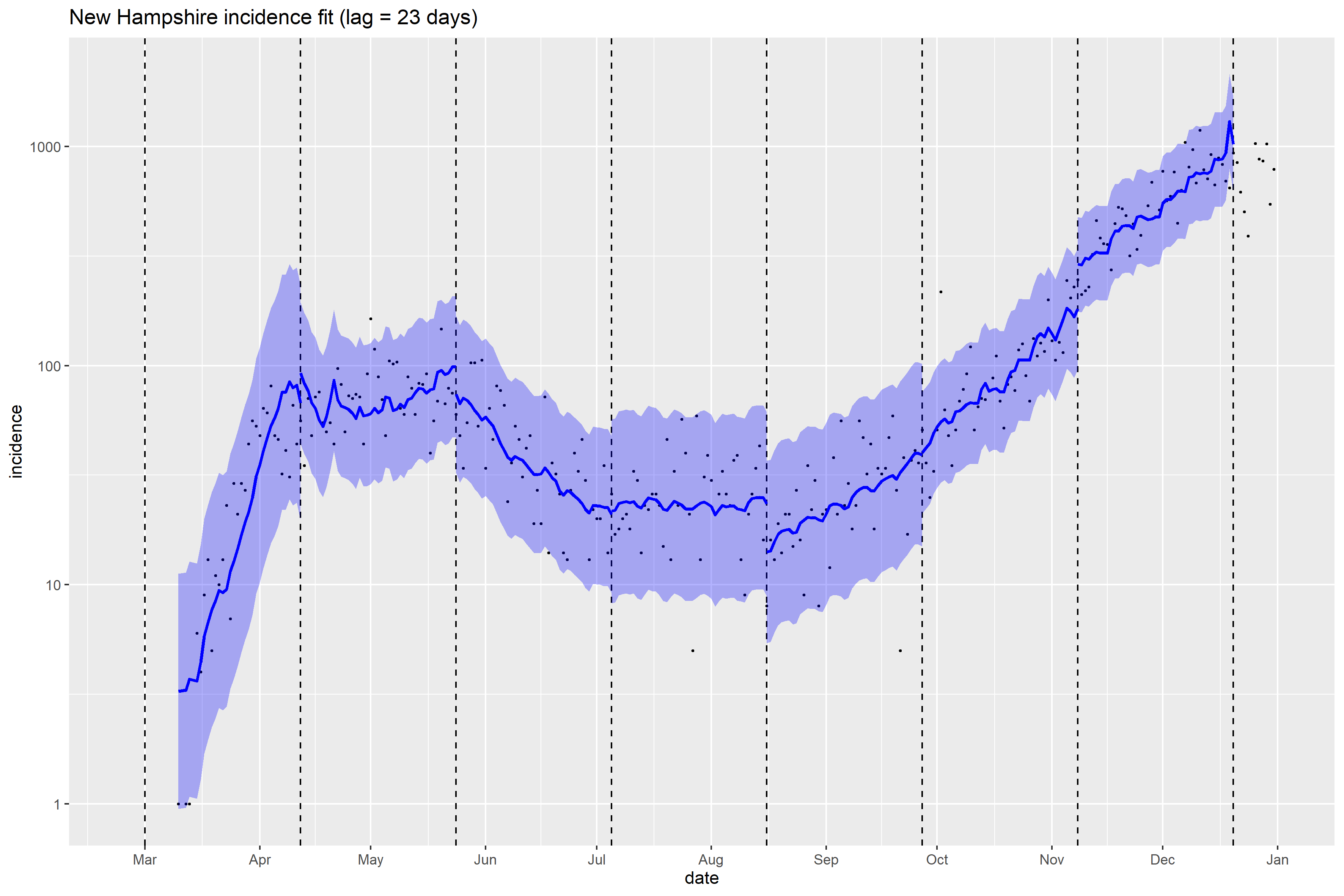

### New Hampshire_Peff_plot.png

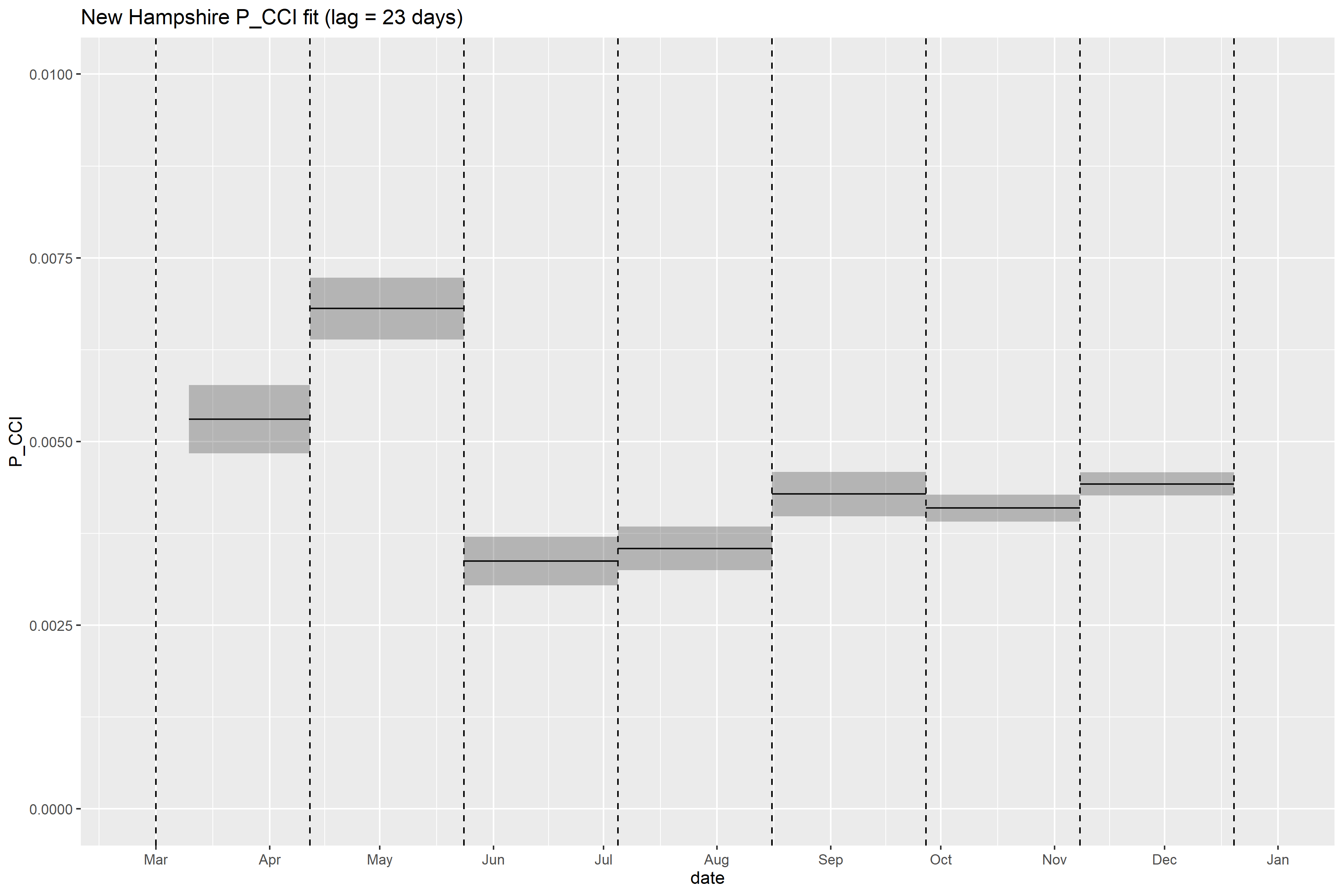

### New Jersey_full_fit_plot.png

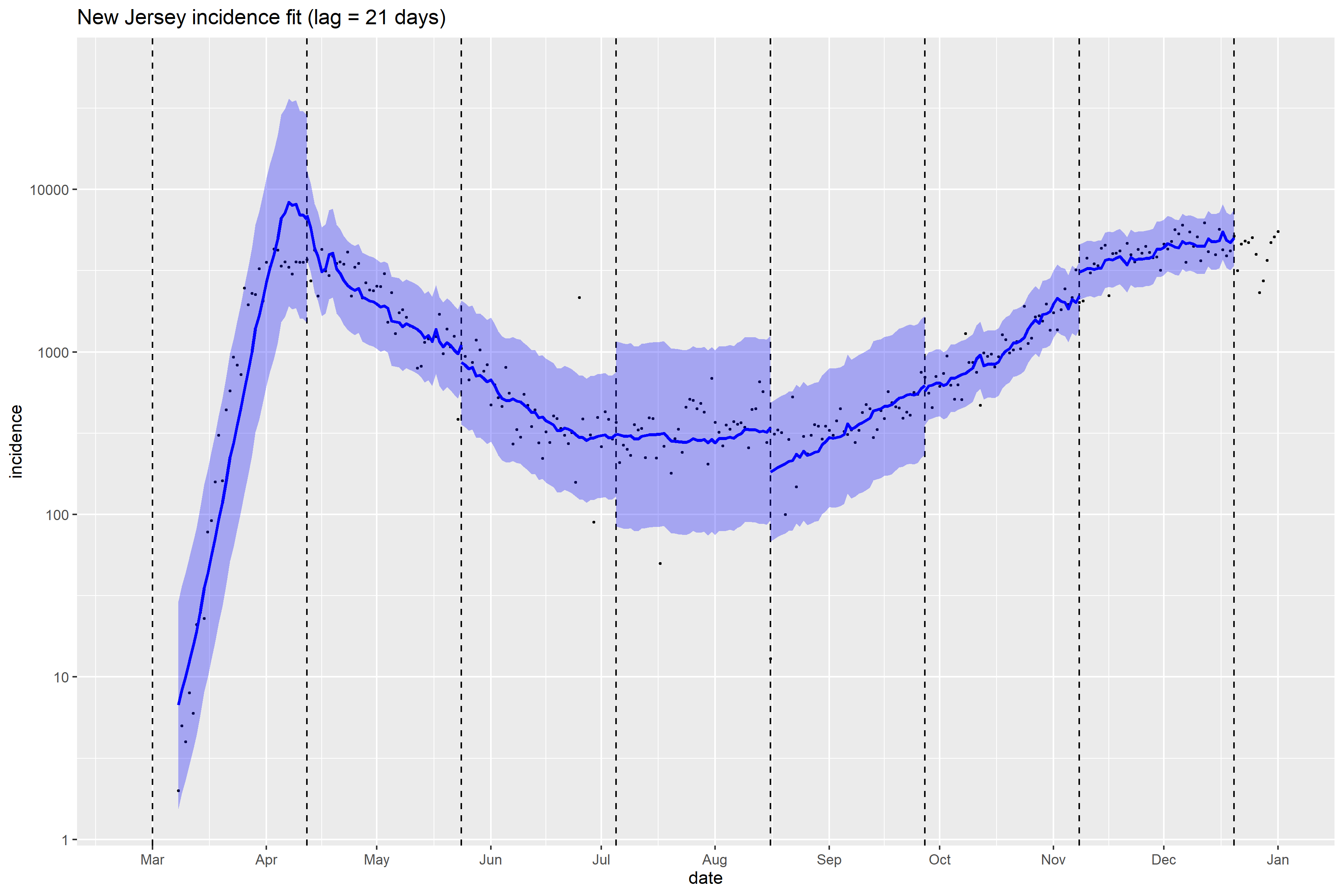

### New Mexico_full_fit_plot.png

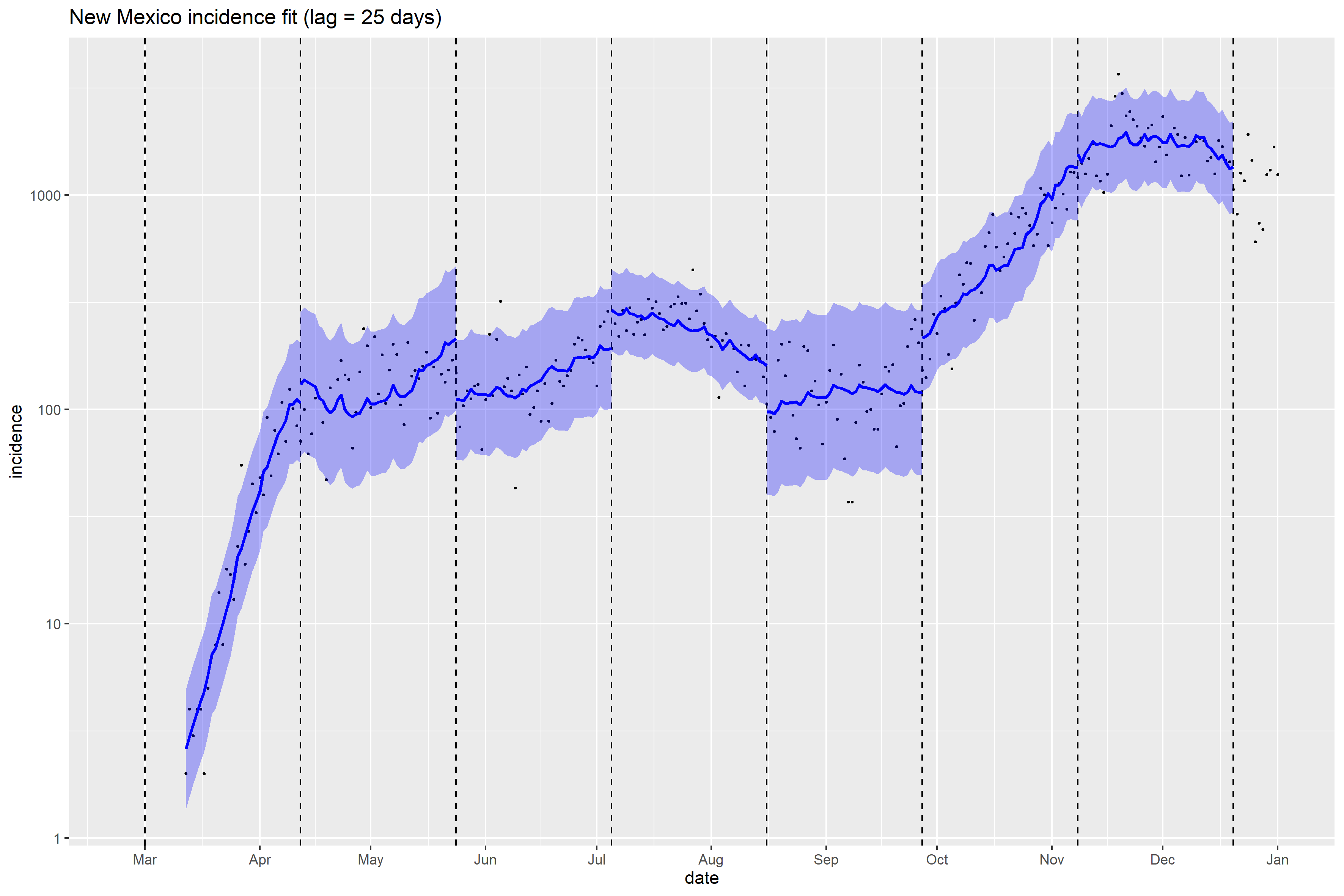

### New York_full_fit_plot.png

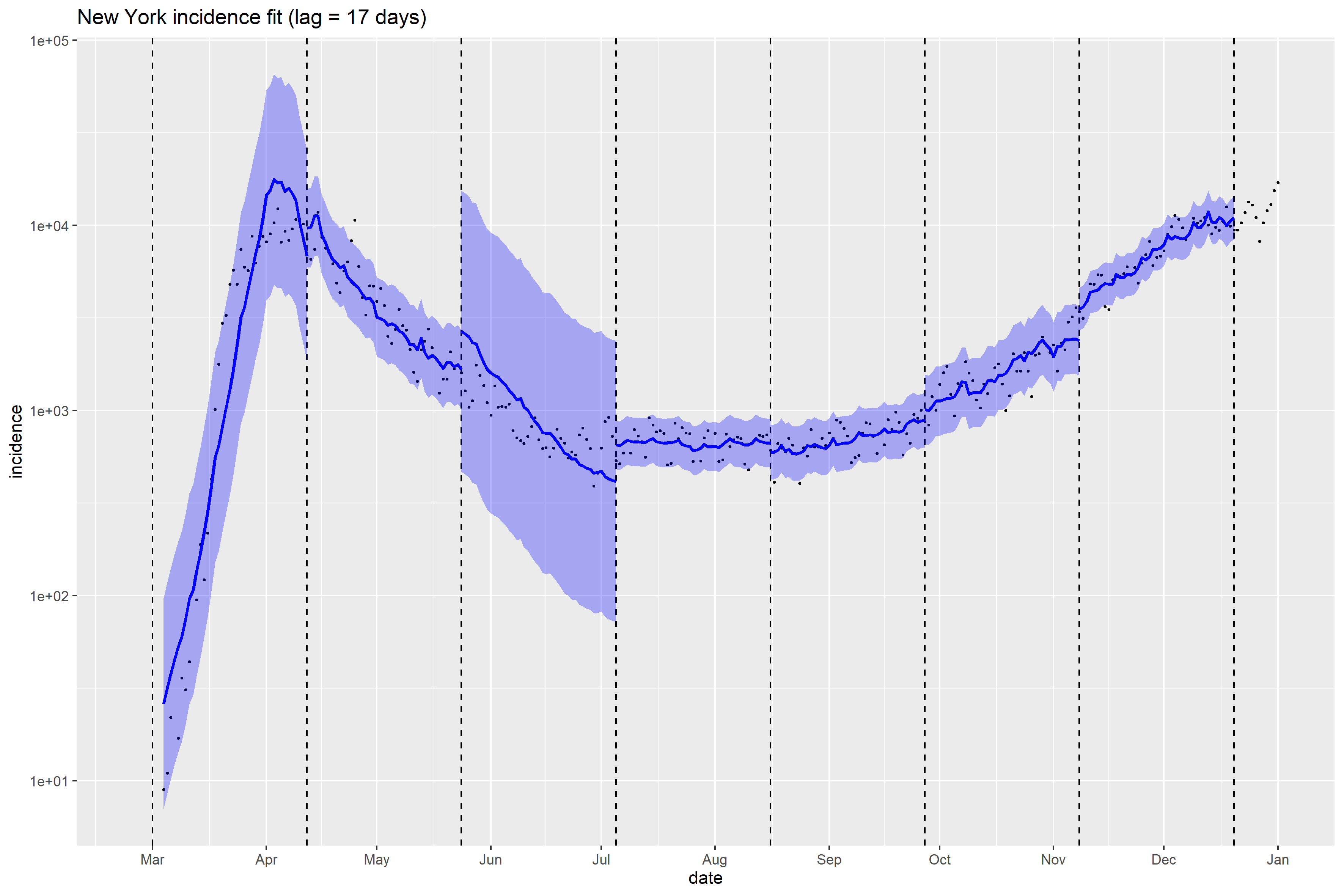

### North Carolina_full_fit_plot.png

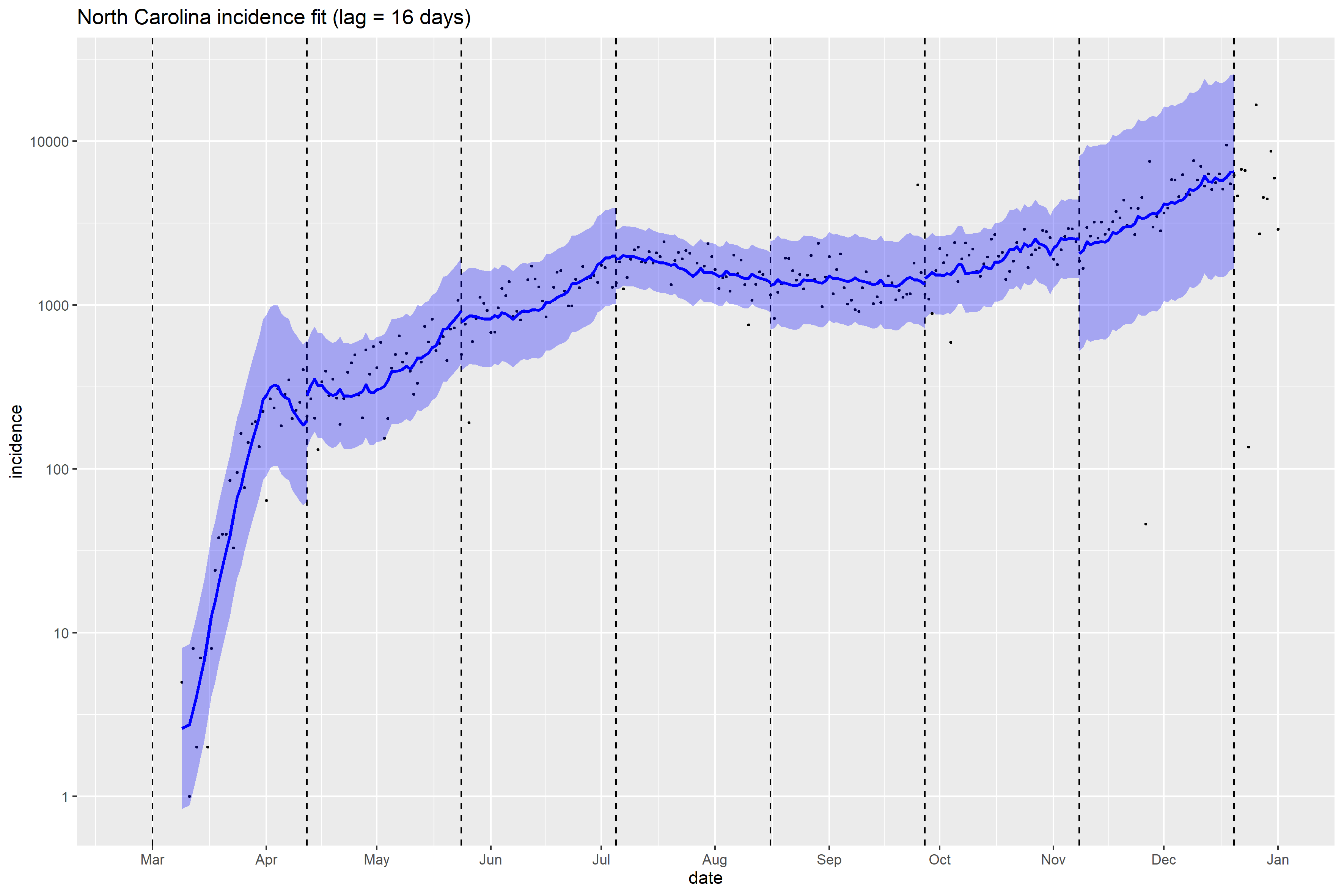

### North Carolina_Peff_plot.png

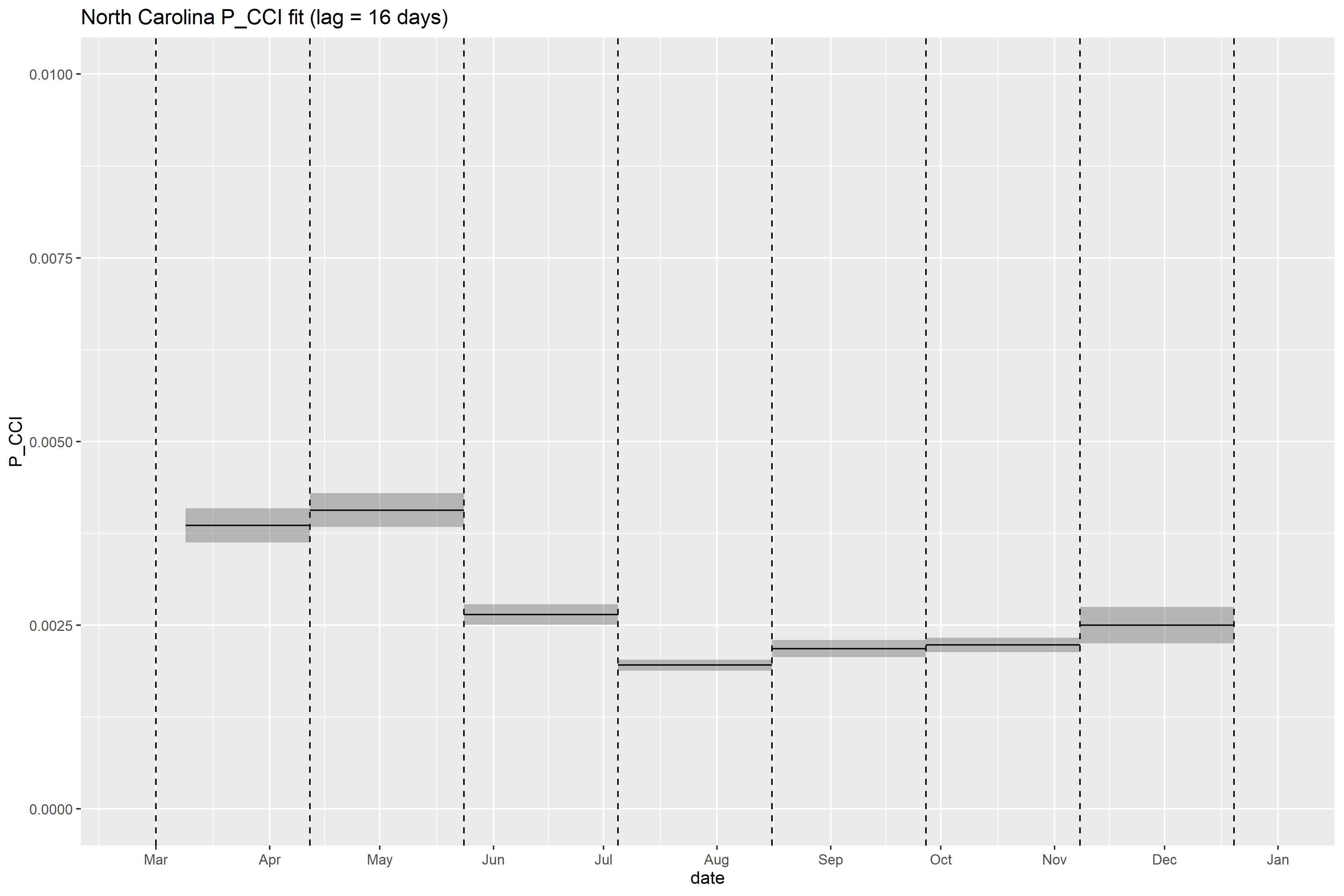

### North Dakota_full_fit_plot.png

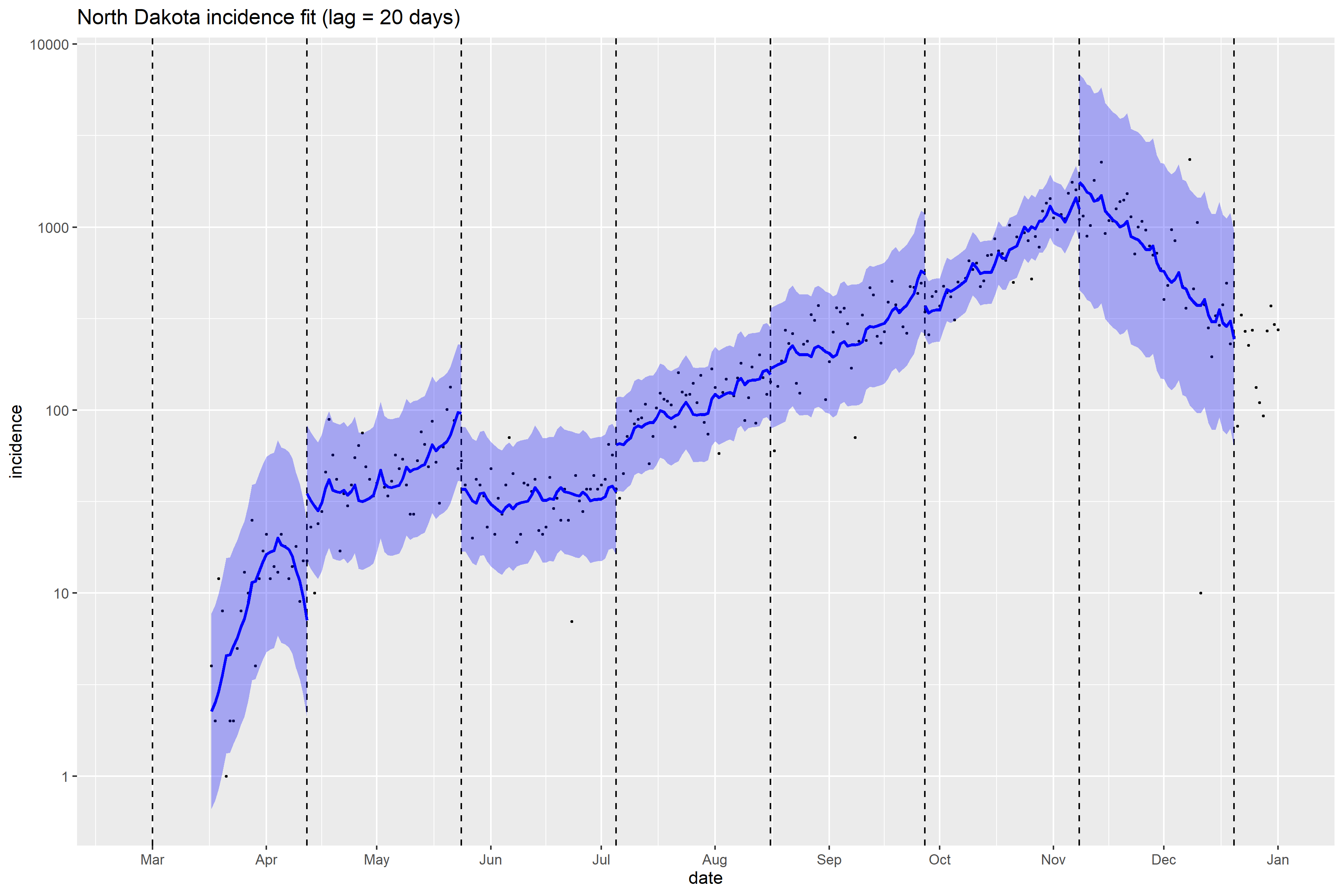

### North Dakota_Peff_plot.png

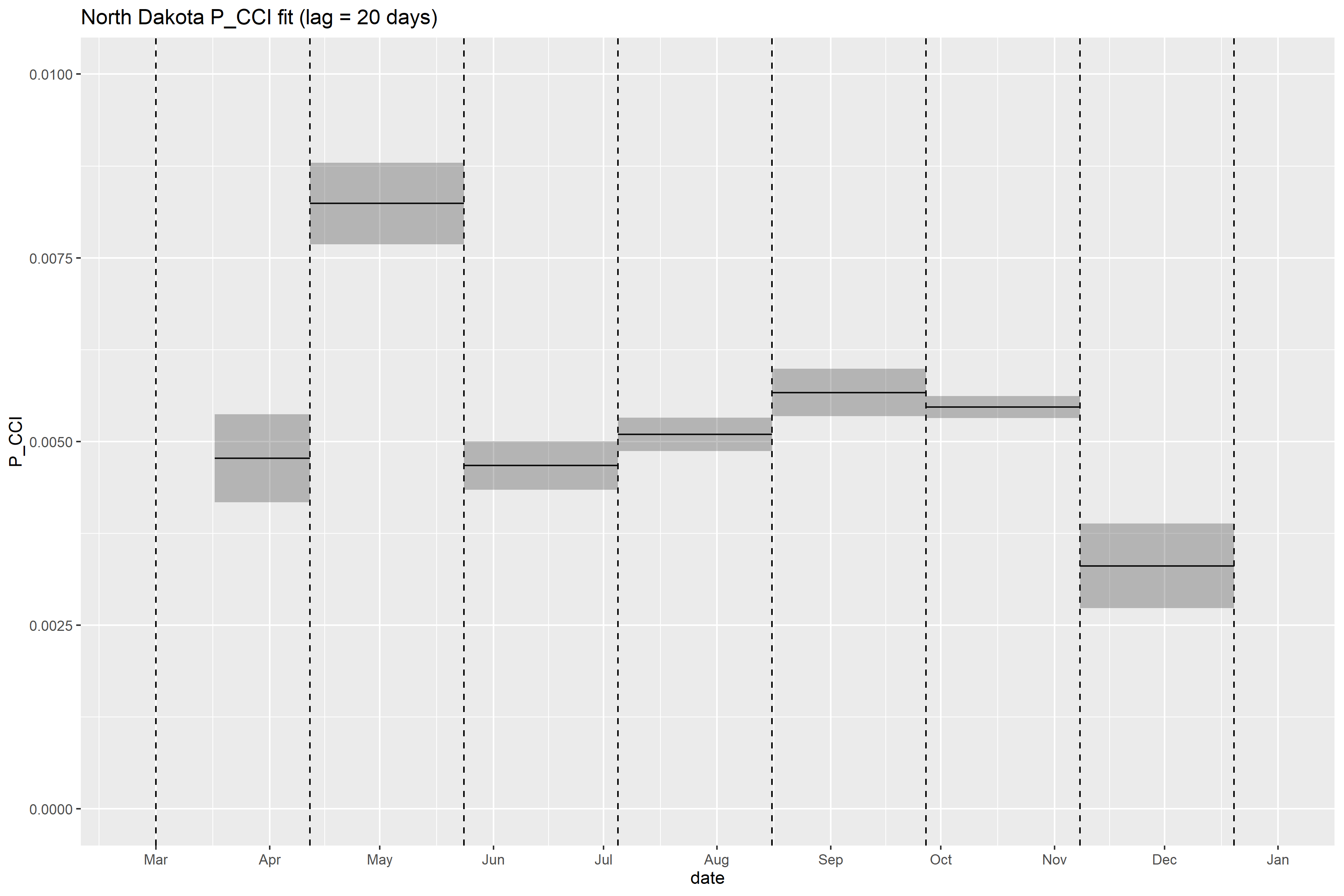

### Oklahoma_full_fit_plot.png

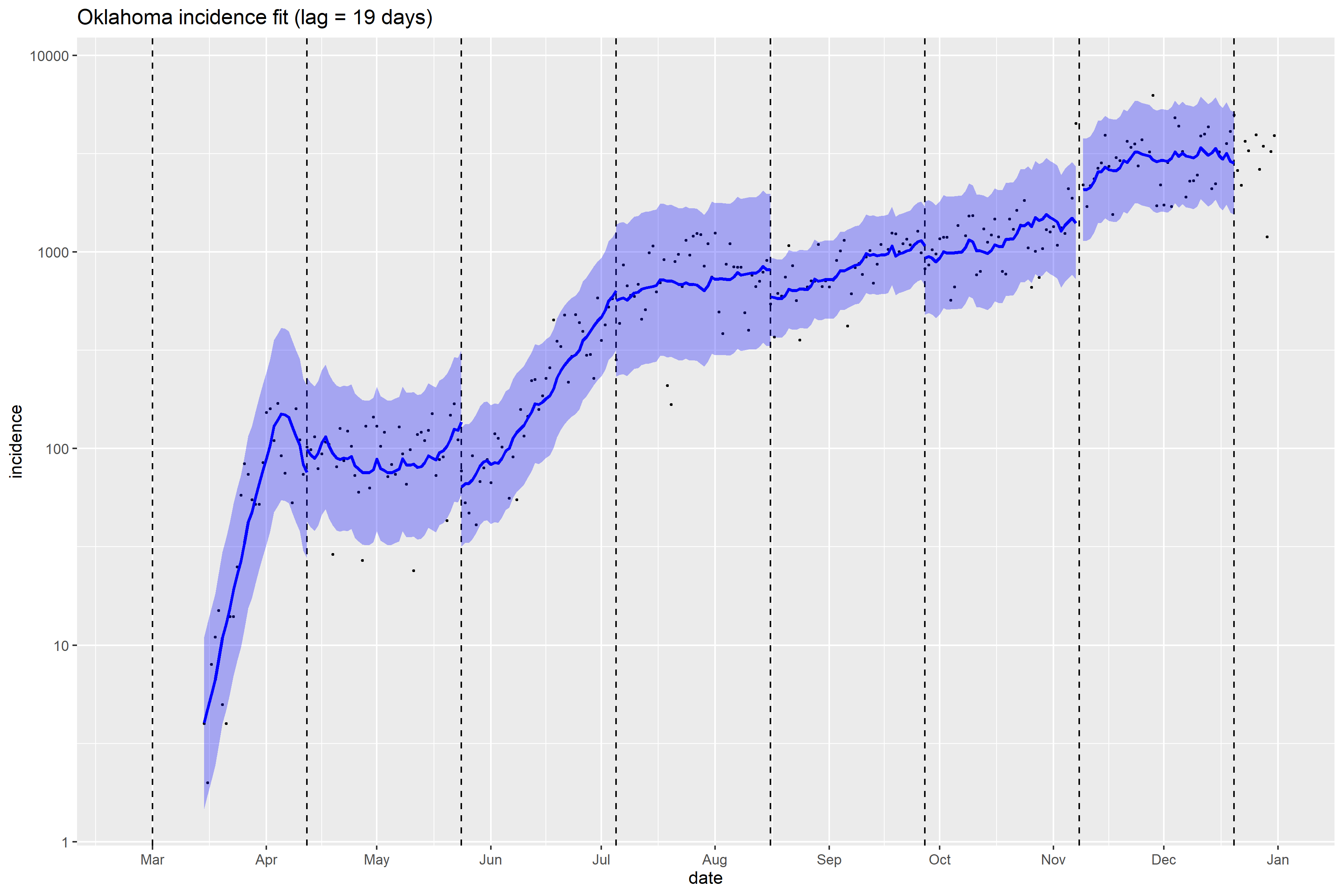

### Pennsylvania_full_fit_plot.png

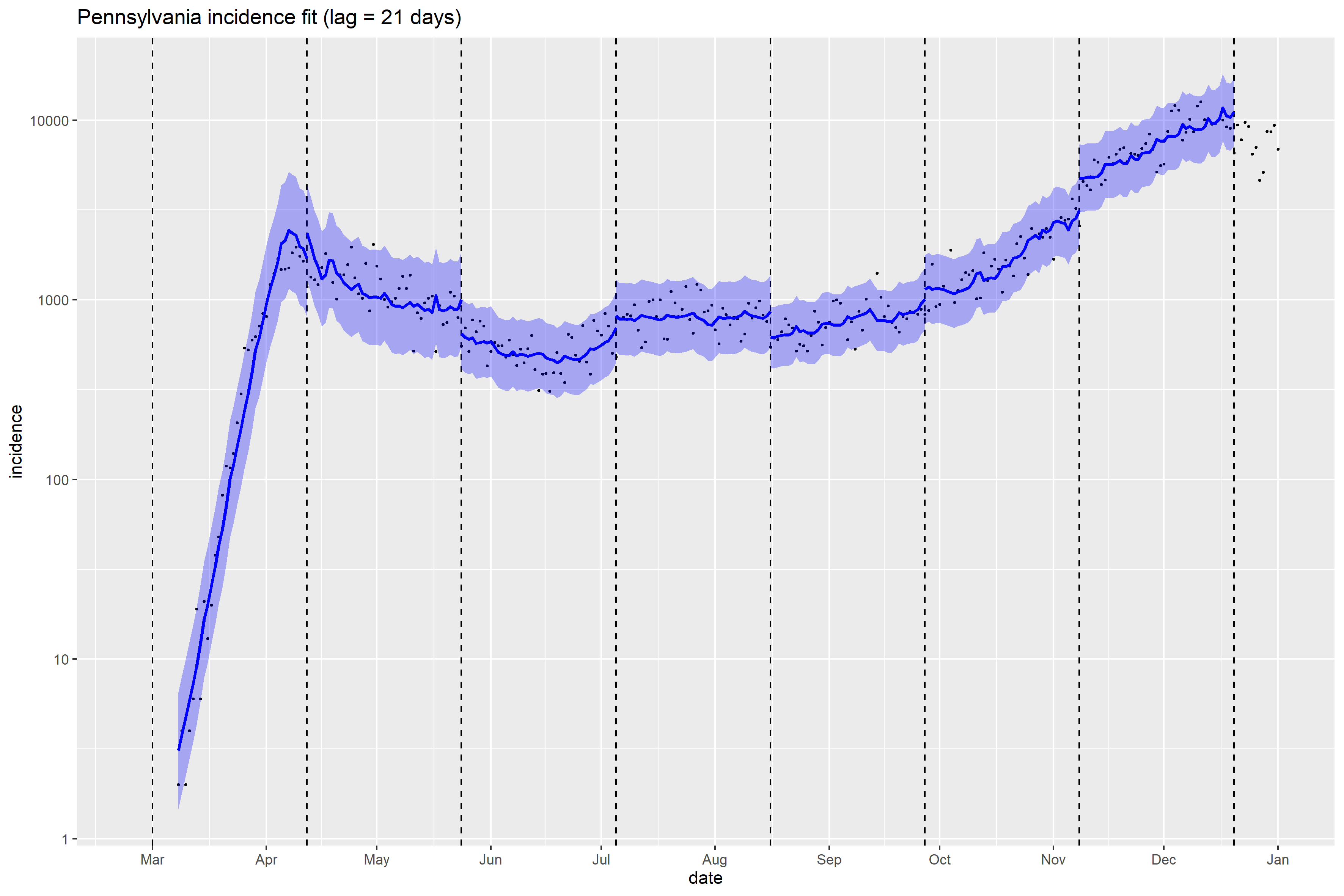

### Pennsylvania_Peff_plot.png

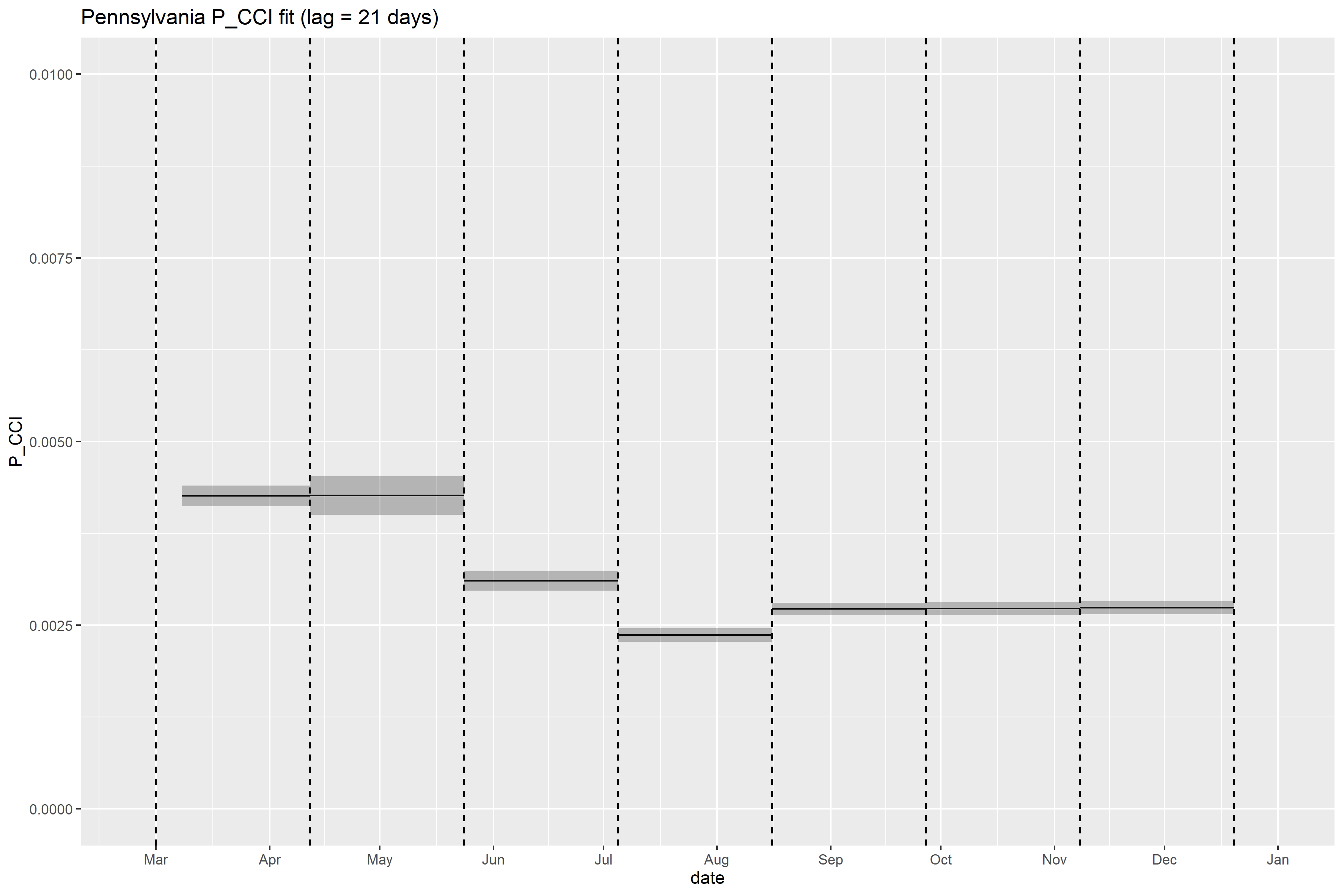

### Rhode Island_full_fit_plot.png

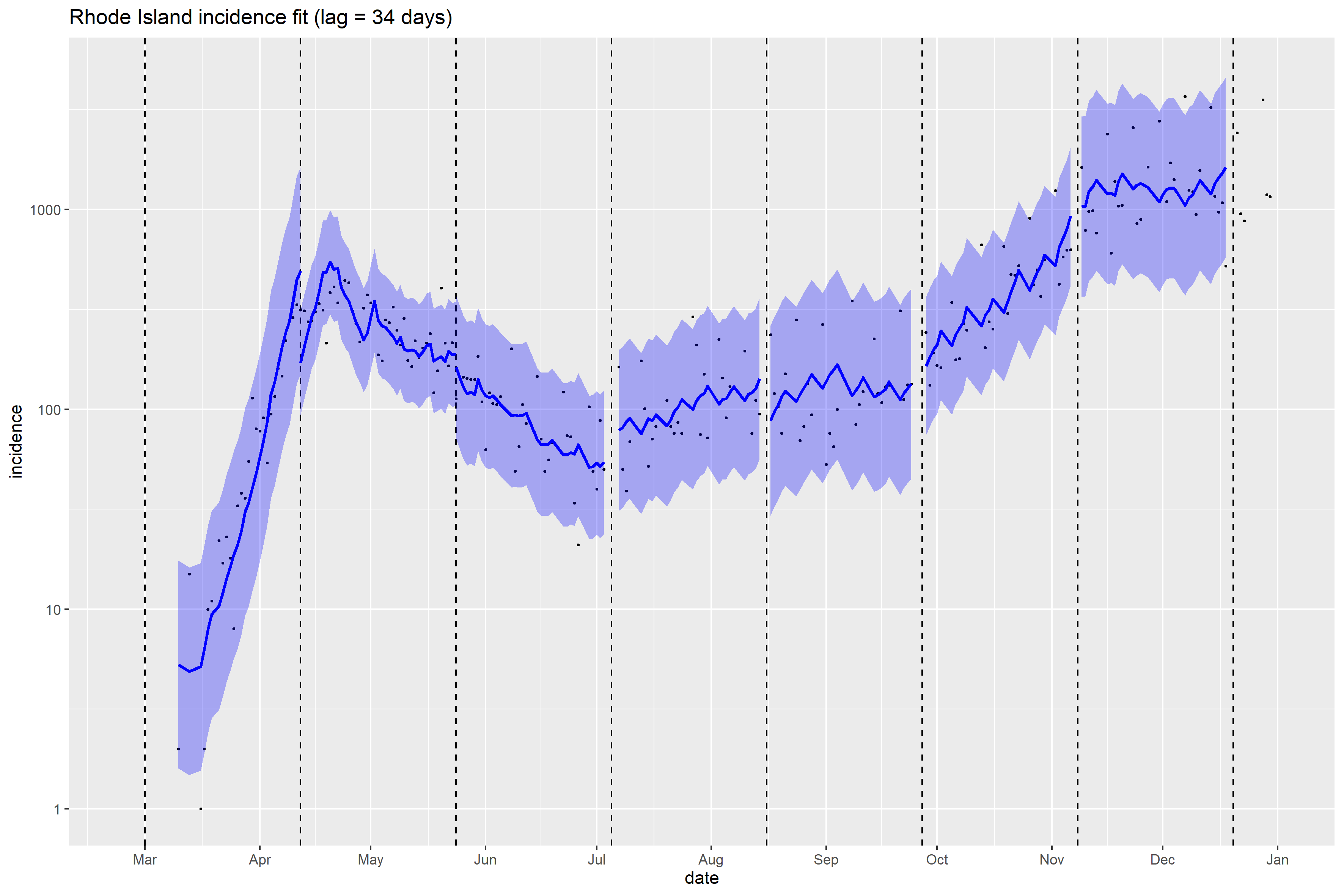

### Rhode Island_Peff_plot.png

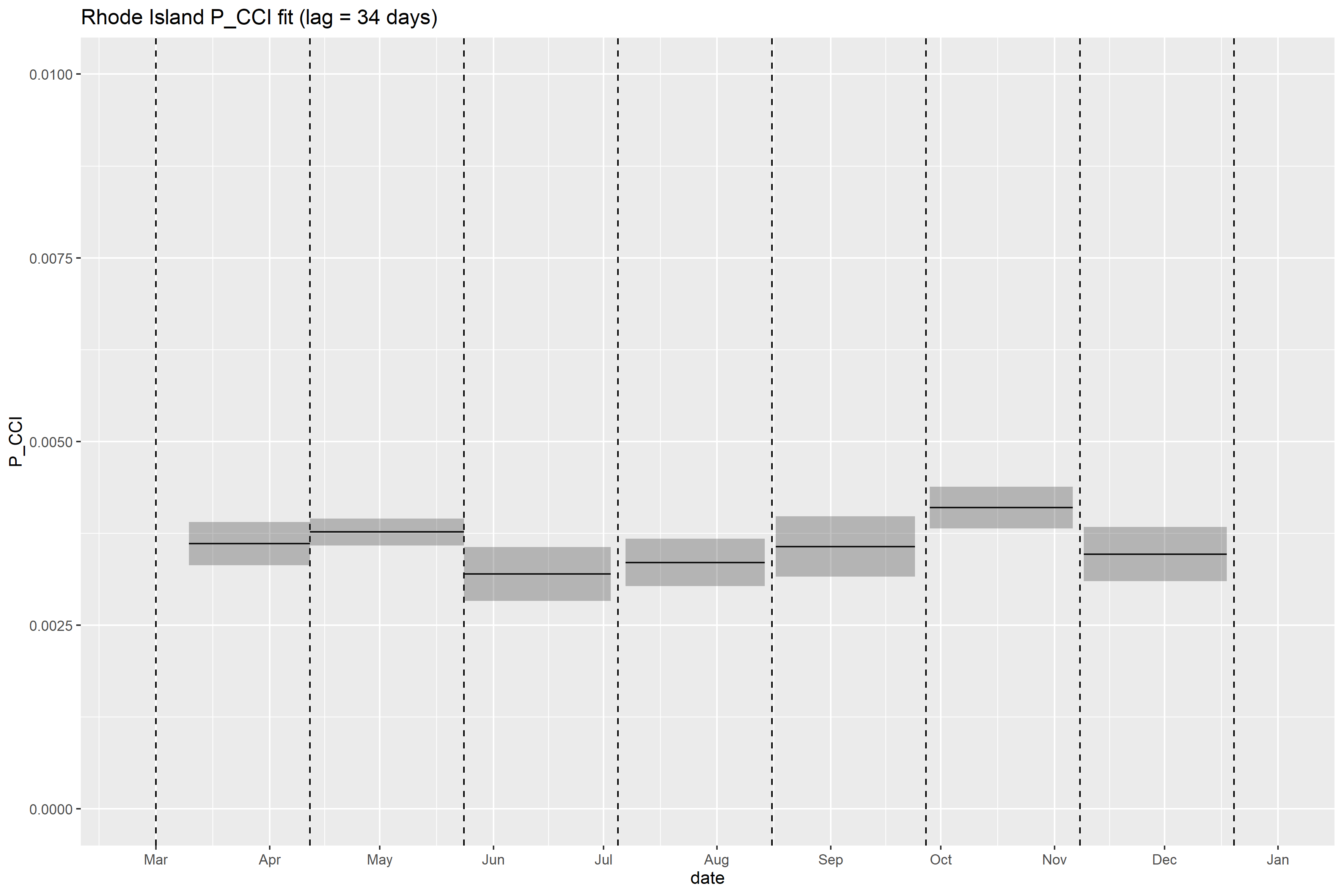

### South Carolina_full_fit_plot.png

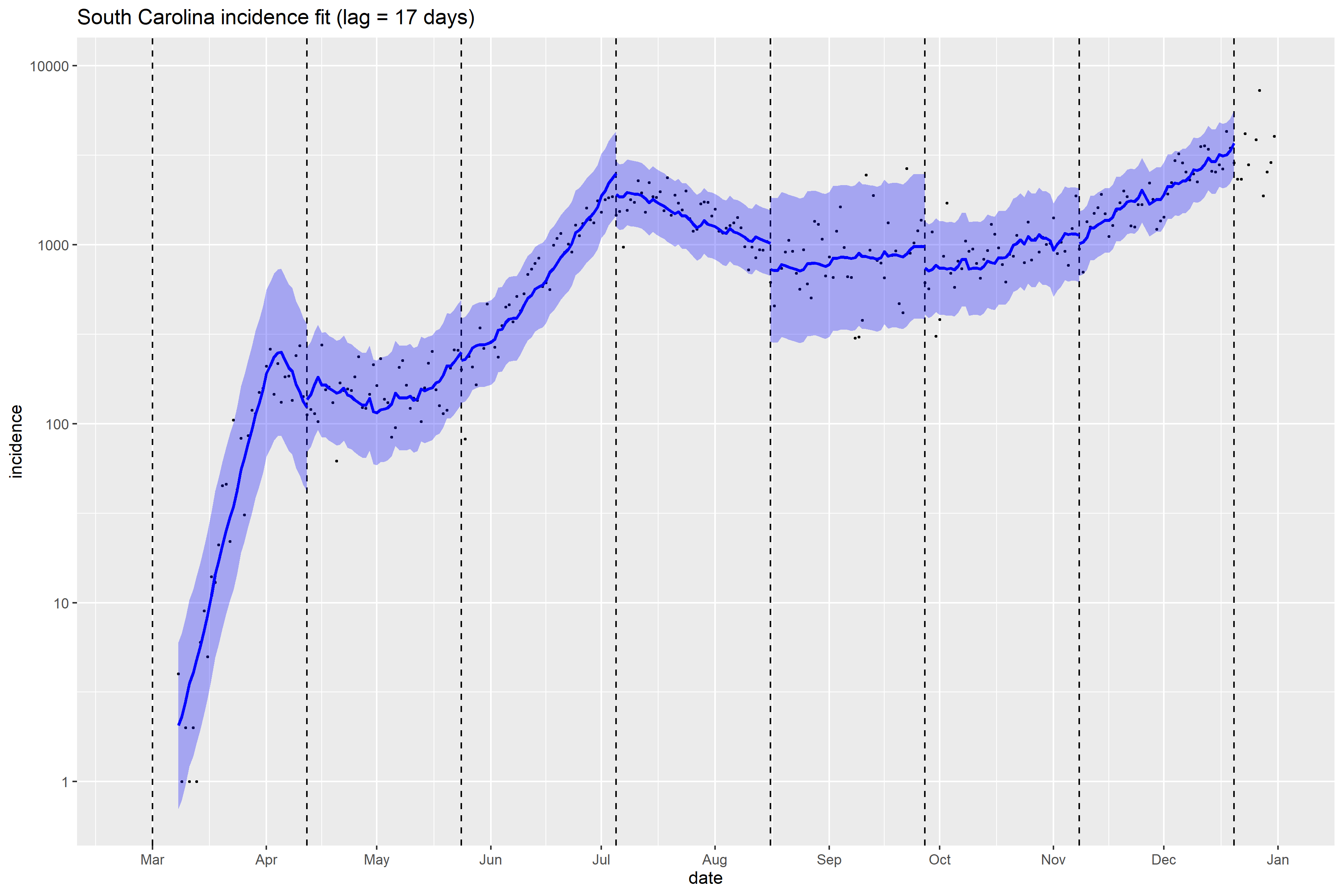

### South Carolina_Peff_plot.png

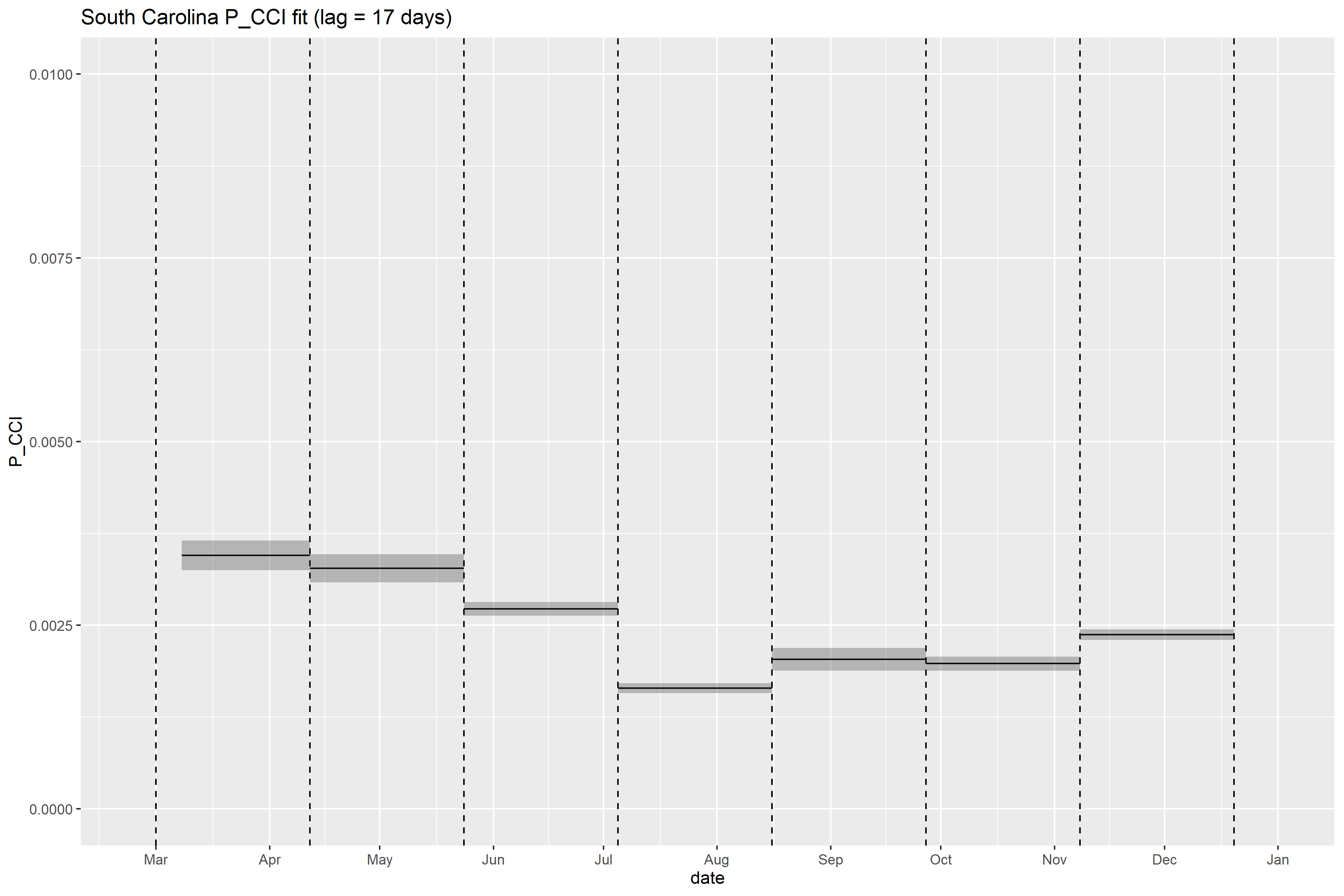

### South Dakota_full_fit_plot.png

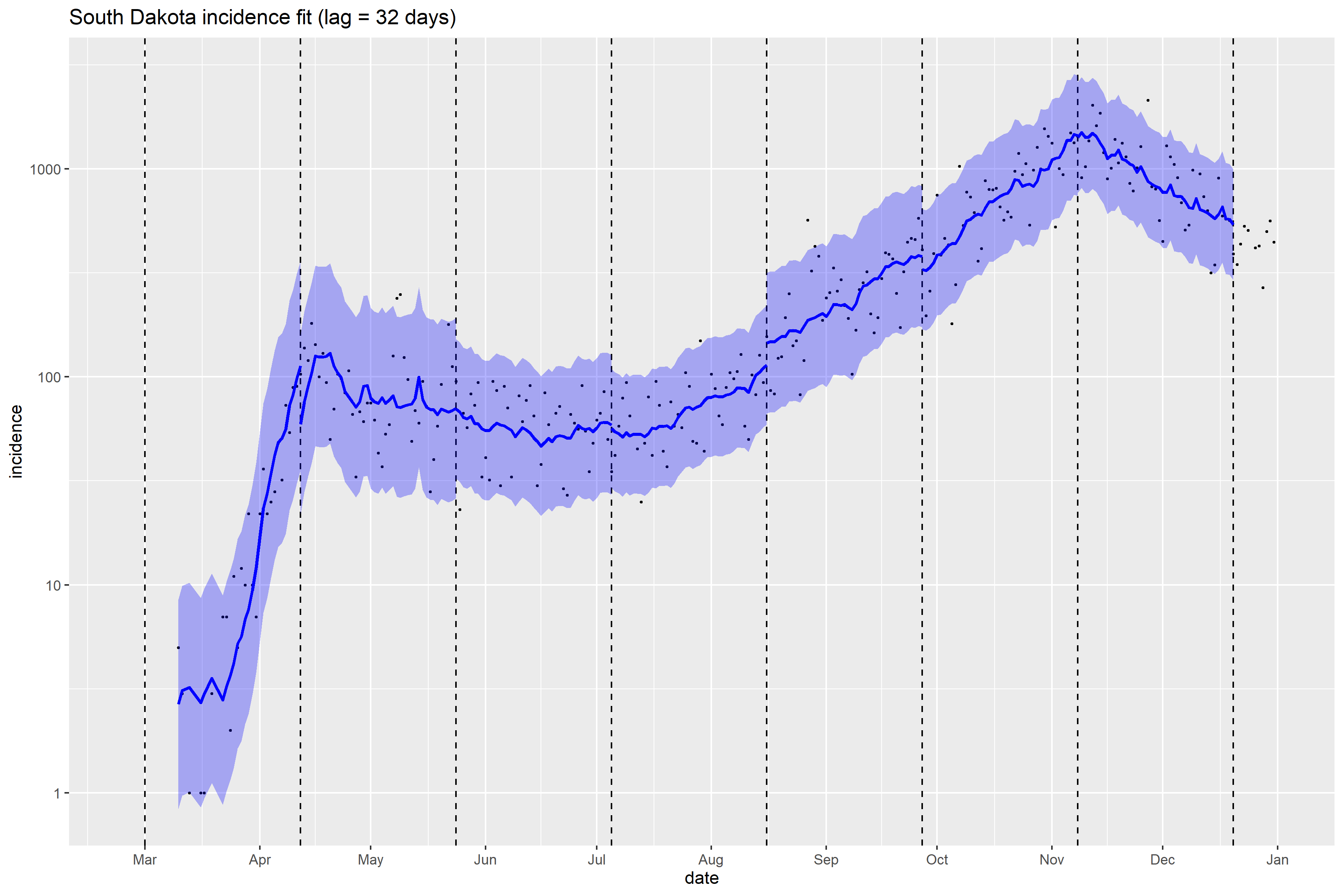

### South Dakota_Peff_plot.png

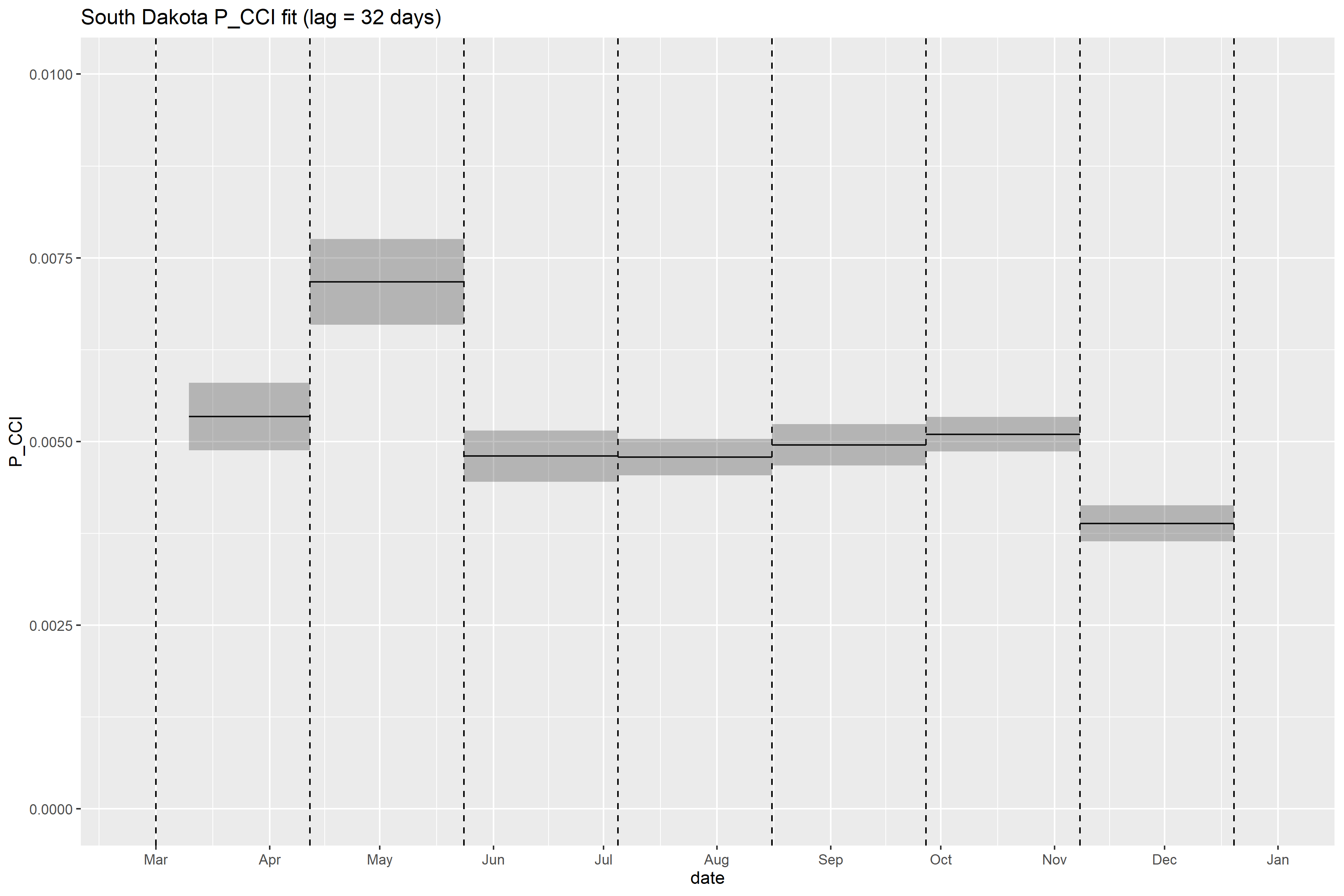

### Tennessee_full_fit_plot.png

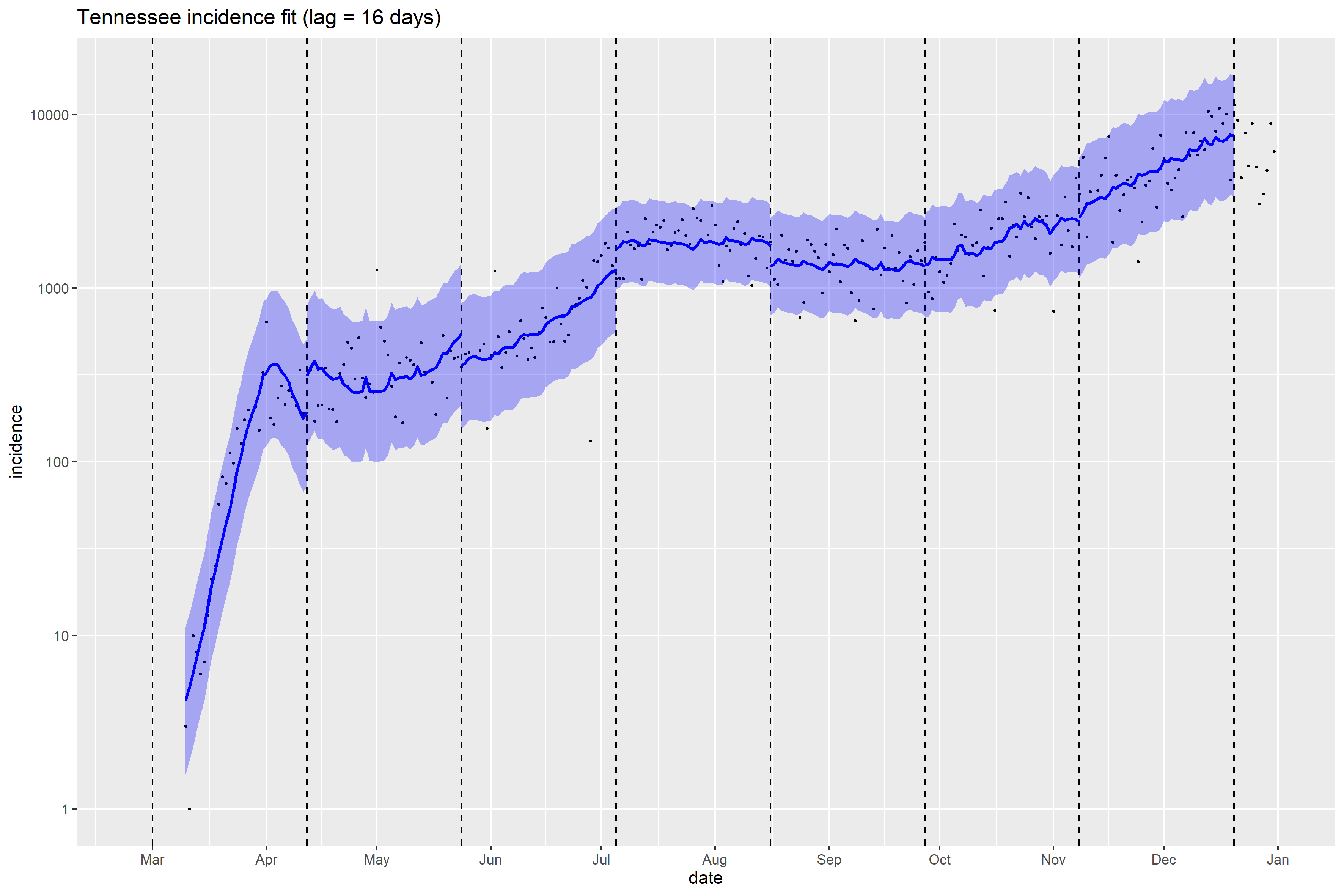

### Vermont_full_fit_plot.png

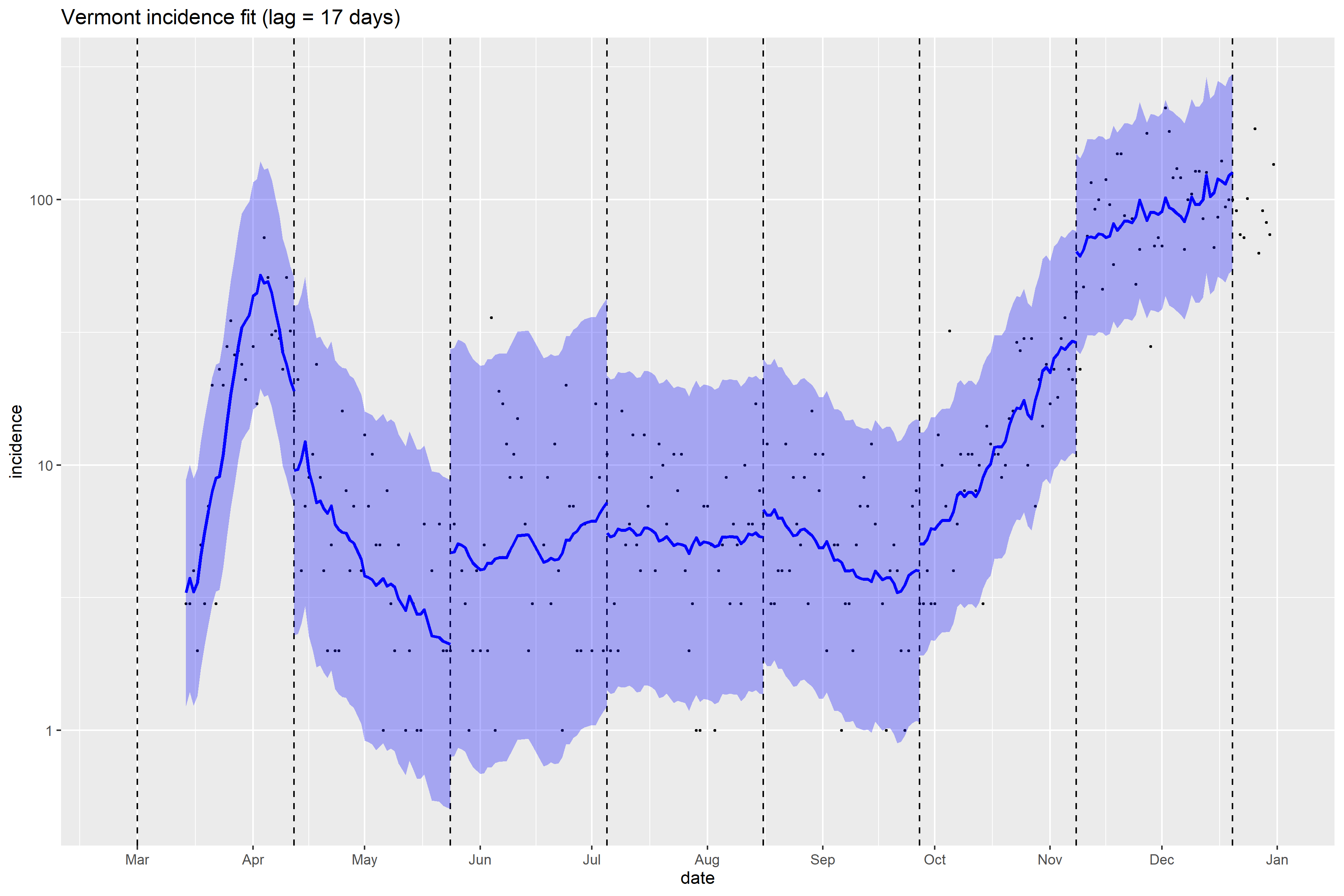

### Virginia_full_fit_plot.png

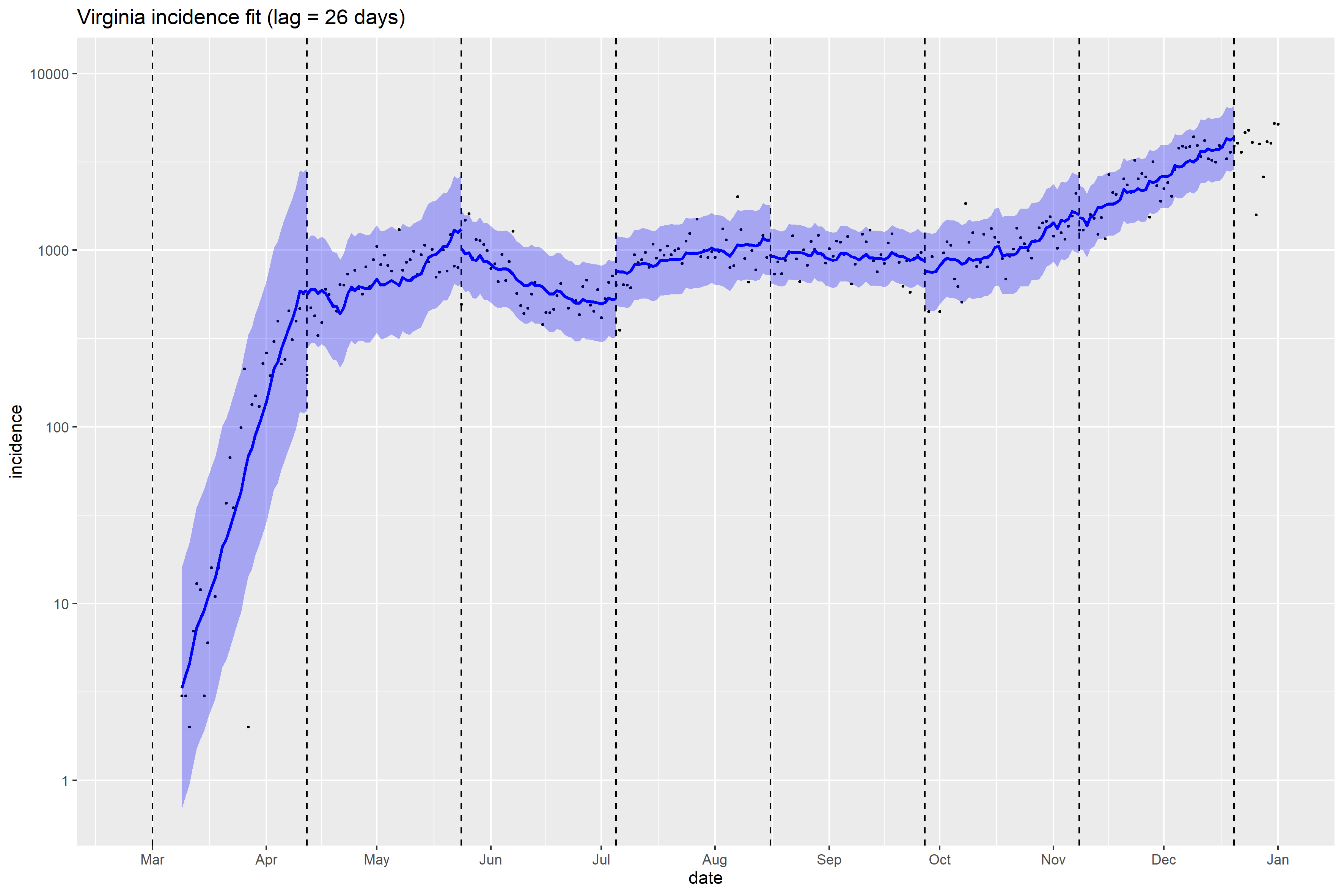

### Washington_full_fit_plot.png

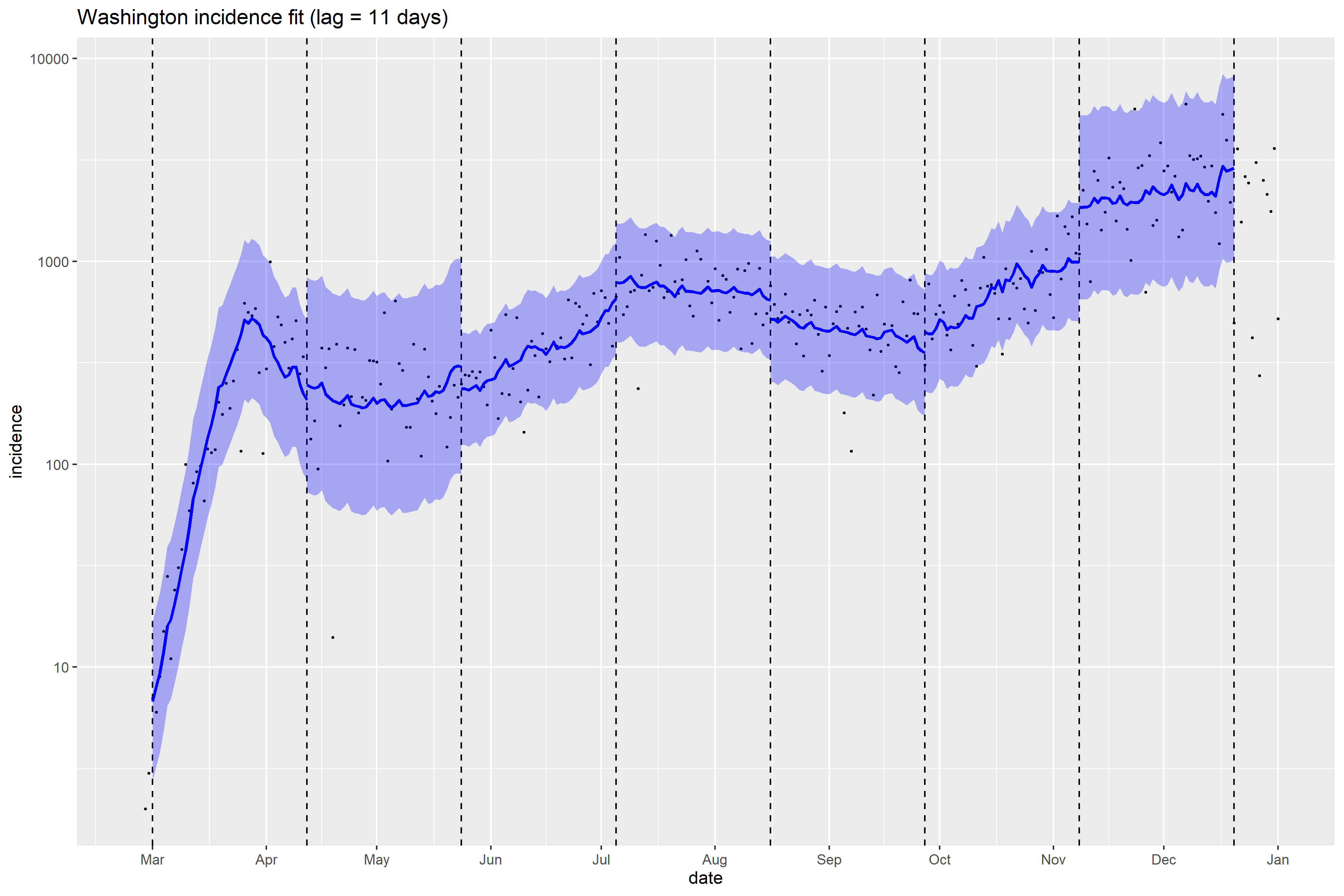

### Washington_Peff_plot.png

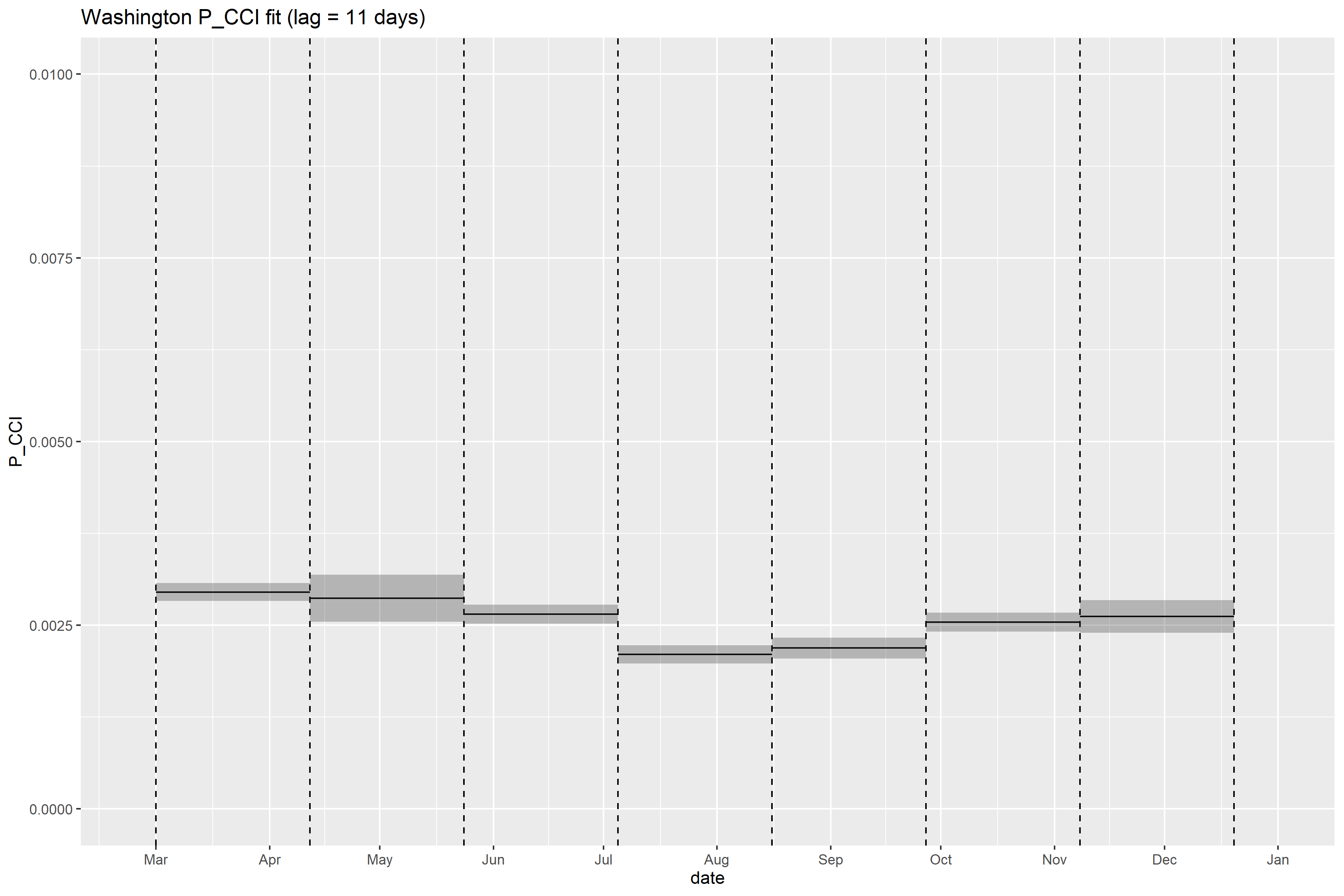

### West Virginia_full_fit_plot.png

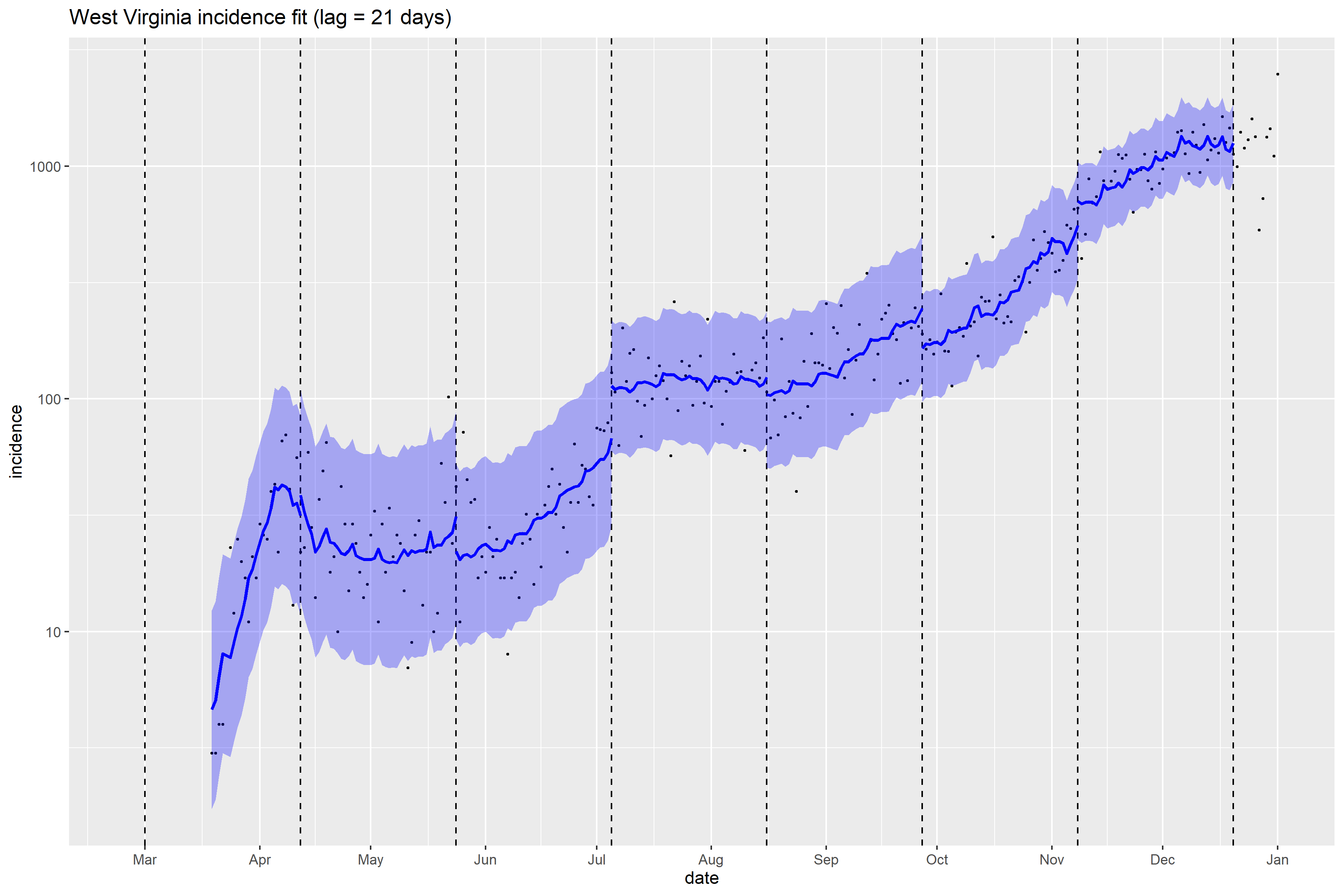

### West Virginia_Peff_plot.png

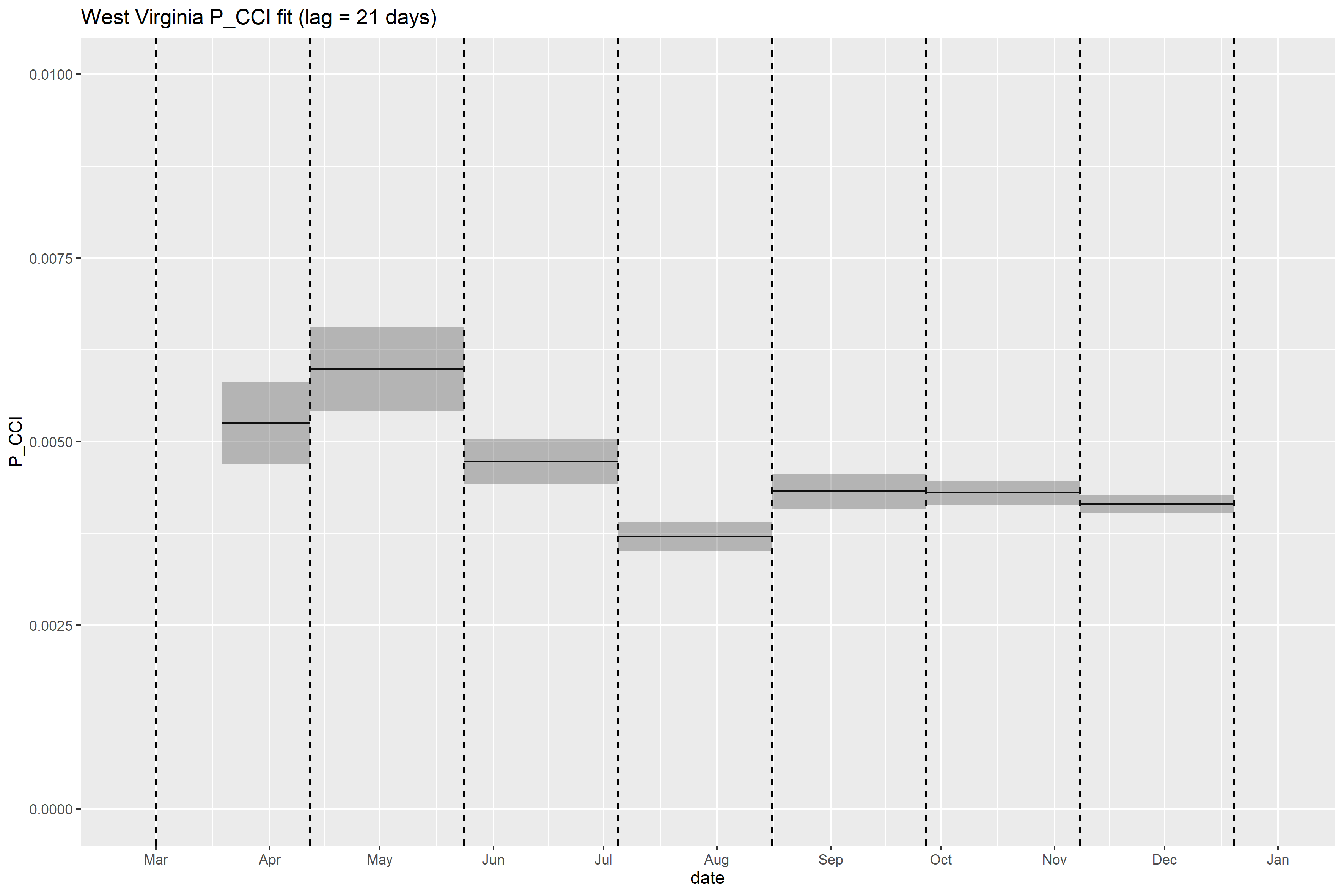

### Wisconsin_full_fit_plot.png

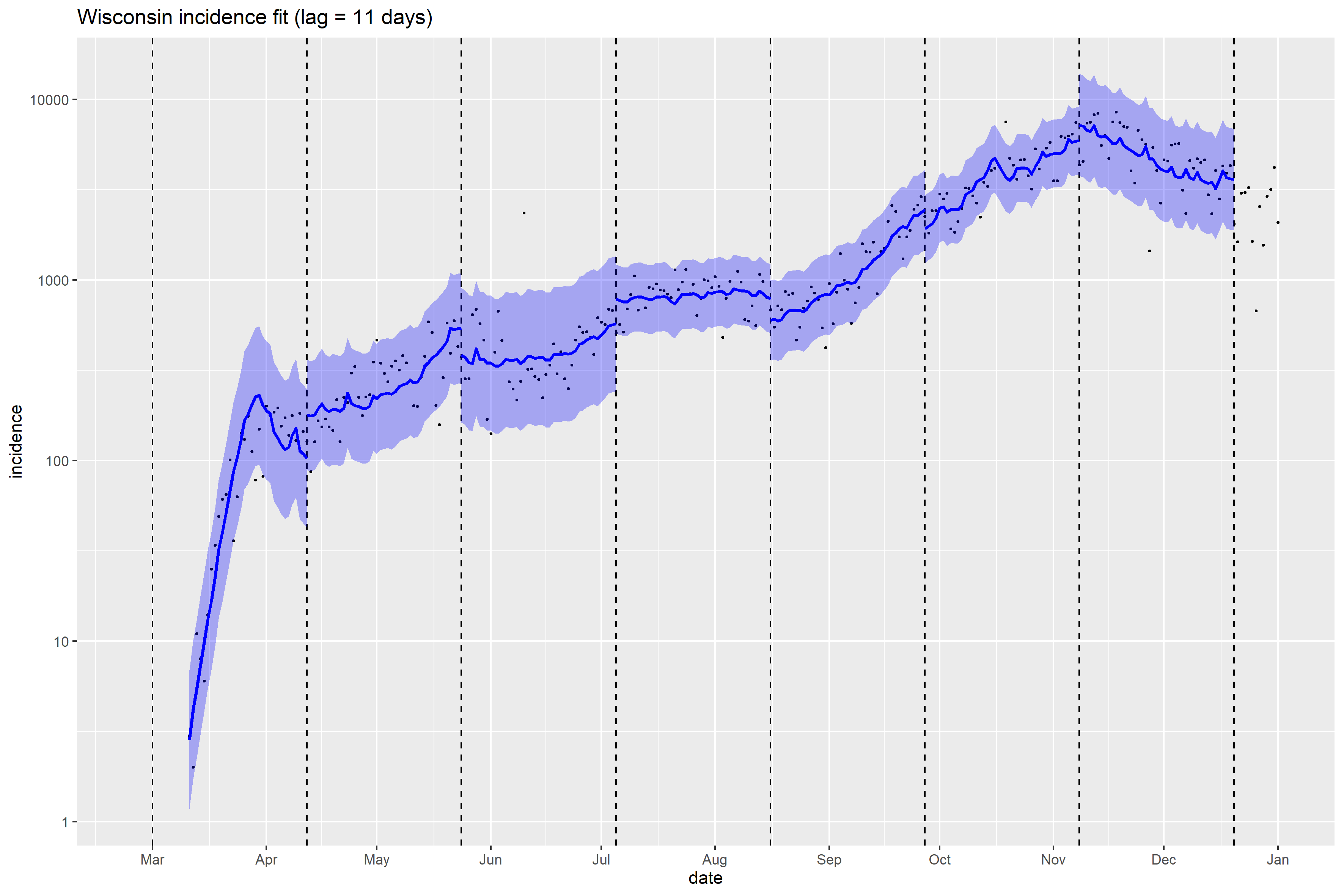

### Wyoming_full_fit_plot.png

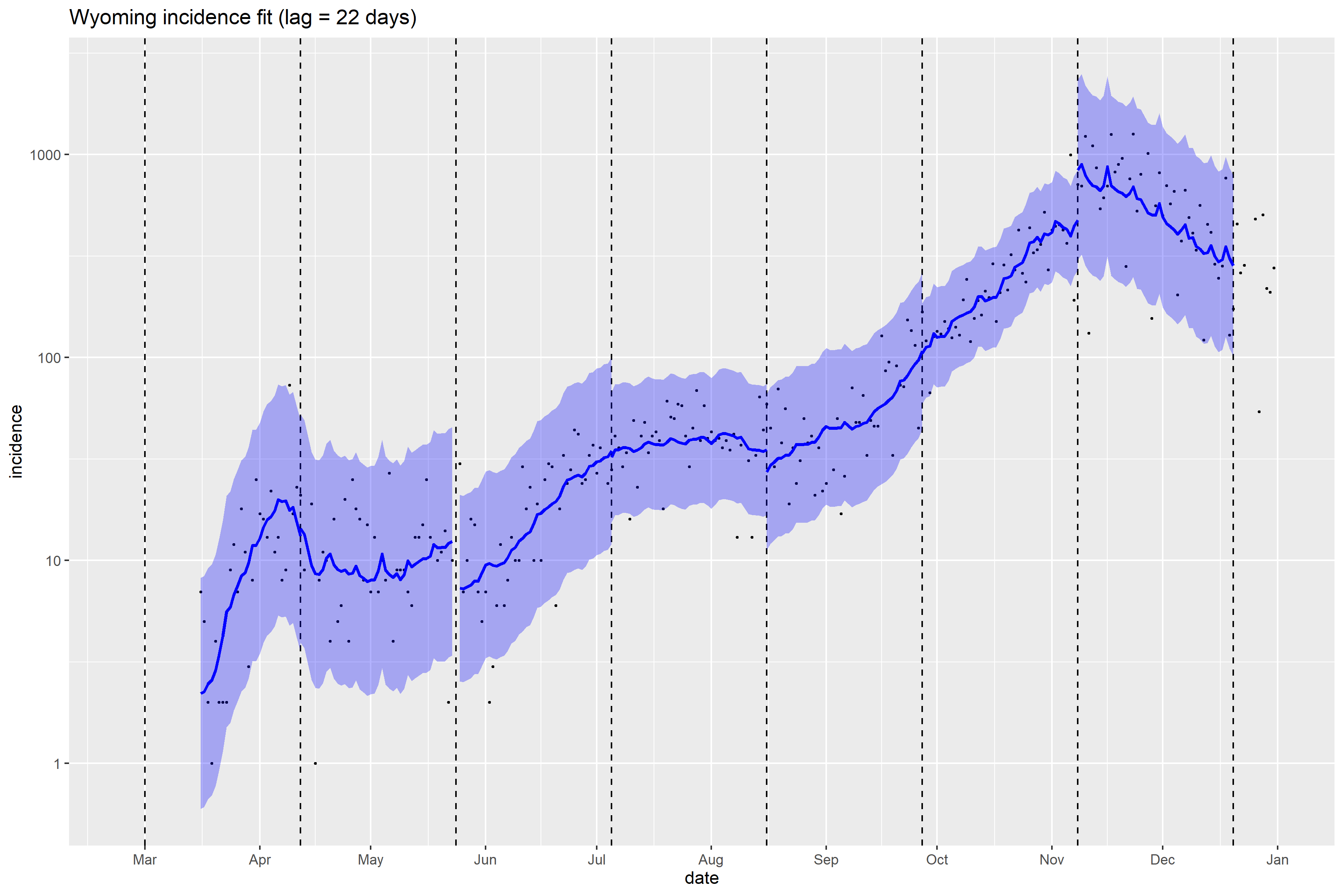
